# Reconstructed influenza A/H3N2 infection histories reveal variation in incidence and antibody dynamics over the life course

**DOI:** 10.1101/2024.03.18.24304371

**Authors:** James A. Hay, Huachen Zhu, Chao Qiang Jiang, Kin On Kwok, Ruiyin Shen, Adam Kucharski, Bingyi Yang, Jonathan M. Read, Justin Lessler, Derek A. T. Cummings, Steven Riley

## Abstract

Humans experience many influenza infections over their lives, resulting in complex and varied immunological histories. Although experimental and quantitative analyses have improved our understanding of the immunological processes defining an individual’s antibody repertoire, how these within-host processes are linked to population-level influenza epidemiology remains unclear. Here, we used a multi-level mathematical model to jointly infer antibody dynamics and individual-level lifetime influenza A/H3N2 infection histories for 1,130 individuals in Guangzhou, China, using 67,683 haemagglutination inhibition (HI) assay measurements against 20 A/H3N2 strains from repeat serum samples collected between 2009 and 2015. These estimated infection histories allowed us to reconstruct historical seasonal influenza patterns and to investigate how influenza incidence varies over time, space and age in this population. We estimated median annual influenza infection rates to be approximately 18% from 1968 to 2015, but with substantial variation between years. 88% of individuals were estimated to have been infected at least once during the study period (2009-2015), and 20% were estimated to have three or more infections in that time. We inferred decreasing infection rates with increasing age, and found that annual attack rates were highly correlated across all locations, regardless of their distance, suggesting that age has a stronger impact than fine-scale spatial effects in determining an individual’s antibody profile. Finally, we reconstructed each individual’s expected antibody profile over their lifetime and inferred an age-stratified relationship between probability of infection and HI titre. Our analyses show how multi-strain serological panels provide rich information on long term, epidemiological trends, within-host processes and immunity when analyzed using appropriate inference methods, and adds to our understanding of the life course epidemiology of influenza A/H3N2.

## Introduction

Patterns of influenza infections in humans are highly varied across time, space and demography [1,2]. Recurrent epidemics occur because influenza viruses undergo an evolutionary process of antigenic drift, whereby new strains escape pre-existing host immunity through the accumulation of mutations in immunodominant surface glycoproteins leading to rapid turnover of lineages, with specific strains persisting for 1-2 years [3,4]. Because individuals are alive at different times and locations, they are exposed to different strains and thus each individual has a distinct immunological history [5,6]. As a result, serological data suggest that humans are infected with a new A/H3N2 influenza strain approximately every 5 years, with less frequent infections, or at least less frequent detectable antibody boosts, as individuals enter middle age [7,8].

A better understanding of who, where and when influenza infections are likely to occur would aid in public health planning, nowcasting and forecasting [9,10]. However, it is not just antigenic variation and evolution that contributes to variation in influenza incidence, but a combination of individual and population level factors [11,12]. Birth cohorts [13–15], contact and movement patterns [16–18], climatic variation [19,20], school terms [21,22], city structure [23,24], and household structure [25,26] have all been shown to be associated with variation in influenza incidence. However, variation in surveillance quality and consistency across locations and over time makes it difficult to identify individual-level or population-specific effects over a longer time period using routine influenza-like-illness surveillance data [27,28]. These limitations may be overcome by using serological data, where unobserved past infections and vaccinations leave a signature in an individual’s measurable antibody profile [29–31].

For influenza, measured antibody levels are the result of complex interactions of immunological responses from all past exposures [6,32]. Hence, accurate inferences of individual infection histories require models of antibody kinetics to determine the number and timing of past exposures to multiple influenza strains [8,13,33–35]. These models can be complicated, as immunological interactions of antigenic drift with immune memory occur through imprinting effects, whereby the set and order of strains in an individual’s previous exposure history influences which epitopes are targeted and the magnitude of their antibody response to subsequent exposures [6,32]. Estimating influenza infection histories from serological data therefore presents a decoding problem, as the space of possible exposure histories which could lead to an observed antibody landscape is large, and observed antibody titres are highly variable due to within-host and laboratory-level effects. Although inferences which account for these mechanisms have provided rich insights into individual-level life course immune profiles, most attempts have been in relatively small cohorts or using small panels of influenza strains, limiting the conclusions which can be drawn about population-level influenza epidemiology [13,36,37].

Here, we applied an infection history inference method to data from a large serosurvey to reconstruct lifetime individual infection histories and population-level incidence of A/H3N2 influenza in Guangzhou, China [35,36,38]. Infection histories were inferred based on individual-level antibody profiles to a panel of 20 influenza A/H3N2 strains representing viruses that first circulated from 1968 onward. The study population comes from a range of age groups, social backgrounds, and geographical areas, thereby providing an ideal dataset to investigate predictors of influenza infection and small-scale spatial variation. In fitting the model, we also obtained parameter estimates for the underlying antibody kinetics model which allows us to elucidate long- and short-term antibody dynamics. Together, these results provide detailed spatiotemporal insights into the historical epidemiological and immunological dynamics of influenza A/H3N2

## Results

### Description of participant data

We measured 67,683 HI titres against 20 A/H3N2 strains isolated between 1968 and 2014 at 2-3 year intervals from serum samples collected between 2009-12-22 and 2015-06-02 (Fig S1) as part of the Fluscape cohort study (1,130 individuals, 2 samples each) (Fig S2 and Fig S3). Sampling was done over four study rounds, with a mean time between serum sample collection of 3.87 years (standard deviation: 0.780; 95% quantiles: 1.68-4.74 years). This cohort covered 40 unique locations and 651 unique households in a 60 kilometer transect from Guangzhou, China. Participant ages at the most recent sampling round ranged from 6 years to 97 years, with a median of 50 years. Vaccination rates were low in this cohort (Table S1), consistent with low reported vaccine coverage rates in mainland China, particularly in older individuals [39,40]. We refer to an individual’s set of antibody titres against all strains in the panel as their antibody profile. All individual antibody profiles and observed changes in titre are shown in Fig S20&S21. Summary statistics of these profiles and full study details have been described elsewhere [38,41].

### Antibody titres vary by age and in space

We saw five broad patterns of seropositivity (HI titre >= 1:40) when stratifying the antibody data by age group and strain (Fig 1). First, individuals had mostly low or undetectable titres against strains that circulated before they were born (cells below the black lines in Fig 1), though many individuals were seropositive against the strain which circulated in the years immediately prior to birth. Second, in the youngest age group (0-10 years old), many individuals were also seropositive to pre-birth strains that circulated further back in time (A/California/2004, A/Fujian/2002, A/Fujian/2000 and A/Victoria/1998), indicating the presence of cross-reactive antibodies as these individuals could not have been exposed to those strains (Fig 1C). Third, seroprevalence tended to be high among individuals who were young when a strain was first isolated (cells just above the black line in Fig 1) compared to individuals who were older at the time that the strain was first isolated. Fourth, the total proportion of individuals who seroconverted between sample collection dates was high against recent strains at nearly 50% across all age groups. Finally, some strains exhibited systematically higher titres than others, for example, titres against A/Fujian/2002 and A/Mississippi/1985 were higher than all other strains and particularly high for individuals who were under 10 years old at the time of their first circulation.

**Figure 1:**
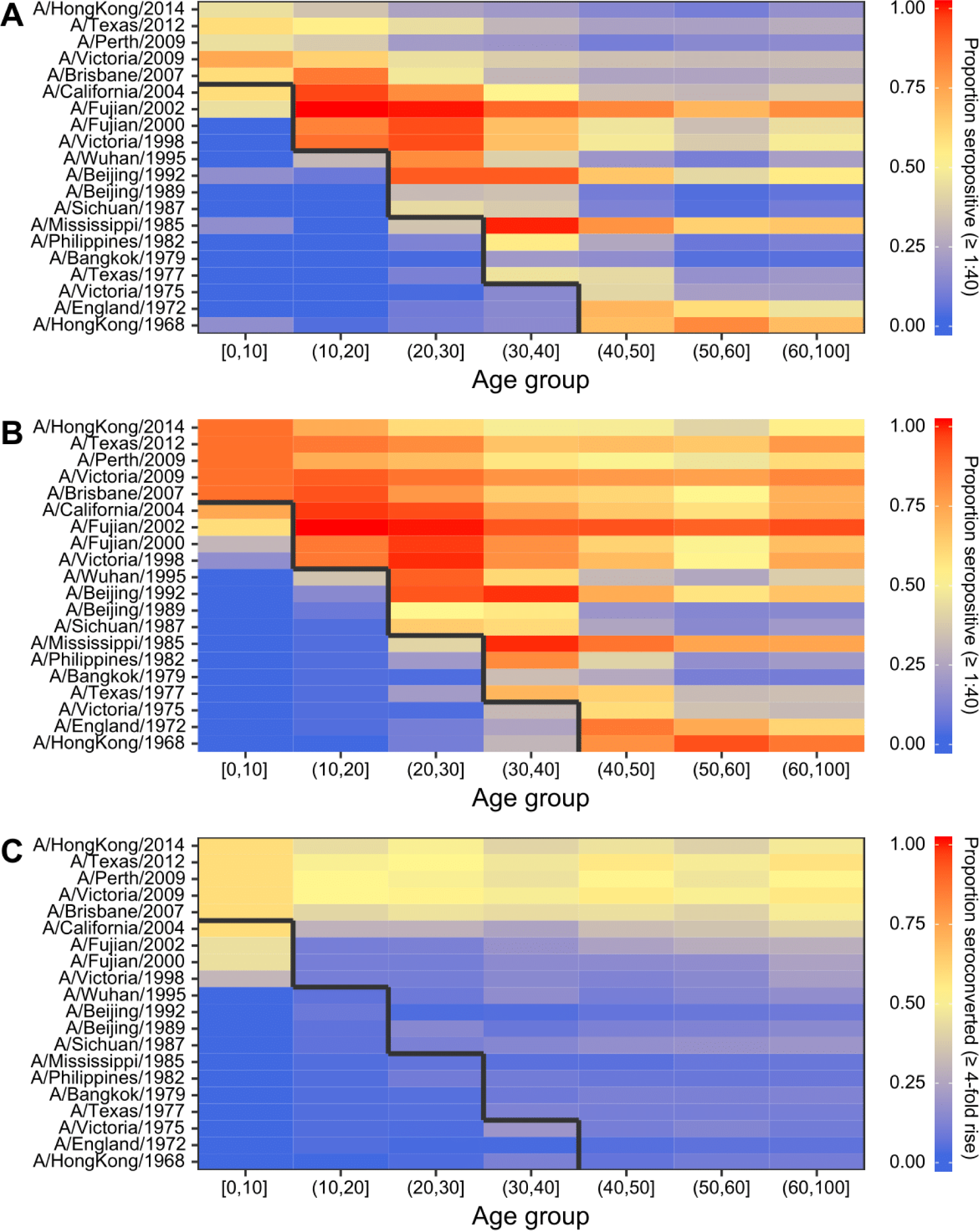
Proportion of individuals seropositive and seroconverted to 20 A/H3N2 strains circulating from 1968 to 2014 stratified by age. Solid black line divides age groups that were alive or not at the time of strain circulation. Seropositivity was defined as having an HI titre of ≥1:40 (a log titre of 3). (**A**) First serum sample. (**B**) Second serum sample. (**C**) Seroconversion between samples, defined as a ≥4-fold increase in HI titre.

### Inferring antibody kinetics and individual infection histories from antibody profiles

To infer the set of A/H3N2 influenza strains each individual was infected with, we further developed and used the *serosolver* R-package to fit an infection history and antibody kinetics model to all individual-level antibody profiles [35]. Briefly, the method finds the combination of influenza strains which an individual is most likely to have encountered conditional on their antibody profile, accounting for cross-reactive, transient antibody boosting and antigenic seniority arising from repeated exposures to antigenically related strains (see Materials and Methods). Individuals can be infected with the strain assumed to be circulating in each time period provided they were alive at the time of circulation. Individuals have distinct infection histories, but the parameters governing post-infection antibody kinetics were assumed to be universal, and individual infection probabilities were assumed to arise from a single population-level infection probability parameter per time period, regardless of location. A crucial component of the model is the observation level, which accounts for the fact that some strains elicit systematically higher or lower titres than others in the HI assay (see Supplementary Text 1).

We routinely fitted the same model to both the Fluscape cohort data and to previously published antibody profile data from a cohort of 69 individuals in Ha Nam, Viet Nam (see Materials and Methods and [37]), and compared the inferred antibody kinetics and epidemic dynamics. Posterior distributions for the model predicted titres and infection histories compared to observed titres are shown for five randomly selected individuals from the Fluscape cohort in Fig S4, and from the Ha Nam, Viet Nam cohort in Fig S5. Overall, we imputed a total of 10,558 (posterior median; 95% CrI: 10,394-10,750) distinct infections (see *Post-processing of infection history posteriors* in Materials and Methods) across all individuals and times from the Fluscape data and 547 (posterior median; 95% CrI: 519-574) infections from the Ha Nam, Viet Nam data. We also ran varied scenario analyses using simulated data closely resembling the Fluscape data, demonstrating that our inference system was able to accurately recover infection histories, attack rates and antibody kinetics parameters under a range of assumptions and model misspecifications (Supplementary Text 2).

### Antibody kinetics parameters

We obtained estimates for the antibody kinetics model parameters assumed to underlie the generation of observed influenza antibody titres. We estimated parameter values consistent with an initial, broadly reactive antibody response that decays within approximately one year to leave an antigenically narrow, persistent antibody response. These estimates are in line with previous estimates using a similar method applied to a smaller pilot serosurvey from the Fluscape cohort [36], as well as from model fits to the Ha Nam, Viet Nam cohort data (Table 1). However, there were some differences in the estimated magnitude of the antibody response between the Fluscape and Ha Nam datasets. Estimates from the Fluscape data suggested that infection elicited an antigenically narrow, long-term antibody boost of 1.42 log HI units (posterior median; 95% CrI: 1.39-1.44) compared to 1.96 log HI units (posterior median; 95% CrI: 1.89-2.03) from the Ha Nam dataset, and an additional antigenically broad, short-term boost of 2.06 log HI units (posterior median; 95% CrI: 1.99-2.14) compared to 2.65 log HI units (posterior median; 95% CrI: 2.43-2.92). The estimated waning rate of the short-term response was similar from the two datasets, suggesting that the short-term response took 0.995 years (posterior median; 95% CrI: 0.963-1.03) to fully subside based on the Fluscape data and 1.33 years (posterior median; 95% CrI: 1.22-1.45) based on the Ha Nam data. Finally, the observation error standard deviation was estimated to be larger for the Ha Nam dataset at 1.29 (posterior median; 95% CrI: 1.27-1.31) vs. 0.636 (posterior median; 95% CrI: 0.631-0.641) (posterior medians and 95% CrI), though this is likely attributable to the different assumed observation model and measurement of more influenza strains in the latter dataset.

**Table 1:**
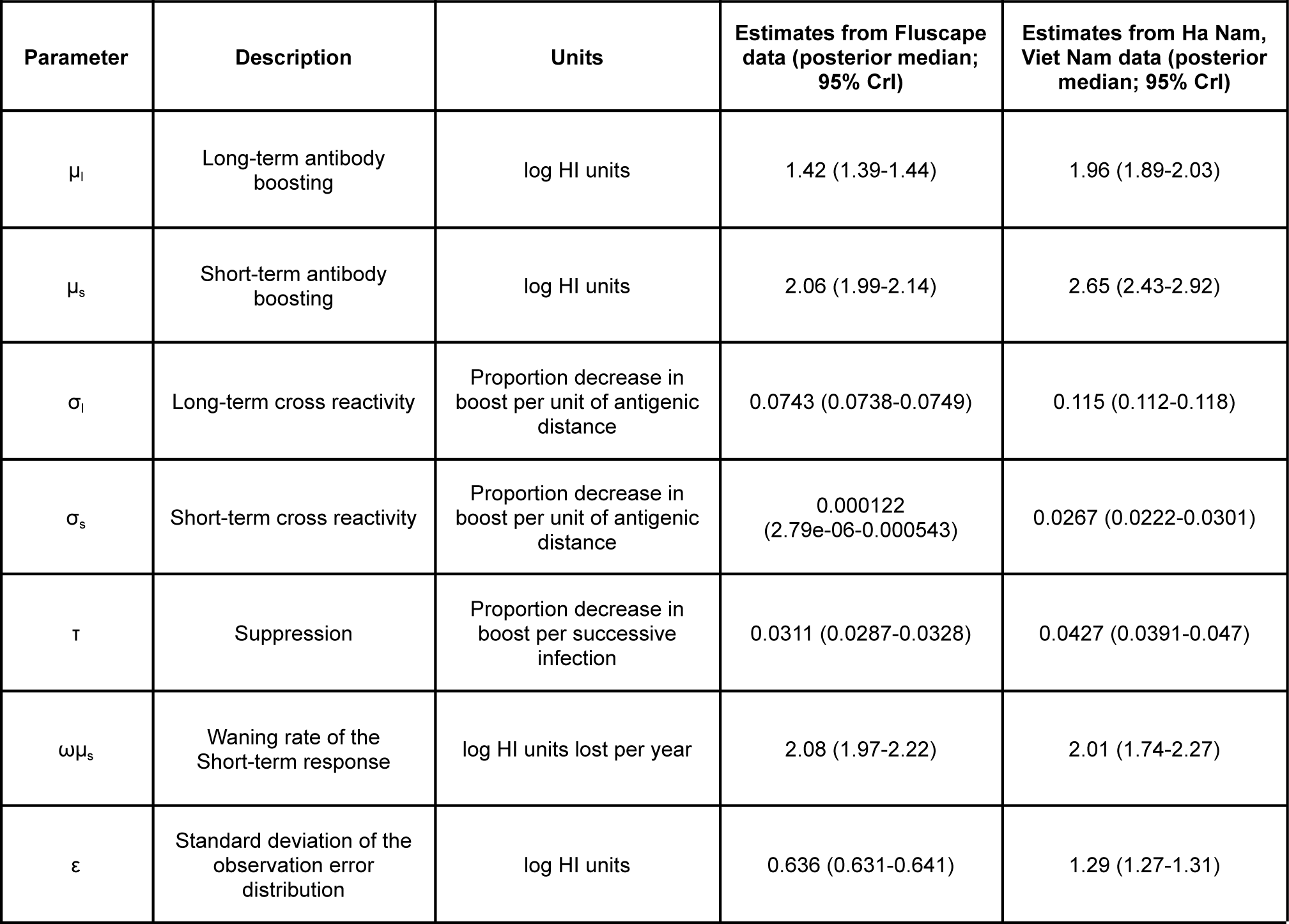
Estimated antibody kinetics parameters.

### Inferred historical and contemporary attack rates

By combining all individual-level inferred infection histories, we obtained A/H3N2 incidence estimates in the Fluscape cohort for each 3-month window since 1968 (Figure 2A). The estimated median quarterly sample attack rate was 3.56% (median across all posterior samples; 95% CrI: 3.13-4.00%). This corresponded to a median annual attack rate of 17.8% (posterior median; 95% CrI: 15.7-19.9%), defined as the proportion of individuals who experienced at least one infection within a calendar year. This was comparable to estimates from the Ha Nam, Viet Nam dataset, which gave an estimated median annual attack rate of 18.6% (posterior median; 95% CrI: 14.5-22.9%).

**Figure 2:**
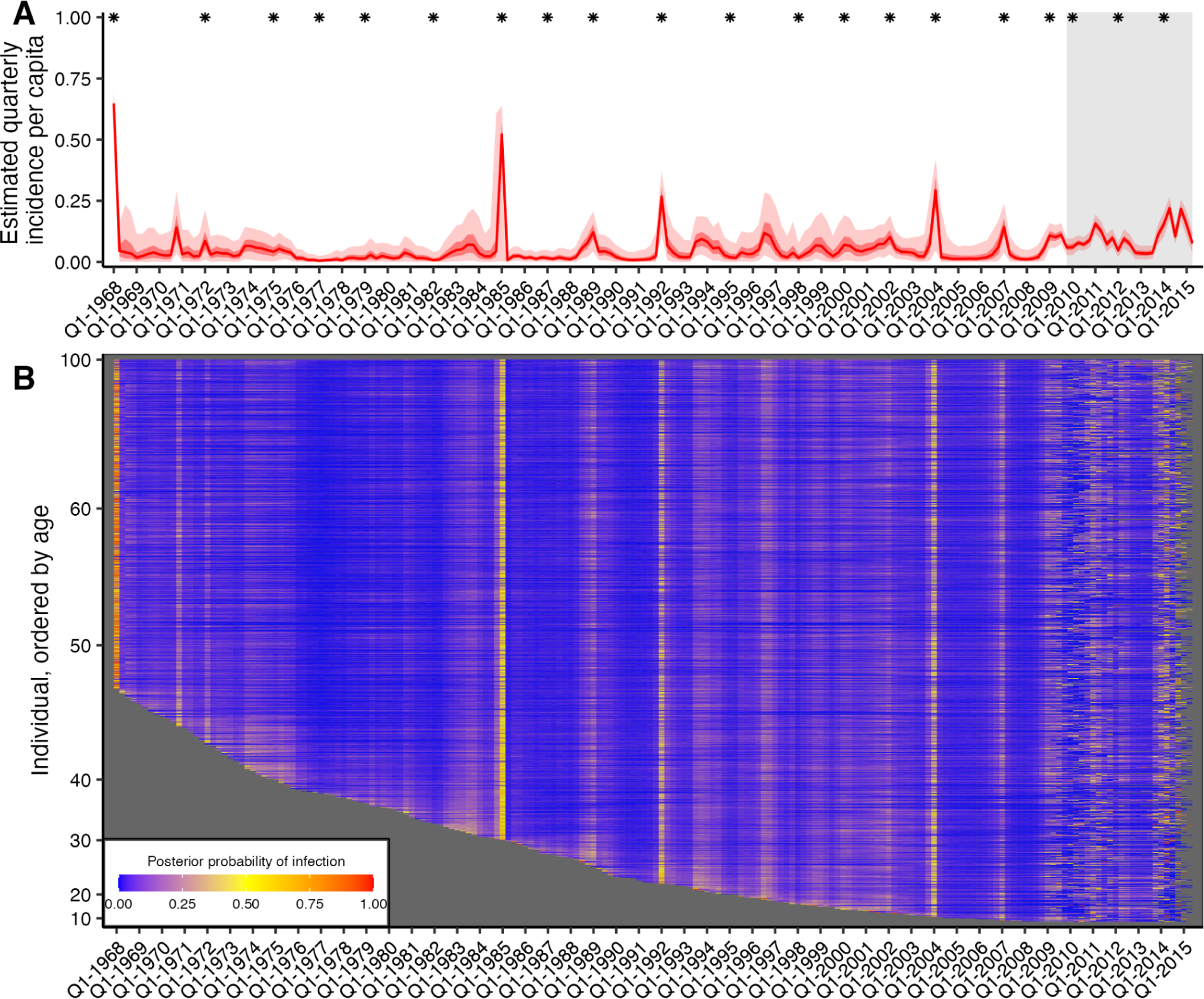
Quarterly incidence and individual infection histories from the Fluscape dataset. (**A**) Model predicted per-capita incidence per quarter. Attack rates were estimated by dividing the number of inferred infections by the number alive in each 3 month period. Red line shows the posterior median estimate from 1000 posterior samples. Dark and light red shaded regions show 50% and 95% credible intervals respectively from 1000 posterior samples. Gray shaded box shows duration of the Fluscape study – the improved precision is due to the inclusion of sera bracketing this time period. Asterisks mark times from which a strain included in the HI panel was first isolated. (**B**) Inferred infection histories for each individual. Each row represents an individual ordered by increasing age in years. Each column represents the time of a potential infection. Cells are shaded based on the proportion of posterior samples with an infection at that time.

Quarterly A/H3N2 attack rate estimates in the Fluscape cohort varied over time, ranging from a minimum of 0.459% (posterior median; 95% CrI: 0.00-2.52%) in Q1-1977 (during the re-emergence of A/H1N1) to a maximum of 64.8% in Q1-1968 (posterior median; 95% CrI: 53.6%-71.1%) (at the beginning of the A/H3N2 pandemic). The attack rate estimate for Q1-1985 was unusually high (54.3% posterior median; 95% CrI: 0.877%-65.5%), suggesting that there might be residual bias from systematically higher titres for the A/Mississippi/1985 virus not captured by the antibody kinetics and measurement models. Periods of high and low attack rates were reasonably well synchronized between the Fluscape and Ha Nam datasets (Fig S6). For example, both datasets gave very high attack rates for 1968, 1989 and 2009, and similar attack rates from 1970 to 1985 (though with substantial uncertainty in the Ha Nam estimates due to the much smaller sample size). There were also periods with clear differences – attack rate estimates were much lower in the Fluscape cohort in the early 2000s, and higher during 2010-2012.

Quarterly attack rate estimates were substantially higher in the Fluscape cohort after the first serum sample in Q4-2009 than the overall average, with the median quarterly attack rate since Q4-2009 estimated to be 8.48% (median of posterior samples for median per-quarter attack rate; 95% CrI: 7.26%-9.93%) versus 2.98% before (95% CrI: 0.748-13.7%). Annual attack rates (defined as the proportion of individuals who were inferred to have been infected at least once per year) fluctuated, with lower attack rates in 2010, 2012 and 2013, and high attack rates in 2011 and 2014 (Table 2). These attack rate estimates were of a similar magnitude to the proportion of individuals that seroconverted between study visits against strains that circulated during that time. Surveillance data collected from the Guangdong Provincial Centre for Disease Control and Prevention influenza surveillance system during the same time period reported slightly different dynamics, with an increased number of A/H3N2 virus isolates in Q2/Q3-2009, Q3-2010, Q4-2011 and Q1/Q2-2012, as well as a higher proportion of A/H3N2 isolates in 2011 and 2012 [42]. However, direct comparison to our estimated attack rates is difficult, as the influenza surveillance network only collects nasopharyngeal swab samples from patients presenting to sentinel hospitals with influenza-like-illness whereas our estimates relate to all exposure events regardless of symptoms.

**Table 2:** Estimated attack rates and infection patterns 2010-2014. Percentages shown are posterior median and 95% credible intervals. “Attack rate” was defined as the proportion of individuals who were infected at least once in that year. “Seroconverted” gives the percentage of individuals that seroconverted to the measured strain, with ranges showing 95% binomial confidence intervals. “Reinfected” gives the percentage of people that were infected more than once in a year. Bottom table shows the percentage of individuals that were infected 0, 1, 2, 3, 4 or 5 times between 2010-2014 inclusive.

| Year | Measured strain | Seroconverted (%)<br>N=1127 | Estimated attack<br>rate (%) | Reinfected within<br>same year (%) |
| --- | --- | --- | --- | --- |
| 2010 | A/Perth/2009 | 47.7% (44.8%-50.6%) | 28.5% (24.8%-31.8%) | 1.68% (0.971%-2.57%) |
| 2011 | - | - | 41.2% (37.9%-44.1%) | 1.52% (0.537%-2.60%) |
| 2012 | A/Texas/2012 | 50.7% (47.8%-53.6%) | 24.1% (20.1%-27.7%) | 1.17% (0.361%-2.26%) |
| 2013 | - | - | 20.5% (16.1%-24.7%) | 3.40% (2.36%-4.54%) |
| 2014 | A/Hong Kong/2014 | 42.6% (39.7%-45.5%) | 56.4% (53.6%-59.3%) | 2.44% (1.64%-3.23%) |

| Proportion of individuals with different total number of infections from 2010-2014 |  |  |  |  |  |
| --- | --- | --- | --- | --- | --- |
| 0 | 1 | 2 | 3 | 4 | 5 |
| 12.4%<br>(11.4%-13.5%) | 33.5%<br>(31.7%-35.7%) | 34.0%<br>(31.8%-36.1%) | 15.9%<br>(14.1%-17.8%) | 3.81%<br>(2.74%-4.96%) | 0.354%<br>(0.0885%-0.708%) |

A substantial proportion of people were estimated to have been reinfected within a single year, with higher reinfection rates in years with higher overall attack rates. 36.5% (posterior median; 95% CrI: 29.7%-43.1%%) of all reinfections between 1968 and 2015 occurred since 2008, whereas only 14.9% of possible infection events were in this time frame, suggesting that reinfections were disproportionately more likely in recent time periods. Annual reinfection rates, defined as having been infected at least once in a year for multiple years, were also high for recent years (Table 2).

### Attack rates varied in space, but were not predicted by proximity

Next, we grouped the posterior draws for the Fluscape infection histories by study location to investigate spatial patterns in attack rates. Attack rates exhibited variation between locations and over time (Fig S7), though the timing of high attack rate periods was synchronized. Fig S8 shows snapshots from a video of attack rates over time across the study region, demonstrating that the periods of high influenza incidence are synchronized across the study locations, but that there is some variation in the timing and magnitude of incidence (full video in Supplementary Material 3). The overall coefficient of variation (CoV) for per-quarter attack rates across all locations and times was 1.12 (posterior median; 95% CrI: 1.04-1.19). This was reduced to 0.493 (posterior median; 95% CrI: 0.458-0.535) after aggregating the infection histories into per-year attack rates, defined as the proportion of individuals infected at least once in a calendar year. There was a strong negative correlation between the estimated posterior median CoV and per-quarter attack rate (Pearson correlation coefficient of −0.829).

To understand if this variation reflects epidemiological differences between locations or simply sampling variation, we generated a comparable null simulation where no significant spatial variation in incidence would be expected (see *Spatial correlation in inferred attack rates* in Materials and Methods). The mean CoV for these simulations was 0.300 (95% quantiles: 0.0307-1.22), suggesting that the attack rates in the Fluscape data exhibited no more variation overall than would be expected by chance if all attack rates were drawn from the same binomial distribution. Fig S9 demonstrates that this pattern is maintained across time, though with higher variation in time periods with low infection rates. Fitting a spatial non-parametric correlation function revealed high correlation in attack rates between locations which did not decline with increasing distance (Fig S9D). This consistent correlation across space was also observed when subsetting attack rates by recent (from Q1-2009 onward) or historical times (from Q1-1968 to Q1-2009), when considering either per-quarter or per-year attack rates, and also when considering the proportion of individuals in a population who seroconverted against all or only recent strains.

### Age-specific infection patterns

Periods of high infection probability were largely synchronized across all individuals regardless of age, though individuals typically experienced more frequent infections in the years immediately following birth (Fig S10). Two age-specific patterns emerged. First, almost all individuals who were alive in 1968 were almost certainly infected in or around 1968, demonstrated by the high posterior probability of infection across all individuals alive at that time. Second, the posterior probability that an individual was infected soon after birth was consistently high, demonstrated by the lower edge of the heatmap in Figure 2B and Fig S10. The posterior mean, median and 95% CrI on the age of first infection was 1.36, 0.75, and 0.00–6.00 years respectively (estimates using 1000 posterior samples for all individuals born since 1968). These results suggest that based on these augmented infection histories, individuals were infected soon after birth, in contrast to previous findings [43].

We calculated each individual’s age at the time of each infection and found that the number of infections per 10 year period decreased through childhood and became stable in adulthood (Figure 3C; Fig S11). Overall, individuals were estimated to be infected 2.12 times per 10 year period (posterior median; 95% CrI: 1.06-7.86), in line with previous estimates (Figure 3D) [36]. Infection frequency patterns and trends with respect to age were similar under different assumptions for the infection history model, suggesting that these findings were driven by features of the data and not an artifact of the model structure (Fig S12; [35]).

**Figure 3:**
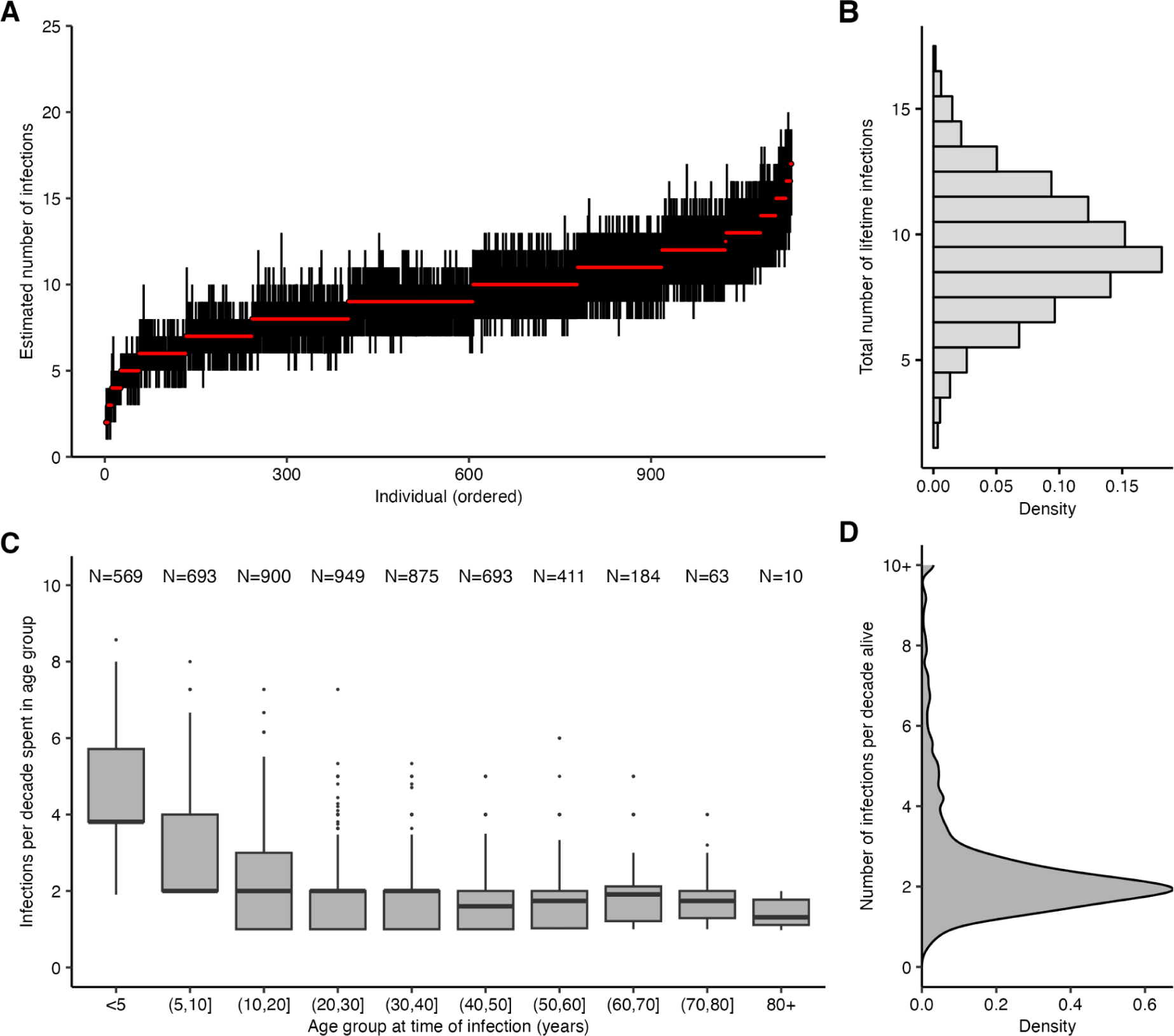
Age-specific patterns of infection. (**A**) Pointrange plot shows posterior median and 95% CrI on the total number of lifetime infections for each individual in the Fluscape cohort, ordered by increasing age at time of sampling. (**B**) Distribution of the total number of infections across all individuals based on the posterior median total number of infections. (**C**) Posterior median number of infections per 10 year period stratified by age group at the time of infection, excluding individuals who spent less than 2 years in that age group and including only time periods prior to the first serum sample in Q4-2009 (see Fig S11 for explanation and comparison using all time periods). Text shows sample size within each age group – note this does not sum to the number of individuals in the sample, as individuals contribute to multiple age groups during their lifetime. (**D**) Distribution of individual posterior median number of infections per 10 years alive.

### Relationship between titre and probability of infection

Our model fits estimated not only infection histories, but also each individual’s expected HI titre against all strains at each point in time, and thus we were able to estimate the relationship between probability of infection and model-predicted latent HI titre (Figure 4). For each sample from the posterior, we found the proportion of time periods (across all times and individuals) where infection was estimated to have occurred, stratified by log HI titre against the circulating strain in that time period. There was a clear pattern of decreasing relative risk of infection as a function of increasing titre (Figure 5). An HI titre of 1:40 (log titre of 3) corresponded to a risk of infection of 0.543 (posterior median; 95% CrI: 0.508–0.591) relative to individuals with no detectable titre, consistent with prior evidence from deliberate infection experiments [44]. This pattern appeared to vary with age. An HI titre of 1:40 gave a relative risk of infection of 0.360 (posterior median; 95% CrI: 0.308–0.399) in the 0–10 age group, but only 0.705 (posterior median; 95% CrI: 0.571–0.834) in the 60+ age group.

**Figure 4:**
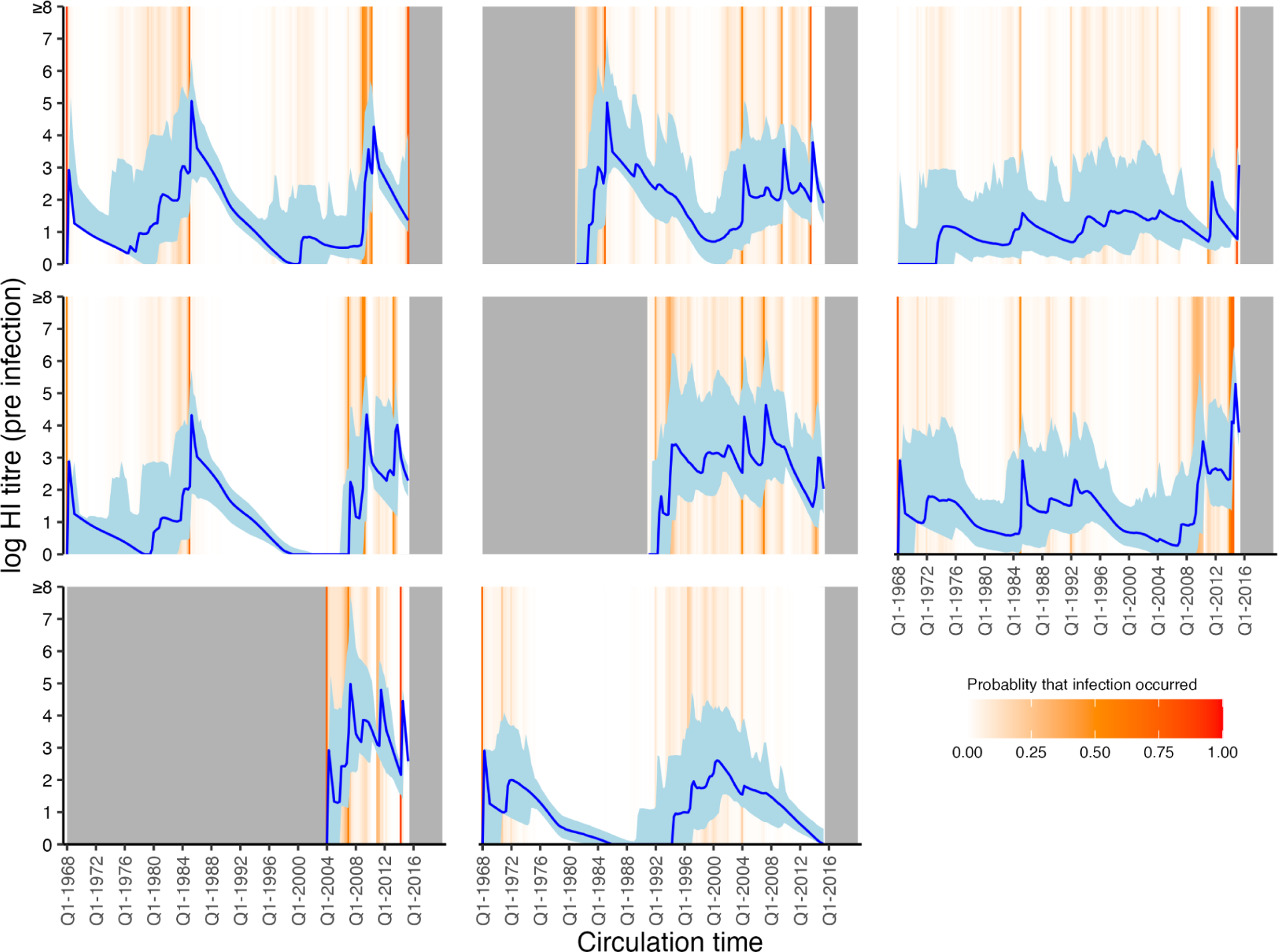
Model predicted titres against circulating strains since birth. Each subplot shows one randomly selected individual. X-axis shows time since birth. Blue line and shaded region show model-predicted, true latent titre against the strain assumed to be circulating at each time period (posterior median and 95% CrI). Note that titres are continuous and represent latent, true values, not observations. Orange lines indicate times of high posterior probability of infection. Grey regions show times before birth and after the last serum sample.

**Figure 5:**
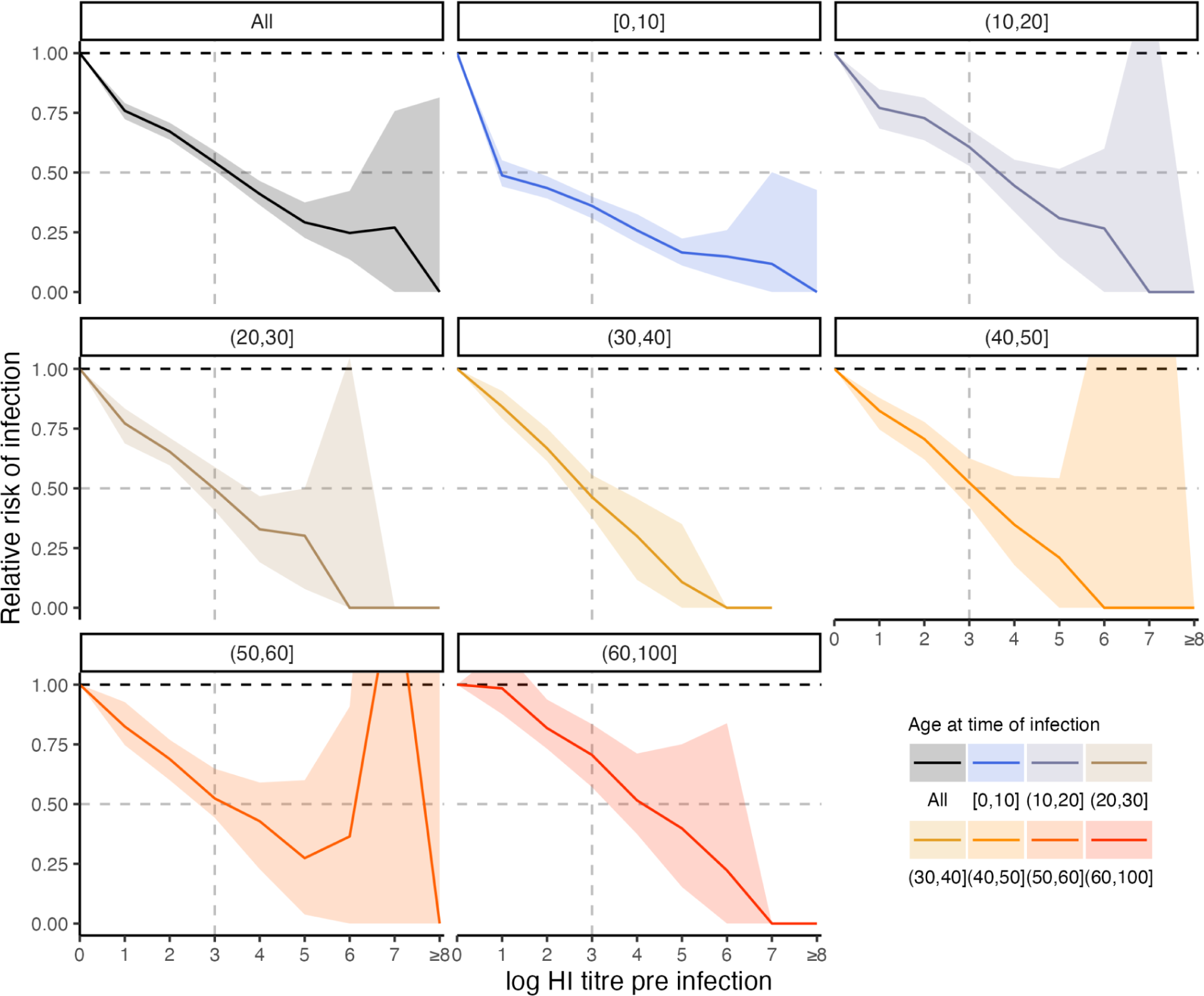
Estimated relationship between HI titre and probability of infection. Top left panel shows the relative risk of infection at all time points stratified by model-predicted log titre against the circulating strain just before infection for all age groups. Remaining plots show the same relationship but stratified by age group in 10-year bands at the time of infection. Solid lines and shaded regions show posterior median and 95% CrI. Note that the uncertainty intervals reflect uncertainty in the imputed infection states and latent antibody titres; the relationships shown here are empirically calculated from the *serosolver* estimates. Vertical dashed line shows HI titre 1:40. Horizontal grey dashed line shows 50% protective titre.

## Discussion

Influenza A/H3N2 infection histories and attack rates exhibited substantial variation across time and locations in a cohort of 1,130 individuals around Guangzhou, China. We considered that variation in influenza antibody titres may be generated in three ways: (i) exposure to different combinations of viruses at different times; (ii) time-dependent antibody kinetics observed at different times relative to an exposure; (iii) random and strain-specific, systematic variation in the HI assay. We showed that accounting for these mechanisms in a modeling framework allowed for the reconstruction of each individual’s complete A/H3N2 infection history since birth conditional on their antibody profile. We reconstructed population-wide and location-specific historical infection, or, more precisely, seroincidence rates from these infection histories, finding that influenza infection incidence may be higher than suggested by routine surveillance. Also, estimates of each individual’s true antibody titre against circulating strains for each 3-month period since birth were generated, showing that elevated antibody titres were associated with a substantial reduction in infection risk that became less effective with increasing age.

We estimated that the incidence of influenza A/H3N2 infections, or at least detectable serological responses, in both the Fluscape cohort and a smaller dataset from Ha Nam, Viet Nam was around 18% on average per calendar year since 1968. These annual seroincidence rate estimates were particularly high during the Fluscape study period in 2010–2014, ranging from 20% to 57% of individuals infected at least once per year, though the cutoff of an influenza season is not as clearly defined for this region as it is for temperate regions [2,45]. These values are higher than previous estimates based on serological data from Hong Kong for the same time period at 7-19% [34,46,47]. However, results from urban and rural South African communities (the PHIRST cohort) demonstrated similarly higher influenza incidence rates under routine RT-PCR testing for influenza A or B [48]. Similarly high incidence rates based on either anti-haemagglutinin or anti-neuraminidase seroconversion were also detected in New Zealand (the SHIVERS cohort) [49]. Our results and those from the PHIRST and SHIVERS studies are at the higher end of annual influenza incidence rate estimates [49–51], and suggest that some individuals may be infected with the same subtype multiple times within a year. Our results additionally demonstrate that incidence rates may have been higher in recent years, and provide estimates for historical A/H3N2 incidence rates back to 1968.

Reconstruction of lifetime infection histories from this large study population covering all age groups revealed that the frequency of influenza A/H3N2 serological responses is initially high and decreases with age and then remains stable at around two infections per decade through most of adult life. The pattern is consistent with previous findings from age-stratified epidemiological data [8,34,48,49,51,52], regression analyses of randomized control trial data [53], theoretical models incorporating age-specific differences in social behavior [54,55], and data from the Fluscape pilot study [7]. Reassuringly, the clear age patterns we find here were maintained under different assumptions for the infection history model, suggesting that these patterns emerge from information in the data rather than through the model structure. However, it is important to note that we have estimated the incidence of detectable antibody responses and not necessarily clinically relevant infections–not all infections lead to seroconversions and not all antibody boosts reflect detectable virus shedding [56,57]. Similarly, although our inference method did not distinguish between vaccination and infection, vaccination coverage in this cohort was very low and thus this assumption is unlikely to bias our infection history estimates substantially.

Although the spatial dynamics of influenza at a large scale are fairly well understood through the use of molecular data, explaining spatial dynamics at smaller scales has revealed contrasting results and remains the focus of ongoing work [12,18,22,58]. Comparable spatio-temporal analyses of seasonal influenza-like-illness (ILI) incidence surveillance data in Norway, Sweden, Denmark and the USA–regions with clearer ILI seasons than Guangdong–showed high spatial correlation in epidemic phase timing and amplitude between locations thousands of kilometers apart that declined with distance [59]. Our estimates show a high correlation of attack rates in space that did not change as a function of distance at a small spatial scale (less than 60km), suggesting that an individual’s life-course of seasonal influenza A/H3N2 infections is largely determined by when they were born and the epidemiology of their wider region, rather than precisely where they live. However, our results are limited as we do not account for the fact that individuals move over their lives, and thus their location recorded at the time their serum sample collected may not match their location earlier in life.

We inferred a negative correlation between log HI titre at the time of exposure and probability of seroresponse in line with the historic deliberate infection data of 50% protection for an HI titre of 1:40 [44,60–62]. We found that this relationship became weaker with age, consistent with recent work showing that although HI titre is a good correlate of protection in children, it is less robust in adults [8]. Non-HI-mediated protection, such as non-haemagglutinin head targeted antibodies or cellular responses, are likely to explain a greater amount of variability in immunity as individuals age [49,63]. The magnitude of our estimates do contrast with other work, which has found that the 50% protective titre threshold may be higher than 1:40 in young children [64,65]. As stated by Hobson et al. in 1972, care must be taken in assigning causality to the titre-mediated infection risk estimated here. In the present model, titres necessarily decreased over time following infection due to antigenic drift and short-term waning. If protection is governed by non-HI immunity that wanes at a similar rate, then the same association between titre and relative risk could be observed. Non-HI protection may also explain the findings of decreased infection frequency at older age despite decreasing titre-mediated protection, for example via non-haemagglutinin head targeted antibodies or cellular responses [49,63].

Finally, we generated estimates for post-exposure antibody kinetics from two serological datasets. There were a number of similarities and differences in the estimated antibody kinetics parameters based on the Fluscape and Ha Nam cohort datasets. Although differences in tested strains and between-lab protocols limits direct comparison [66], both datasets gave qualitatively similar estimates suggesting a transient, antigenically broad antibody boost that waned within two years following exposure to leave a persistent, antigenically narrow antibody boost. The antigenic breadth of the short-term boost was broader in the Fluscape estimates, though this may be due to the inclusion of fewer recent strains and more older individuals than in the Ha Nam cohort, requiring the model to give greater weighting to boosting of historical strains (back-boosting) [37]. We estimated that the transient arm of the seroresponse waned within one year based on the Fluscape data compared to 1.33 years based on the Ha Nam cohort data, which are broadly in line with previous observations of antibody titres waning to near baseline approximately one year post vaccination [37,67–69]. Overall, this supports previous work suggesting that accounting for sub four-fold rises in titre, issues arising from non-bracketed sera, and measurement effects may lead to greater sensitivity in identifying infections from paired titres [33,70,71].

Our study has a number of limitations. A key challenge underpinning almost all efforts to analyze quantitative antibody titre data is the variation in titres arising from laboratory processes rather than underlying epidemiology and immunology [66]. Although we adjusted our observation model to account for systematically lower or higher observed titres to particular strains, which may lead to under- or over-estimation of attack rates, unaccounted for biases may still remain. External validation or estimation of the offset terms would be useful, but is difficult to do across studies given variability in laboratory protocols and serum potency [66]. Also, our data were limited in their strain coverage, as we measured titres to only one strain for each ∼2 year period and assumed that that A/H3N2 antigenic evolution followed a smooth rather than punctuated trajectory through antigenic space, and thus we may infer infections with antigenic variants that an individual has never been exposed to [3]. As a sensitivity analysis (not shown), we did attempt to fit the *serosolver* model instead assuming punctuated changes through antigenic space, but we were unable to produce converged model fits due to the discretization of the parameter space and thus we do not present these results. However, we did explore the impact of this potential model misspecification using simulation-recovery experiments, and found that fitting a model assuming continuous antigenic changes over time when data were generated under a punctuated antigenic evolution model did not substantially bias antibody kinetics parameter or attack rate estimates, with the exception of slightly overestimating the short-term antibody waning rate (Supplementary Text 2). These assumptions may also partially explain why the inferred seroresponse rates did not strictly align with timings of high incidence based on viral isolate data in Guangdong and Hong Kong for the same time period [42,46].

Another limitation is the simplification of the antibody kinetics model used here. The assumption of a fixed effect term on boosting and waning for all individuals and infection events masks a substantial amount of individual-level and strain-level variation in kinetics which would be better described by random-effects terms. This is a necessary simplification to ensure identifiability of the post-exposure kinetics parameters whilst simultaneously inferring hundreds of thousands of latent infection states. Similarly, there are a number of immunological mechanisms, such as titre-dependent boosting and titre ceiling effects which we did not include in the model due to identifiability issues [37,72]. Our model is also limited in explaining back-boosting, as we assumed that cross-reactivity extends linearly across antigenic space. In the short-term antibody boosting arm, cross-reactivity was found to boost the entirety of antigenic space regardless of an individual’s age to account for the clear back-boosting of titres against strains encountered early in life. However, this model may be inappropriate for younger individuals, whose immune systems may not have encountered any of these historical antigens and would therefore have no targeted memory B-cells to stimulate. A model that distinguishes boosting of heterologous antibodies through back-boosting of the memory response as opposed to cross-reactive antibodies from a *de novo* response through targeting shared epitopes would provide a more realistic model of the observed, antigenically broad short-term boost.

Antibody landscapes based on traditional HI assays, as well as multiplex antigen array and deep mutational scanning data, are useful tools for understanding how immunity develops following repeated infection and vaccination to antigenically related viruses [73–78]. The approach and results shown here demonstrate how these antibody profiles can be used to reconstruct lifetime infection histories at a fine spatial scale, providing a new source of augmented data with which to understand long-term epidemiological trends for influenza and other antigenically variable pathogens such as SARS-CoV-2 [79–81].

## Materials and Methods

### Ethics statement

Study protocols and instruments were approved by the following institutional review boards: Johns Hopkins Bloomberg School of Public Health, University of Hong Kong, Guangzhou No. 12 Hospital, and Shantou University. Written informed consent was obtained from all participants over 12 years old, and verbal assent was obtained from participants 12 years old or younger. Written permission of a legally authorized representative was obtained for all participants under 18 years old.

### Cohort description – Fluscape

Influenza haemagglutination inhibition (HI) titres were obtained from a previously described cohort in Guangzhou, China, called the Fluscape study [38,41]. The Fluscape study is a serological, contact and demographic survey in and near Guangzhou, China. The study covers 40 locations randomly selected from a 60km cone-shaped transect extending from Guangzhou city center into the surrounding rural area. Latitudes and longitudes of each study location were assigned based on a central place (e.g., a street or village committee center). 60 households within 1km of each chosen location were randomly selected and contacted one-by-one until 20 households with at least one member willing to provide a blood sample and answer the survey questionnaire were contacted. The majority of locations were classified as rural (30/40), were between 20 and 80 minutes travel time from Guangzhou city center, and had a population density between 256 and 367,346 persons per 9 km grid cell. All household members aged 2 or above were eligible to participate. 5 ml of blood was taken for each blood sample from each visit. Here, we used data from a subset of serum samples, capturing 1,130 individuals who had two serum samples taken and analyzed from two different rounds of sampling between 2009 and 2015 inclusive. The month of sampling, age of participant, vaccination history and other socioeconomic covariates were available for all individuals.

### Serological data

Haemagglutination inhibition (HI) assays were performed for each sample to measure antibody titres against 20 A/H3N2 strains that circulated between 1968 and 2014 inclusive with approximately 2 year spacing between circulation years. Repeat titres were available for all of the strains tested from the second serum sample (23,686 repeat measurements in total). Titres were tested in serial 2-fold dilutions from 1:10 to 1:1280, with undetectable titres recorded as <1:10. The recorded titre reading was the highest dilution at which haemagglutination was still inhibited. For all analyses, titres were transformed to a *log_2_* scale (i.e., 2-fold dilutions), where *y=log_2_(D/5)*, giving log titres between 0 and 8 (undetectable titres were treated as a 0 log titre). Seroconversion between study visits was defined as a 4-fold rise in titre, equivalent to a ≥2 unit increase on the log scale. Seropositivity was defined as having a titre of ≥1:40 (log titre ≥ 3). Further details on laboratory testing have been described previously [41].

### Summary of model

The overall inference task is to obtain estimates for the joint posterior distribution of antibody kinetics parameters (***Θ***), infection histories (***Z***) for all *n* individuals, and the attack rate within each of *m* possible discrete infection periods conditional on the set of observed HI titres (***Y***). Crucially, only ***Y*** is observed, so we must infer (or augment) the values of ***Z*** as latent features. Throughout the remainder of the methods, we use capital letters to represent random variables and bold letters to represent vectors of random variables. A detailed description of the inference problem and approach is described in [35], but can be summarized as sampling from the posterior distribution:

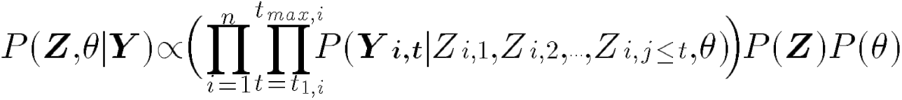

where ***θ*** is the vector of antibody kinetics parameters that describes the link between ***Z*** and ***Y***. The set of times *t_1,i_* to *t_max,i_* gives the time periods when a serum sample was obtained from individual *i. P(**Y** | **Z, θ**)* is defined by the antibody kinetics and observation model, and 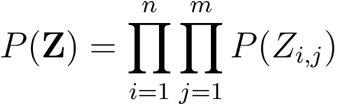 is the infection history prior, described below.

An individual’s entire infection history is given as a vector of unobserved binary variables, ***Z_i_*** *= [Z_i,1_, Z_i,2_, … , Z_i,j_]*. Each infection event, *Z_i,j_*, is the outcome of a single Bernoulli trial, where *Z_i,j_ = 1* indicates that individual *i* was infected with the strain circulating in discrete time period *j*, *Z_i,j_ = 0* indicates that they were not. The entire infection history matrix ***Z*** for all *n* individuals across all *m* time windows was therefore represented by an *n* by *m* binary matrix. We estimated infection histories at a 3-monthly resolution, such that each individual had an unobserved infection state for each 3-month period *j* since birth. Individuals could be infected from the first quarter after they were born. We did not attempt to impute infection states occurring after an individual’s last serum sample. The attack rate is then given as 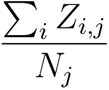, where *N_j_* gives the number of individuals alive in time period *j*.

Infections lead to the production of antibodies (“seroresponses”), that undergo longitudinal and cross-reactive kinetics. The vector of true, latent antibody titres across all time periods is given as ***A_i_*** *= [A_i,1_, A_i,2_, … , A_i,j_]*, which is generated from an antibody kinetics process with parameters ***θ*** described below. The process generating measured titres from the latent antibody titres is modeled through an observation level. The vector of observations is given as ***Y_i_*** *= [**Y_i,1_**, **Y_i,2_**, … , **Y_i,t_**]*. Note that the time index for ***Y_i_*** is different to ***A_i_*** and ***Z_i_***, as observations are only made at a subset of *t* times whereas latent infection states and antibody titres must be represented at all *j* times. ***Y_i,t_*** is also itself a vector, as it contains HI titres against all strains measured from that serum sample. A schematic of the full model is shown in Fig S13 and an example simulated antibody landscape over time is shown in Fig S14.

### Antibody kinetics, antigenic map and observation model

We used an existing deterministic model to describe the generation of observed antibody titres following exposure to an influenza strain [35,36]. The model has three components: (i) the antibody kinetics model; (ii) the antigenic map; and (iii) the observation model. The antibody kinetics model describes linear short- (*µ_s_*) and long-term (*µ_l_*) antibody boosting on the log scale immediately following infection. The short-term boost wanes over time such that the remaining short-term boost *j − k* time periods after infection at time *k* is given by *µ_s_w(k,j) = µ_s_max{0, 1 − ω(j-k)}*. Long-term boosting is persistent. In addition to boosting antibodies against the infecting strain, infection also elicits the production of cross-reactive antibodies against antigenically related strains. Cross-reactivity is assumed to decrease linearly with antigenic distance by a factor of *d_l_(k, j)=max(0, 1 − σδ_k,j_)*, where *δ_k,j_* represents the antigenic distance between the infecting strain *k* and the measured strain *j*. The short- and long-term boost for a given strain is therefore given by *µ_l_d_l_(k, j) = max{0, µ_l_(1 − σ_l_δ_k,j_)}* and *µ_s_d_s_(k, j) = max{0, µ_s_(1 − σ_s_δ_k,j_)}* respectively. Finally, antigenic seniority by suppression was included, wherein the full amount of boosting decreased linearly by a proportion *τ* for every infection following the first. Boosting was scaled by *s(Z_i,j_, j) = max{0,(1 − τ(N_j_ − 1))}* after each infection, where *N_j_ − 1* is the number of previous infections before strain *j*. The full model, *f(**A**|**Z**,**θ**)* (where ***θ****={µ_s,_ µ_l,_ ω, σ_l,_ σ_s,_ τ}),* for expected titre for individual *i* measured at time *t* against strain *j*, *A_i,j,t_*, is given by:

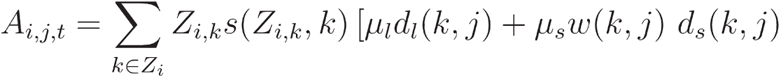

A key component of the model is the antigenic map specifying the antigenic distance between A/H3N2 strains and thus their cross-reactivity measured by the HI assay. That is, infection with strain A generates cross-reactive antibodies which also recognise epitopes on strain B, where the degree of cross-reactivity can be modeled as the antigenic distance between the two strains. In the *serosolver* model, antigenic distance between strain *k* and strain *j*, *δ_k,j_*, was given by the Euclidean distance between them on the antigenic map. In theory we might jointly estimate the antigenic map alongside the other model parameters, but at present this is computationally infeasible and thus we assume a fixed antigenic map for model fitting.

We used the antigenic coordinates of the 20 strains measured in the Fluscape study to represent the strains circulating in each time period (Fig S15). We fit a cubic smoothing spline with low amounts of smoothing (smoothing parameter = 0.3) through the coordinates to provide a comparable model to previous analyses and to smooth over large jumps in the posterior surface, which greatly aids in model convergence by smoothing over multiple posterior modes (see Supplementary Text 2) [36]. We also attempted to fit a version of the model assuming punctuated rather than continuous evolution through antigenic space, placing strains into clusters based on previous analyses of A/H3N2 strains in China [82]. Although this punctuated version of the cross-reactivity model may be more realistic [13], the posterior distribution under this model was multi-modal and thus we were unable to produce reliably converged model fits.

We assumed that log HI titres observed at time *t* were normally distributed with mean *A_i,j,t_* and variance *ε* as in [35,36], with censoring to account for the upper and lower bounds of the assay. The probability of observing an empirical titre at time *t* within the limits of a particular assay *Y_i,j,t_* ∈ *{0, …, q_max_}* given expected titre *A_i,j,t_* is given by:

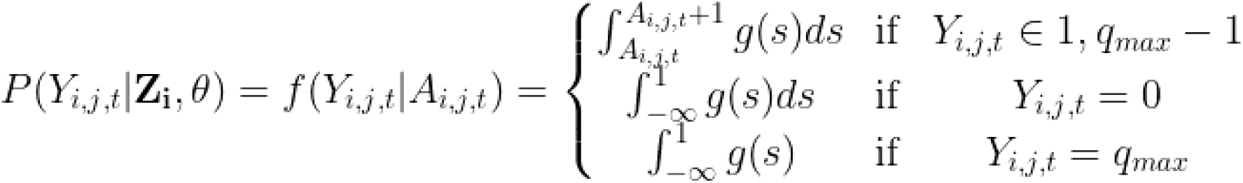

where *q_max_=8* and 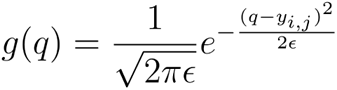, the probability density function of the normal distribution and *ε* is the standard deviation.

Fitting the model as described so far to the HI titre data led to systematic under- or over-estimation of titres to certain strains (Fig S16&S17). We therefore introduced fixed, strain-specific measurement offsets similar to [83]. The procedure for estimating these offsets is described in Supplementary Text 1. In short, we added fixed offsets to each predicted titre as:

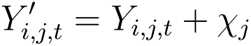

where *χ_j_* is the measurement offset for strain *j*. These additional offset parameters aim to capture the residual observation error not explained by the estimated infection histories, antibody kinetics and normally-distributed observation error.

### Infection history prior

The choice of prior for the infection history matrix ***Z*** is discussed in detail in [35]. The crux of the problem is that the choice of prior *P(**Z)*** determines not only *P(Z_i,j_=1)*, but also the prior distribution of total number of lifetime infections, attack rate in a given time period, and time between infections. As we are interested here in reconstructing historical attack rates, we chose to place a Beta prior on the probability of infection in a given time window (prior version 2 in *serosolver*). The infection history matrix ***Z*** is then a Beta-Bernoulli distributed variable such that:

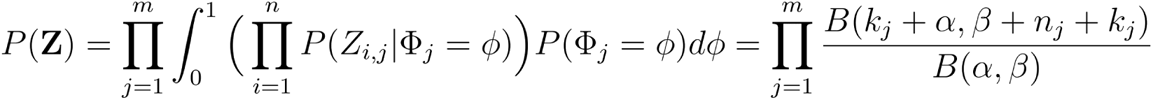

where *B* is the Beta function; 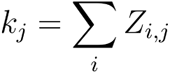 is the total number of infections across all individuals during time period *j*; and *n_j_* is the number of individuals that could be infected during time period *j*. Values for *α* and *β* can then be set to give known priors and variance on the total number of infections as 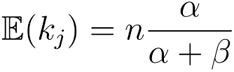 and 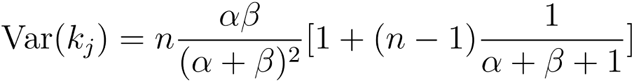. When *α=β*, the attack rate prior has an expectation of *0.5n*, and the variance may be decreased by increasing *α* and *β*. Here, we set *α=β=1*. This choice of prior also implicitly assumes that the total number of lifetime infections for an individual is binomially distributed with success probability *p=α/(α+β)* and *N=m_i_, where m_i_* is the number of time periods that individual *i* could be infected.

### Inference using Markov chain Monte Carlo

All models were fitted using the Markov chain Monte Carlo (MCMC) algorithm implemented in the *serosolver* R package. This is a custom, adaptive Metropolis-Hastings algorithm with alternating univariate normal proposals for the model parameters ***θ*** and custom proposals ***Z*** for the infection history states. Step sizes for all antibody kinetics parameter proposals were also scaled automatically during the burn in to achieve an acceptance rate of 0.44 for all parameters. Uniform priors were placed on all antibody kinetics parameters ***θ*** shown in Table S4. Infection history state proposals were randomly chosen between one of two options: (i) select two potential infection times 12 time periods apart and swap infection states for all individuals that could have been infected at these times; (ii) randomly select 20% of individuals and for each individual, with 50% probability, either sample new infection state values for all possible infection times, or choose two potential infection times 12 time units apart and swap their values.

For the main model results, 5 chains were run for 50,000,000 iterations, with the first 20,000,000 discarded as burn-in, to achieve an effective sample size of >200 for all inferred parameters. Convergence was assessed visually and using the Gelman-Rubin diagnostic criteria (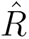) with the *coda* R-package [84]. Note that the huge number of iterations is due to the need to impute nearly *4*N*T = 4*1130*190 = 858,800* latent binary variables in ***Z*** (i.e., the infection state for each individual in each 3-month window since 1968, slightly less as individuals could not be infected prior to birth). The ability for the model to recover ground-truth parameters was explored through simulation-recovery, described in Supplementary Text 2.

### Post-processing of infection history posteriors

When fitting to the Fluscape dataset, the *serosolver* model occasionally imputed continuous runs of repeated infections in adjacent 3-month windows (e.g., imputing infections in Q1-1968, Q2-1968 and Q3-1968), reflecting either genuine repeat infections or the model explaining titres that were higher than a single antibody boost could explain. These infection runs were relatively rare, but were most common for infection in young children and occasionally for recent time periods (Fig S18&S19). Runs of repeat infections were not estimated when fitting to simulated data, suggesting that their occurrence in the Fluscape data represents either genuine repeat infections early in life or the model explaining titres that were higher than a single antibody boost could explain. Although these reinfections may be real, particularly amongst children [85,86], this may be a limitation of the model, which sometimes explains high antibody titres through multiple infections rather than from a single infection eliciting a large boost. Our pipeline therefore included a post-processing step to count these runs as single infection events starting on the date of the first infection in the run. This reduced the total number of distinct infection events from 11,544 infections (posterior median; 95% CrI: 11,358-11,736) to 10,558 (posterior median; 95% CrI: 10,394-10,750). The attack rate estimates and reinfection rates from the posterior samples prior to removing the runs of consecutive infections are shown in Table S2, showing that although the post-processing step only slightly reduced overall attack rate estimates, it substantially reduced the reinfection rate estimates, which were as high as 8% in 2013 prior to removing consecutive infections.

### Fitting to longitudinal HI titre data from Ha Nam, Viet Nam

To provide a comparison for the model fits to the Fluscape data, we fit the same model to a publicly available dataset from Ha Nam, Viet Nam. This dataset consisted of 69 participants, each with between 1 and 6 (inclusive) serum samples taken annually from 2007-2012 as described previously [37]. In this cohort, HI assays were performed against a panel of up to 57 A/H3N2 strains isolated between 1968 and 2008, with greater sampling of titres against more recent strains. This represents a comparable dataset with different dimensions: a much smaller sample size, but far more titres and samples tested per individual. Unlike the Fluscape data, we only had access to the year of sample collection, and thus we could only estimate infection histories at an annual resolution. Furthermore, we did not have dates of birth available for the individuals, and thus we assumed that all individuals were born prior to 1968, noting that age signals will still be detected based on the individual’s antibody profile. For this analysis, we did not include the strain-specific measurement offsets in the observation model, as there were multiple strains tested for each time period. We ran the Markov chain Monte Carlo algorithm for 5 chains each for 1,500,000 iterations and discarded the first 500,000 iterations as burn-in, which was sufficient to achieve effective sample sizes of >200 and upper 95% confidence intervals of the 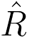 values of less than 1.1 for all estimated parameters.

### Spatial correlation in inferred attack rates

To test for patterns of influenza incidence in space, nonparametric correlation as a function of distance was tested using the *Sncf* function from the *ncf* R-package [87], where observations were the quarterly attack rate estimates stratified by location ID (40 locations, 190 observations per location). We fit these spline correlograms to 100 samples from the posterior distribution of attack rates, resampling the 40 locations with replacement for each sample to generate posterior medians and 95% credible intervals (CrI). This analysis was repeated using estimated attack rates since 1968, since 2009, and pre-2009. To provide a comparable null simulation where no significant spatial variation would be expected, we simulated 40 draws from a binomial distribution with n=25 (i.e., 1,000 individuals across 40 locations) and success probability drawn from a uniform distribution between 0 and 1, and repeated this process 10,000 times to calculate the mean and 95% quantiles of the resulting coefficients of variation.

### Relationship between titre and probability of infection

For the analyses investigating the relationship between titre at time of infection and probability of infection, we drew samples from the estimated posterior distribution of antibody kinetics parameters and infection histories, and calculated model-predicted pre-infection latent antibody titres against the circulating strain for each individual at each possible infection time. Predicted titres were converted to integers, with titres ≥8 assumed to be 8 to match the observation process. We then calculated the proportion of time periods where infection occurred (*Z_i,j_ = 1*) (the overall probability of infection) stratified by log titre at the time of infection relative to the overall probability of infection with a log titre of 0. As discussed above, some of the inferred infection histories had runs of infections in consecutive time periods (e.g., ***Z_i_****=*[0, 0, 0, 1, 1, 1, 0, 0]). We removed these consecutive infections from the probability of infection statistics (e.g., [0, 0, 0, 1, 1, 1, 0, 0] becomes [0, 0, 0, 1, 0, 0, 0, 0]). We repeated this process for 1000 posterior samples to generate median and 95% credible interval estimates on the relationship between titre and relative risk of infection.

## Data Availability

All code and data required to reproduce the analyses are available online at https://github.com/jameshay218/fluscape_infection_histories

## Supplementary text, figures and tables

**Figure S1:**
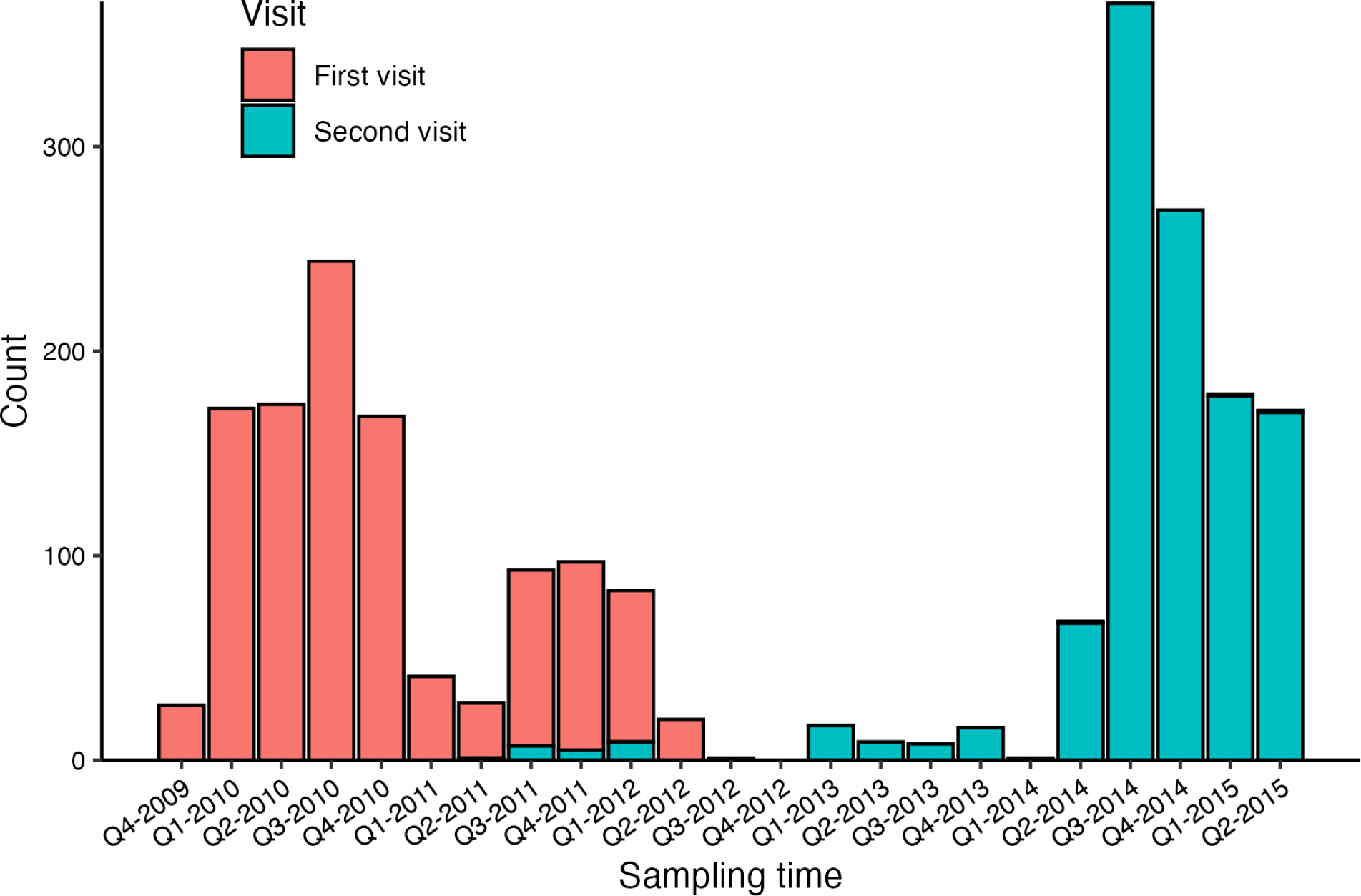
Distribution of serum sampling times from the Fluscape cohort. First and second visits refer to an individual’s serum sample order, which may differ from the sample collection round of the overall study.

**Figure S2:**
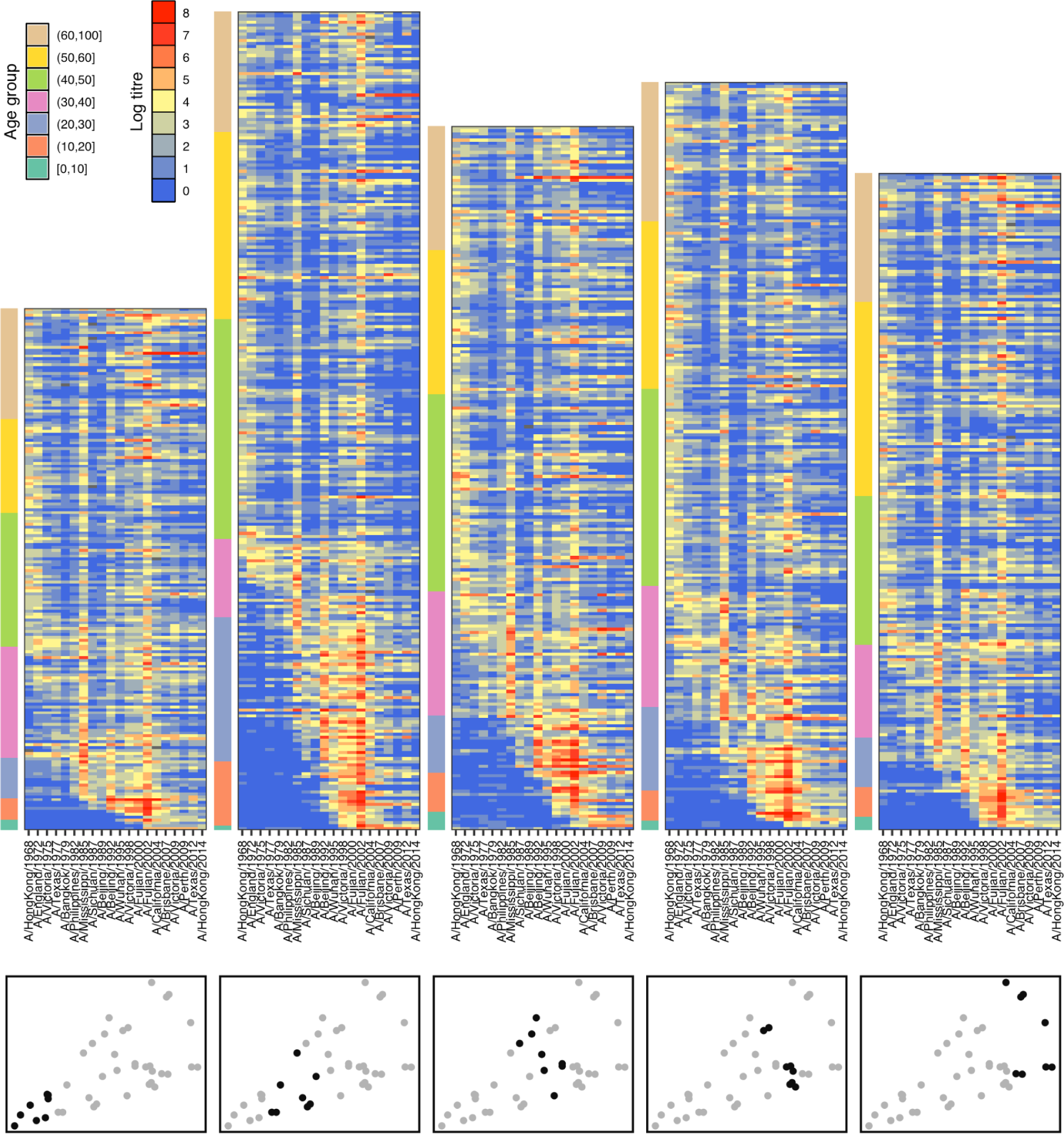
Distribution of log HI titres by study location at first serum sample. Each cell represents the log HI titre for one individual measured against one strain, shown on the x-axis. Locations were grouped into quintiles based on increasing distance from Guangzhou city center (bottom panels). Individuals were grouped by age and plotted with increasing age. Colors to the left of each subplot show age group.

**Figure S3:**
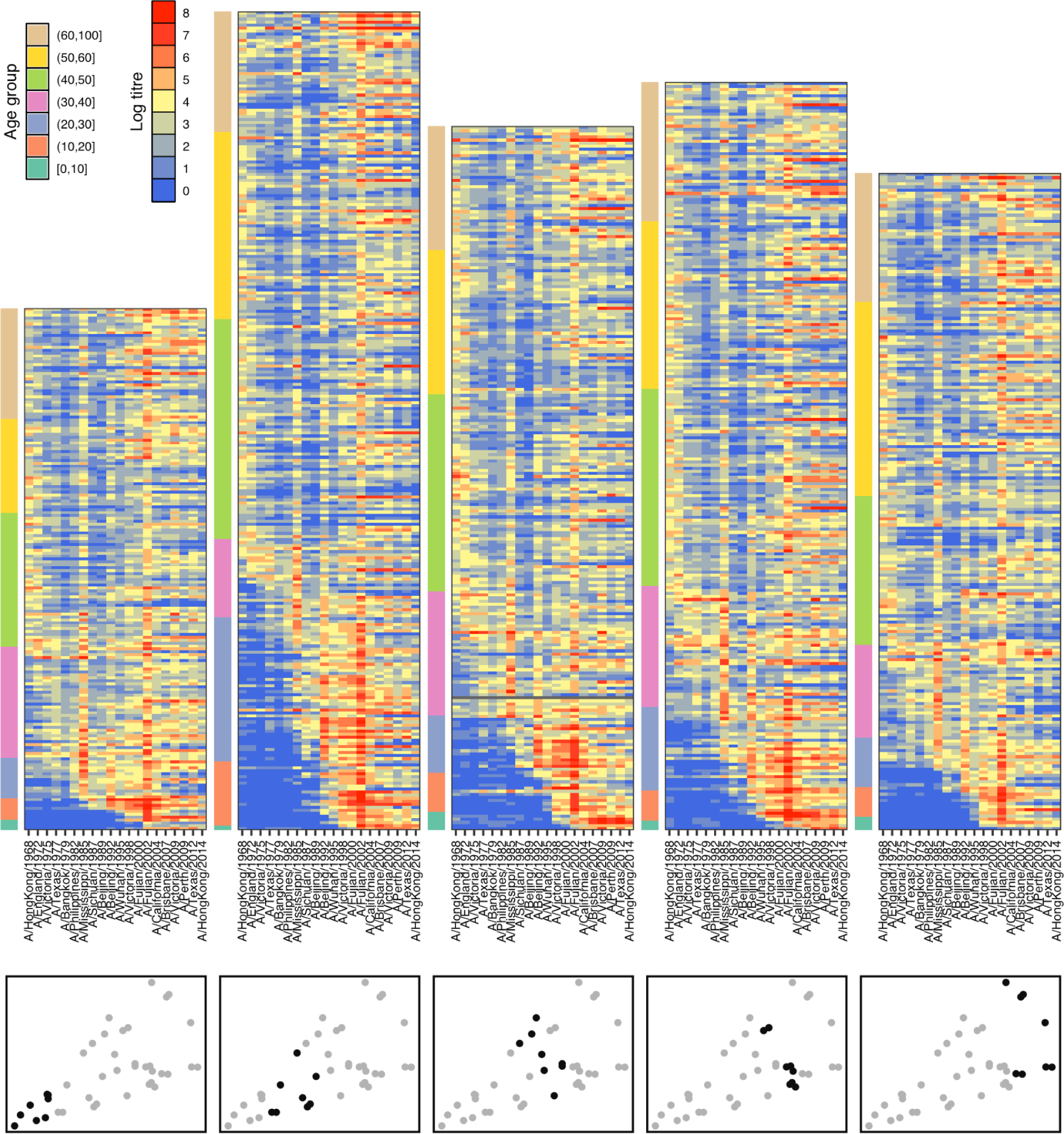
Distribution of log HI titres by study location at second serum sample. Each cell represents the log HI titre for one individual measured against one strain, shown on the x-axis. Locations were grouped into quintiles based on increasing distance from Guangzhou city center (bottom panels). Individuals were grouped by age and plotted with increasing age. Colors to the left of each subplot show age group.

**Figure S4:**
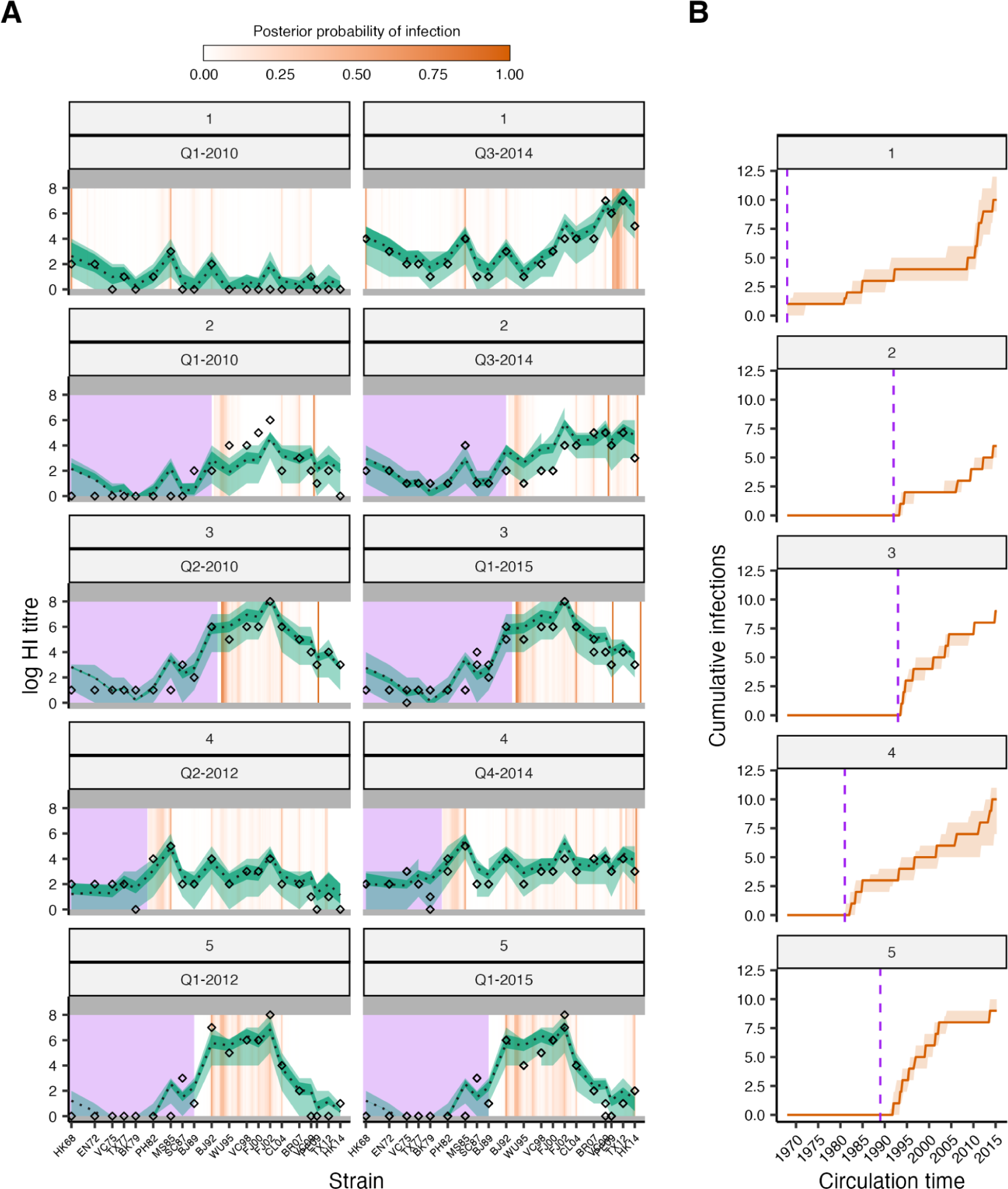
Example inferred latent antibody titres and infection histories. (**A**) Model-predicted titres compared to observed HI titres at each sampling time for 5 randomly selected individuals. Rows represent individuals. Subplots show antibody titres based on serum samples taken at that time. X-axis represents a position along the antigenic summary path. Black diamonds show observed titres. Black line and green shaded regions show posterior median and 95% credible intervals (CrI) on model-predicted latent titres (dark green) and 95% prediction intervals (light green). Orange bars show posterior probability of infection in that 3-month window. Gray rectangles denote the limit of detection of the HI assay. Purple rectangles show time periods before birth. (**B**) Posterior median and 95% CrI for the cumulative number of infections over time from birth (purple dashed line).

**Figure S5:**
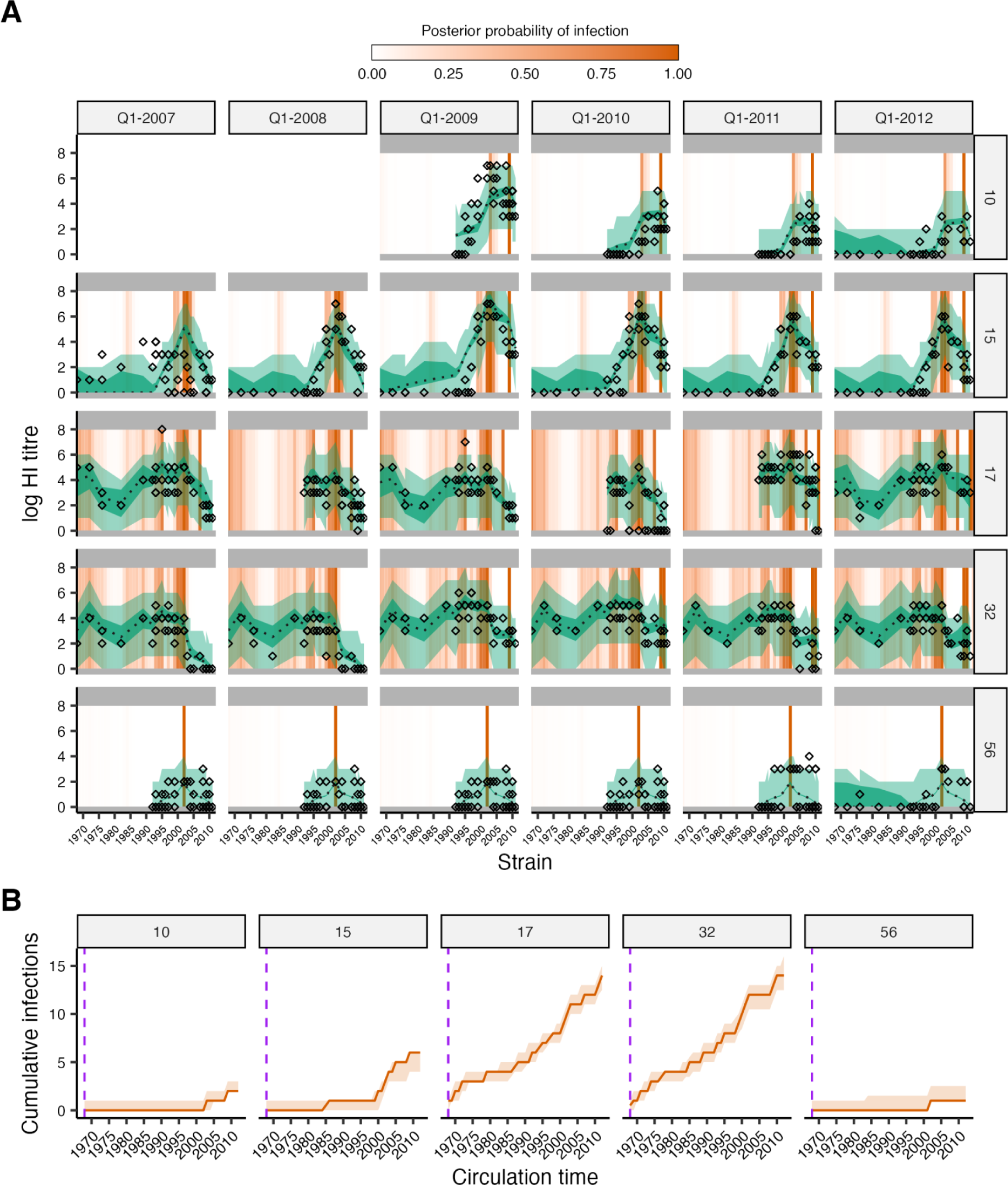
Model fit to data from Ha Nam, Viet Nam. (**A**) Model-predicted titres compared to observed HI titres at each sampling time for five randomly selected individuals, as in Fig S4. Diamonds show titre measurements; green shaded region shows 95% CrI and 95% prediction intervals; dashed line shows posterior median; orange bars show posterior probability of infection in a given time window. (**B**) Posterior median and 95% credible intervals (CrI) for the cumulative number of infections over time from birth (orange). Note that date of birth information was not available for these individuals.

**Figure S6:**
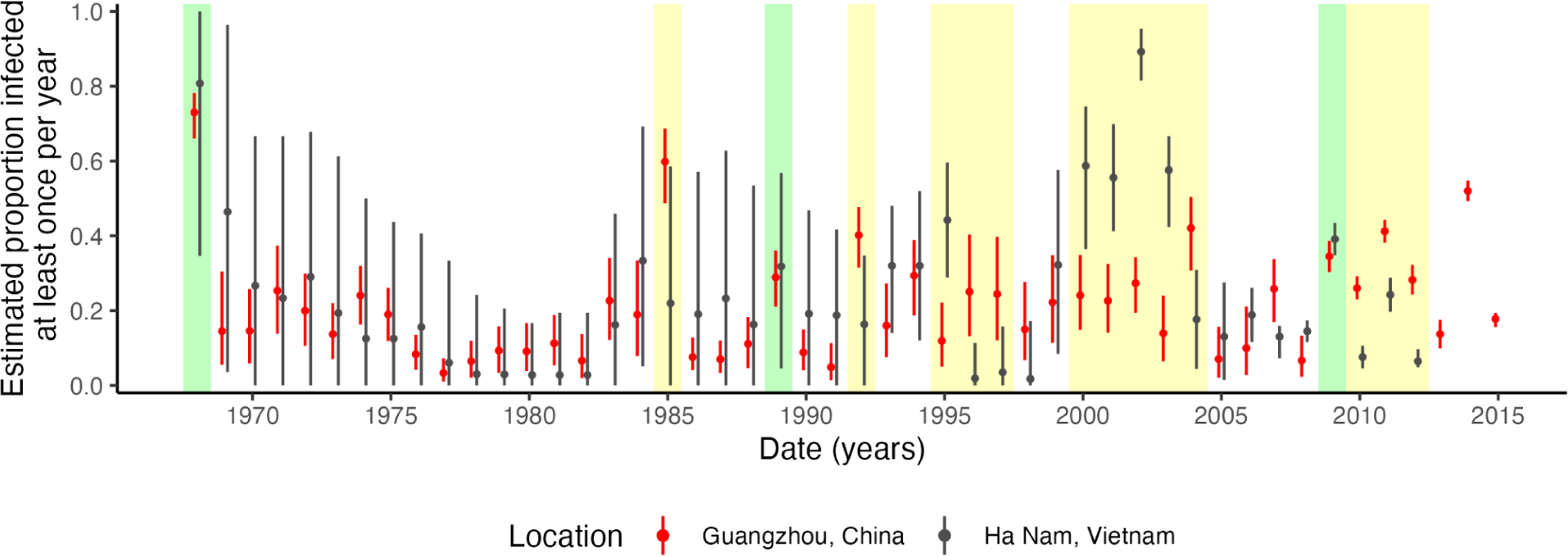
Comparison of annual attack rates using data from Ha Nam, Viet Nam and the Fluscape study in Guangzhou, China. Annual attack rates were defined here as the proportion of individuals who experienced at least one infection per year. Green shaded regions show notable time periods where the two datasets return similar attack rate estimates, whereas yellow shaded regions show time periods with notably different attack rate estimates. Attack rates in 1968, 1989 and 2009 were remarkably similar between the two locations. Some time periods showed high uncertainty for the Ha Nam dataset, as few individuals in the sample were alive during that time (e.g., 1969-1980). Attack rate estimates were very different from 2000 to 2004, with much higher attack rates estimated for the Ha Nam cohort. This might reflect a genuine different in A/H3N2 epidemiology during that time, but may also be partially driven by systematic biases in titre measurements to strains isolated during that time period – the fits to the Fluscape data include a positive offset term for titres against A/Fujian/2002, which leads to lower attack rate estimates in that time period, whereas fits to the Ha Nam data do not. The time period from 2010-2012 is also interesting, showing similar relative patterns but much higher attack rate estimates overall in the Fluscape cohort.

**Figure S7:**
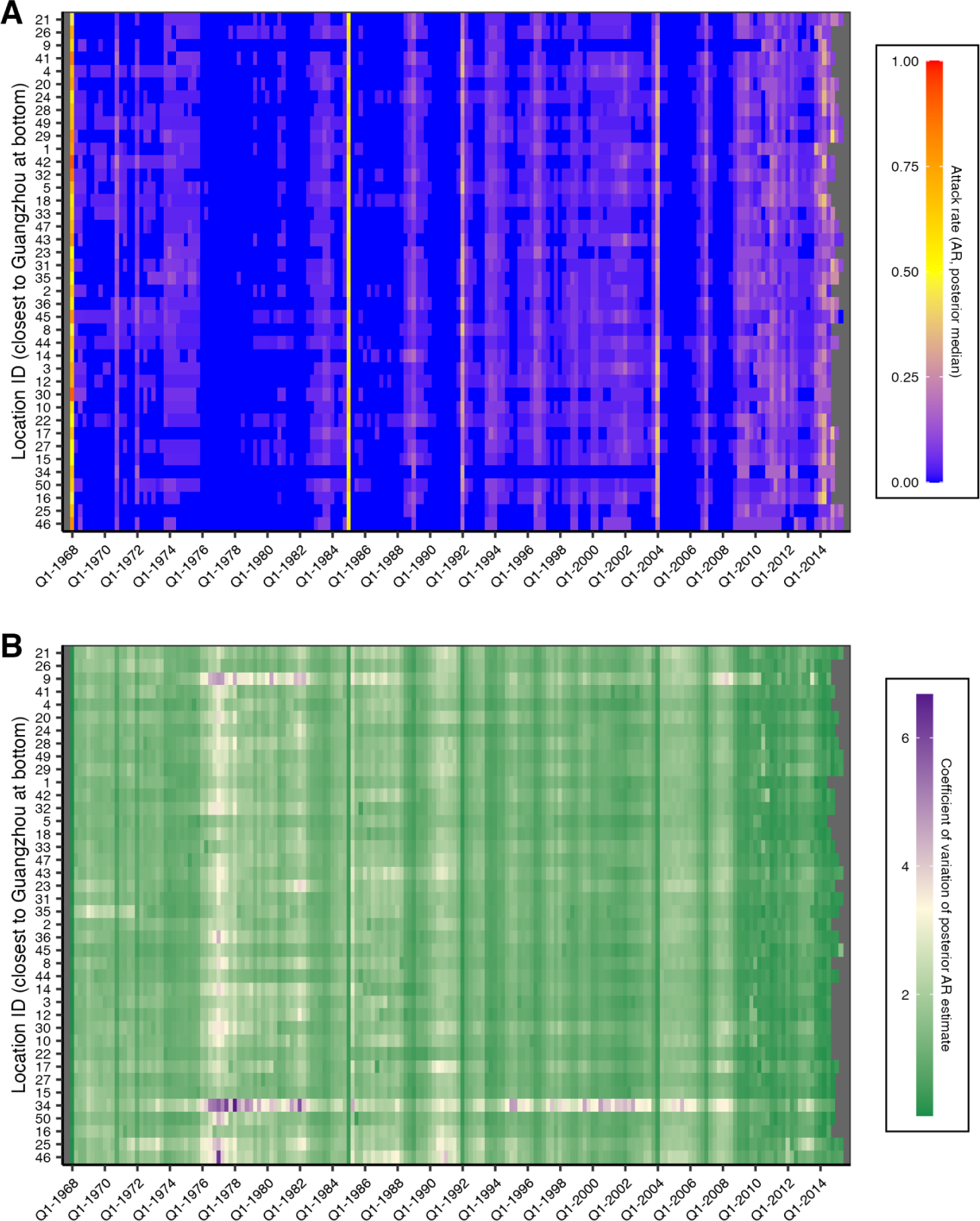
Quarterly attack rates across the 40 Fluscape study locations. Each row represents one study location ordered by increasing distance from Guangzhou city center. Each column represents a 3 month period. Cells are shaded by (**A**) the posterior median inferred attack rate or (**B**) the coefficient of variation of the posterior quarterly attack rate estimate for each location.

**Figure S8:**
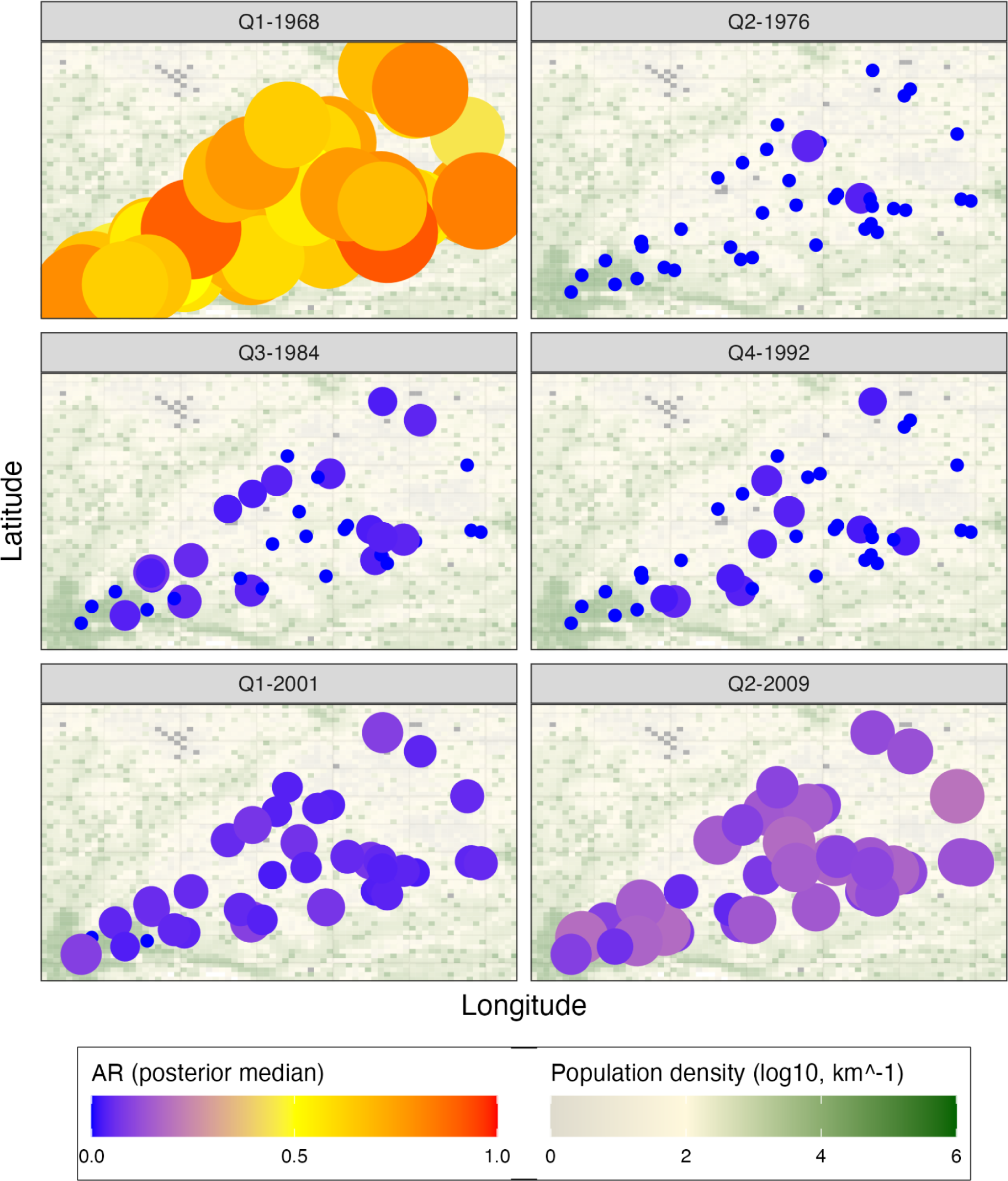
Distribution of quarterly attack rates by location over time. Each panel is one frame from a full animation available in Supplementary Material 3. Each colored point shows the inferred attack rate in each of the 40 locations, with size and shading reflecting the posterior median attack rate. Underlying the plot is a map of the study area, with each grid cell shaded by its *log_10_* population density.

**Figure S9:**
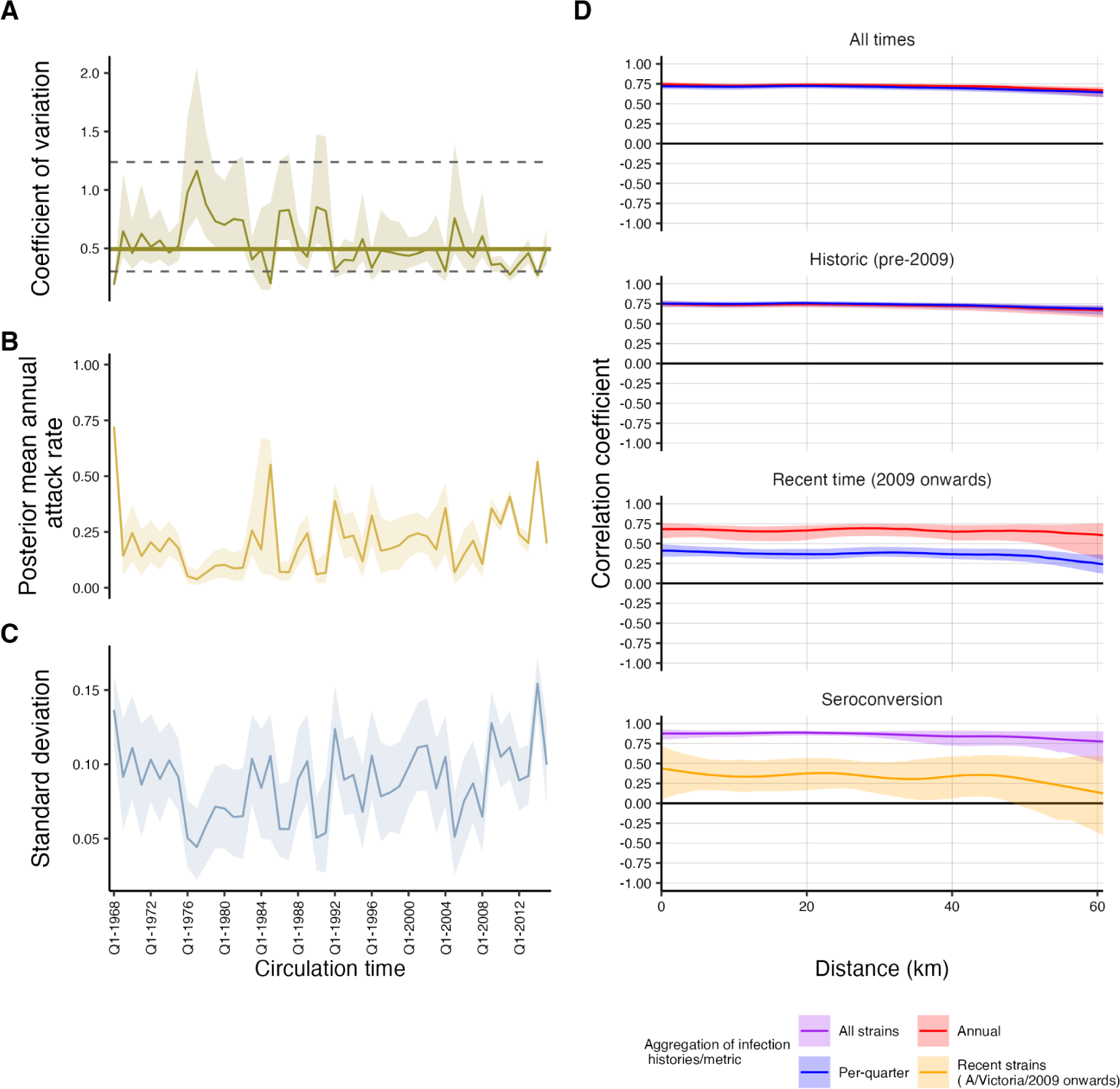
Spatial variation in annual attack rate estimates over time and correlation between nearby locations. **(A)** Coefficient of variation, (**B**) overall mean and (**C**) standard deviation of posterior median annual attack rate estimates from the 40 study locations. Solid lines and shaded regions show posterior medians and 95% credible intervals. In (**A**), the solid horizontal line shows the overall mean coefficient of variation across all time. Dashed horizontal lines show 95% quantiles of simulated coefficients of variation under the assumption that attack rates are the same across space. (**D**) Fitted spline correlograms showing spatio-temporal correlation in attack rates and proportion seroconverted with increasing distance. The first three plots show the spline correlogram calculated using the *Scnf* function from the *ncf* R-package for each of 100 posterior samples for the 40 location-specific attack rates. Solid lines and shaded regions show median and 95% quantiles of the predicted covariance function for these 100 samples, coloured by the level of aggregation used to calculate the attack rates. Each subplot shows the same calculation using either attack rates from all times, prior to Q1-2009 or Q1-2009 onwards. For the final plot (“Seroconversion”), we calculated the spatial correlation in the proportion seroconverted in each of the 40 study locations, treating strain isolation time as the time variable. Solid lines and shaded regions show median and 95% quantiles of 1000 bootstrapped observations. We repeated the analysis using either seroconversion to all strains, or only A/Victoria/2009, A/Perth/2009, A/Texas/2012 and A/HongKong/2014.

**Figure S10:**
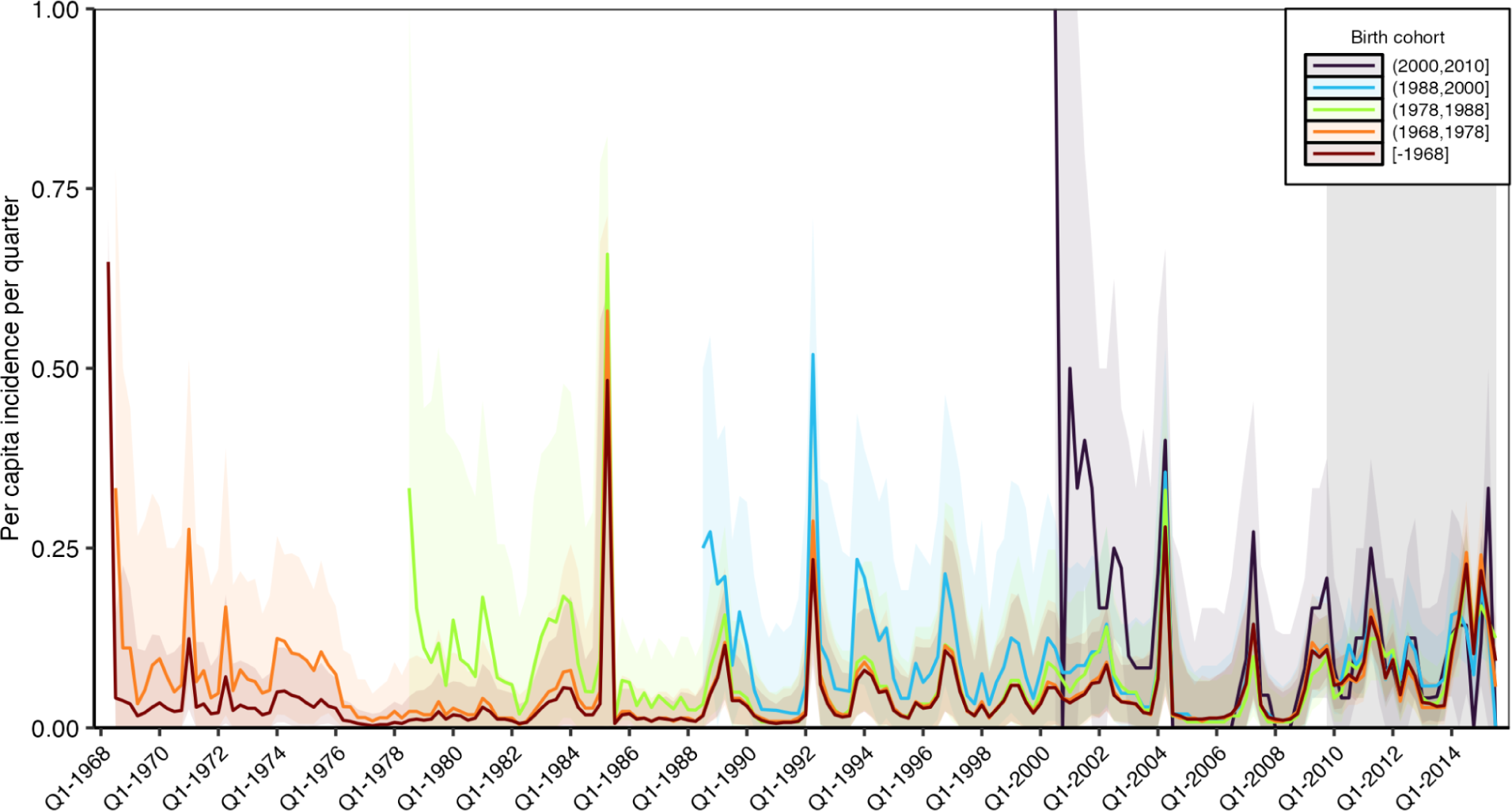
Quarterly attack rates by birth cohort. Model predicted per-capita incidence per quarter stratified into 5 birth cohorts. Attack rates were estimated by dividing the number of inferred infections by the number alive in each birth cohort in each 3 month period. Solid lines show the posterior median estimate from 1000 posterior samples. Shaded regions show 95% credible intervals from 1000 posterior samples. Gray shaded box shows duration of the Fluscape study.

**Figure S11:**
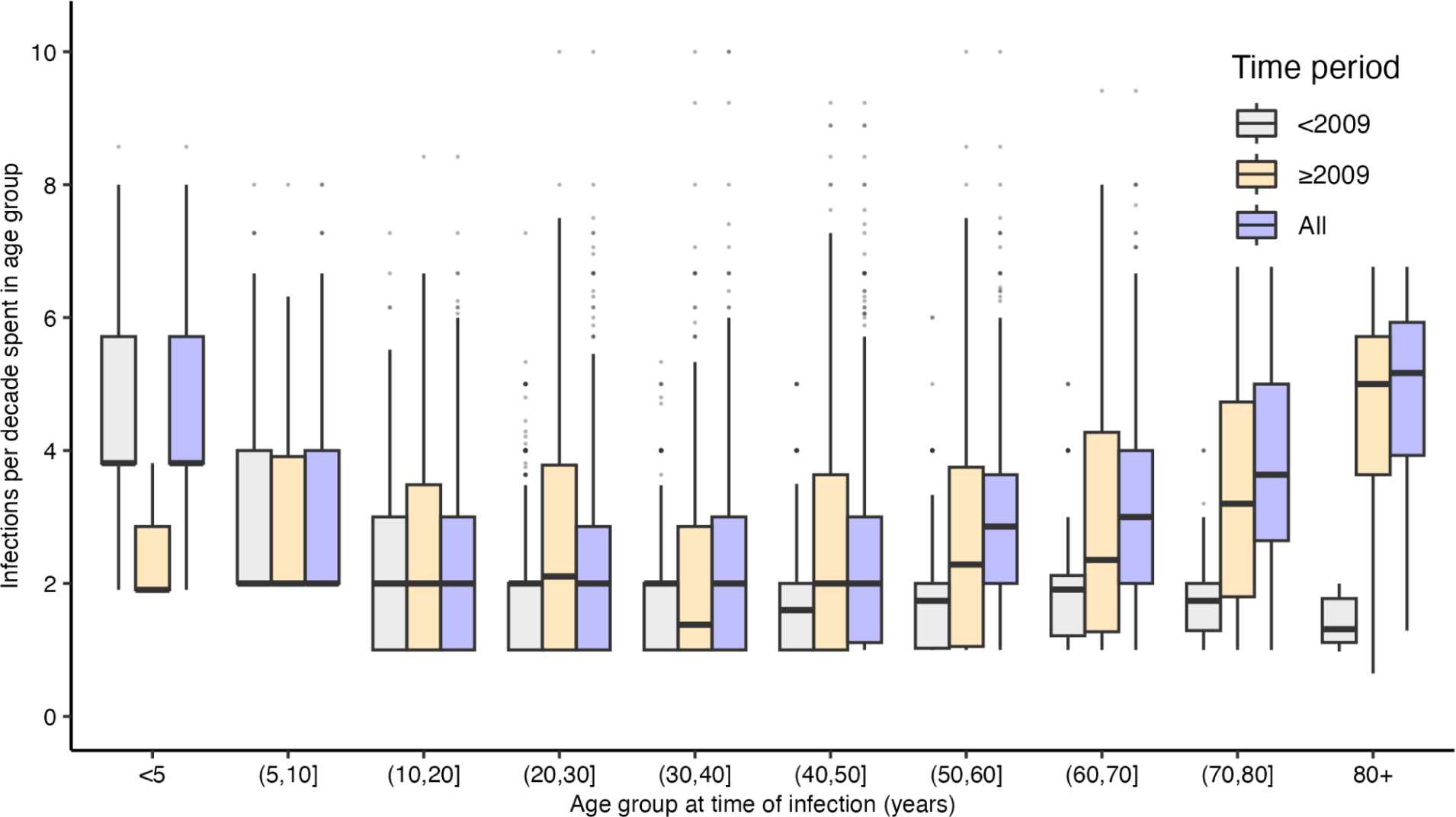
Posterior median number of infections per 10 year period stratified by age group at the time of infection, including either all time periods (blue), time periods during the Fluscape study period (Q4-2009 onward; orange) or only time periods prior to the Fluscape study (prior to Q4-2009; grey). Excludes infection states for individuals who spent less than 2 years in that age group. We present infection rate estimates using only infections from time periods prior to the first serum sample in Q4-2009 in Figure 3C. This is because there are many individuals representing the oldest age group at time of infection for time periods post Q4-2009, but relatively few from pre Q4-2009 (as individuals who were very old in historical time periods are no longer alive). In contrast, younger age groups are better represented across historical time periods (as those individuals are still alive at the time of sampling). Combining this biased representation of older individuals with much higher estimated incidence rates in recent time periods weighs the infection rate estimates for the older age groups much higher simply because most of their infections come from this time period. Therefore, we present age-stratified infection rate estimates using only pre Q4-2009 infections in the main text.

**Figure S12:**
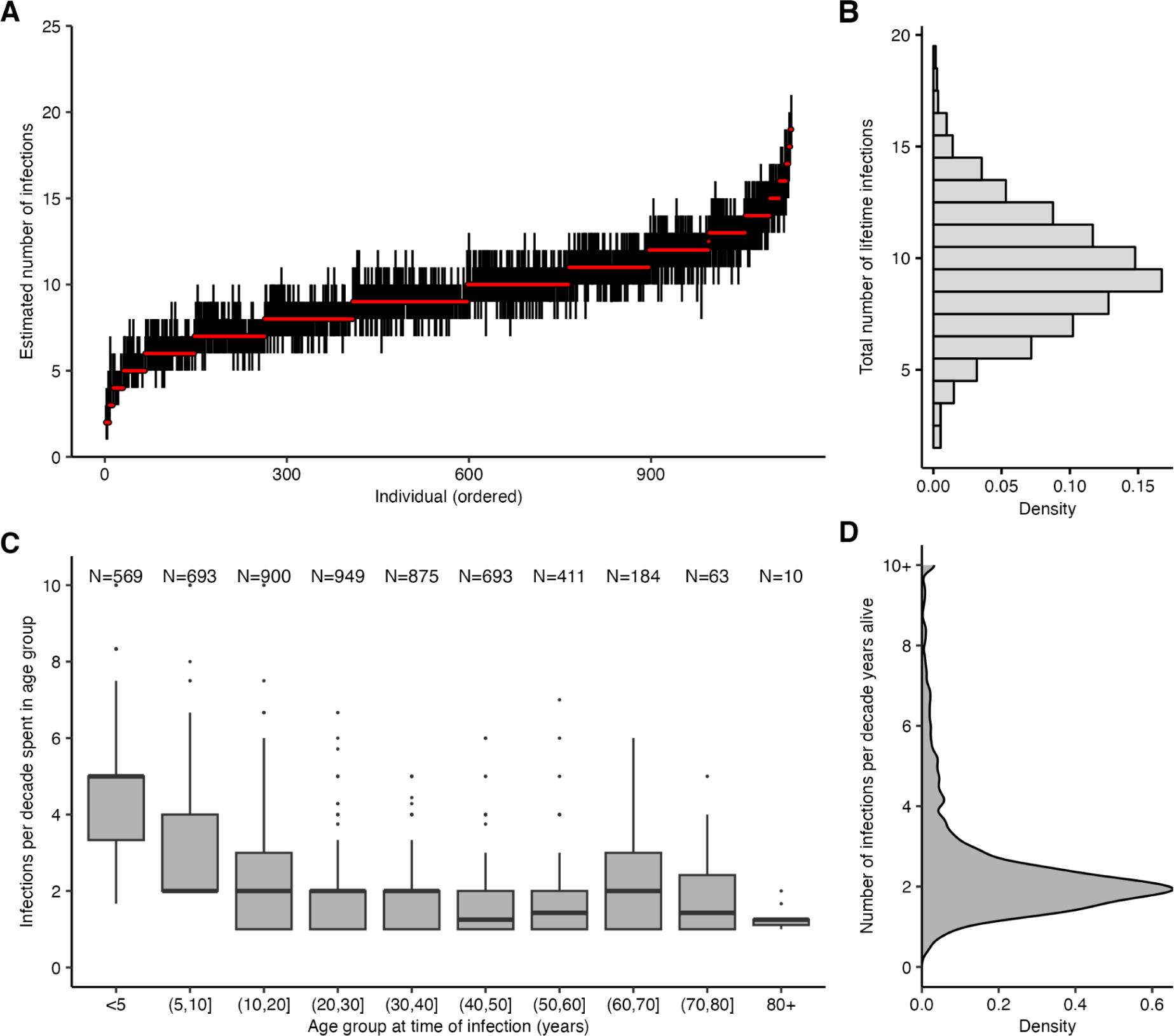
Age-specific patterns of infection under an alternative infection history model using estimates from the model version described in Supplementary Text 1. Results shown are identical to those in Figure 3, but assuming that (i) individuals can only be infected once per year (i.e., annual resolution infection histories rather than quarterly); (ii) the infection history model is placed upon an individual’s total number of lifetime infections and not their per-time probability of infection (see [35] for further detail on implications of different prior assumptions); (iii) we did not remove runs of continuous infections from the posteriors. (**A**) Pointrange plot shows median and 95% CrI on the total number of lifetime infections for each individual, ordered by increasing age. (**B**) Distribution of the total number of infections across all individuals based on the posterior median total number of infections. (**C**) Posterior median number of infections per 10 year period stratified by age group at the time of infection, excluding individuals who spent less than 2 years in that age group, and including only time periods prior to Q4-2009. Text shows sample size within each age group – note this does not sum to the number of individuals in the sample, as individuals contribute to multiple age groups during their lifetime. (**D**) Posterior median number of infections per 10 years alive across all individuals. Under this prior, we estimated that individuals are infected 2.08 times per 10 year period (posterior median; 95% CrI: 1.04-7.62).

**Figure S13:**
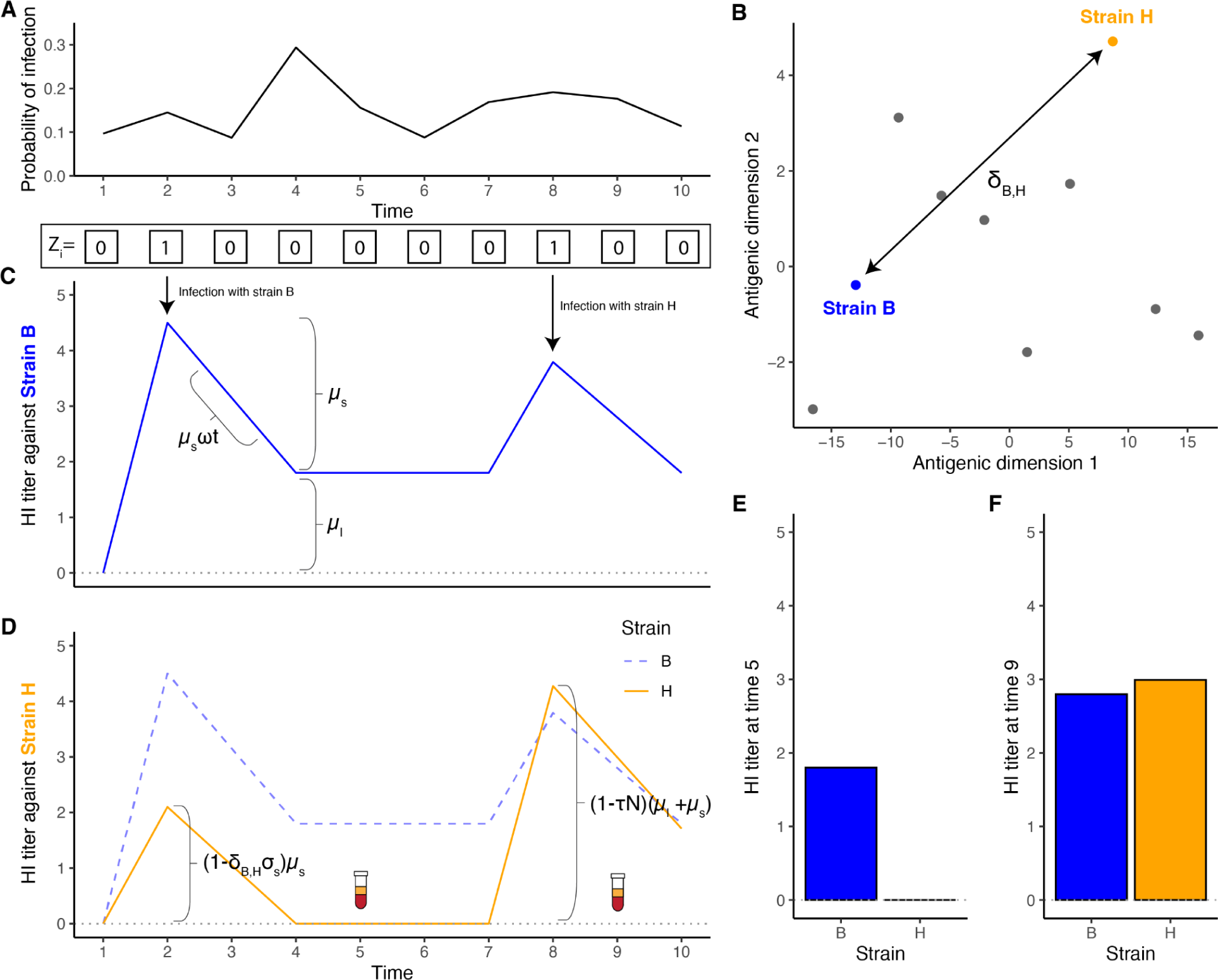
Schematic of the full *serosolver* model representing a single individual infected with two strains, B and H, over a 10 year time period. (**A**) Example, randomly generated population-level infection probabilities. At the population level, the model describes a per-time-period probability of infection applied to the whole whole population. These probabilities are used to simulate a vector of latent binary infection states, *Z_i_*, for each individual as a series of independent Bernoulli trials (shown as a vector of 1s and 0s). (**B**) The antigenic relatedness of A/H3N2 strains is given by an antigenic map, where the degree of cross reactivity between any two strains is given by their euclidean distance on the map. (**C**) Antibody levels against the infecting strain (strain B) are boosted and wane, given by the summation of transient short-term boosting and persistent long-term boosting. (**D**) Infection with strain B also induces cross-reactive antibodies against all other strains, here showing antibody levels to strain H. The degree of cross-reactivity is proportional to the antigenic distance between the infecting and measured strain. Later on, the individual is infected again, this time with strain H, inducing further antibody boosting and waning. An antigenic seniority parameter, τ, reduces each successive boost as a function of the number of previous infections, N. Snapshots of these underlying antibody kinetics are observed through serum samples (blood vials; (**E**) and (**F**)) distributed according to a truncated, discretized normal distribution with standard deviation parameter ε.

**Figure S14:**
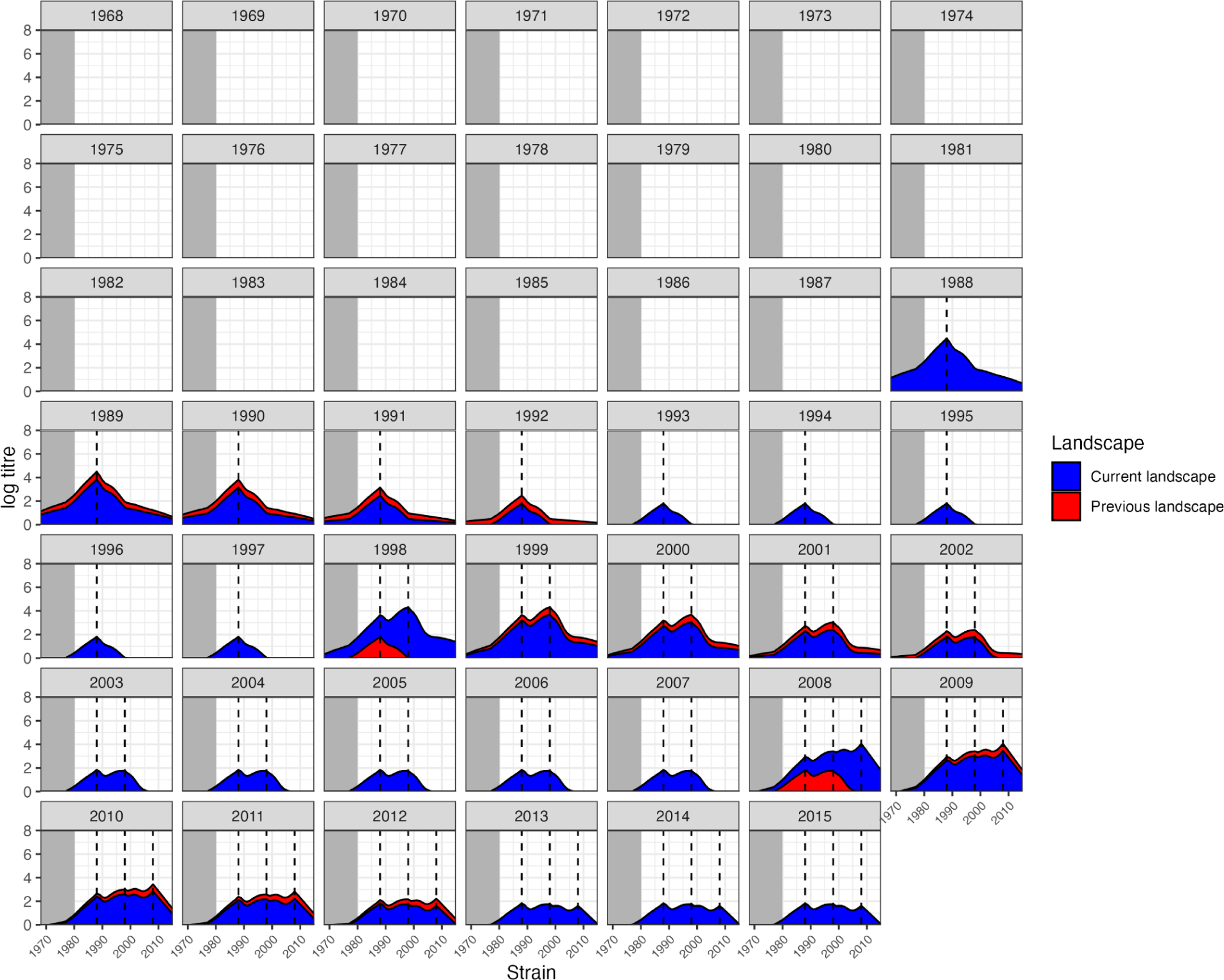
Simulated antibody landscapes and infection histories over time for one individual using the antibody kinetics model. Each subplot shows the antibody landscape for that time period. The blue region gives the antibody landscape in that time period, whereas the red region gives the antibody landscape in the preceding time period. The x-axis of each subplot gives the identity of the strain assumed to be circulating in that year. The grey region shows the time period and thus strains that circulated before the individual was born. Vertical dashed lines give the timing/strains the individual was infected with.

**Figure S15:**
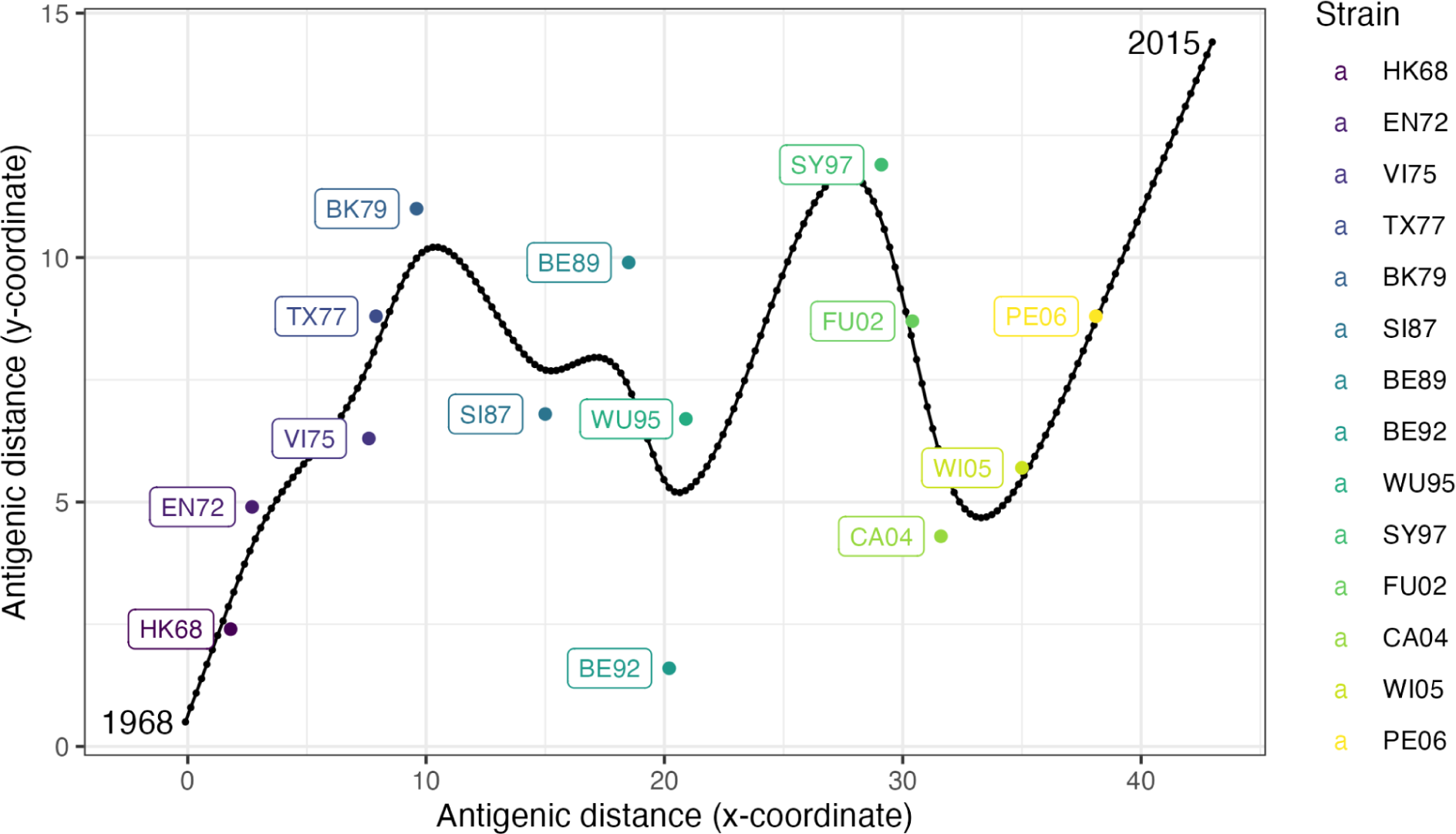
Antigenic coordinates of measured strains and strains assumed to be circulating in each time period. Each colored point shows the location of the labeled strain on the antigenic map given in [37]. Strains which are further apart are less antigenically similar, and therefore exhibit less cross-reactivity following a seroresponse. The smoothing spline shows the inferred coordinates of each strain *j* assumed to have circulated in each 3-month time period, where each black point shows the assumed location in successive time periods. First, a cubic smoothing spline was fitted to the locations of the measured strain with smoothing parameter 0.3. Second, a linear model was fitted to predict the x-coordinate as a function of the strain isolation time. Finally, we generated predicted x-coordinates for each possible *j* given the circulation time from the linear model, and then used the predicted x-coordinate to predict the y-coordinate from the fitted smoothing spline. The antigenic distance between each pair of strains *k* and *j* was then calculated based on their Euclidean distance.

**Figure S16:**
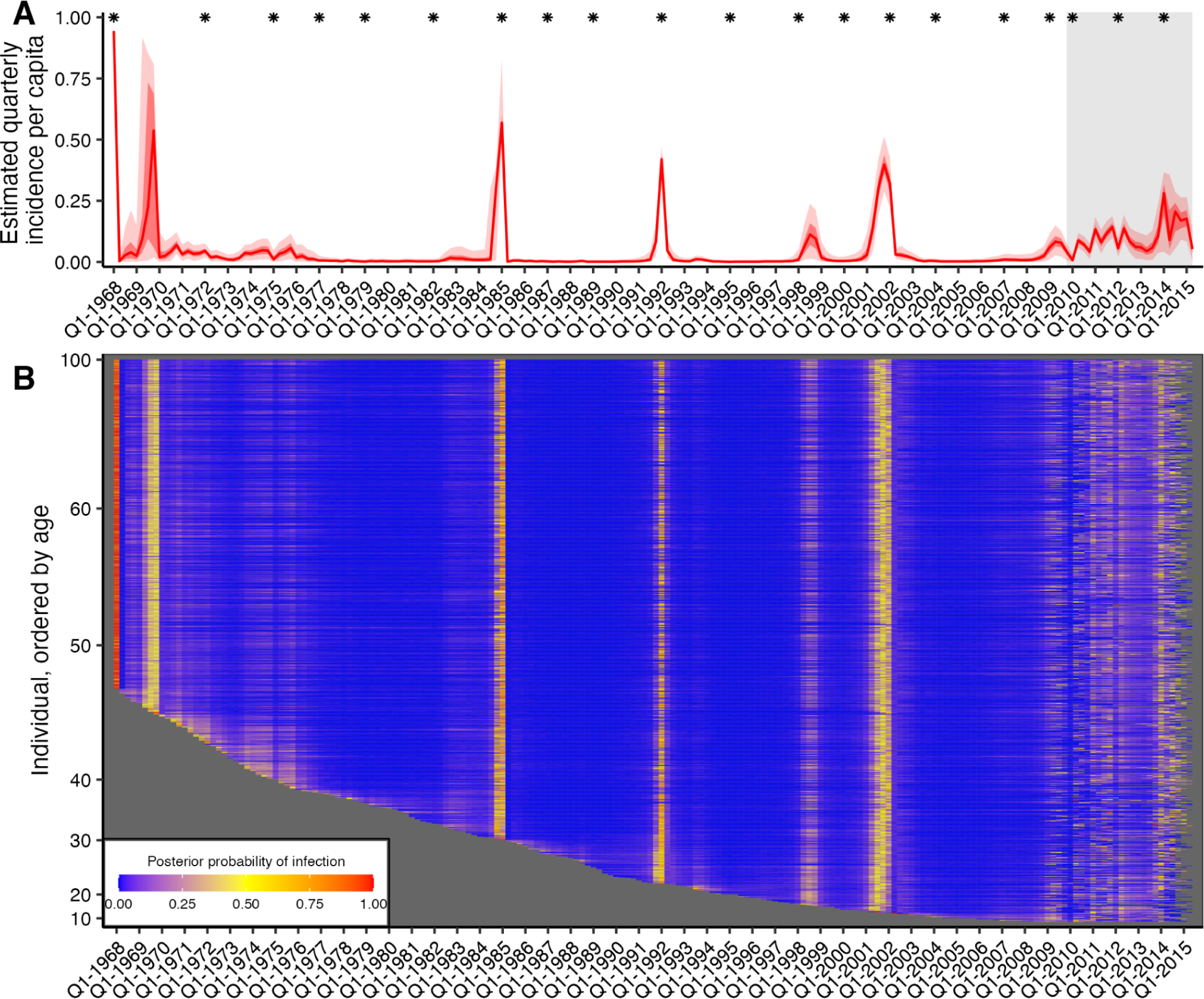
Quarterly incidence and individual infection histories from the Fluscape dataset without strain-specific measurement offsets. Identical to Figure 2, but without the inclusion of strain-specific measurement offsets in the observation model. (**A**) Model predicted per-capita incidence per quarter. Attack rates were estimated by dividing the number of inferred infections by the number alive in each 3 month period. Red line shows the posterior median estimate from 1000 posterior samples. Dark and light red shaded regions show 50% and 95% credible intervals respectively from 1000 posterior samples. Gray shaded box shows duration of the Fluscape study. Asterisks mark times from which a sample circulating strain was tested. (**B**) Inferred infection histories for each individual. Each row represents an individual ordered by increasing age in years. Each column represents the time of a potential infection. Cells are shaded based on the number of the posterior samples with an infection at that time divided by the total number of posterior samples for that infection state.

**Figure S17:**
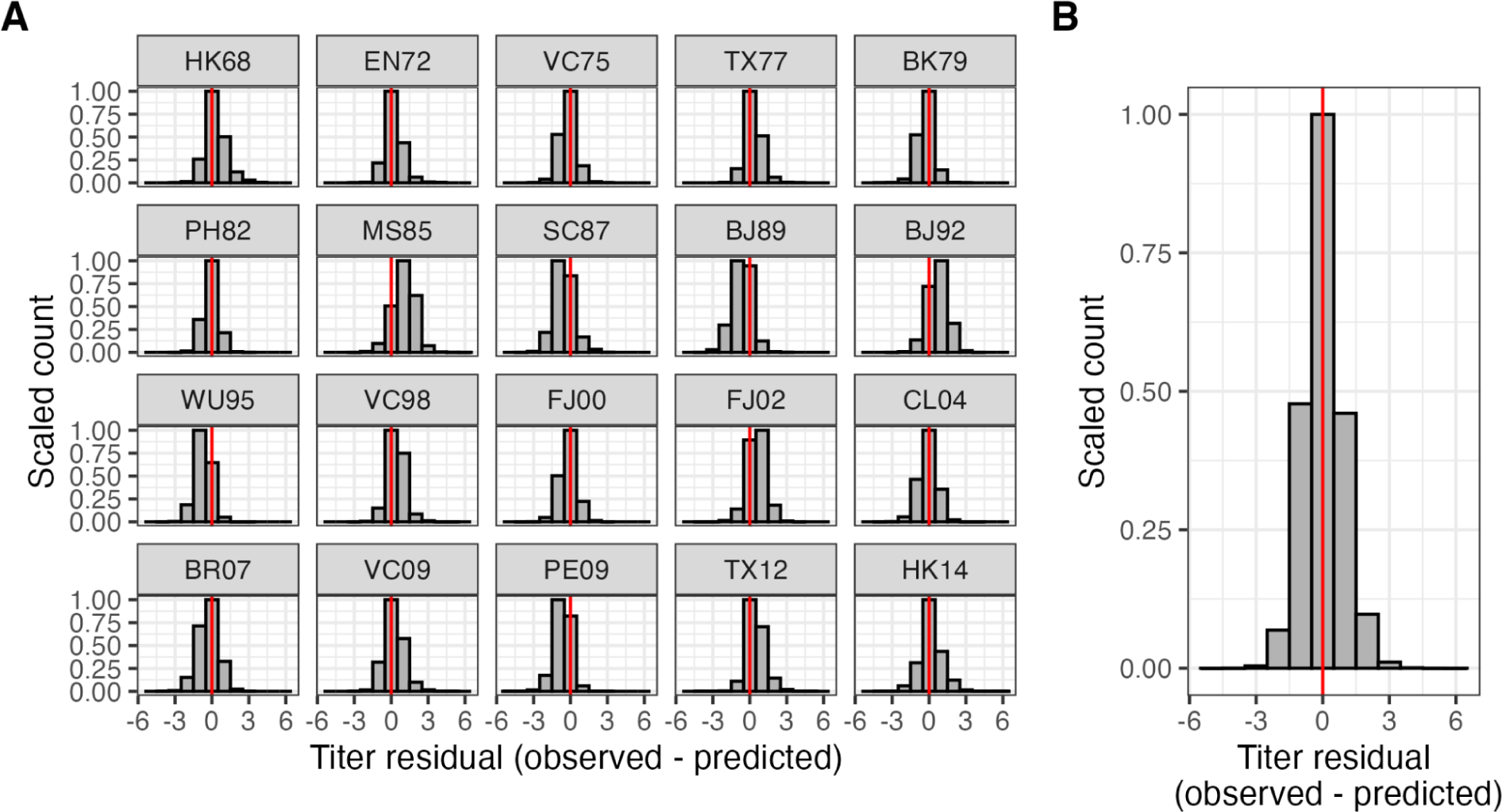
Distribution of antibody titre prediction errors (observed - model predicted) when fitting the *serosolver* model ignoring strain-specific measurement offsets. (**A**) Distribution of titre prediction errors stratified by tested A/H3N2 strain. (**B**) Overall distribution of titre prediction errors across all measured viruses. Vertical red line shows *x=0*; buckets to the right of the red line suggest underestimation of titres; histograms to the left of the red line buckets suggest overestimation of titres.

**Figure S18:**
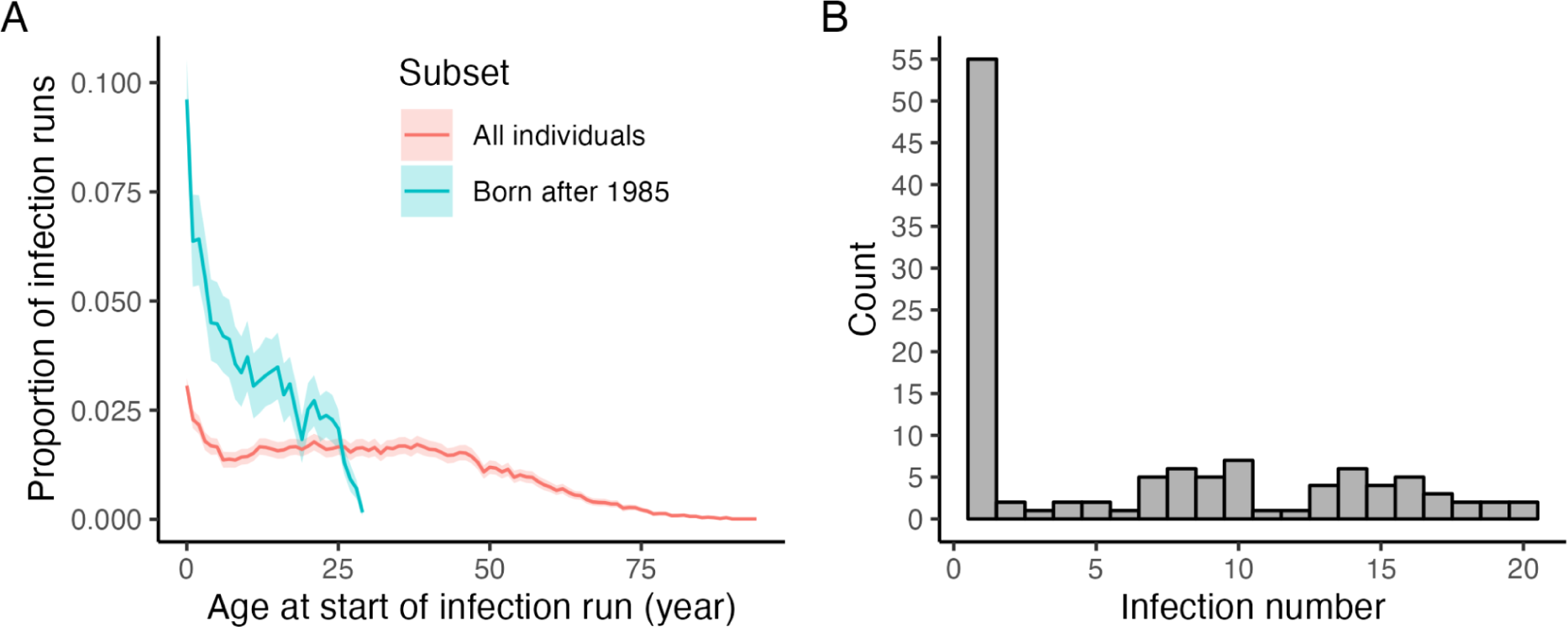
Runs of repeated infections are disproportionately more likely to occur early in life. (**A**) Proportion of infection episodes which are estimated as runs of consecutive infections by age at start of infection run, using either all infection episodes or only those from individuals born after 1985. (**B**) Number of inferred infection episodes which are estimated as runs of consecutive infections stratified by the infection order in each individual’s infection history. For example, if the run is the first infection an individual has experienced, this is given an infection number of one. Of the 10,558 distinct infection episodes, 757 (posterior median; 95% CrI: 676-861) were runs of two consecutive infections, 79 (posterior median; 95% CrI: 62-97) were runs of 3 consecutive infections and 19 (posterior median; 95% CrI: 13-27) were runs of 4 or more.

**Figure S19:**
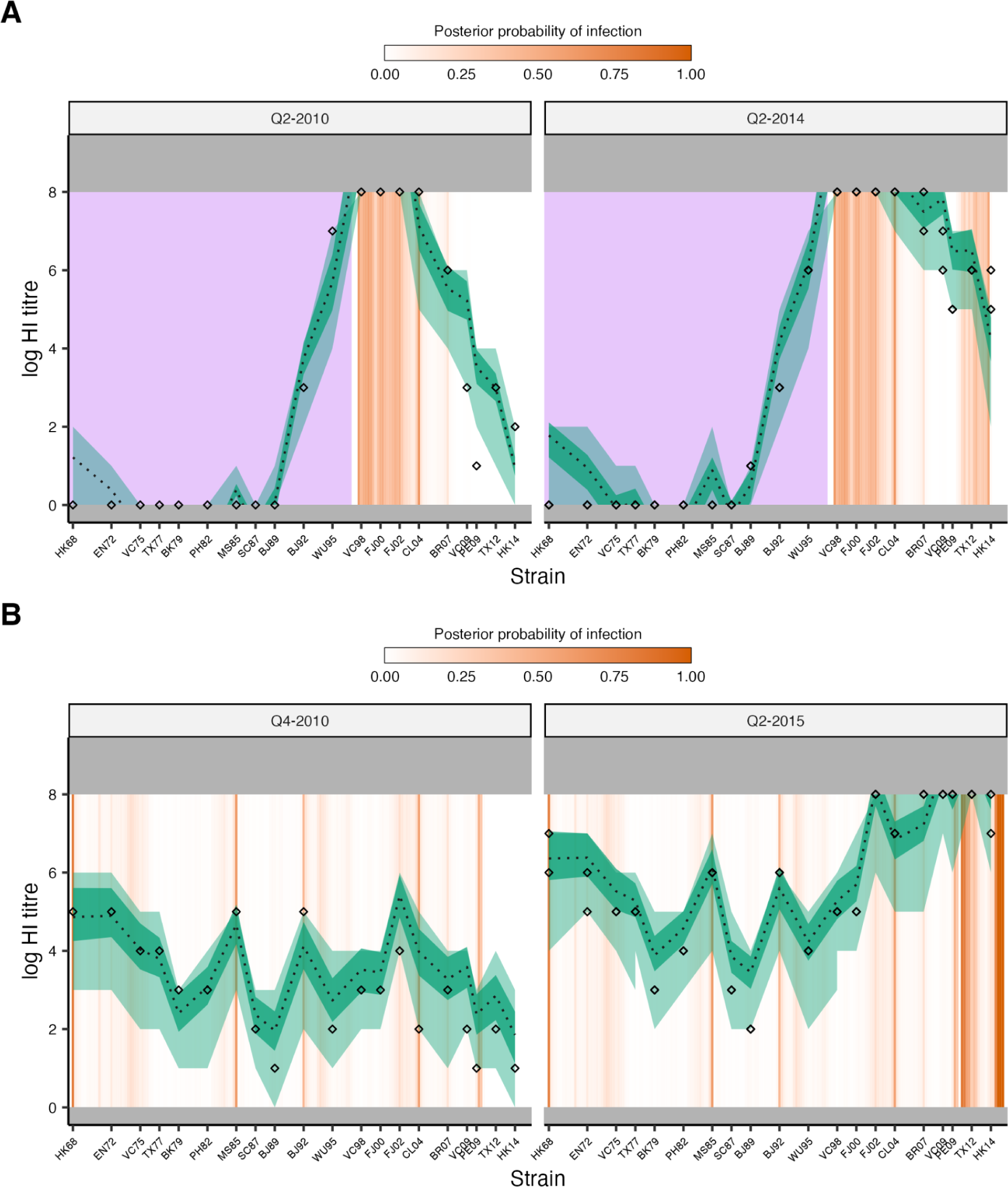
Model fits demonstrating typical profiles where consecutive infection runs were imputed. Model-predicted titres compared to observed HI titres at each sampling time for two individuals, as in Fig S4. Diamonds show titre measurements; green shaded region shows 95% CrI and 95% prediction intervals; dashed line shows posterior median; orange bars show posterior probability of infection in a given time window. Purple rectangles show time periods prior to birth. (**A**) Example of an individual estimated to have experienced multiple consecutive infections immediately following birth to explain high titres. (**B**) Example of an individual estimated to have experienced multiple consecutive infections between the two serum sampling times to explain the drastic increase in titres to recently circulating strains.

**Figure S20:**
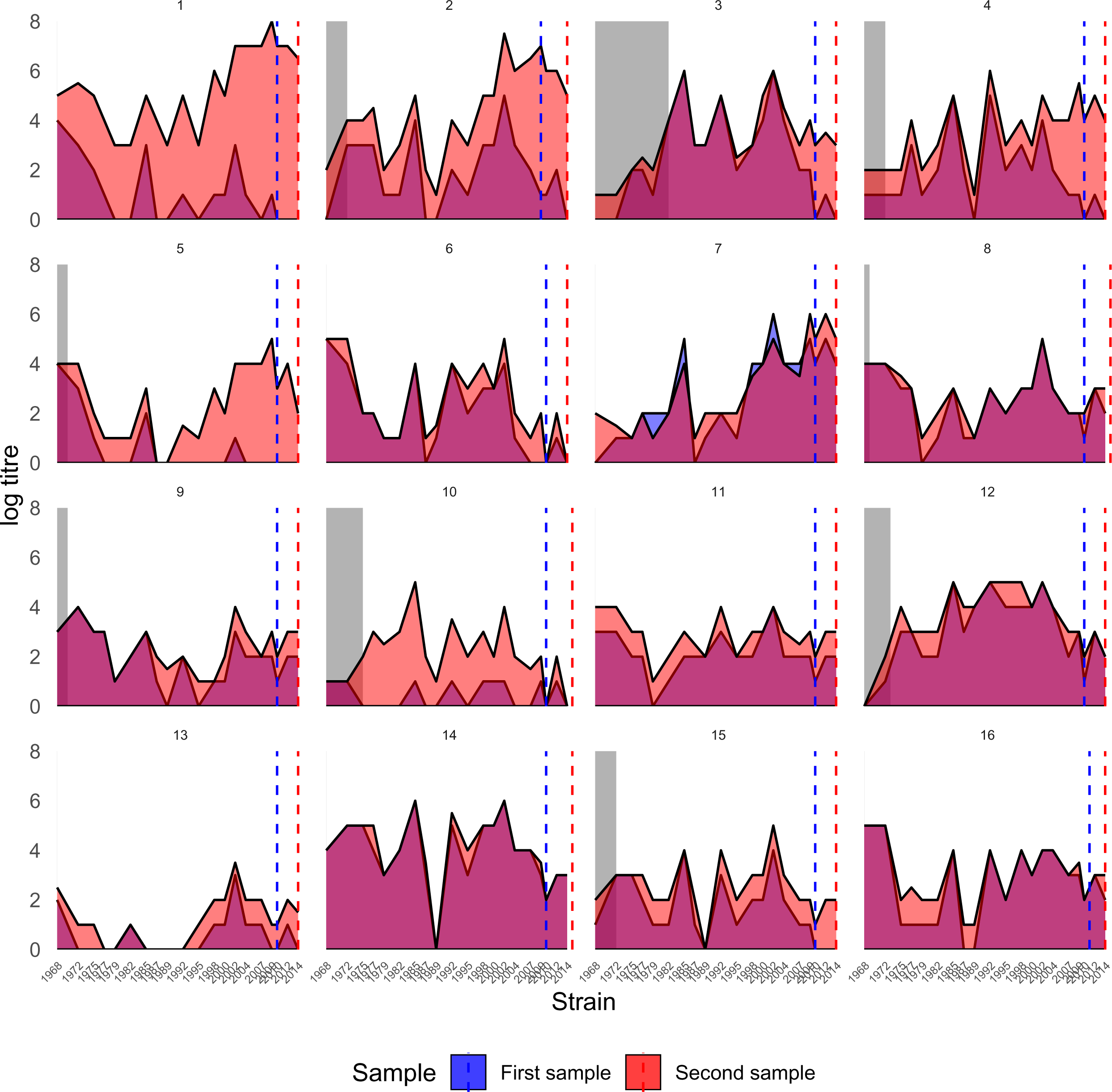
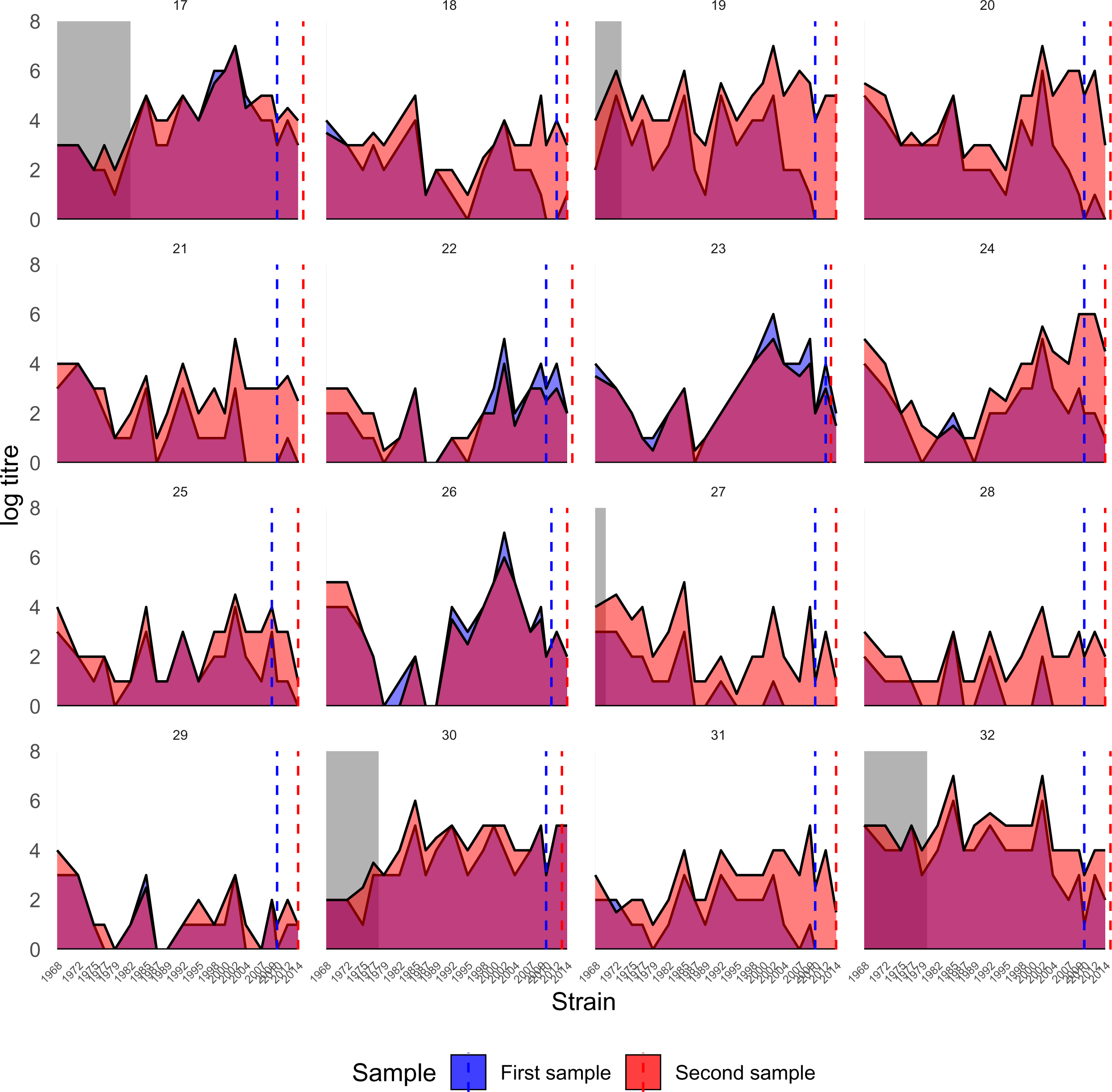
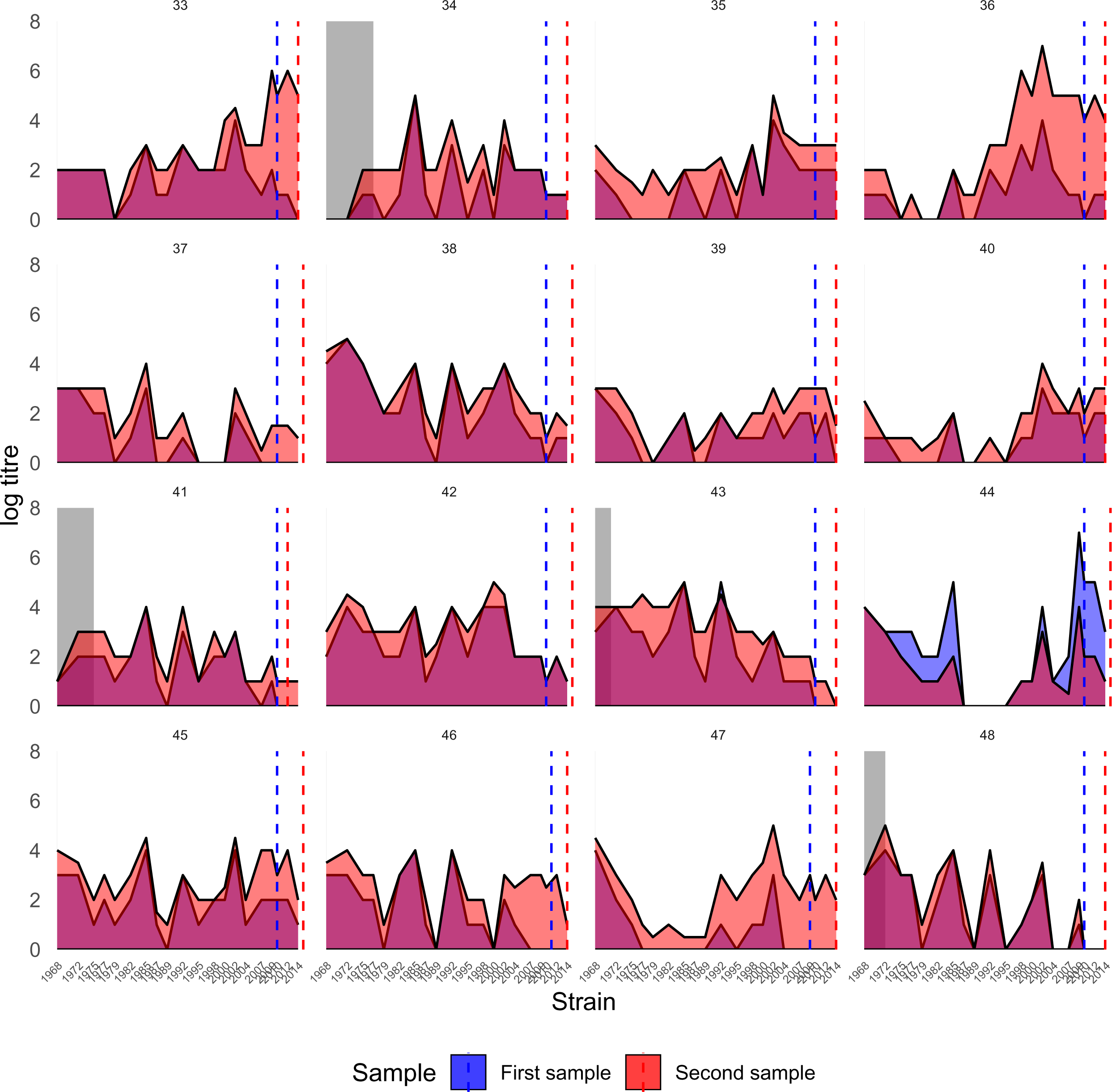
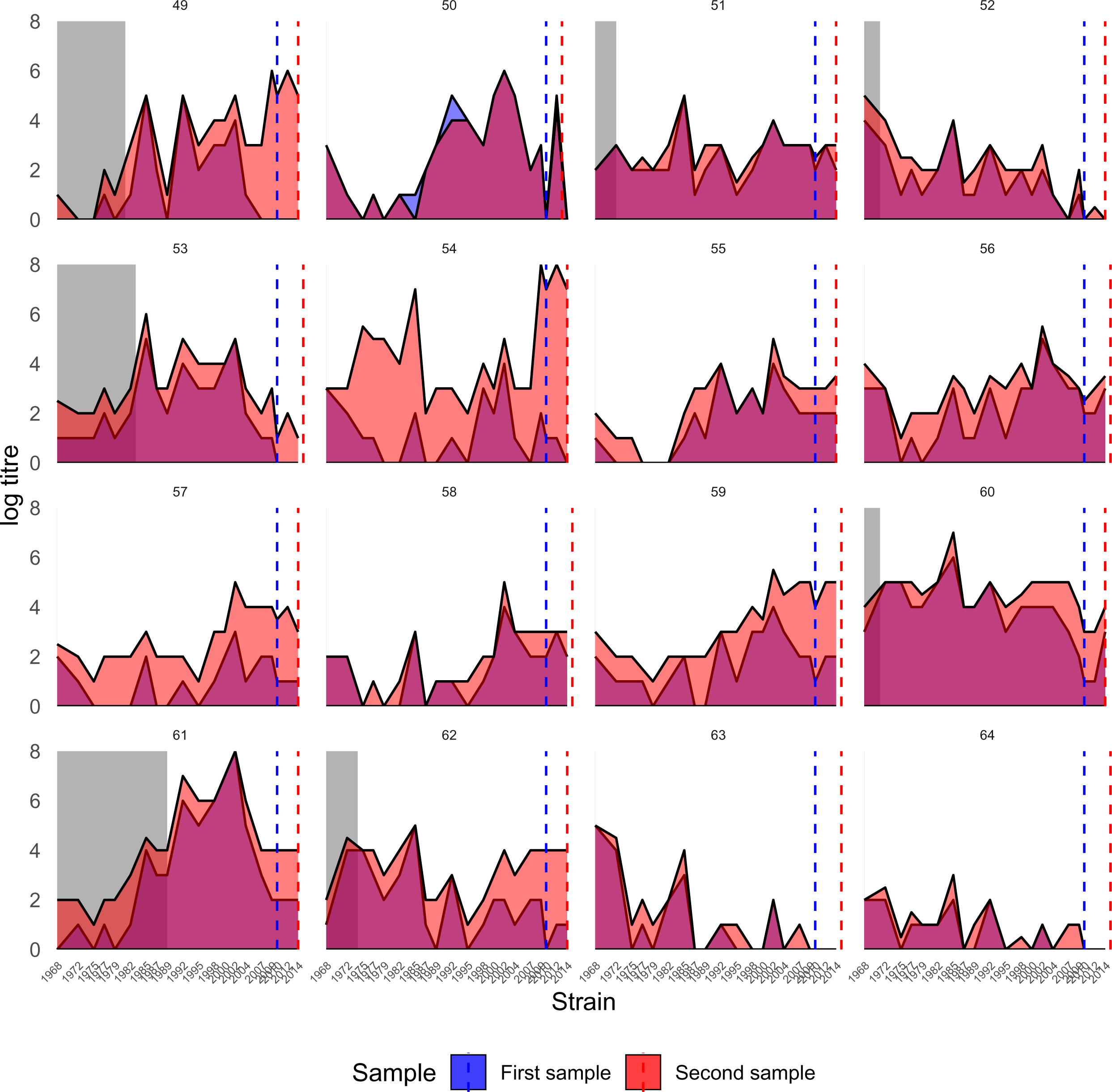
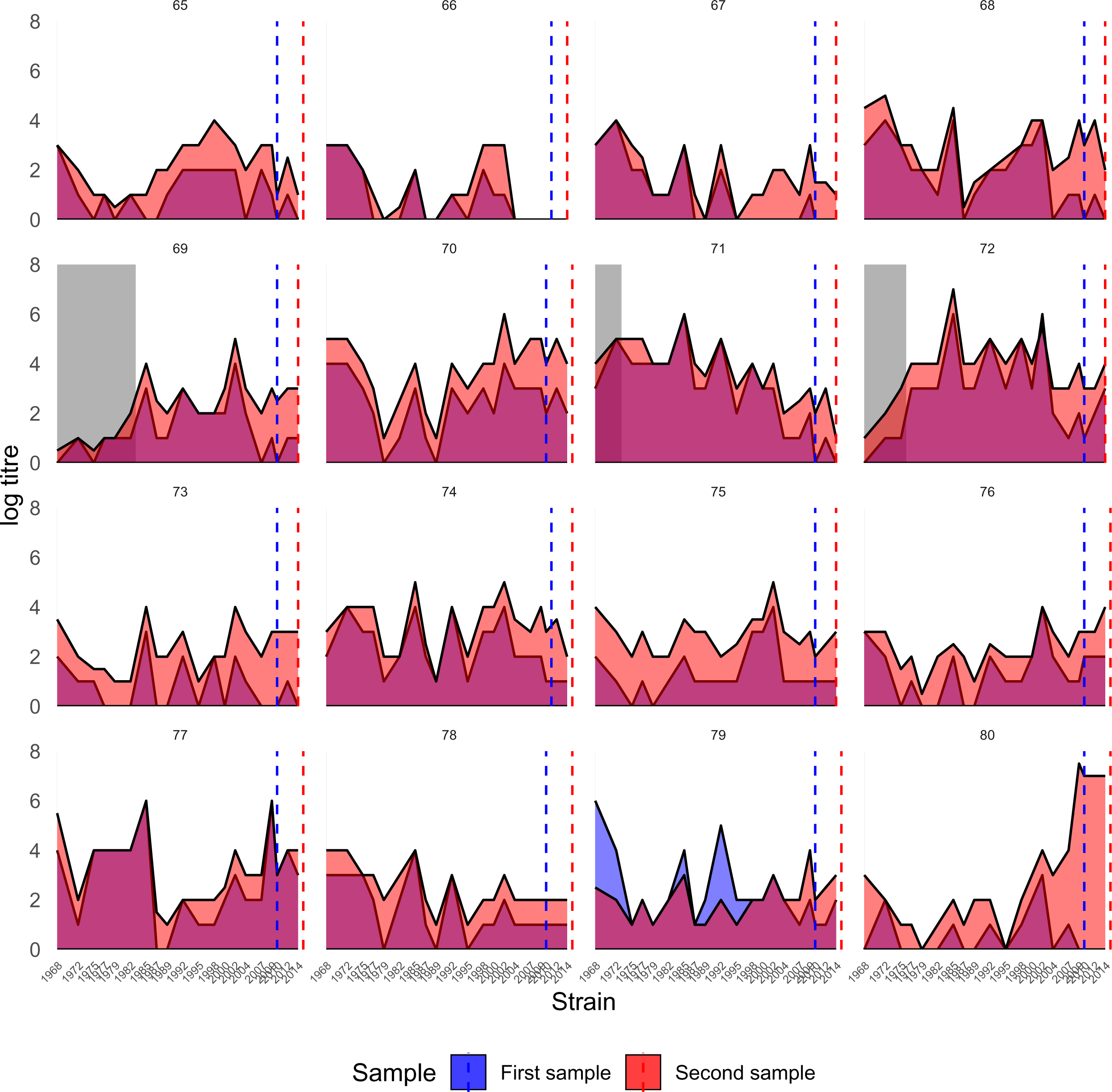
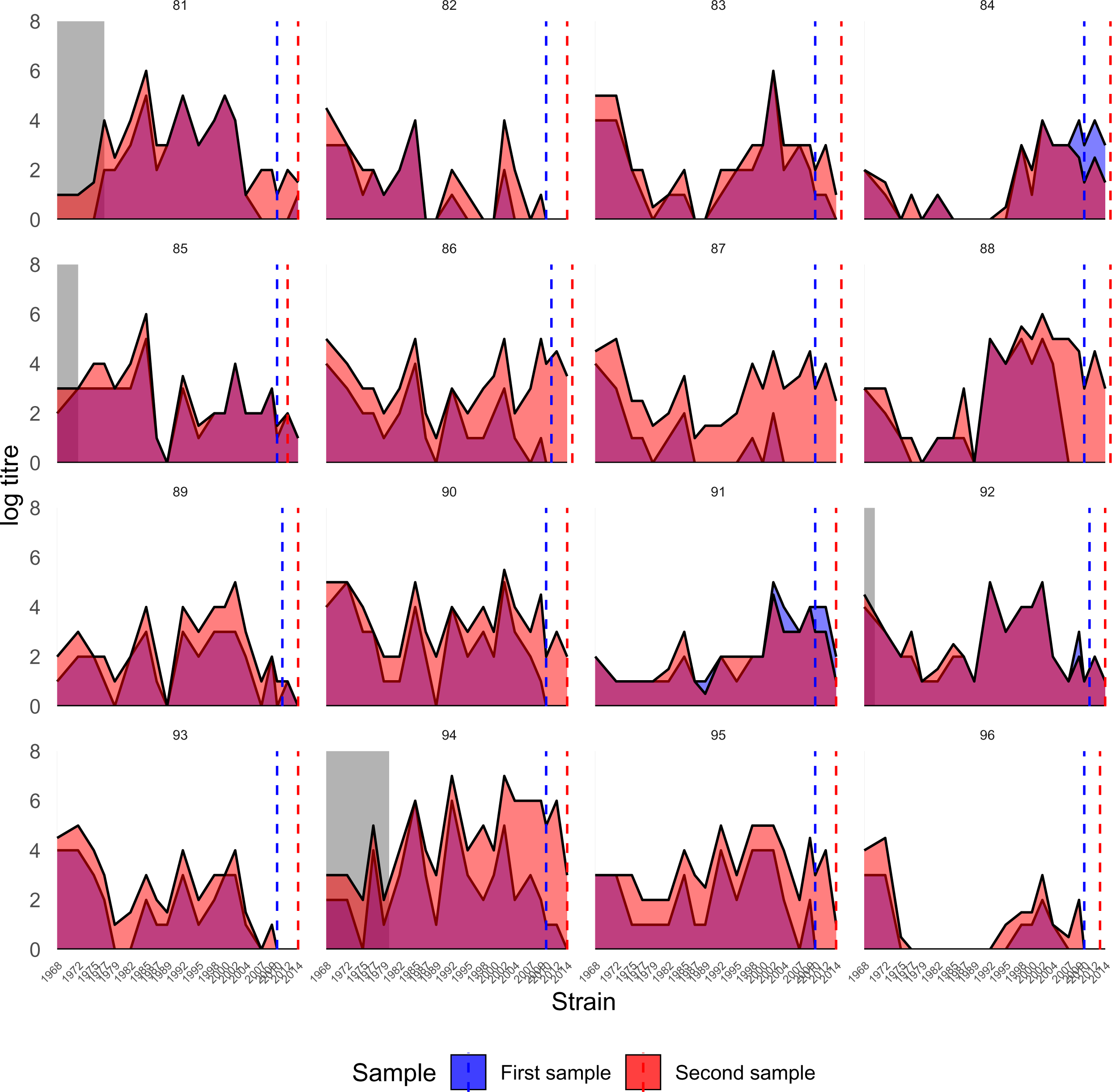
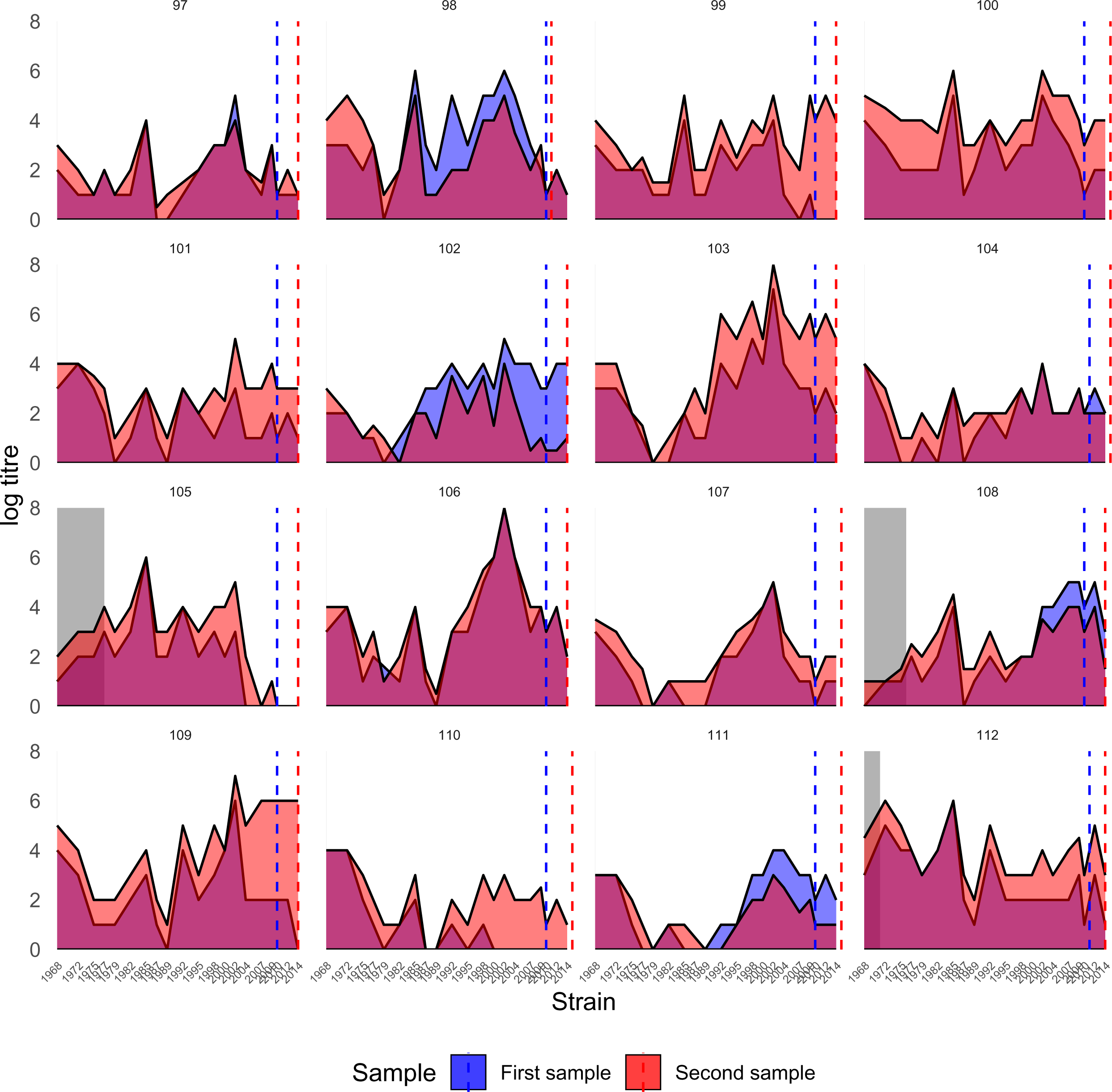
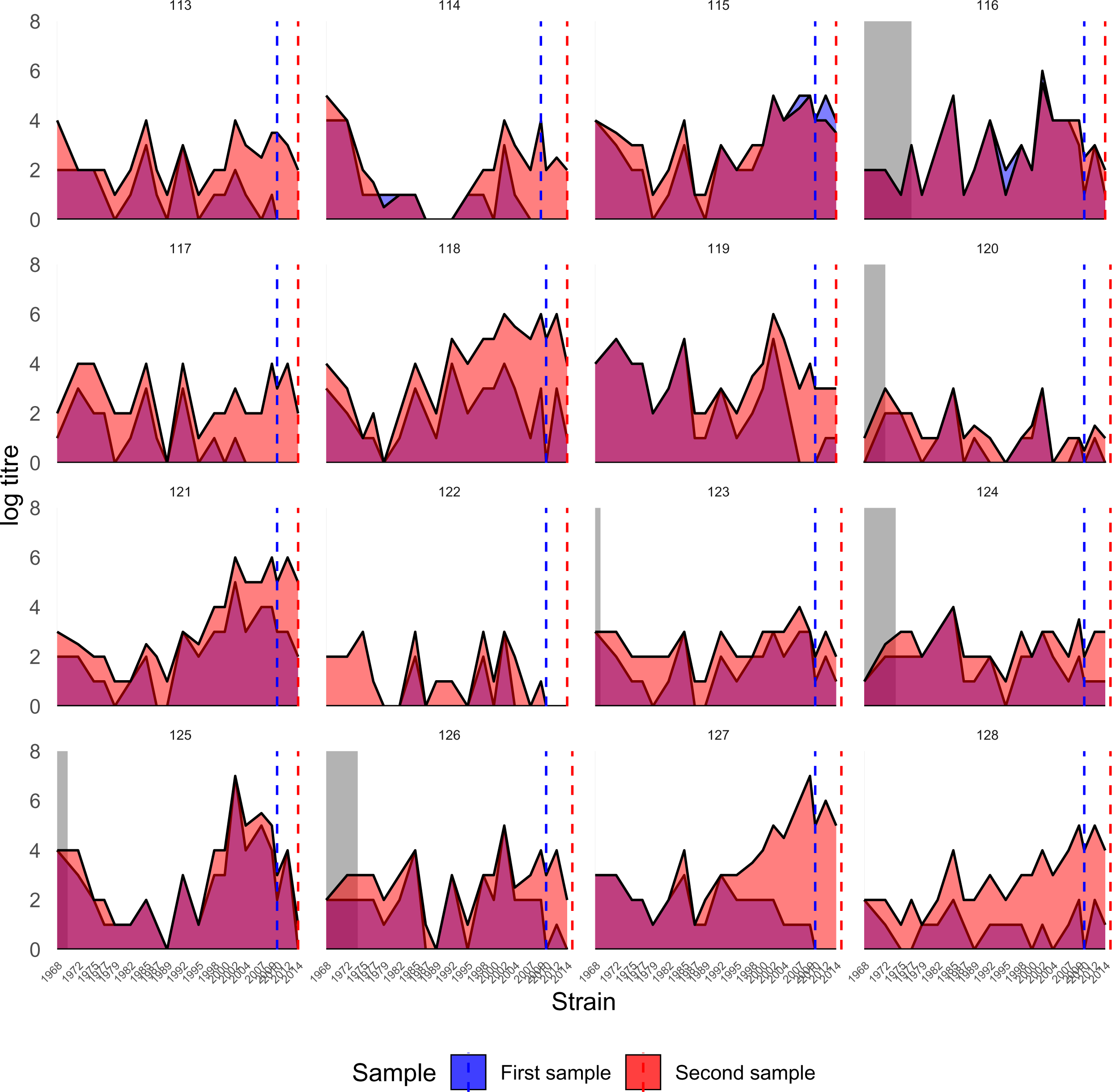
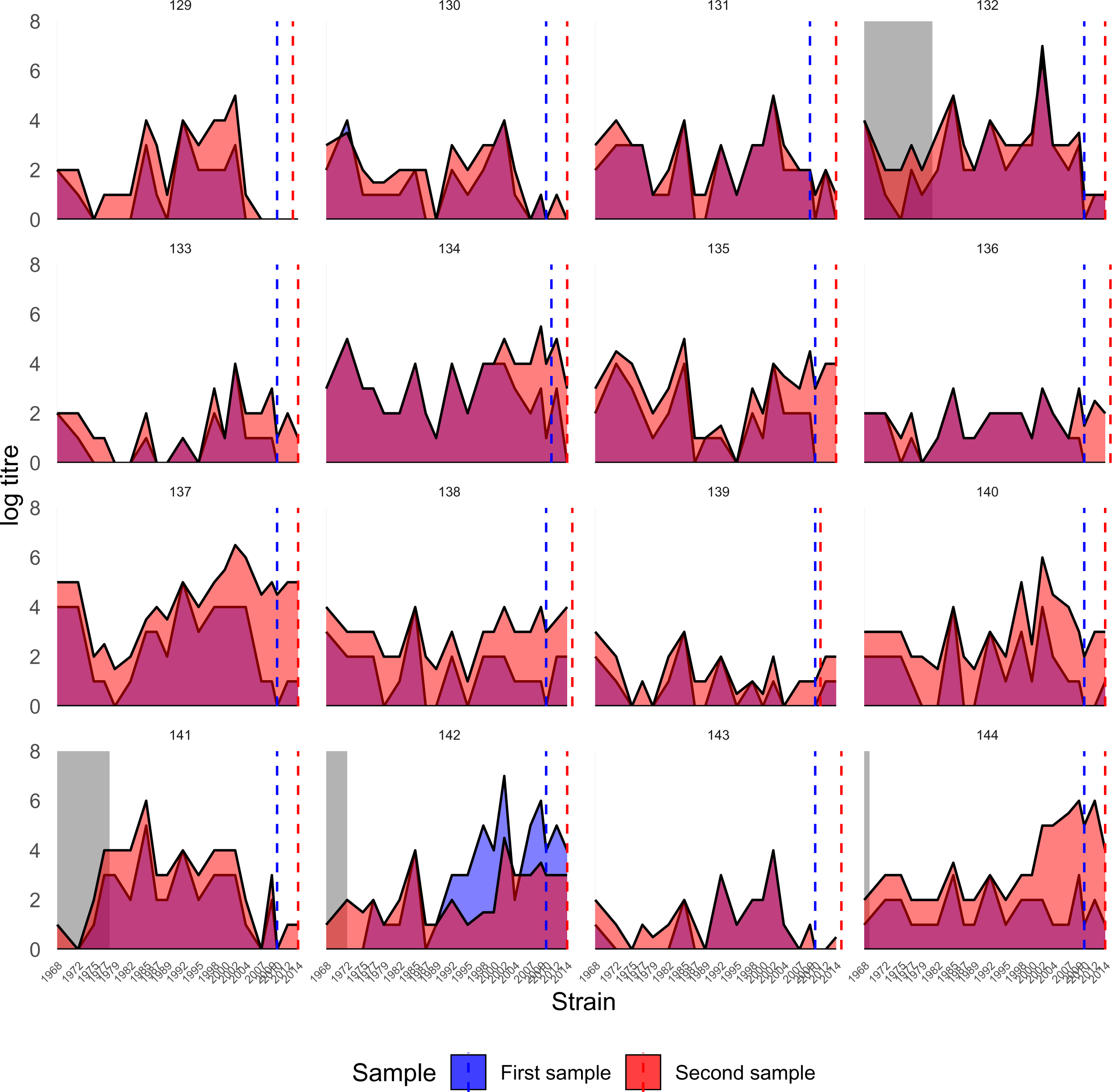
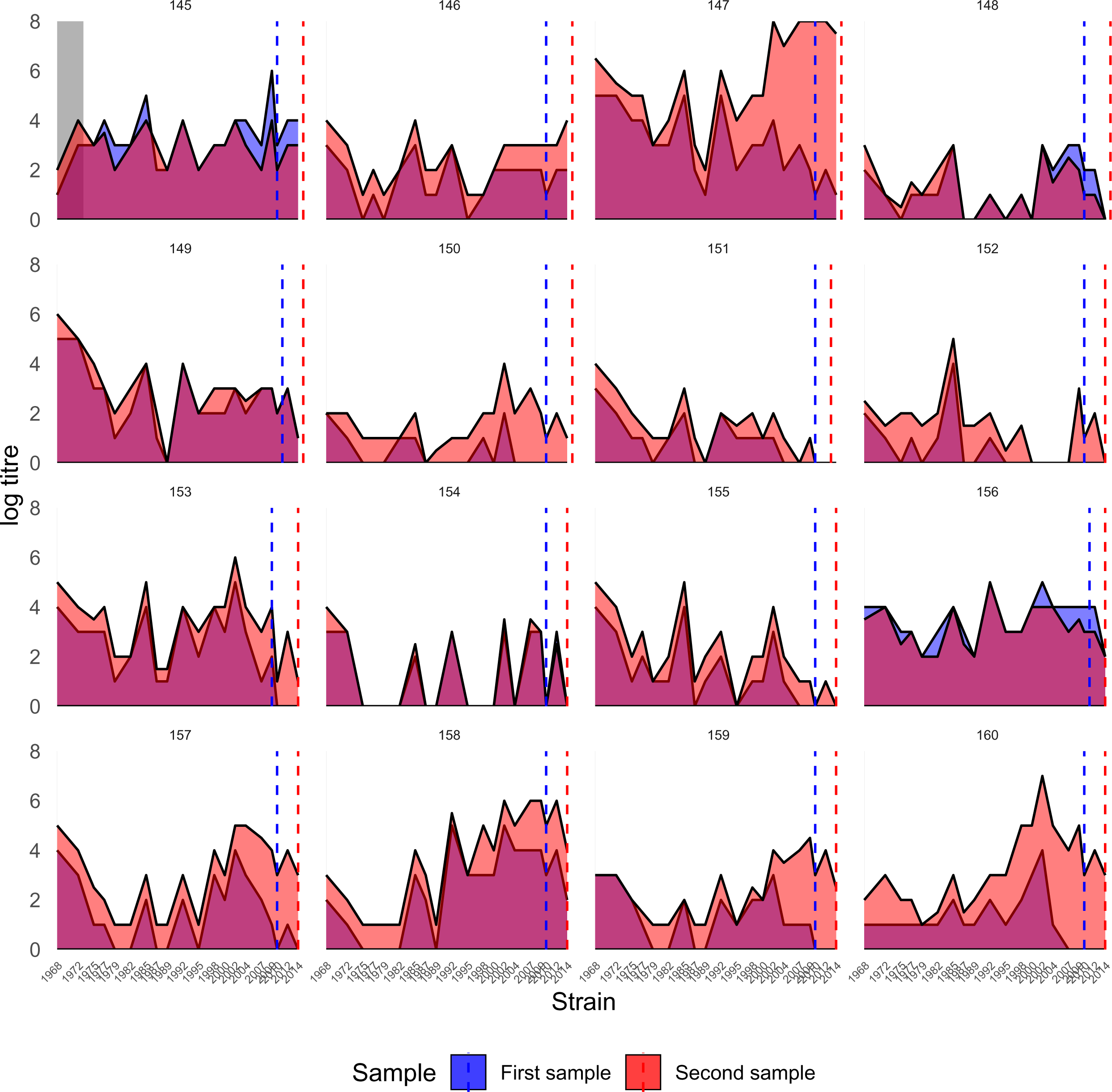
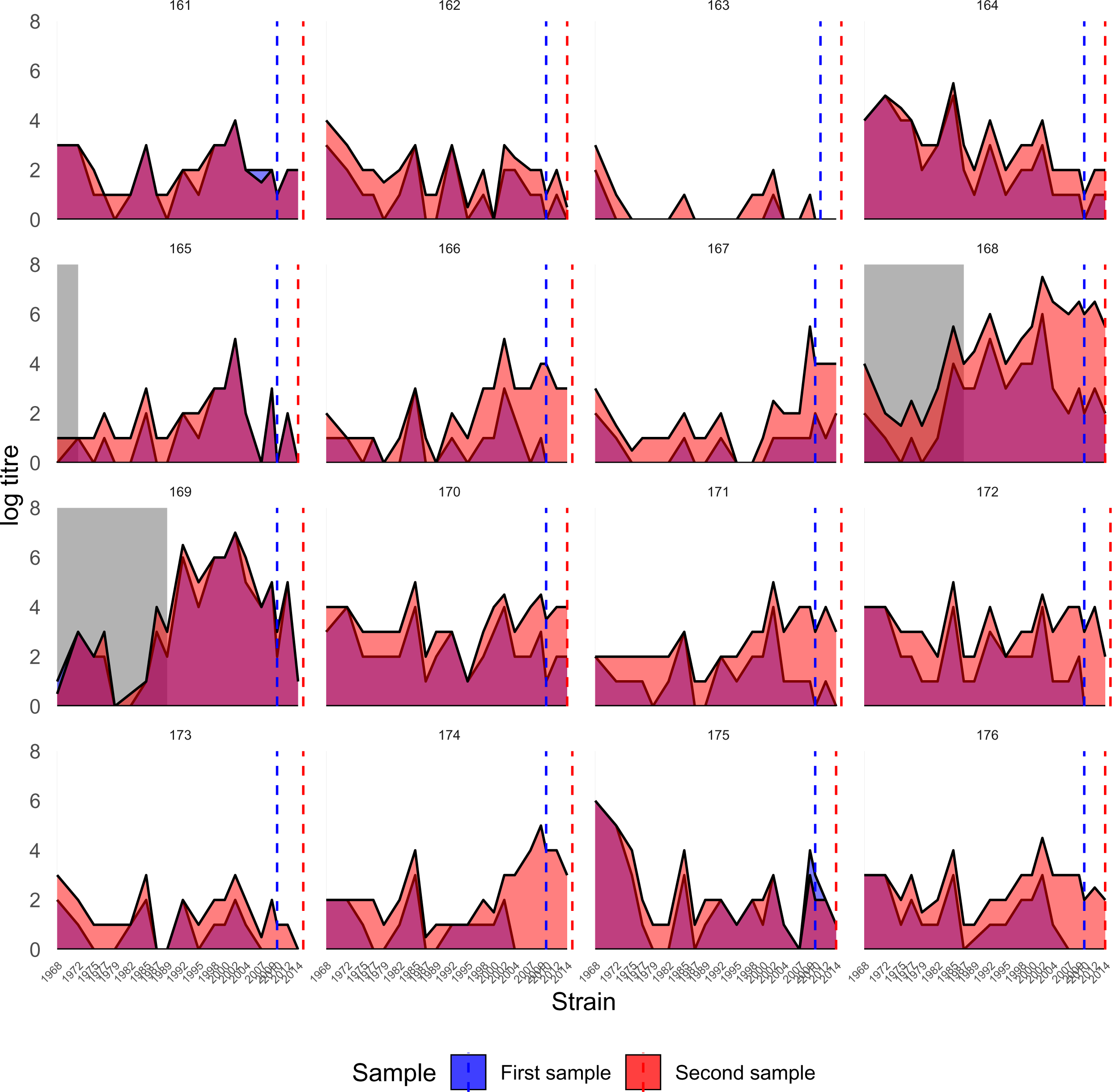
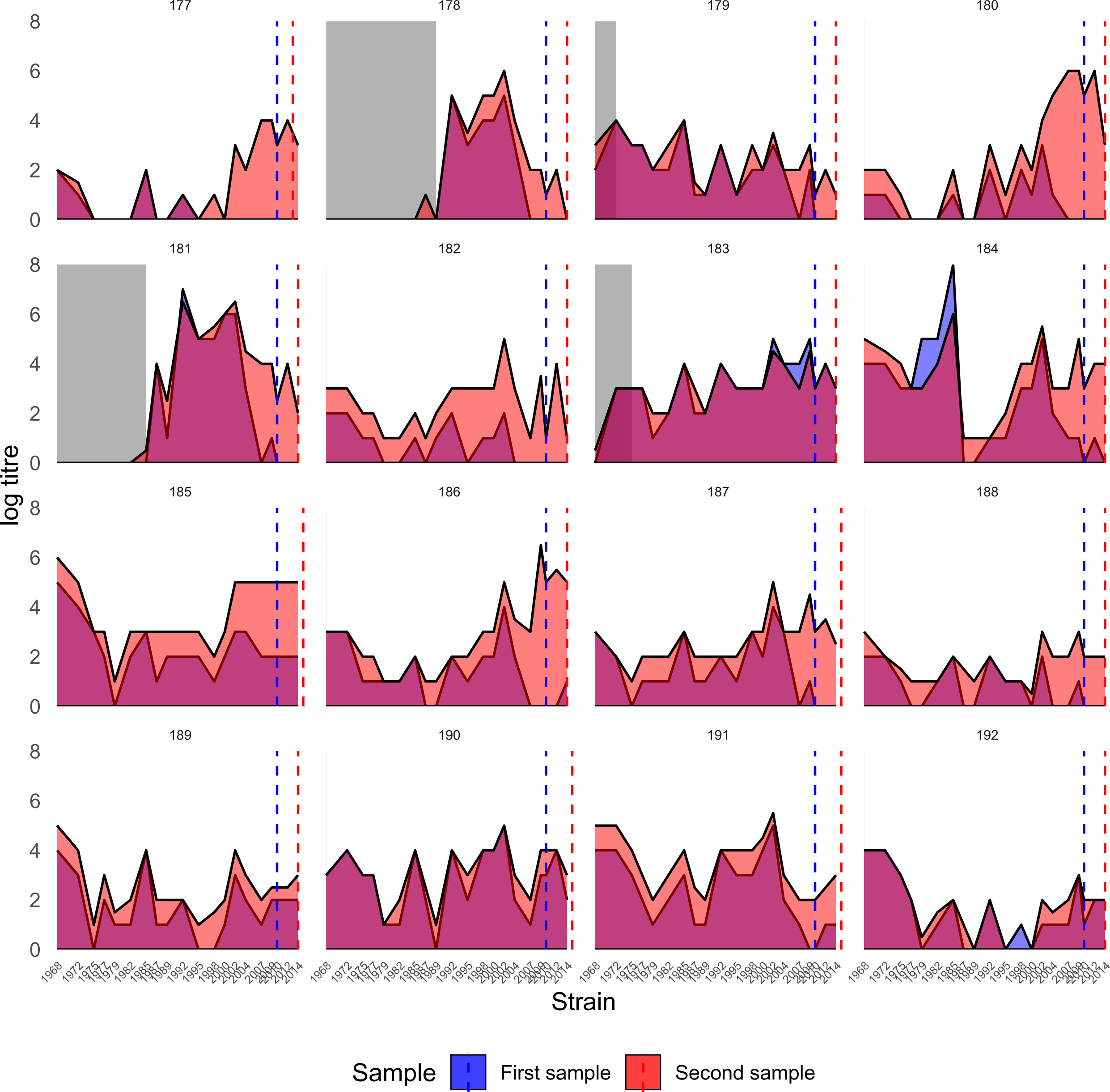
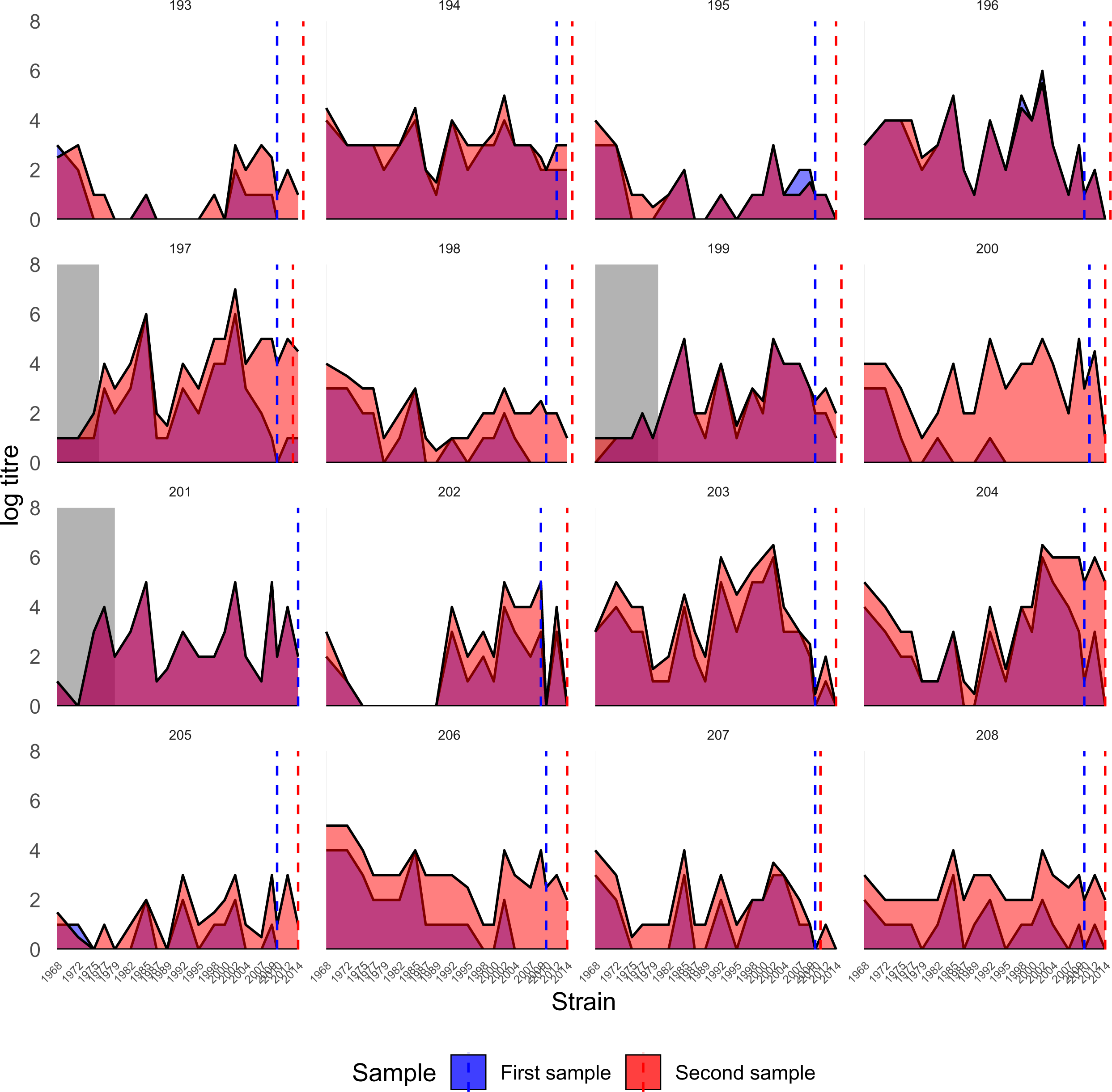
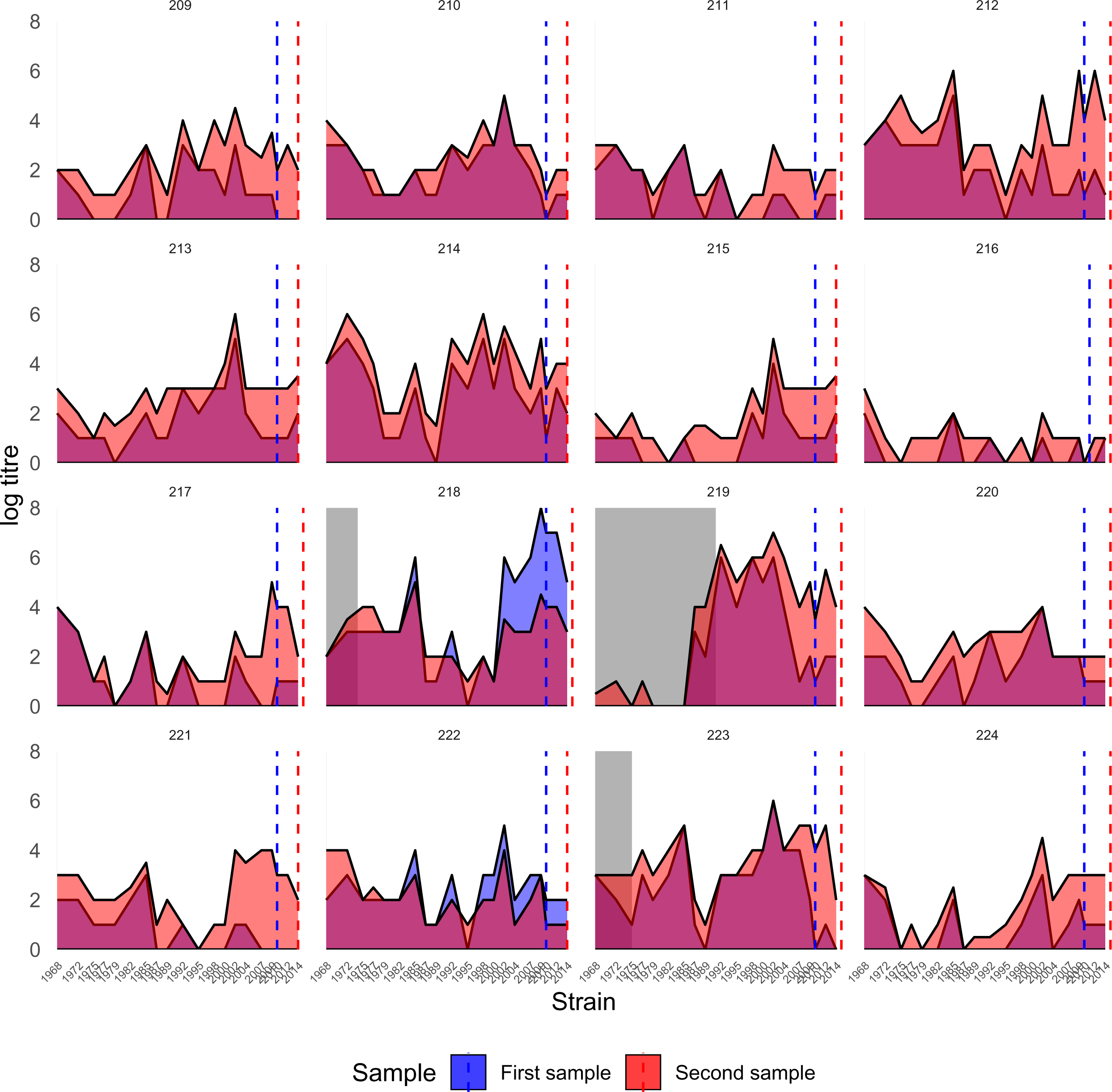
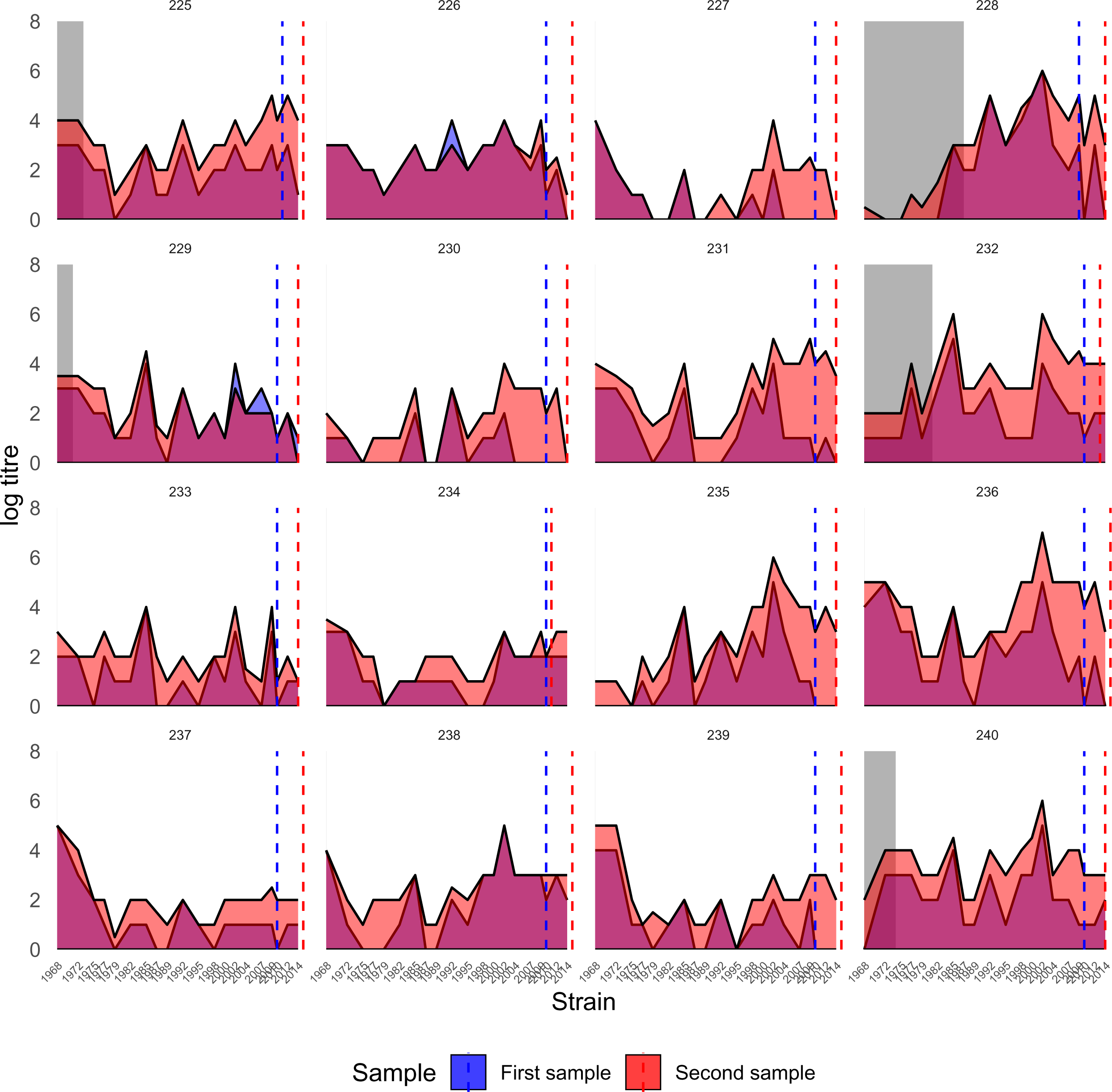
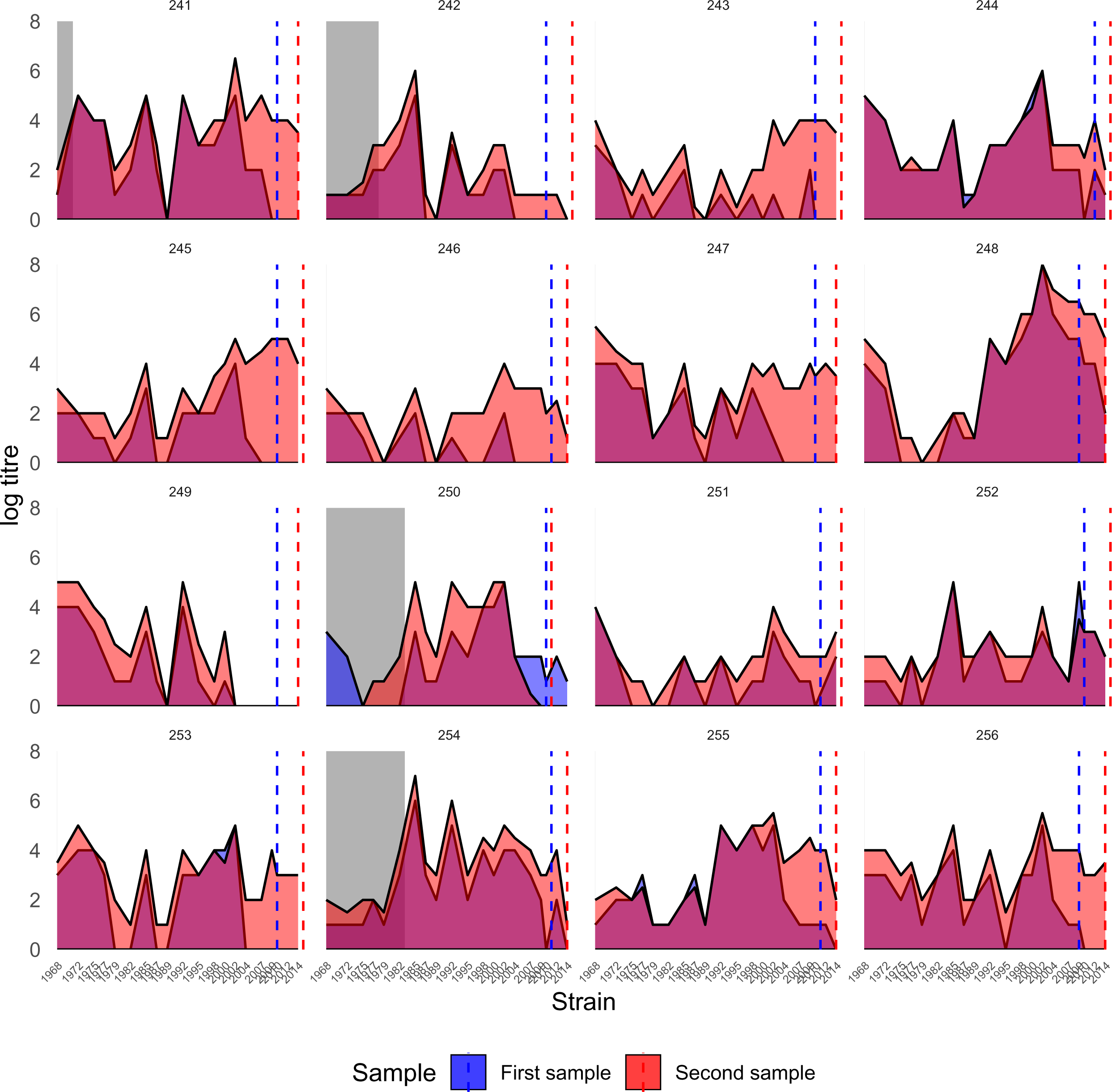
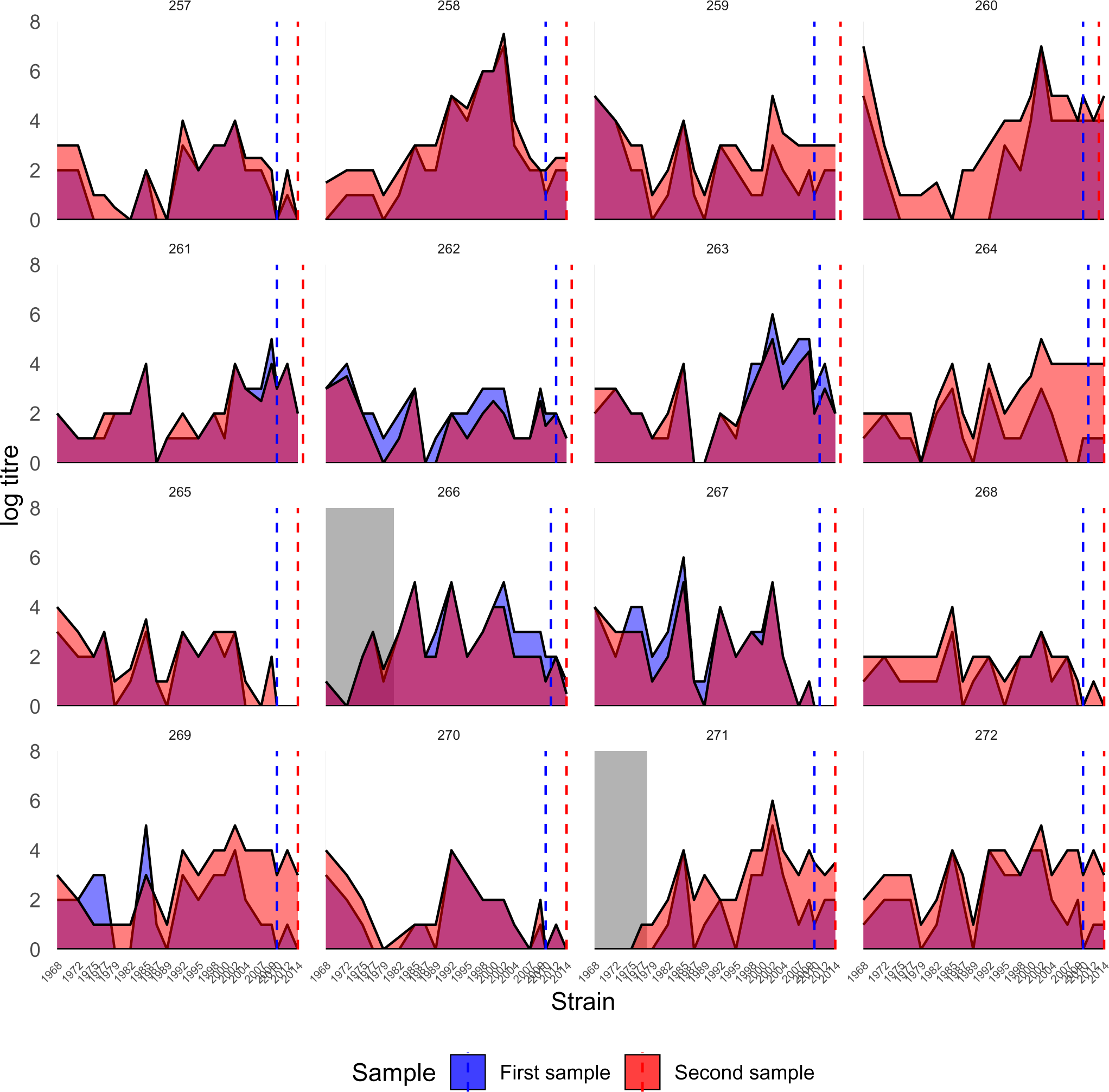
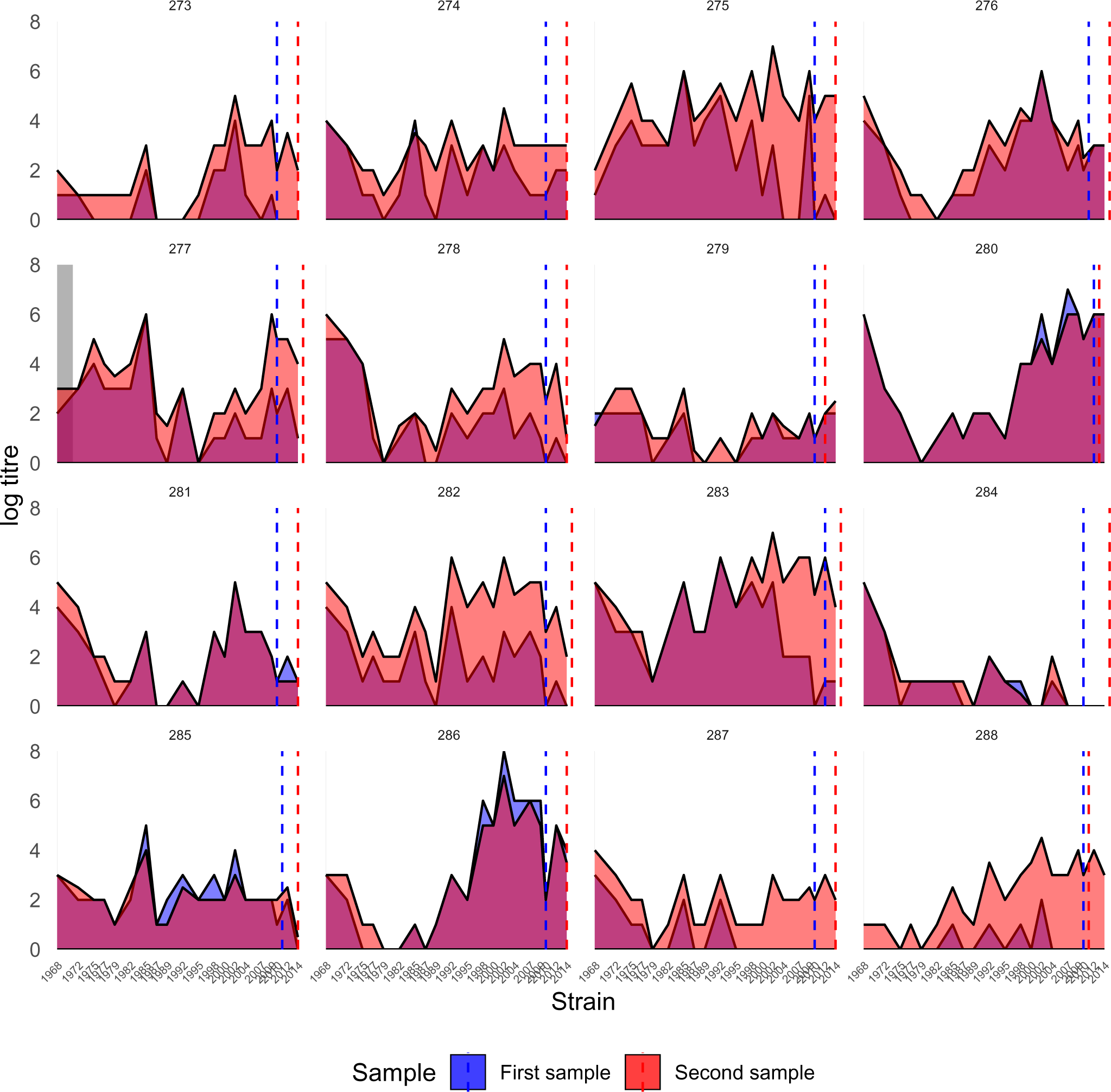
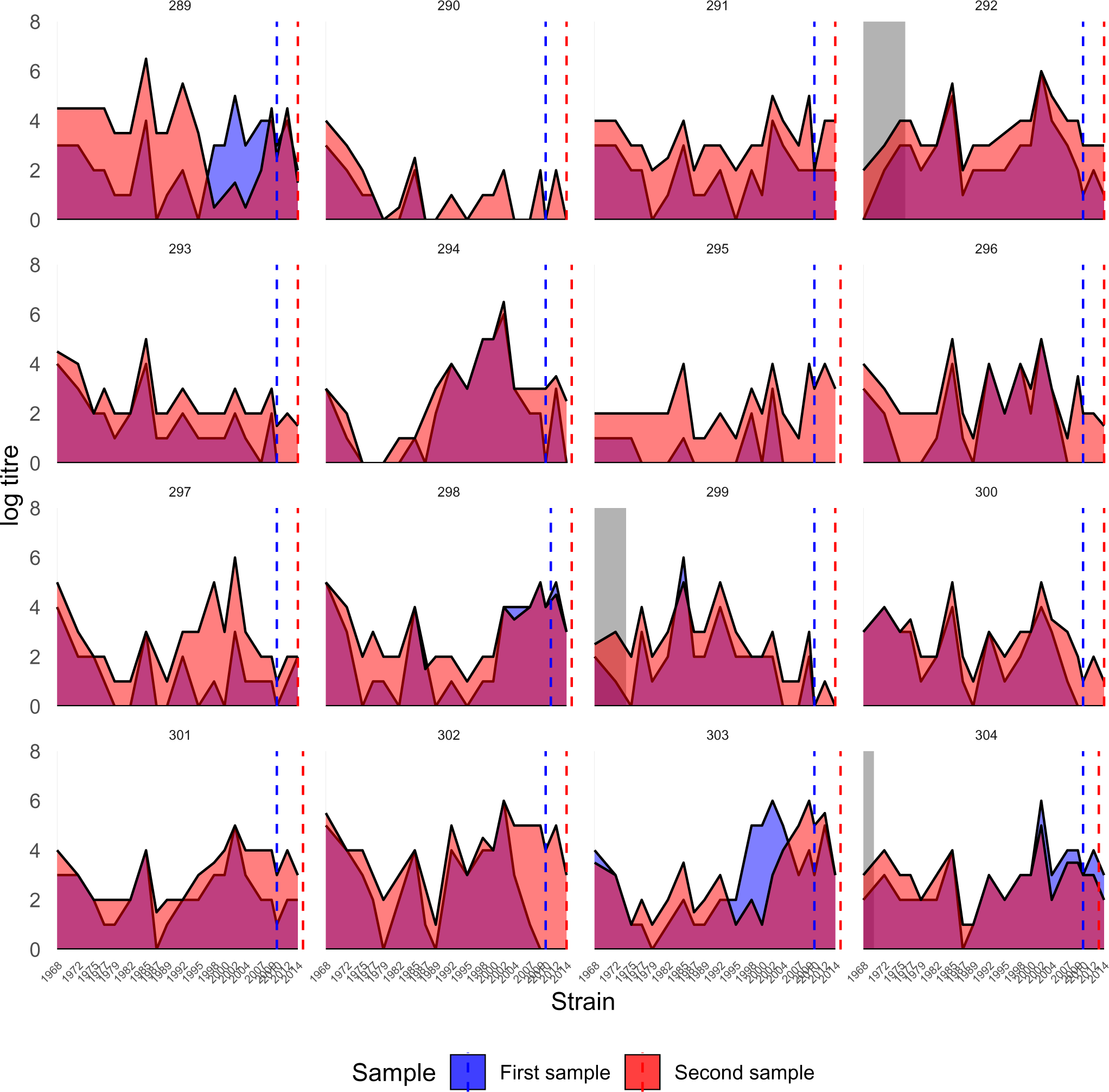
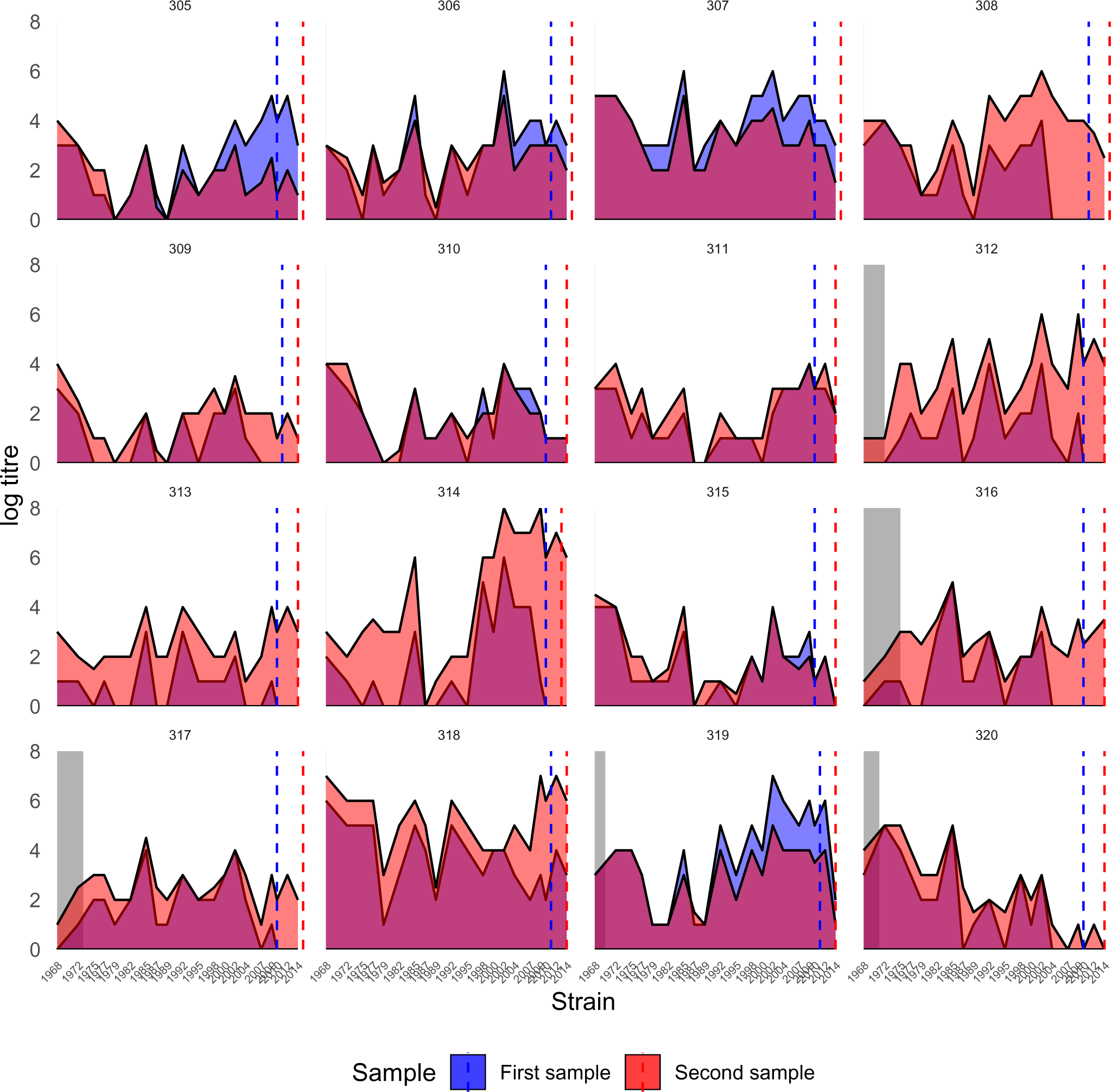
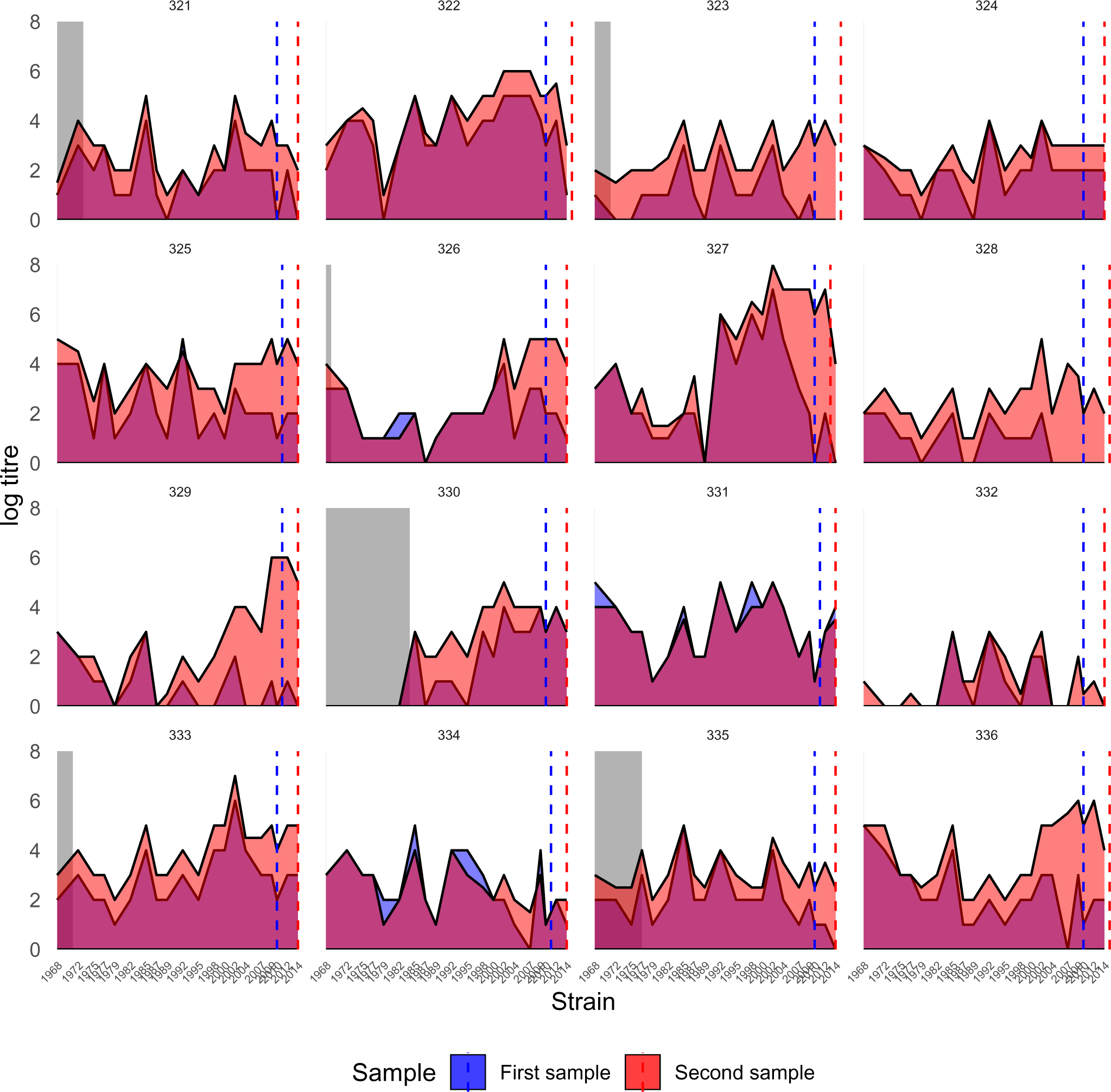
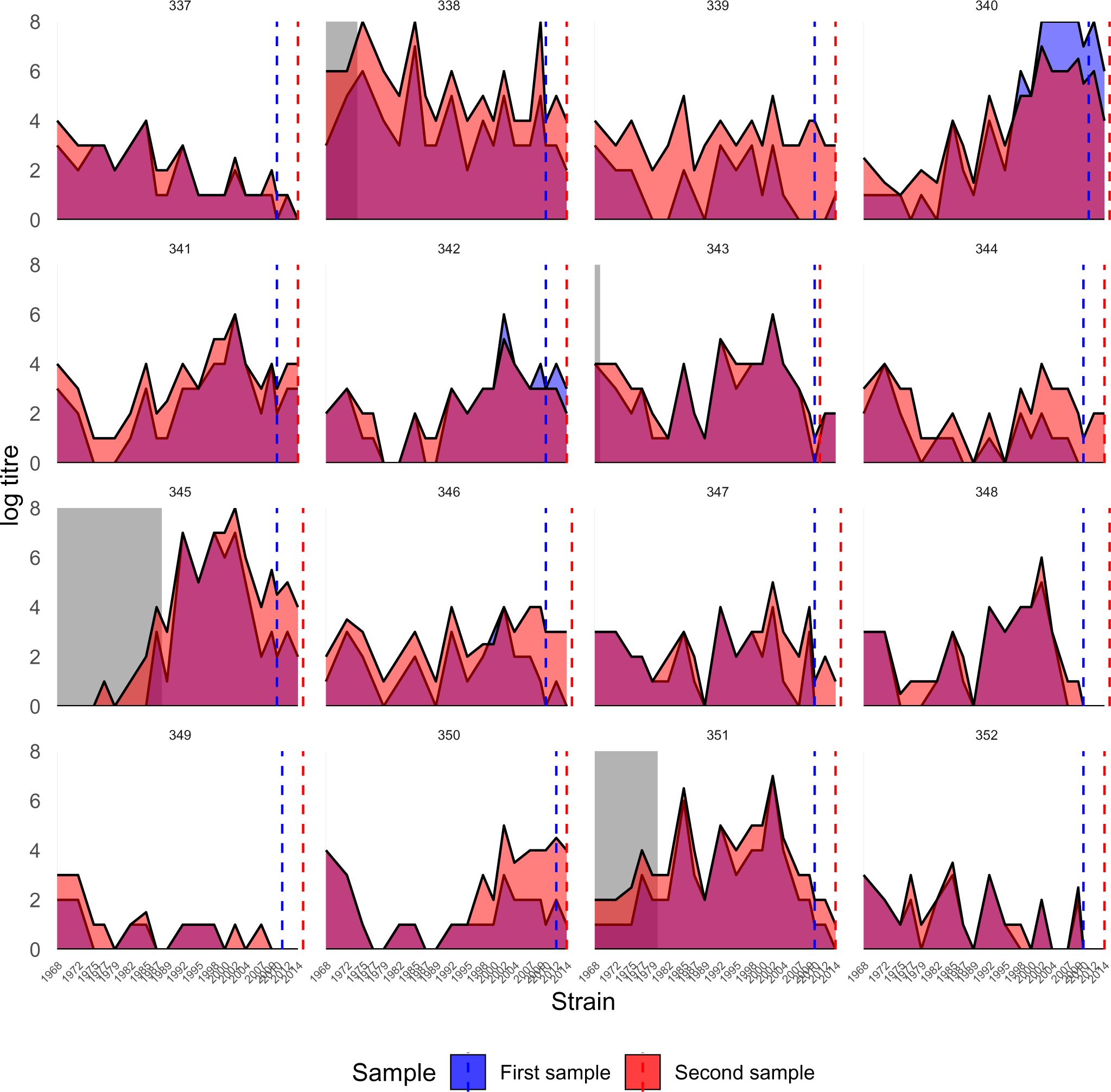
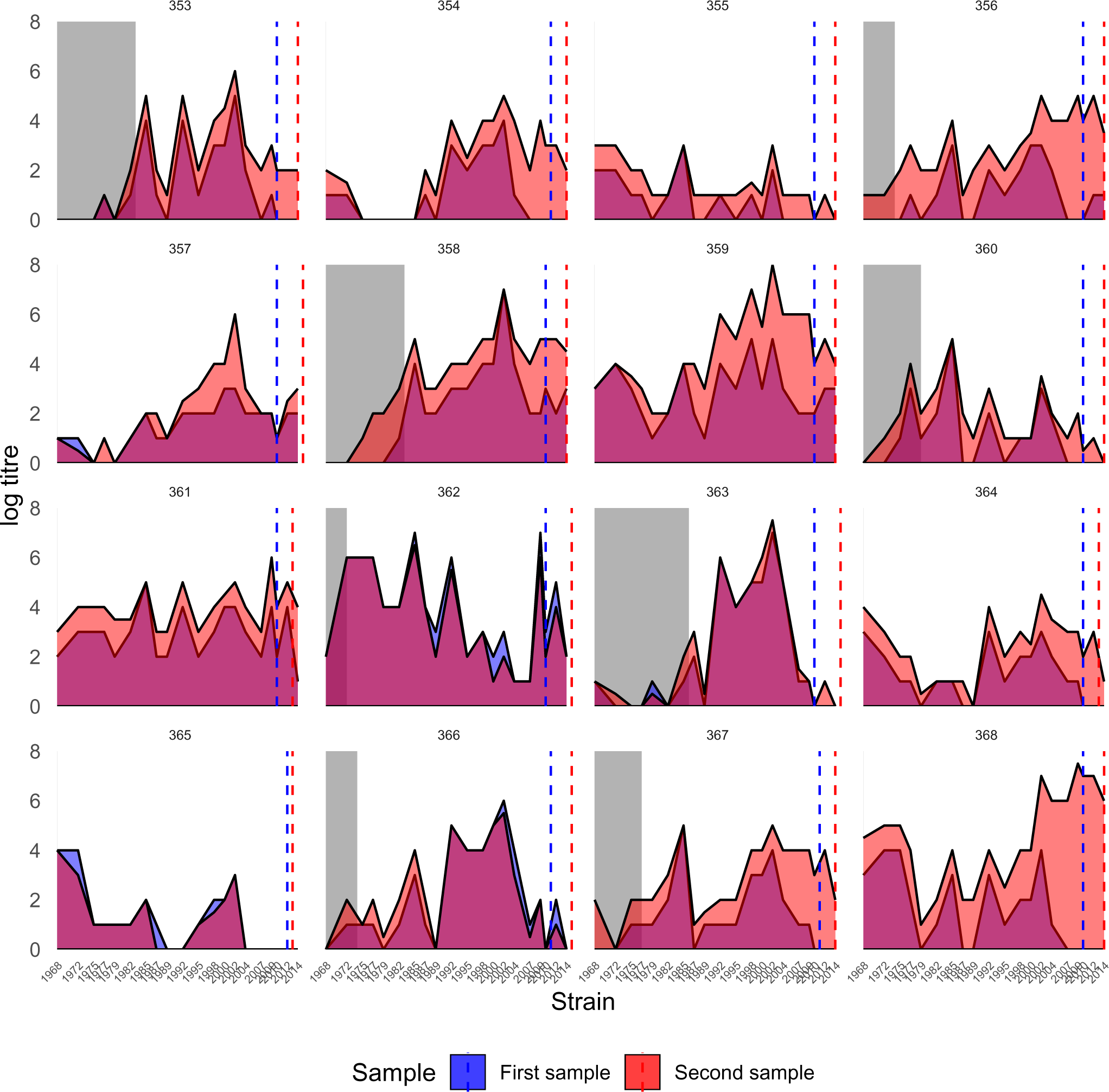
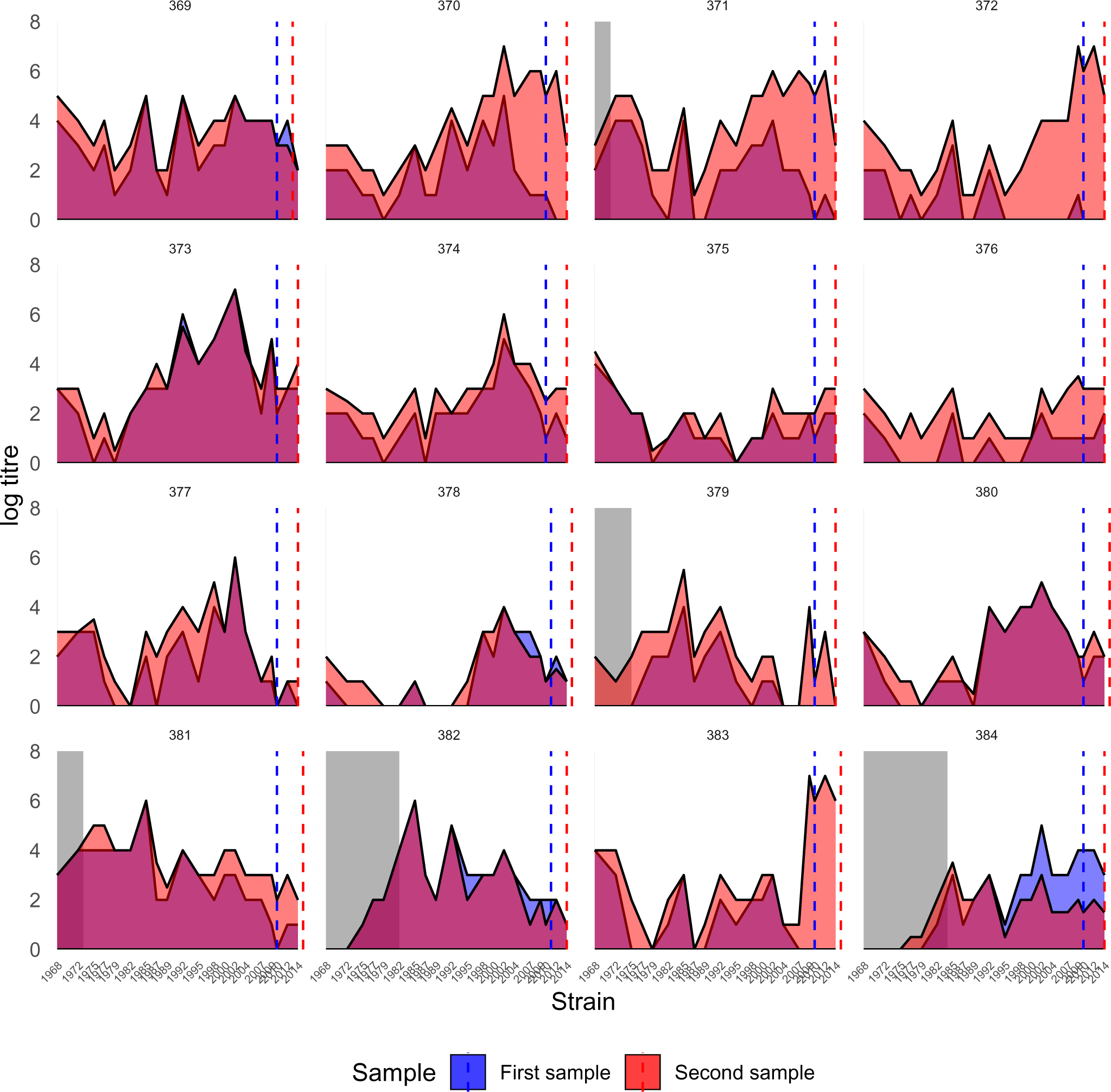
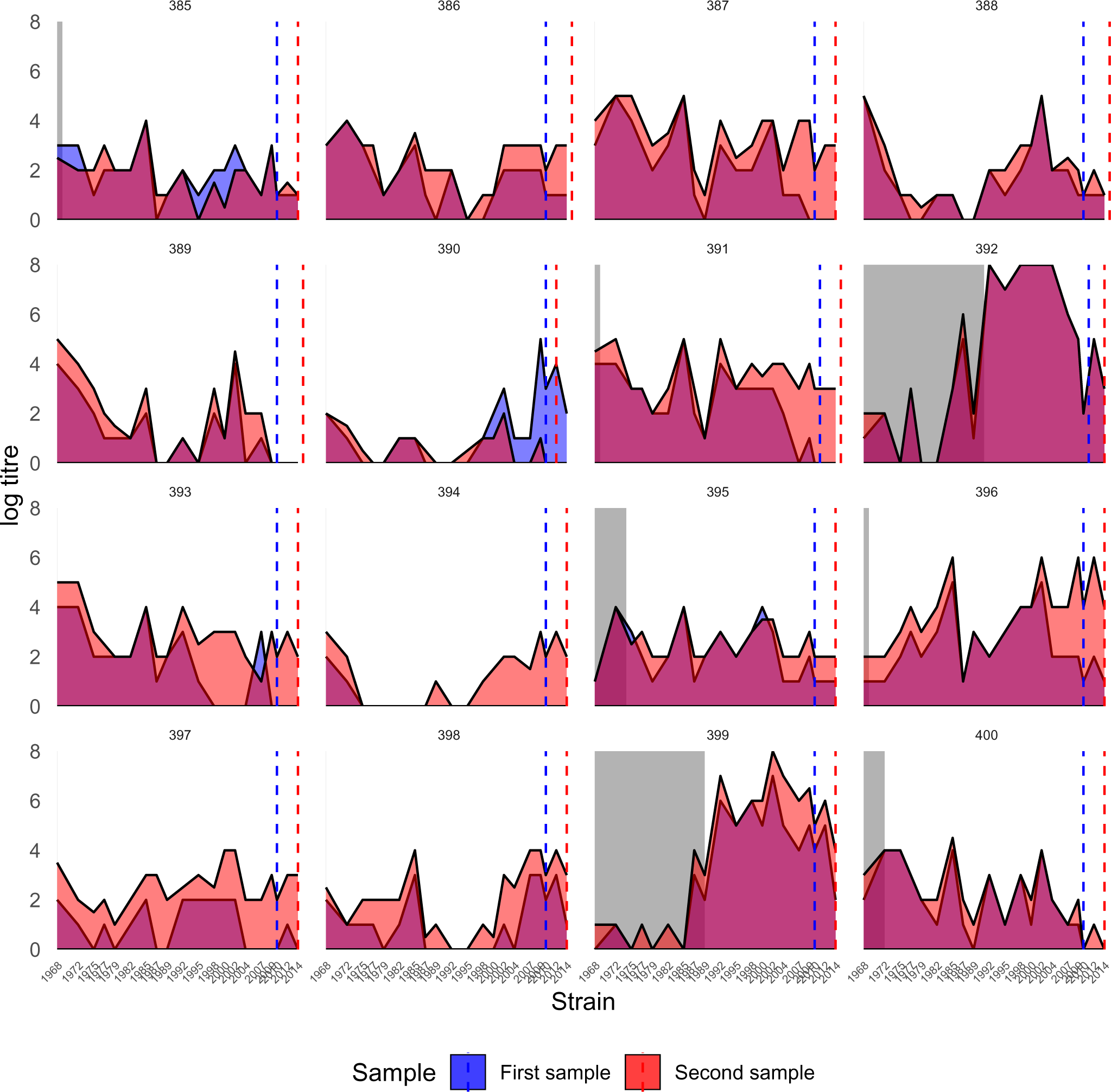
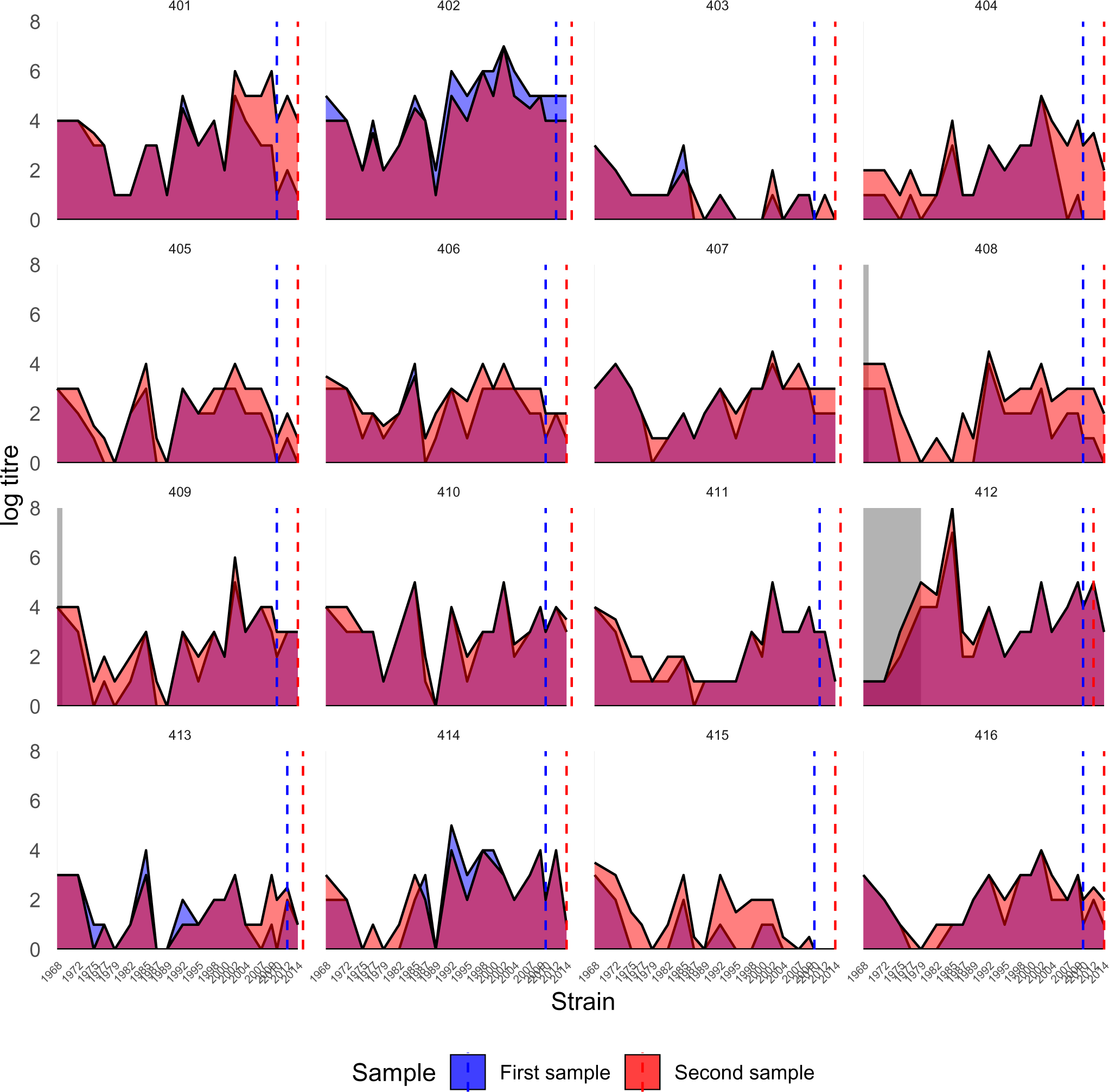
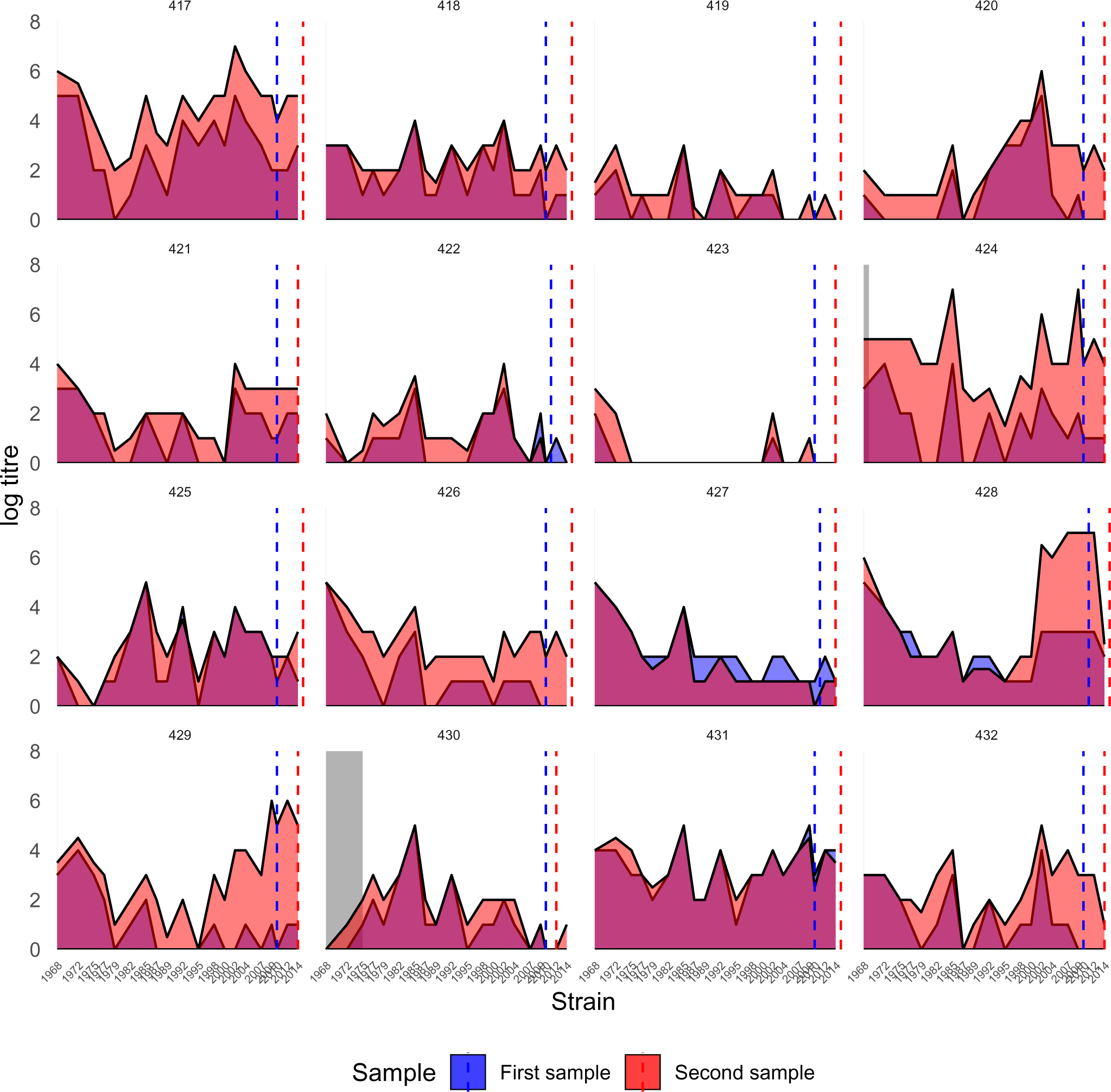
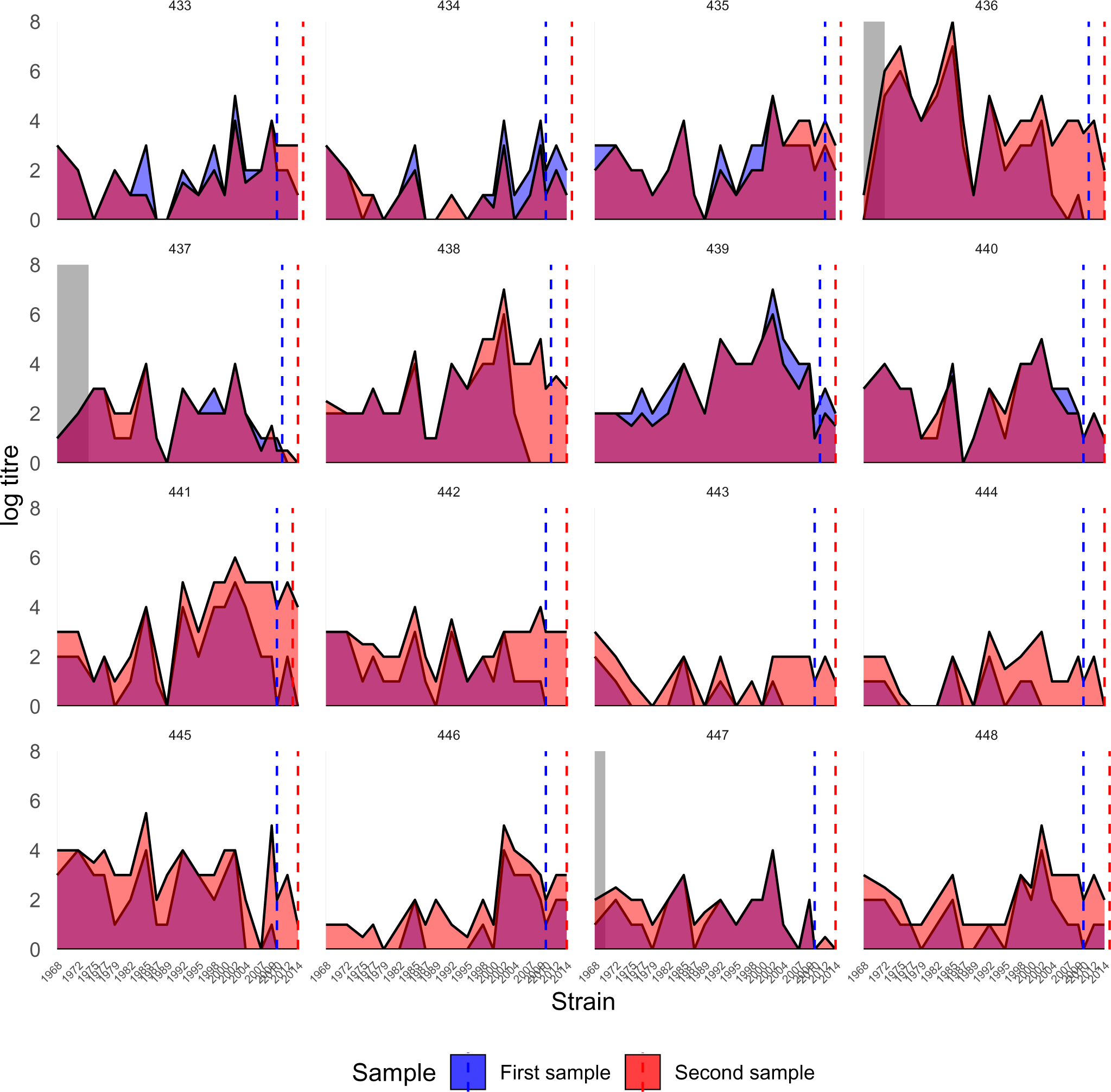
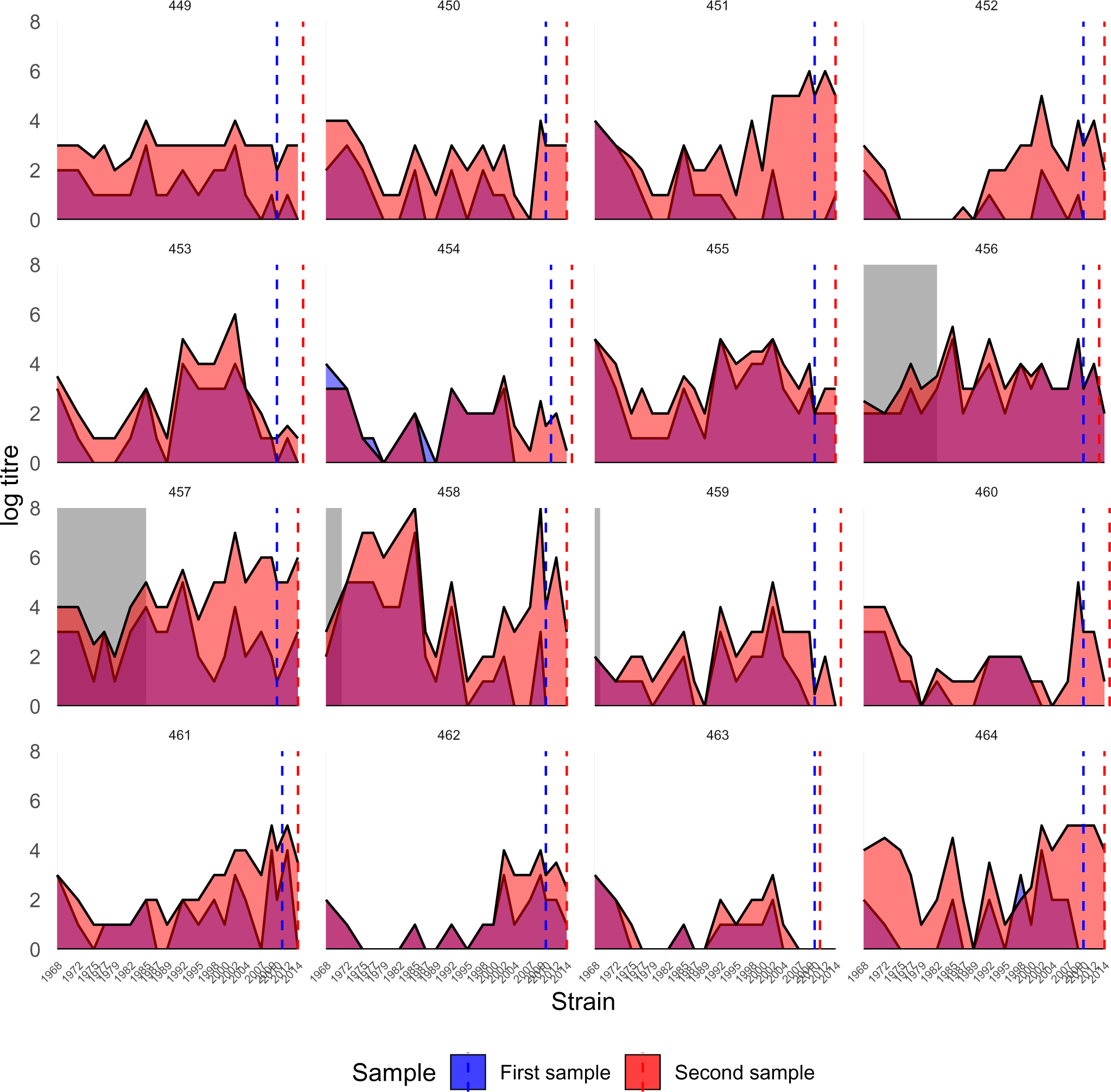
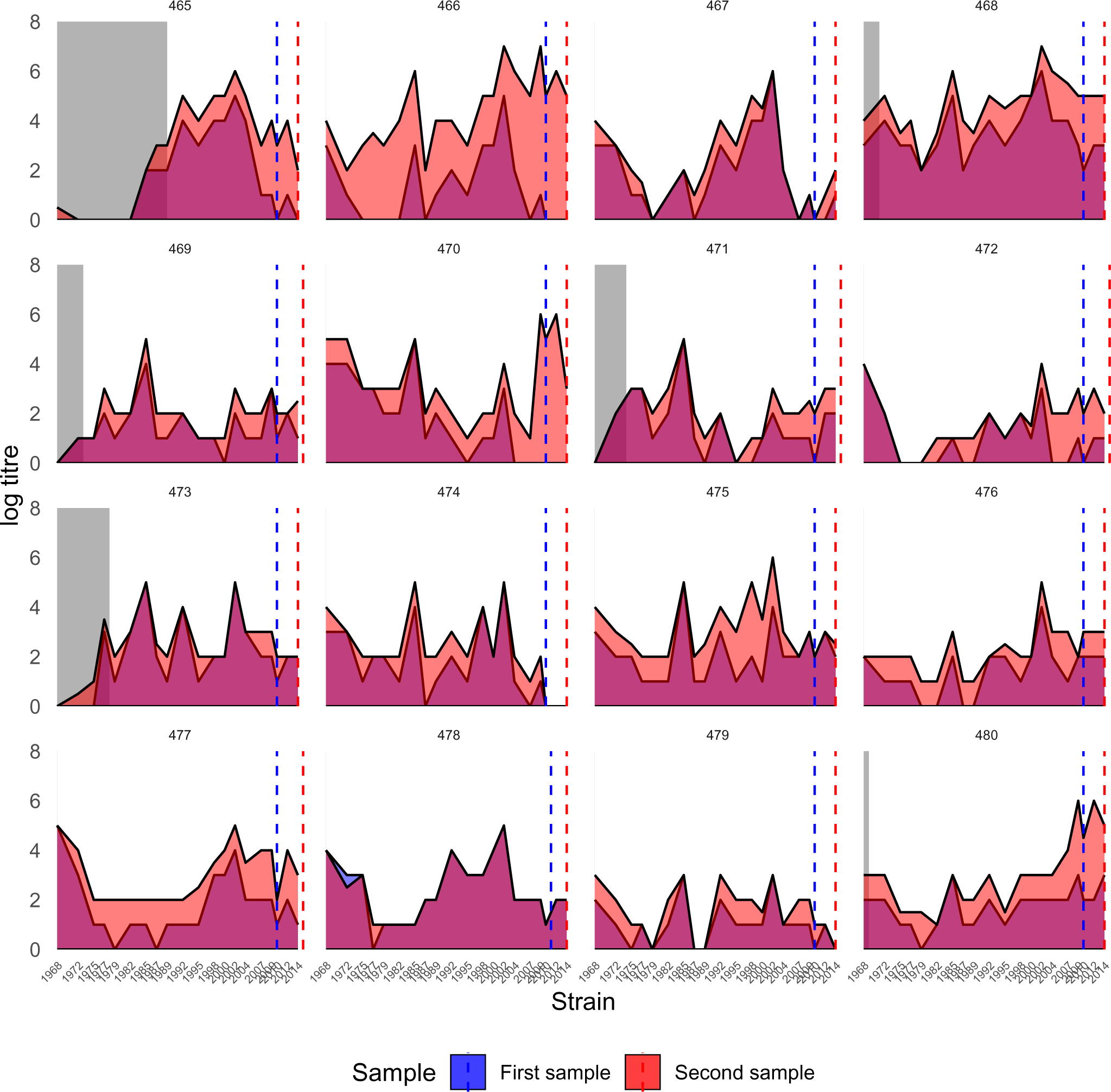
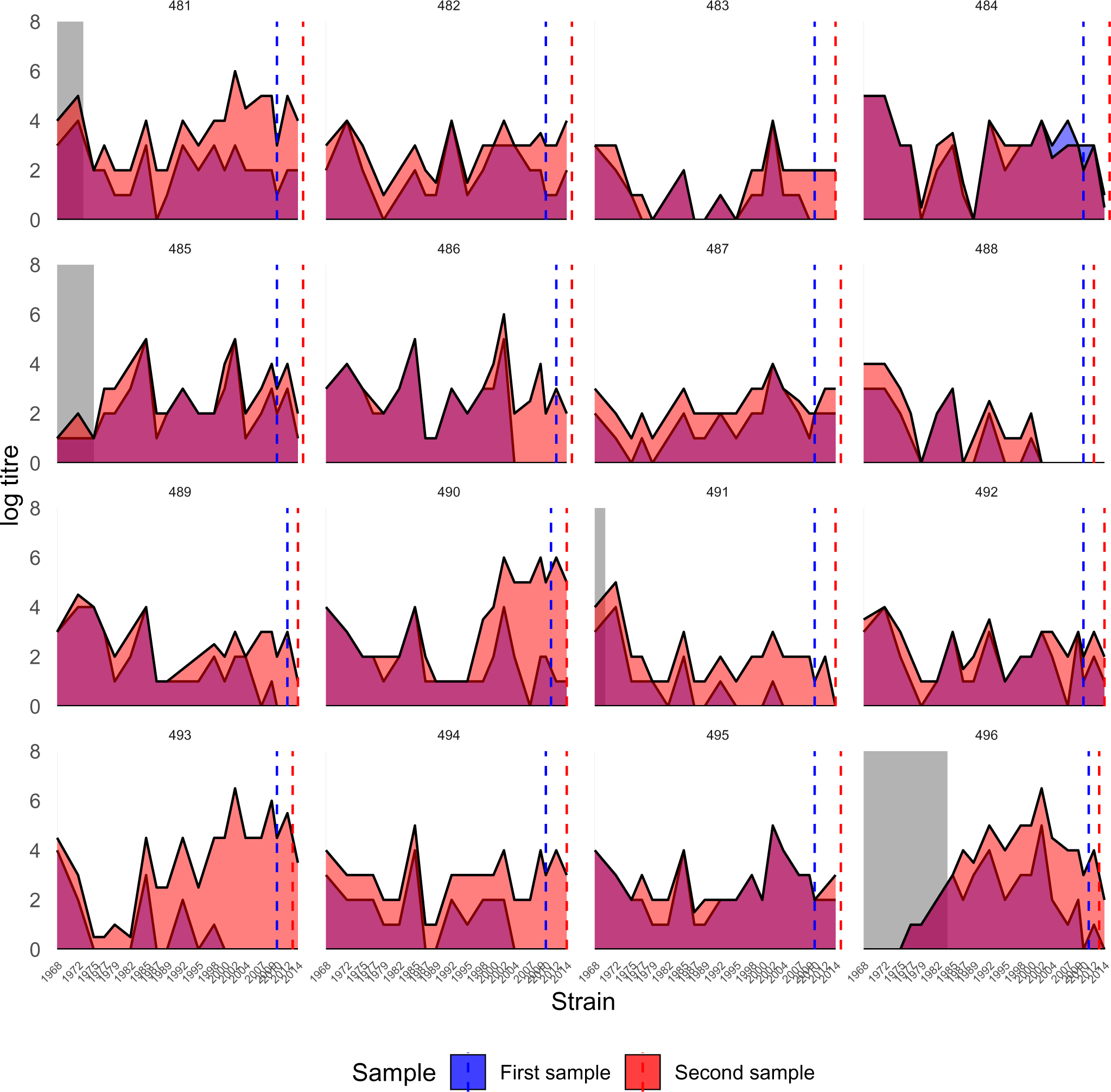
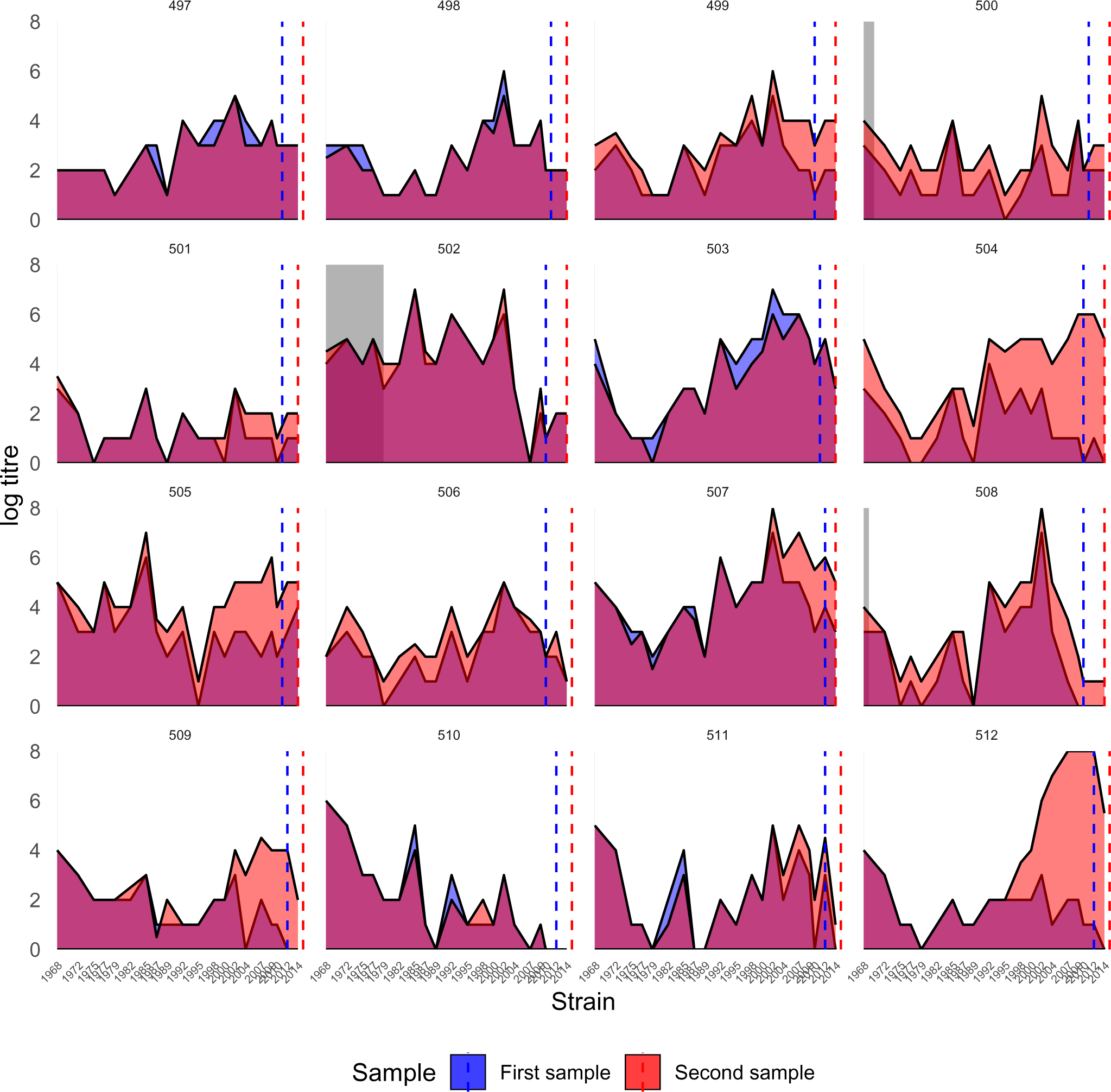
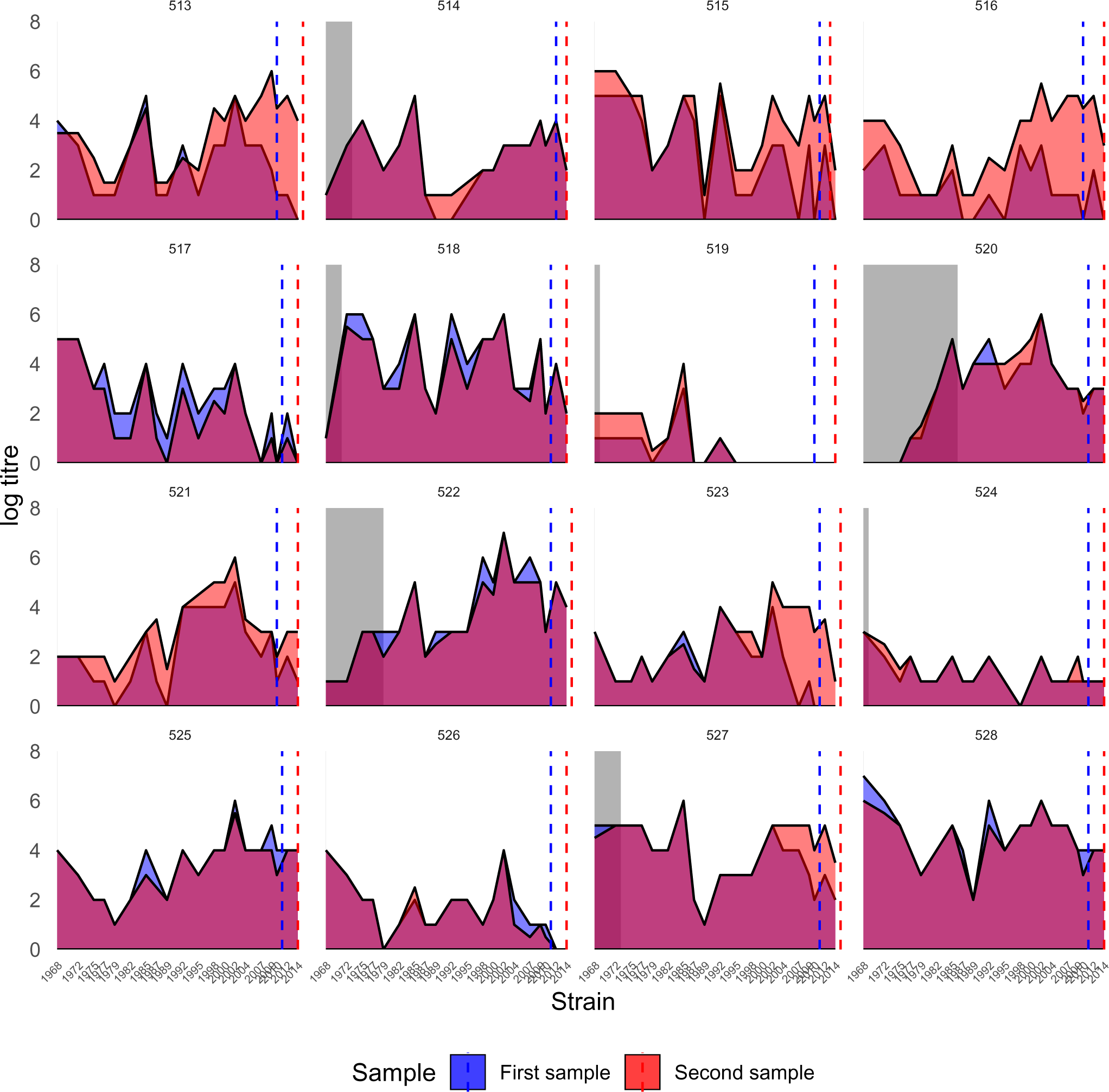
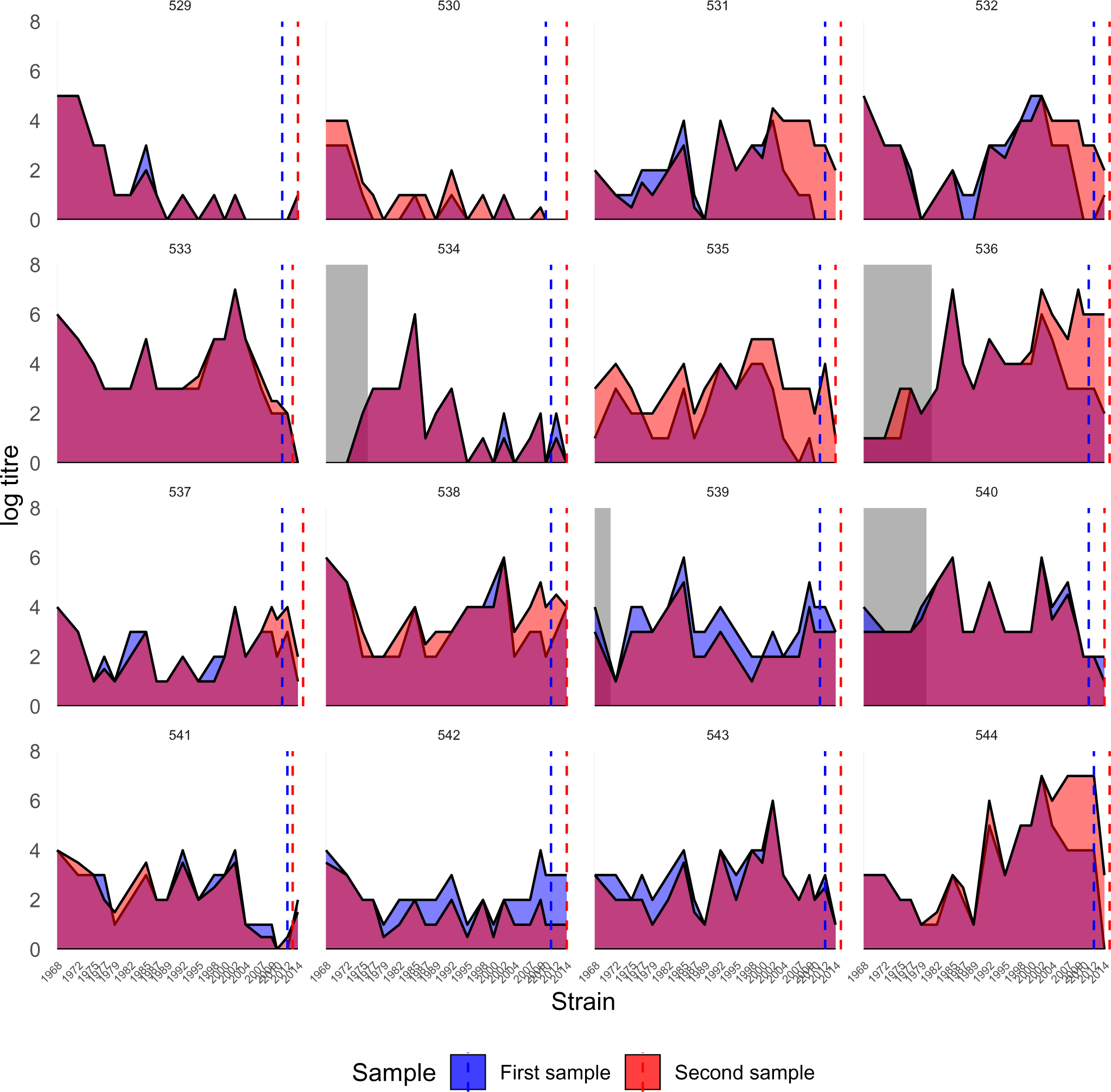
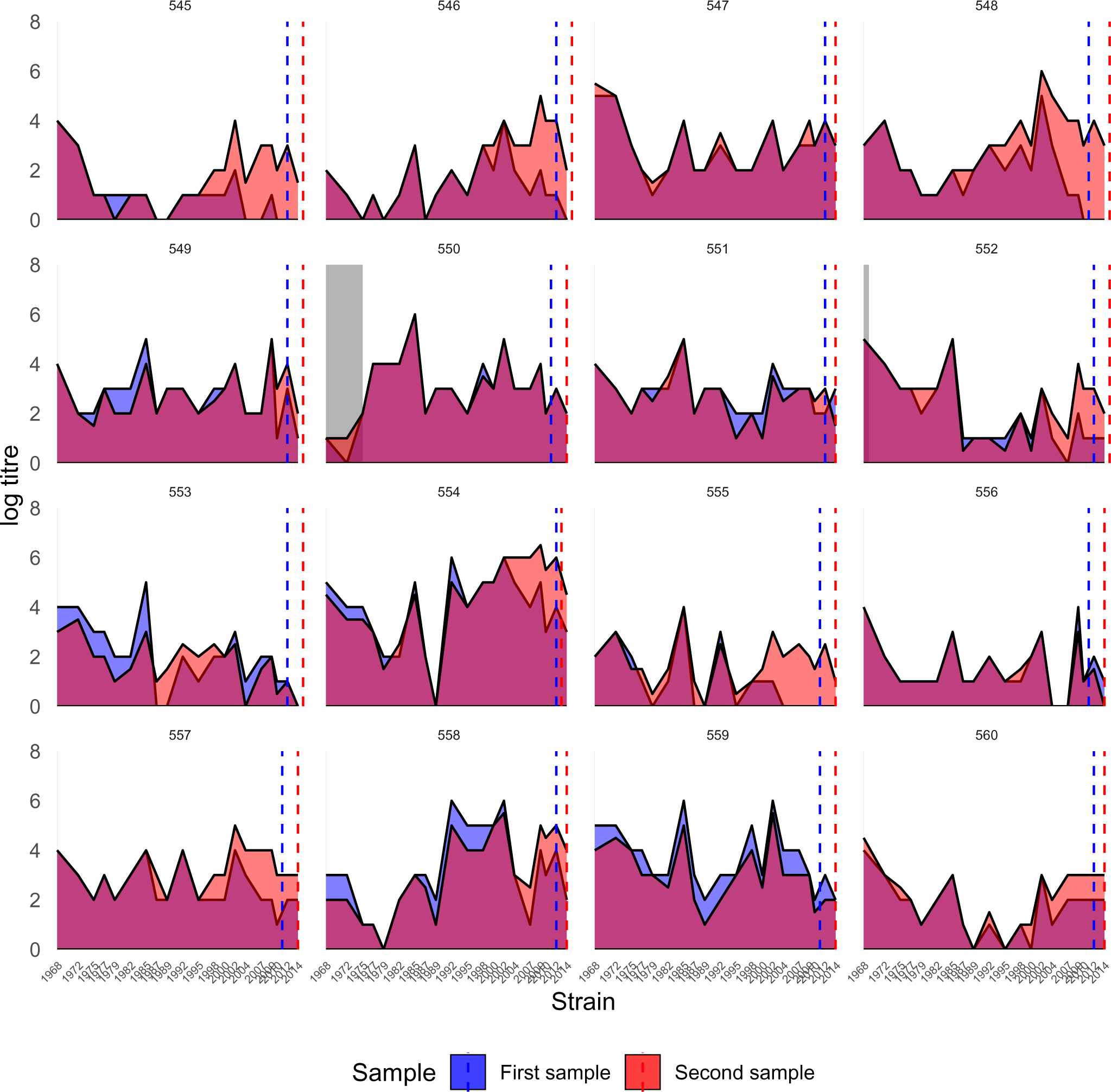
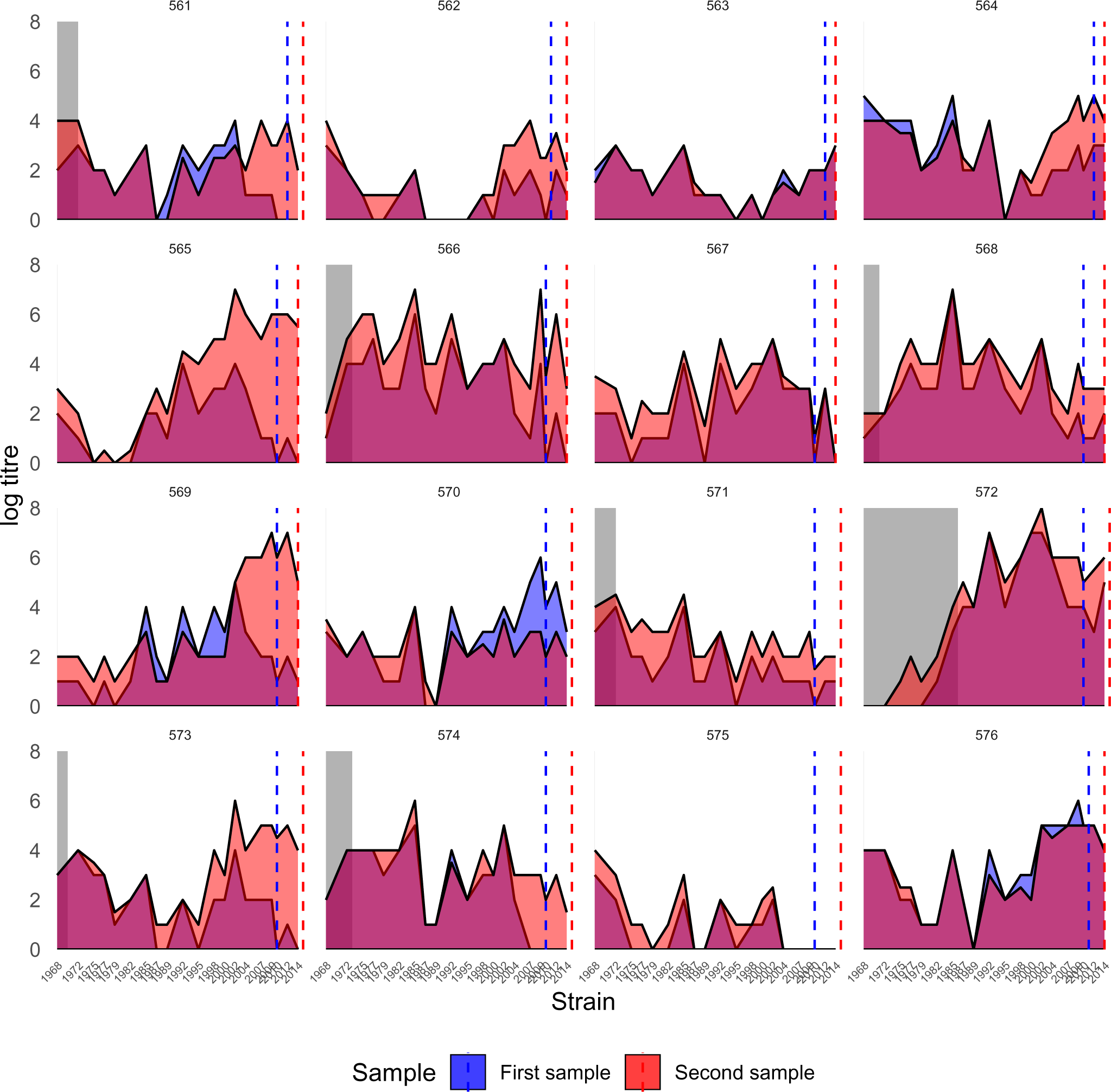
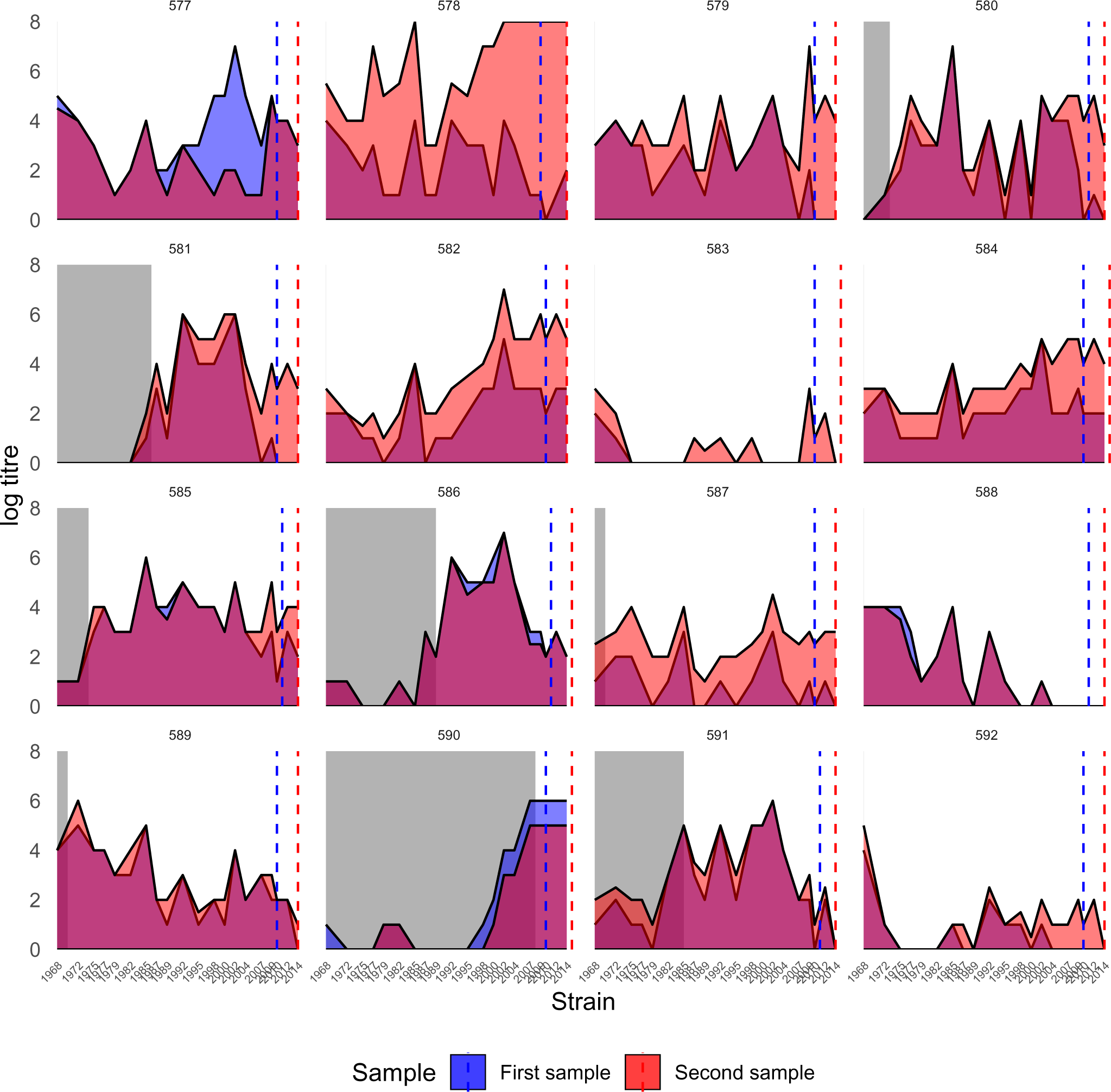
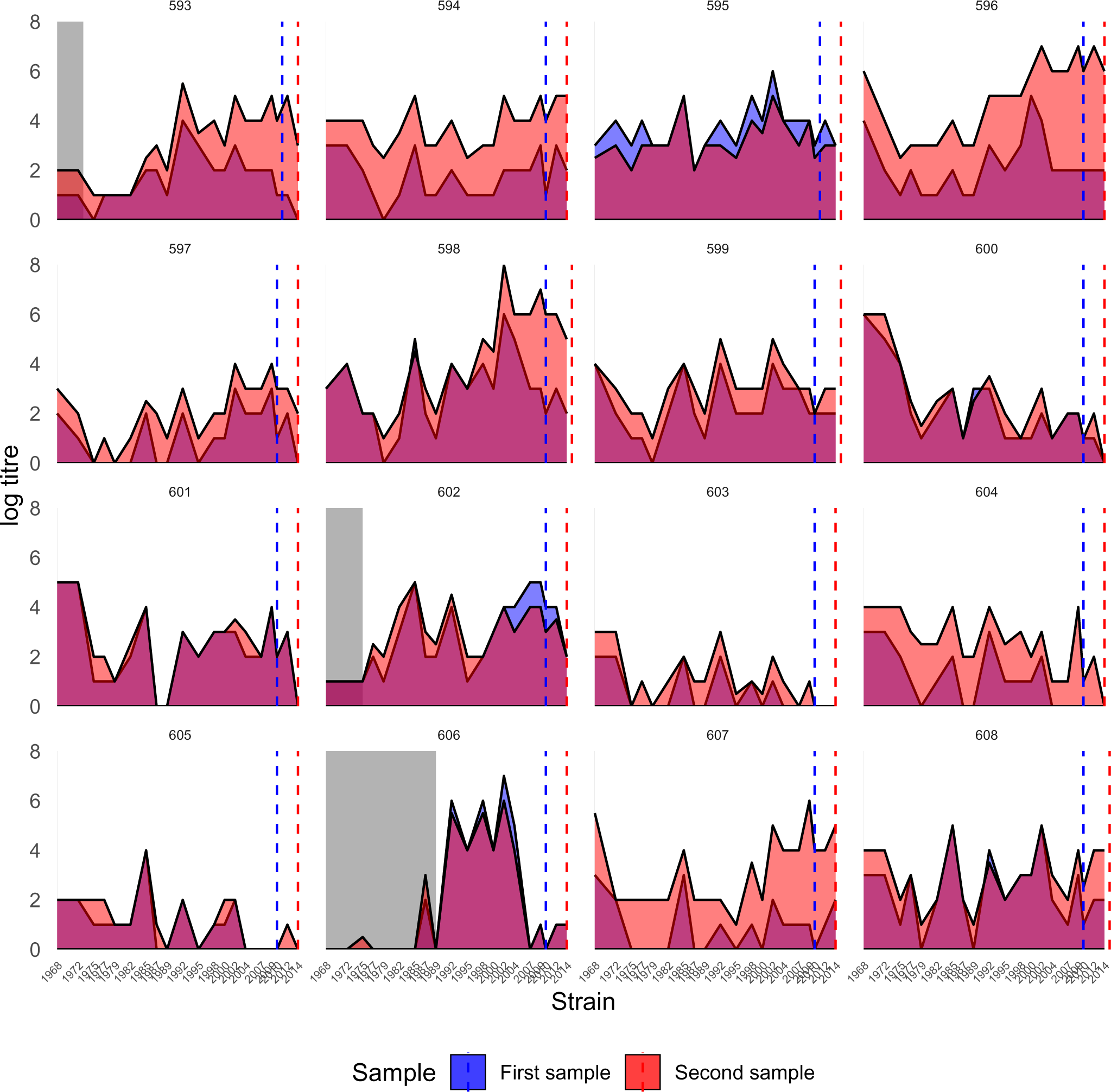
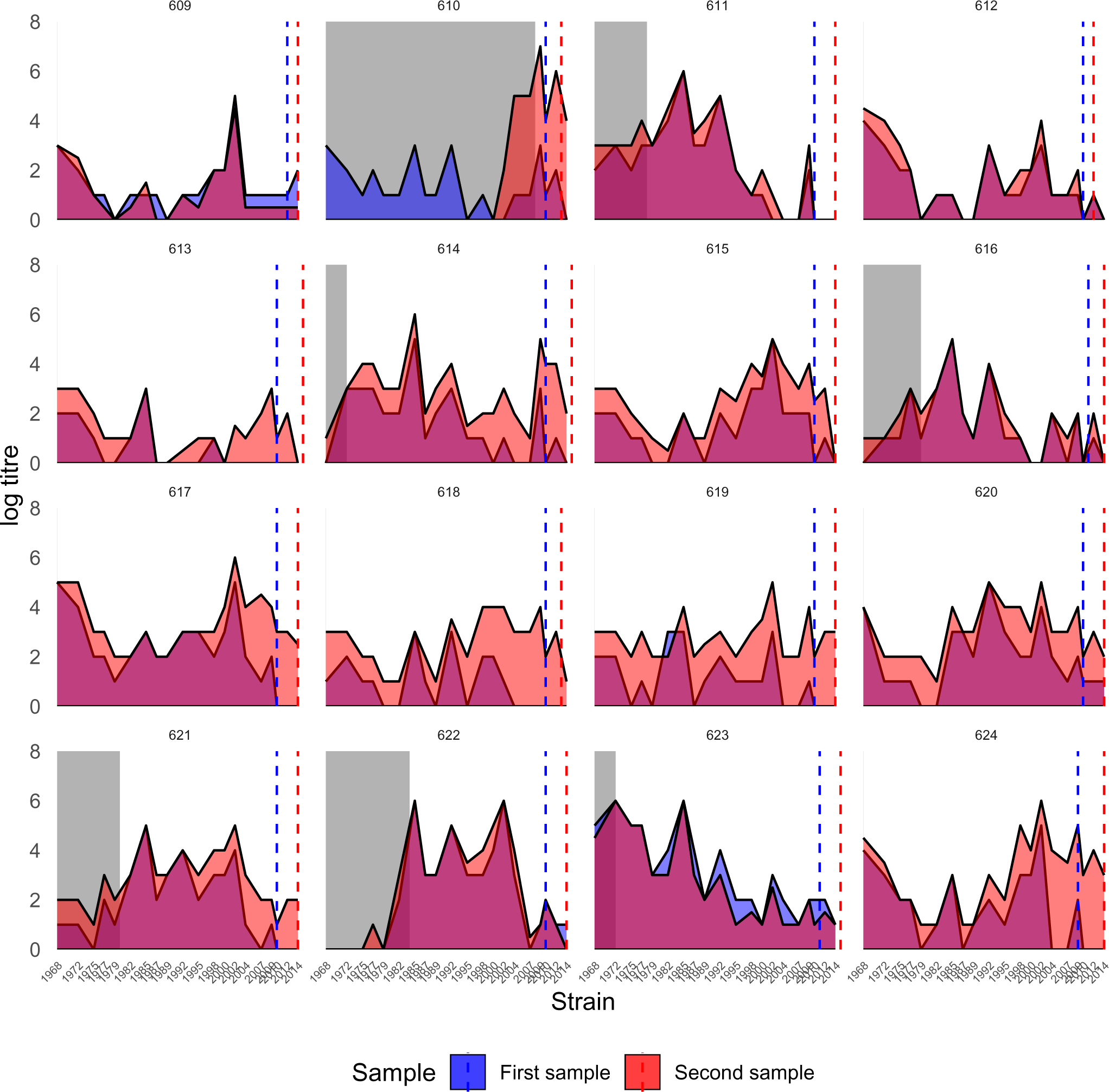
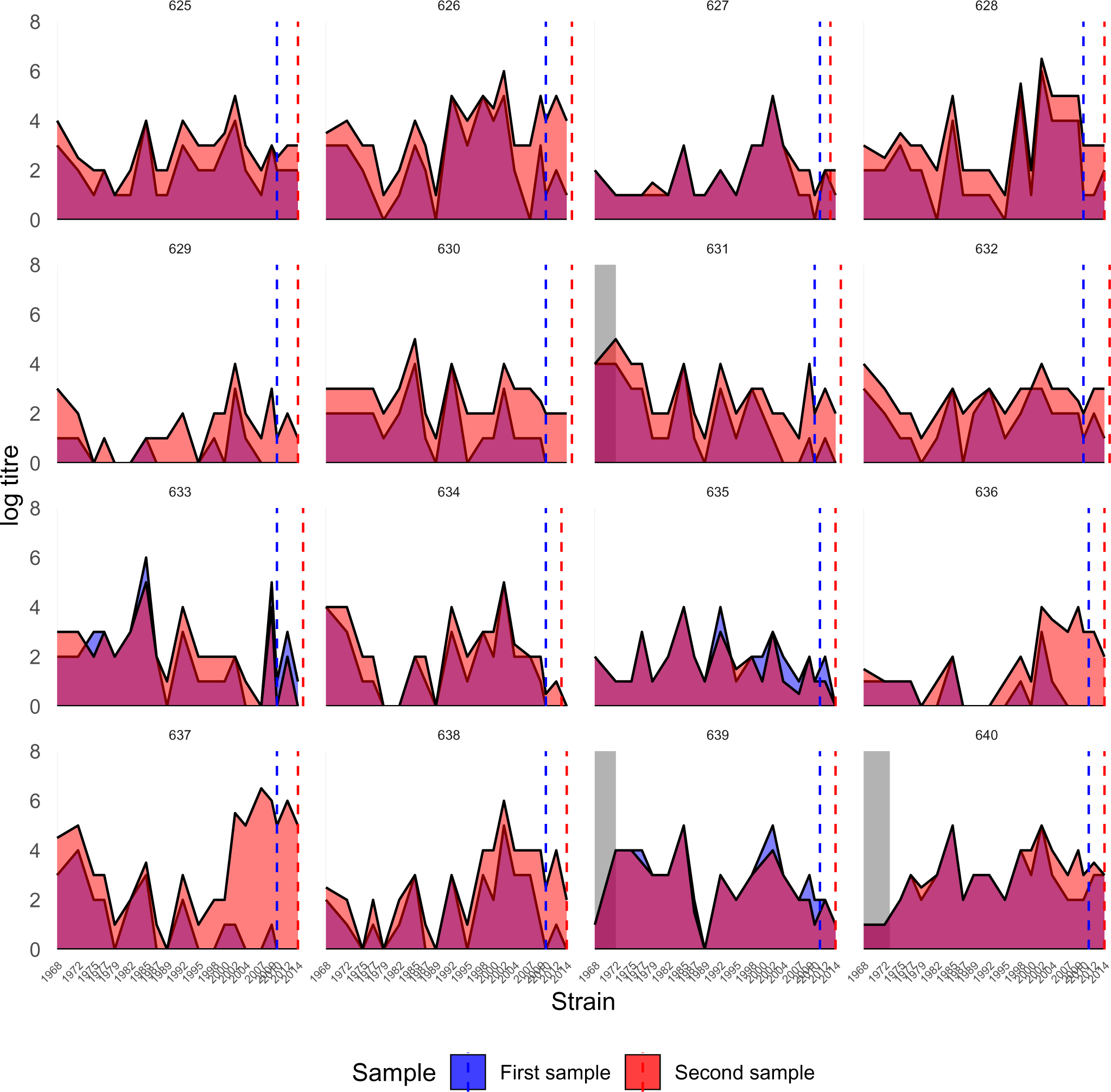
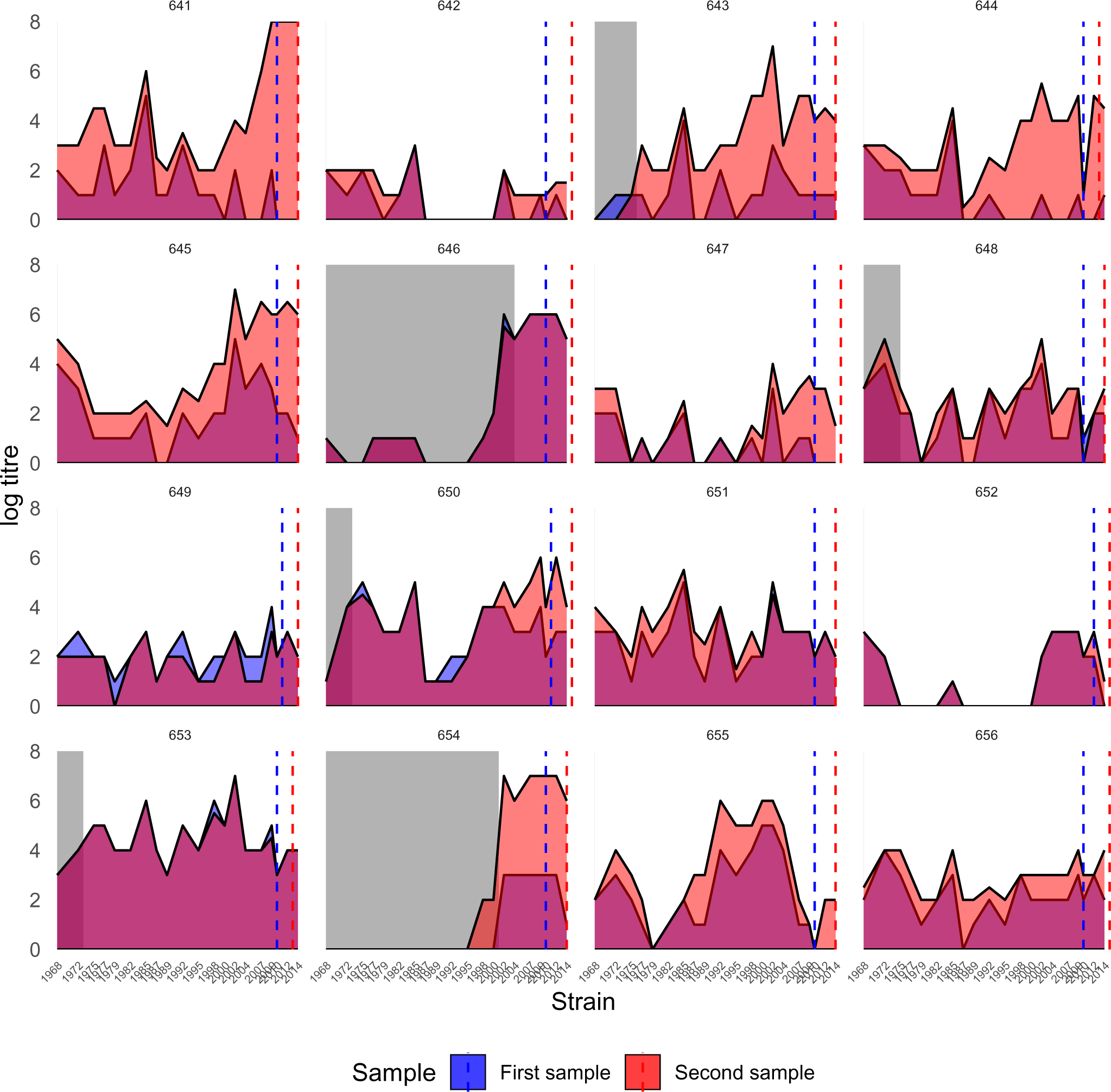
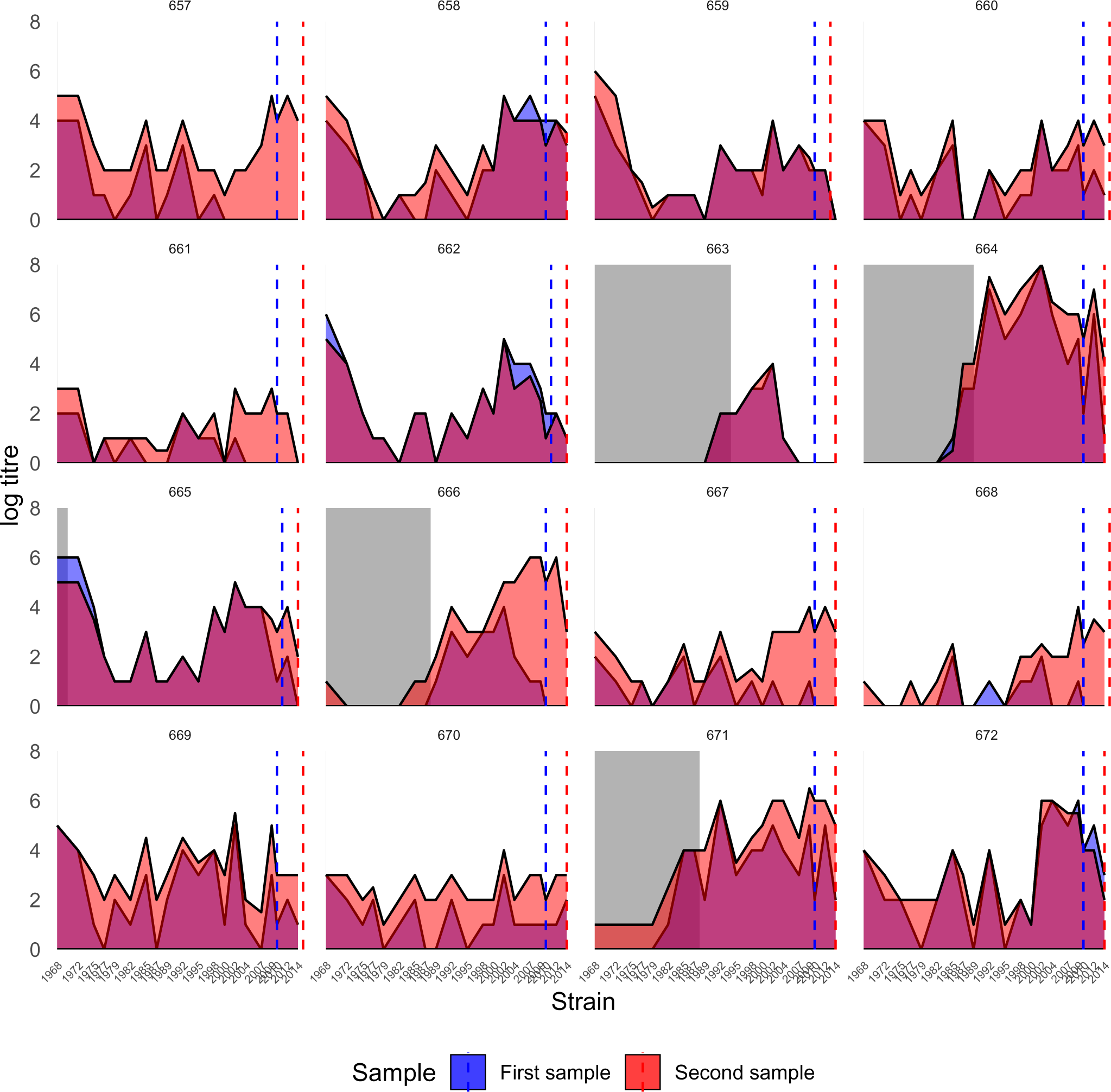
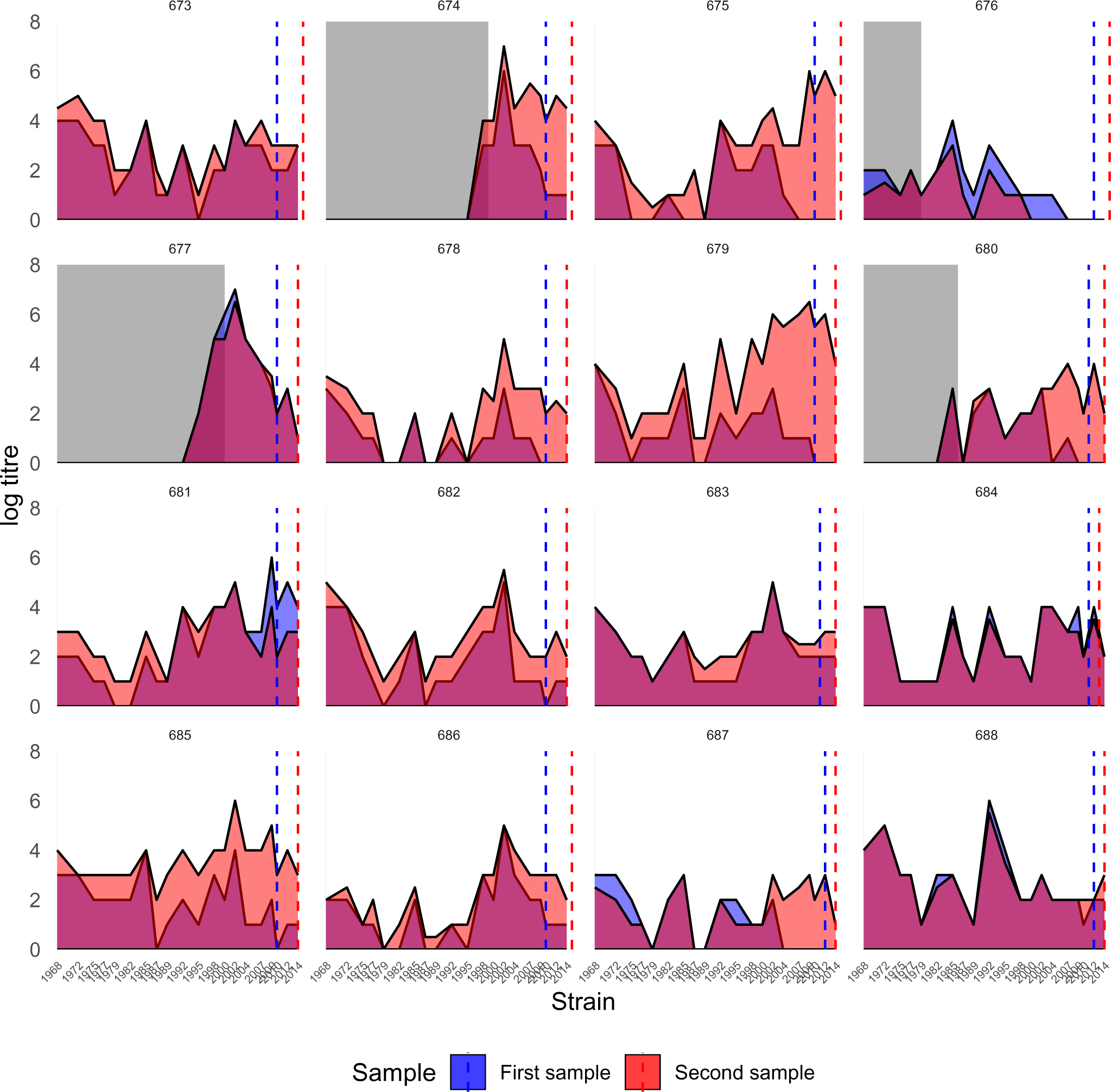
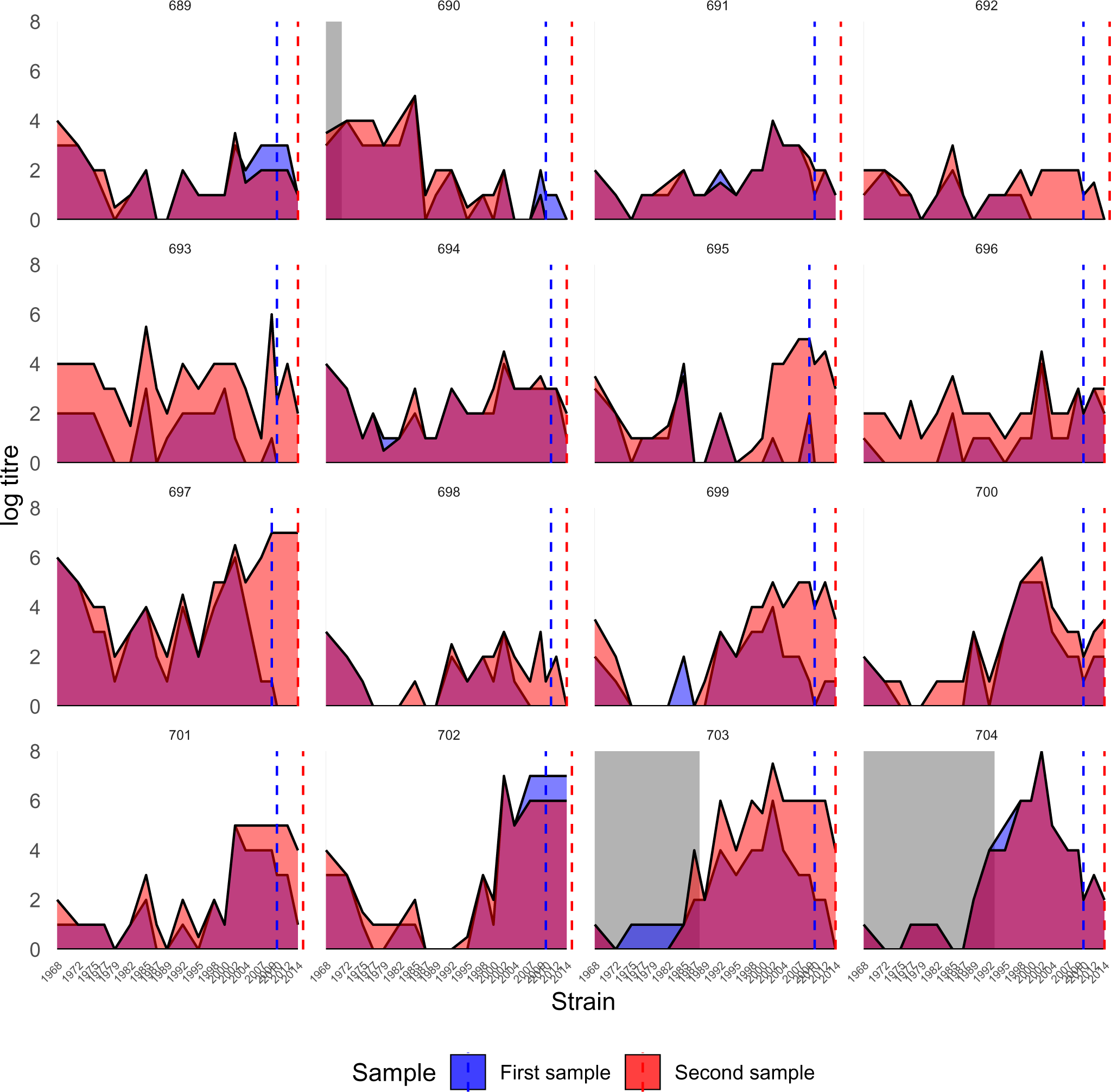
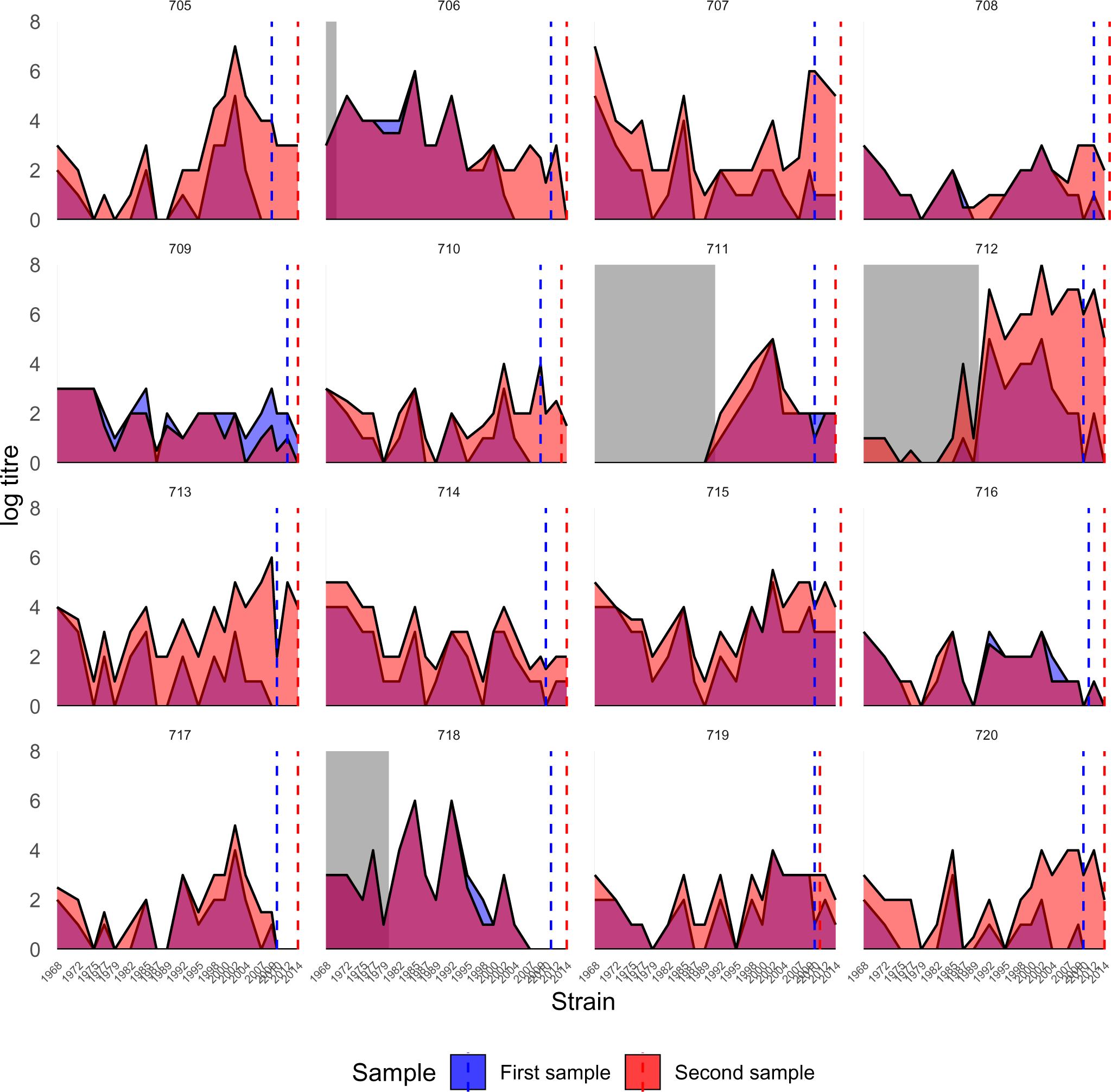
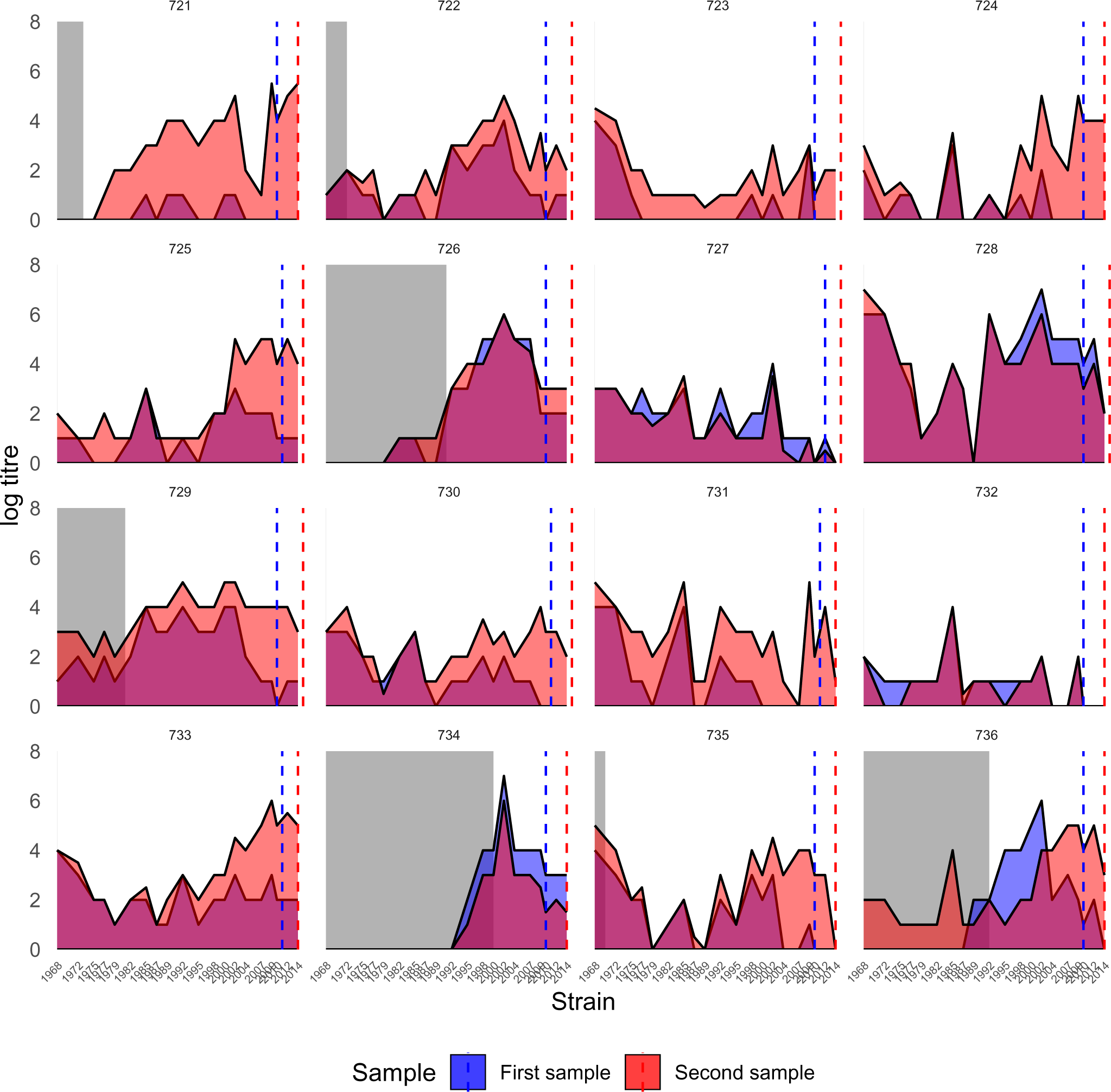
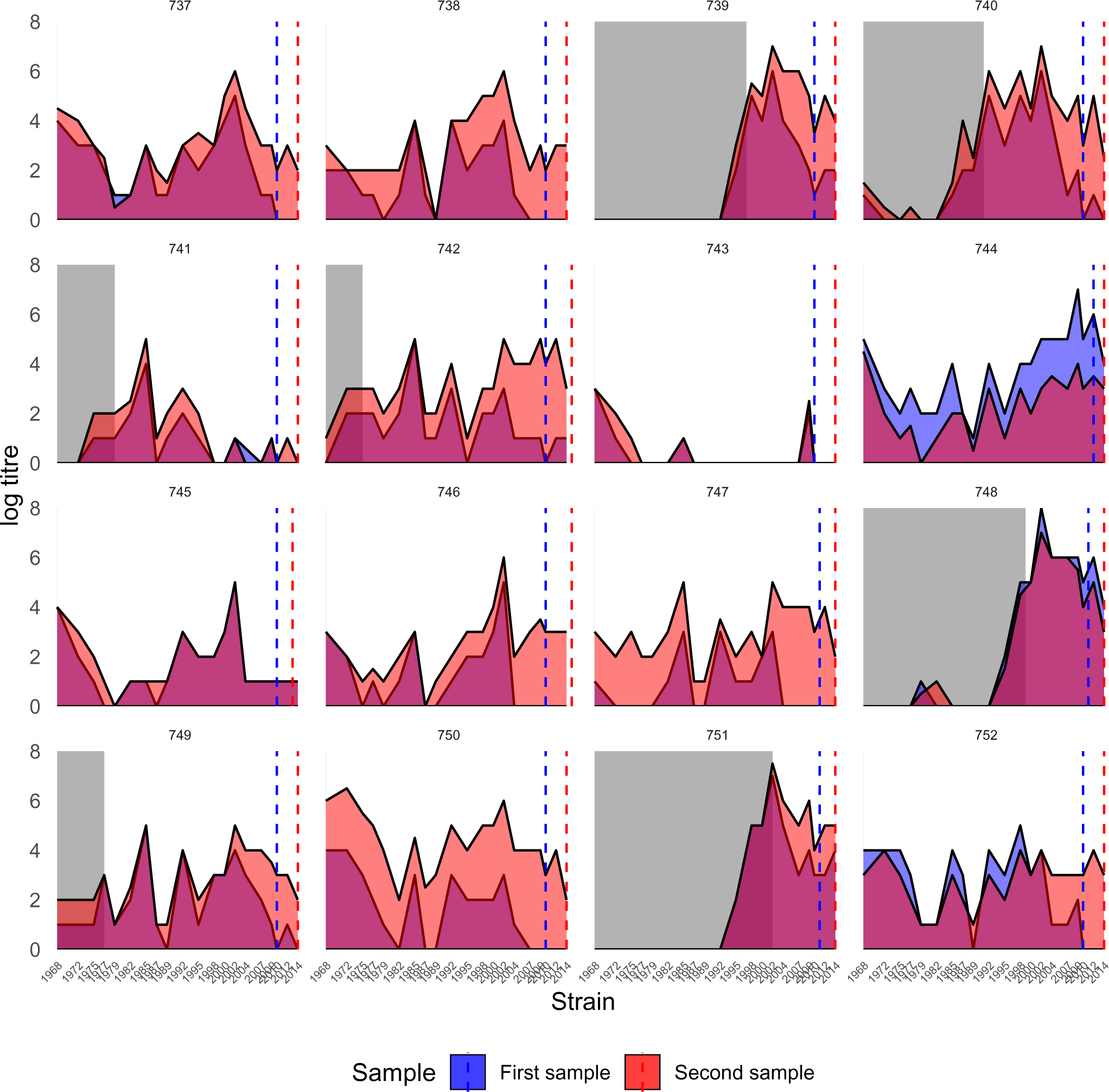
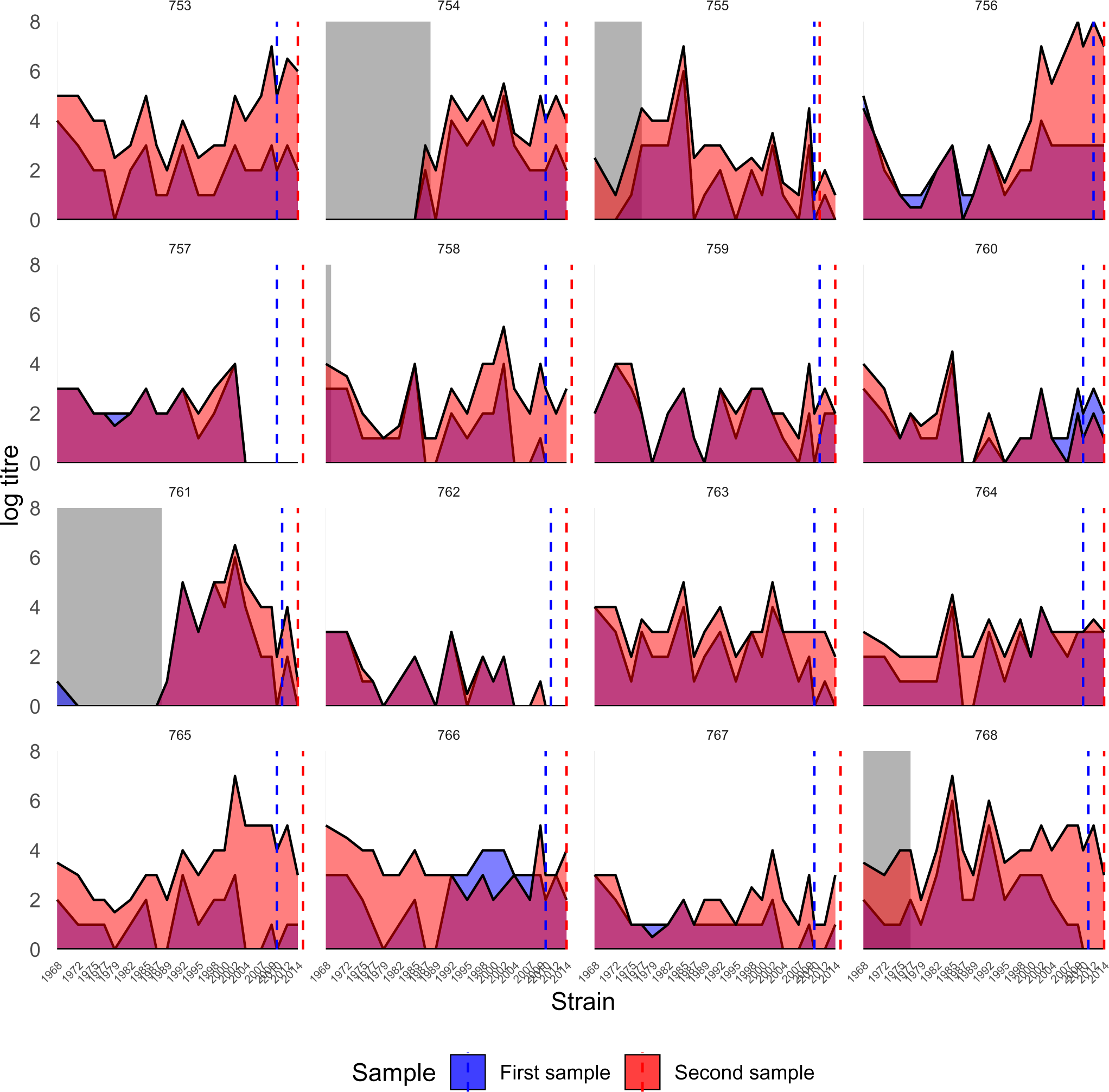
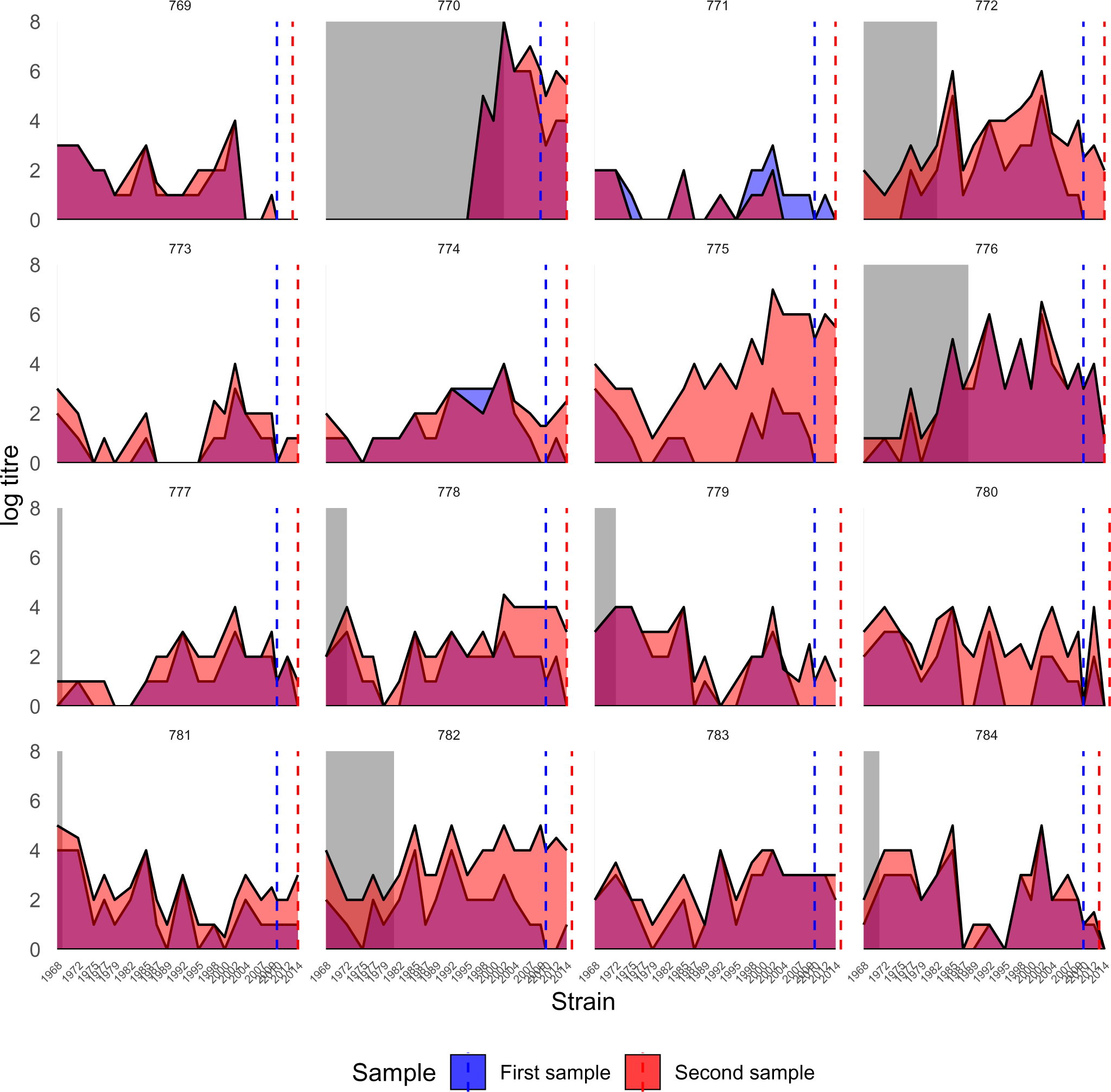
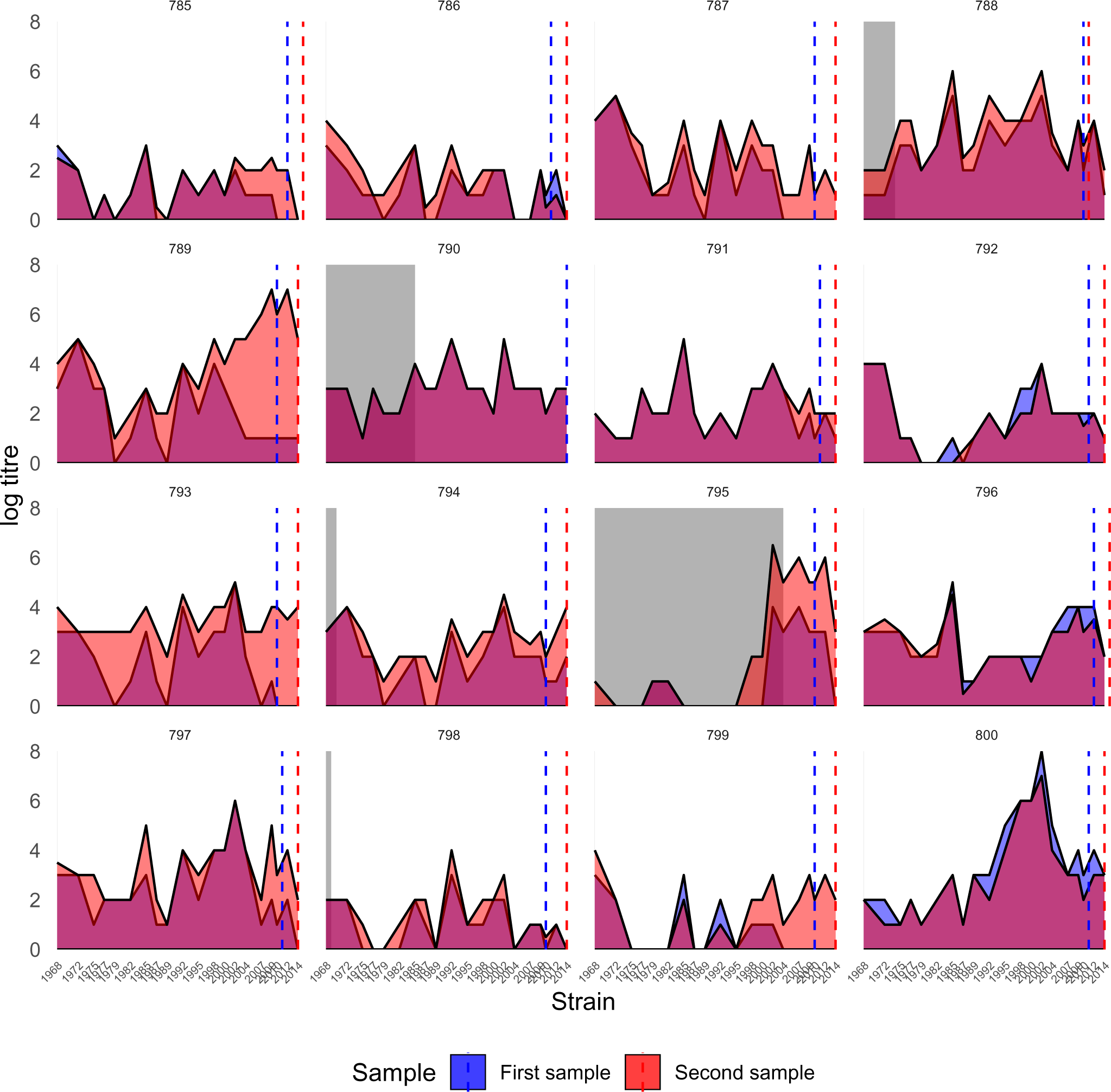
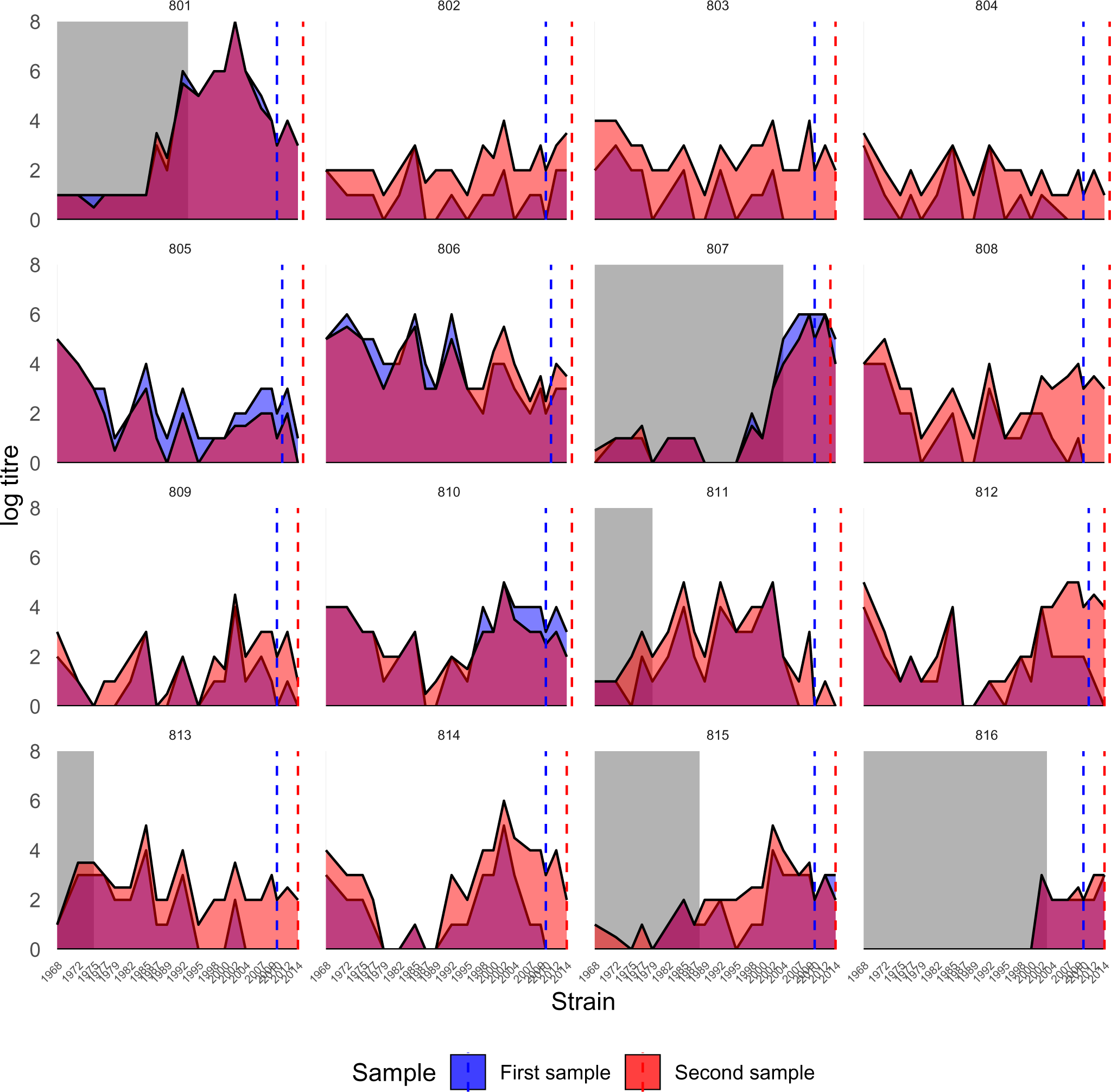
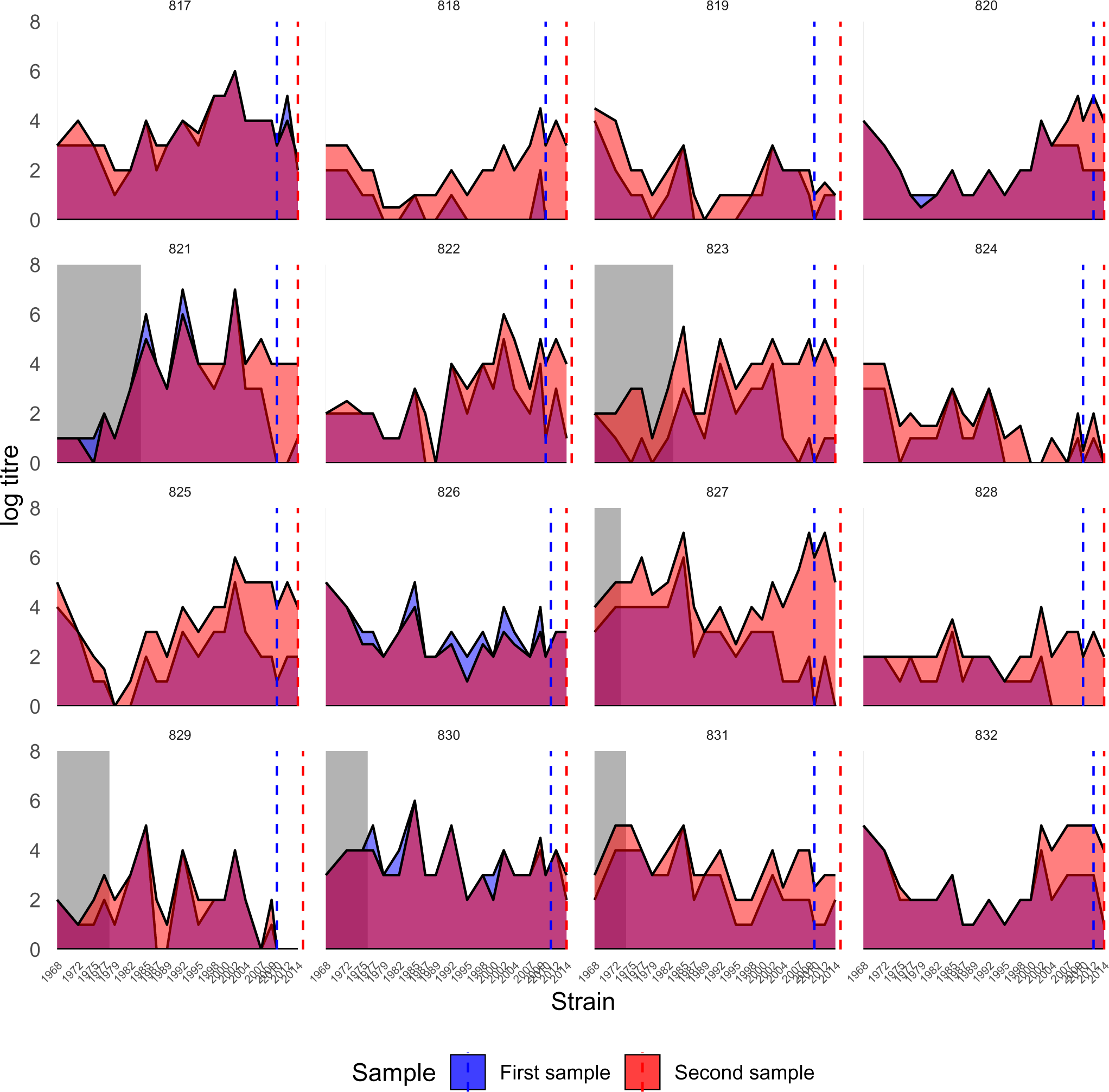
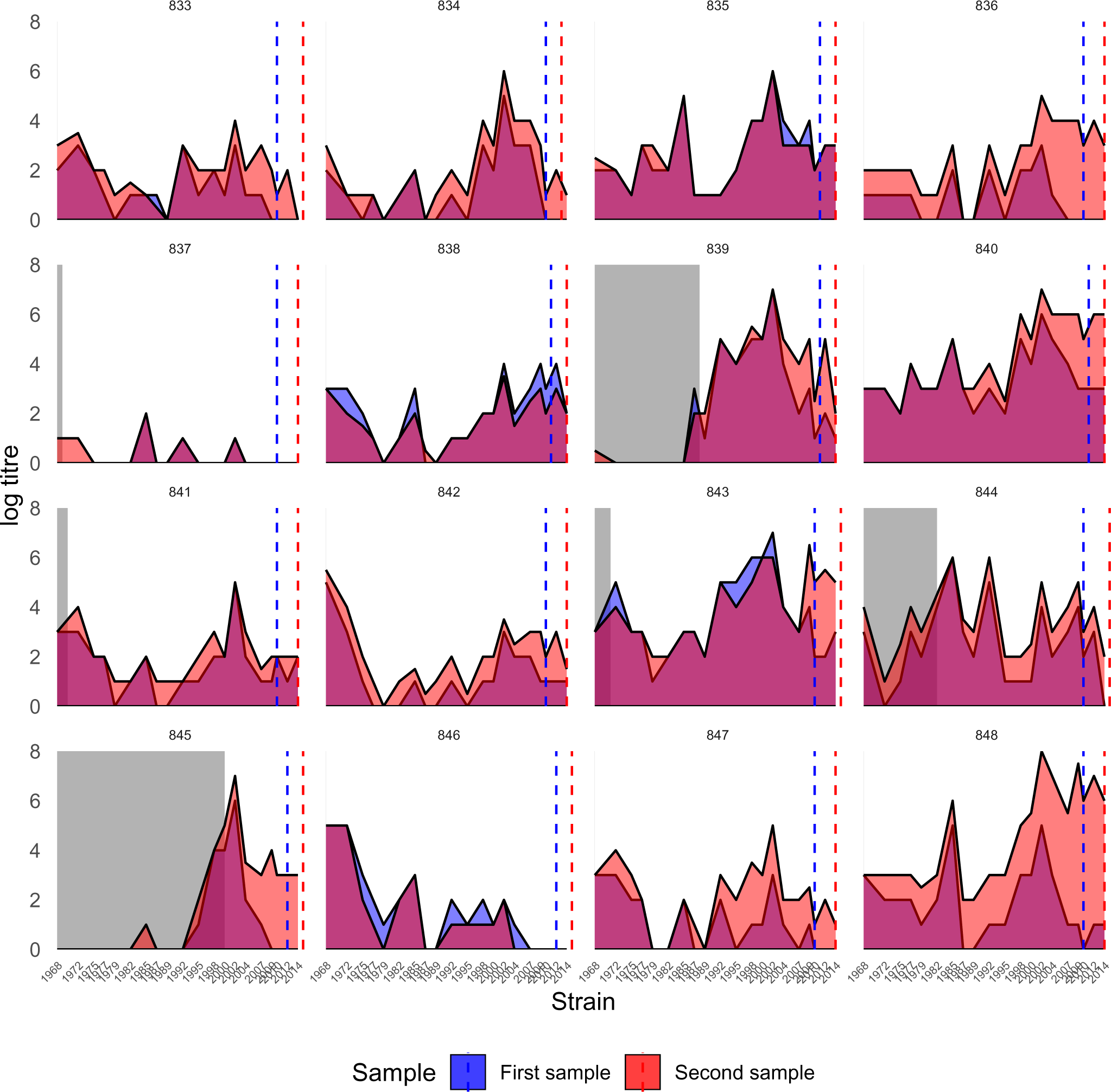
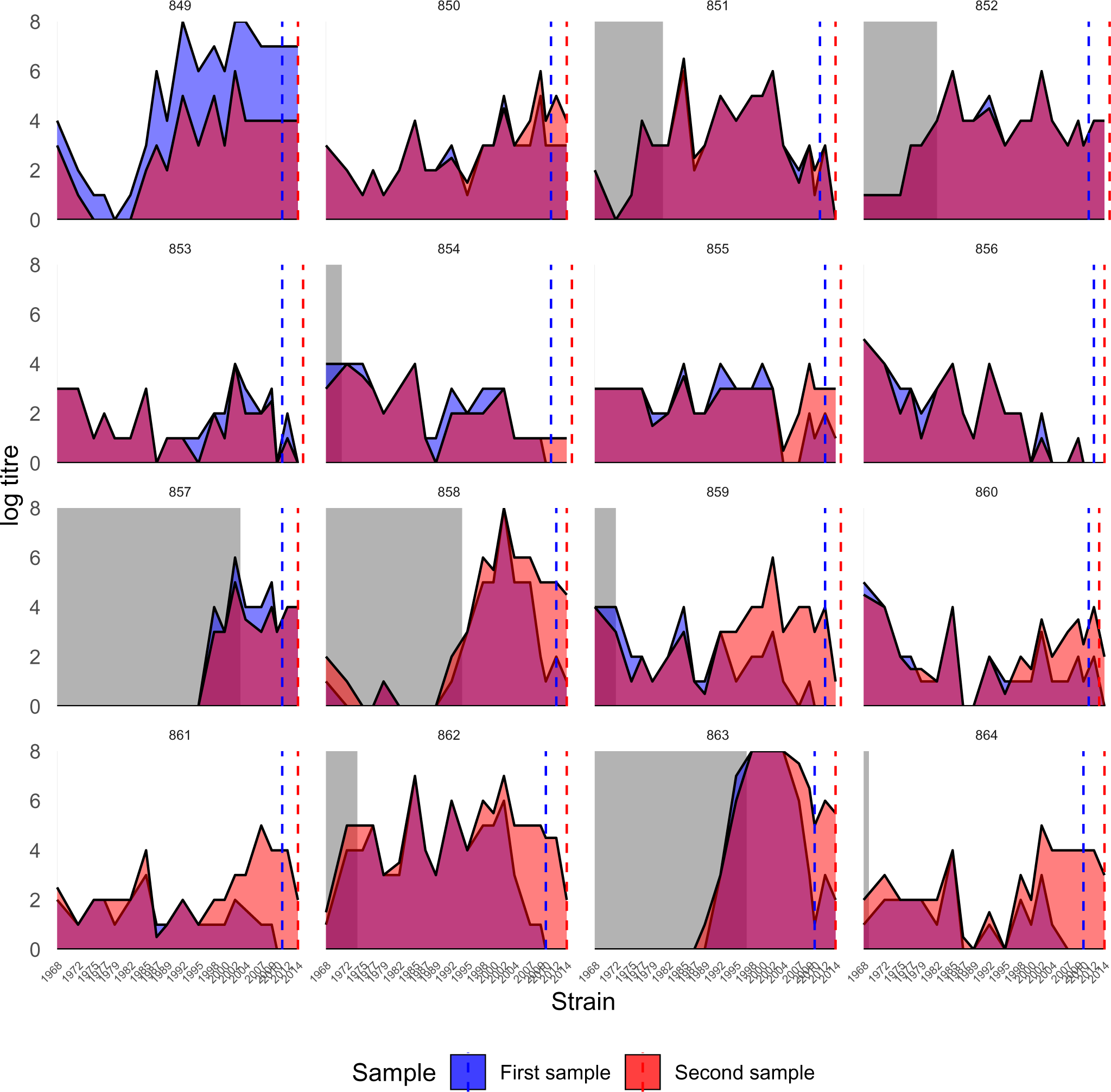
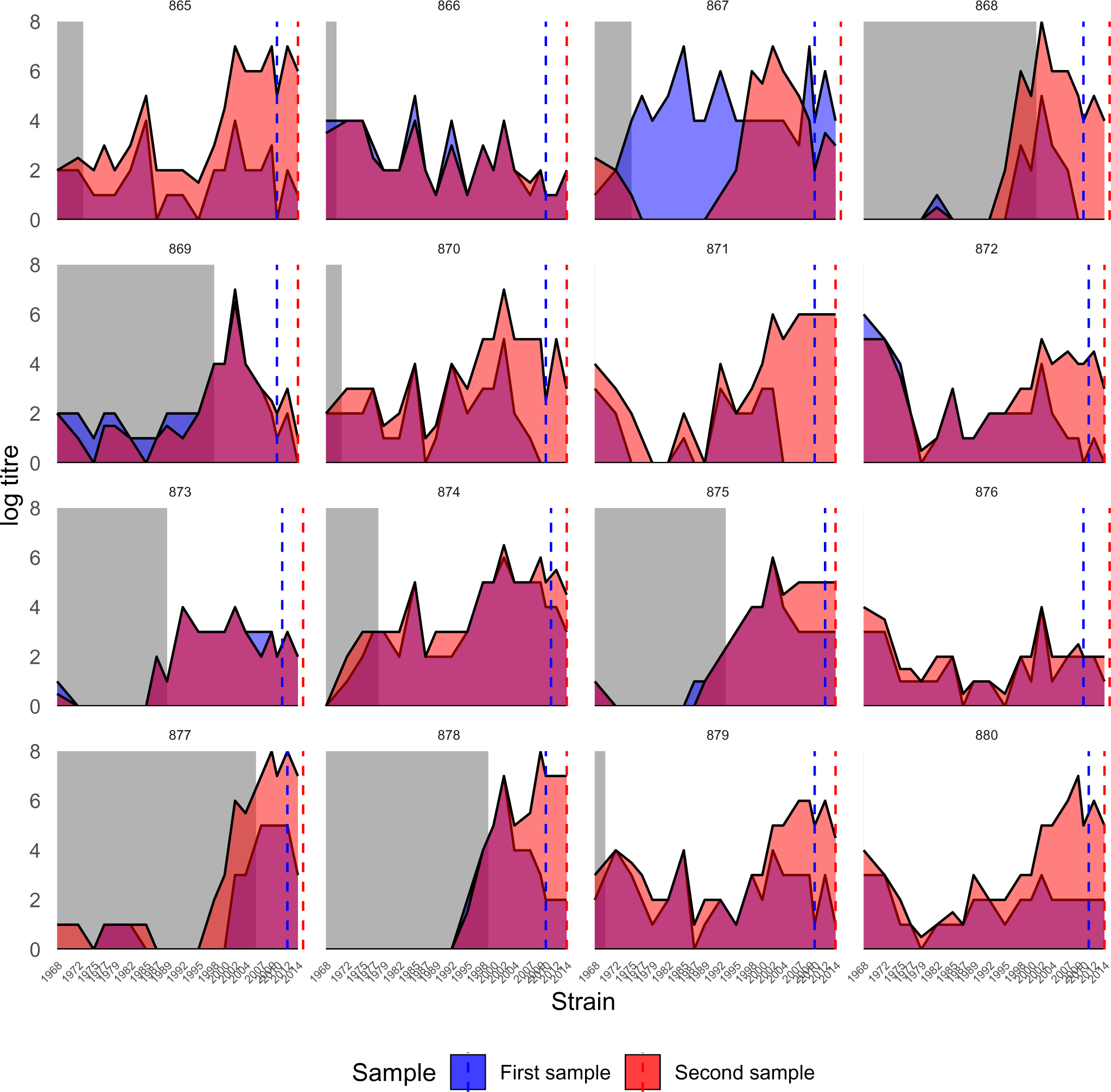
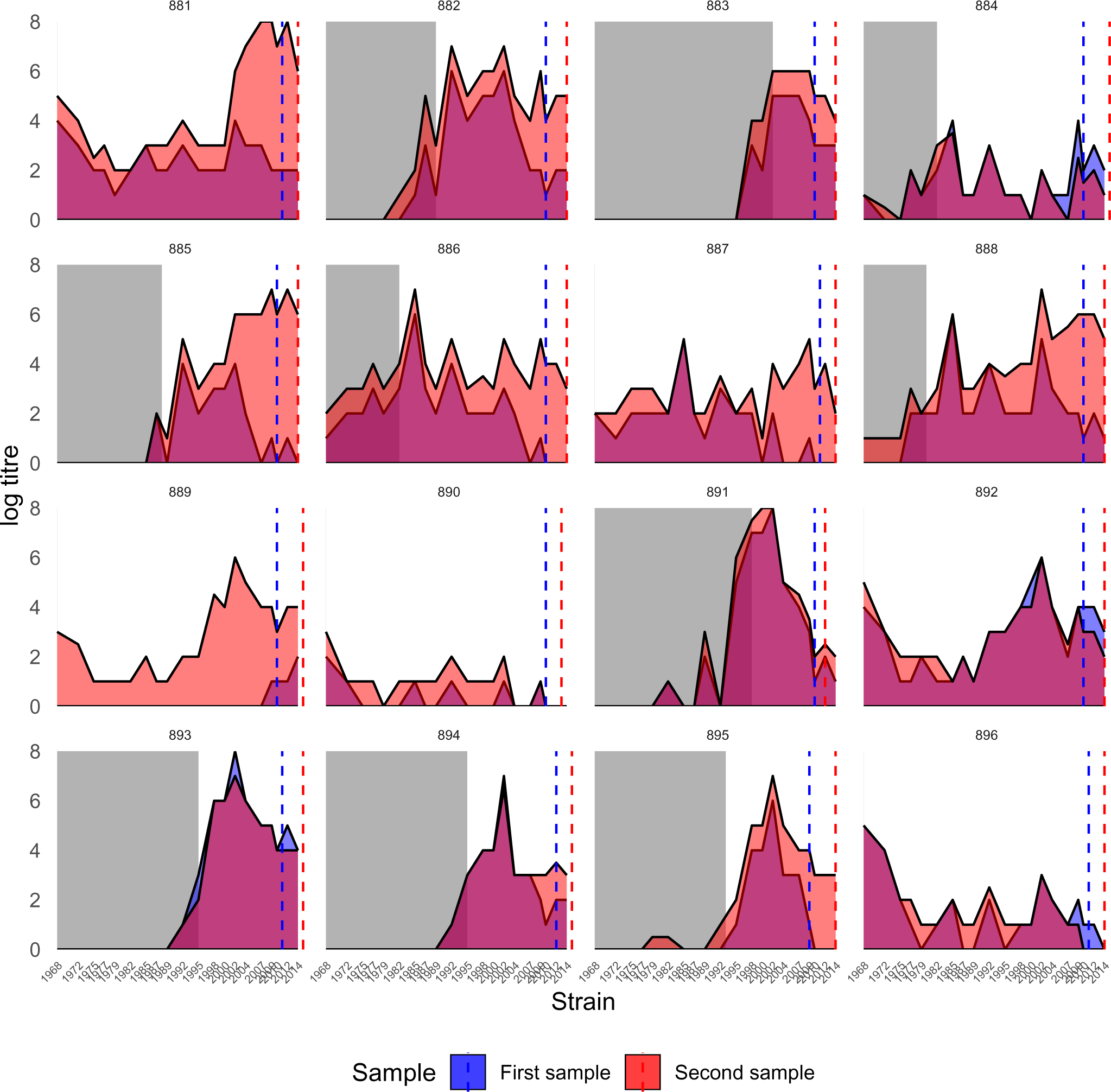
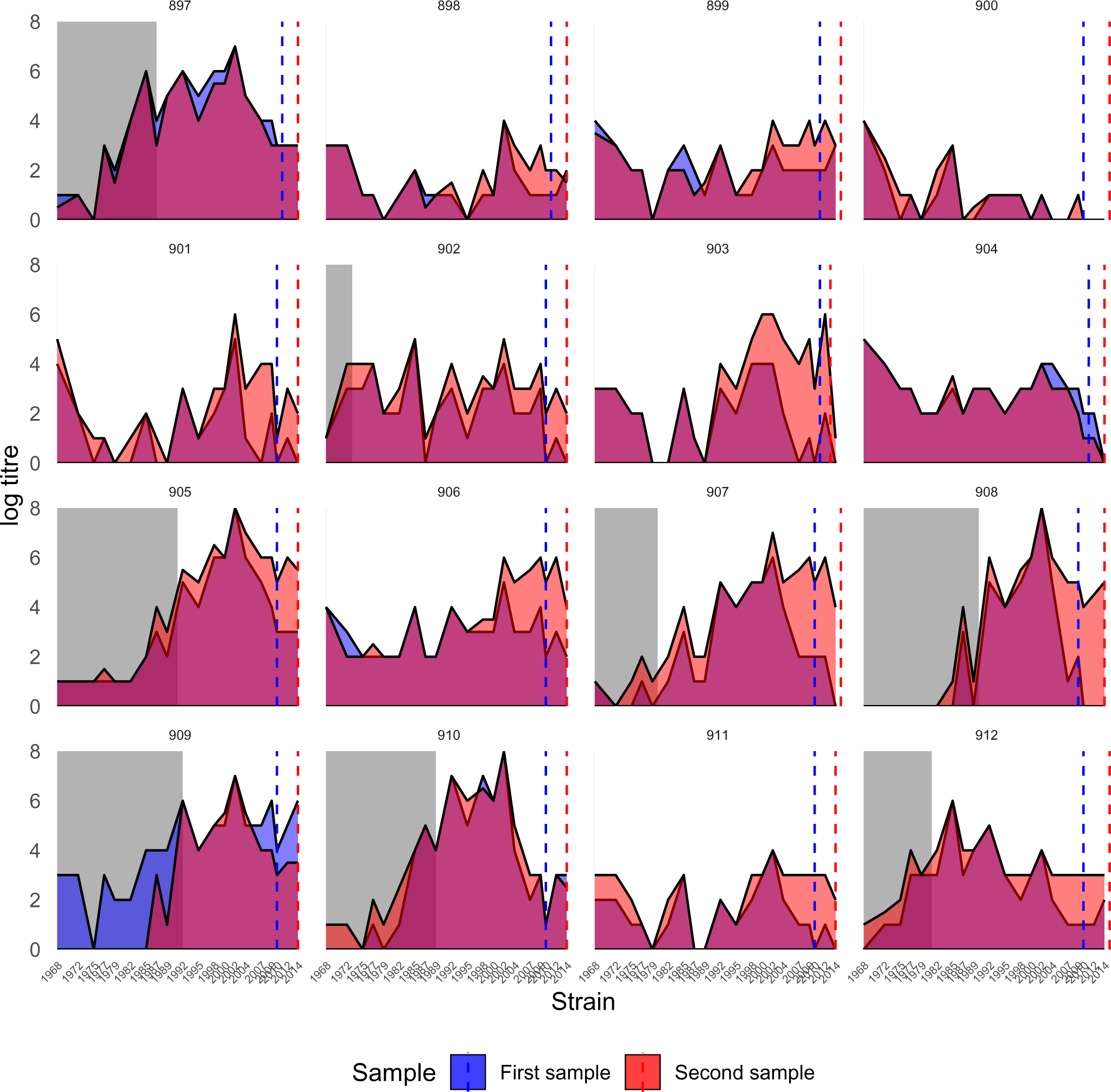
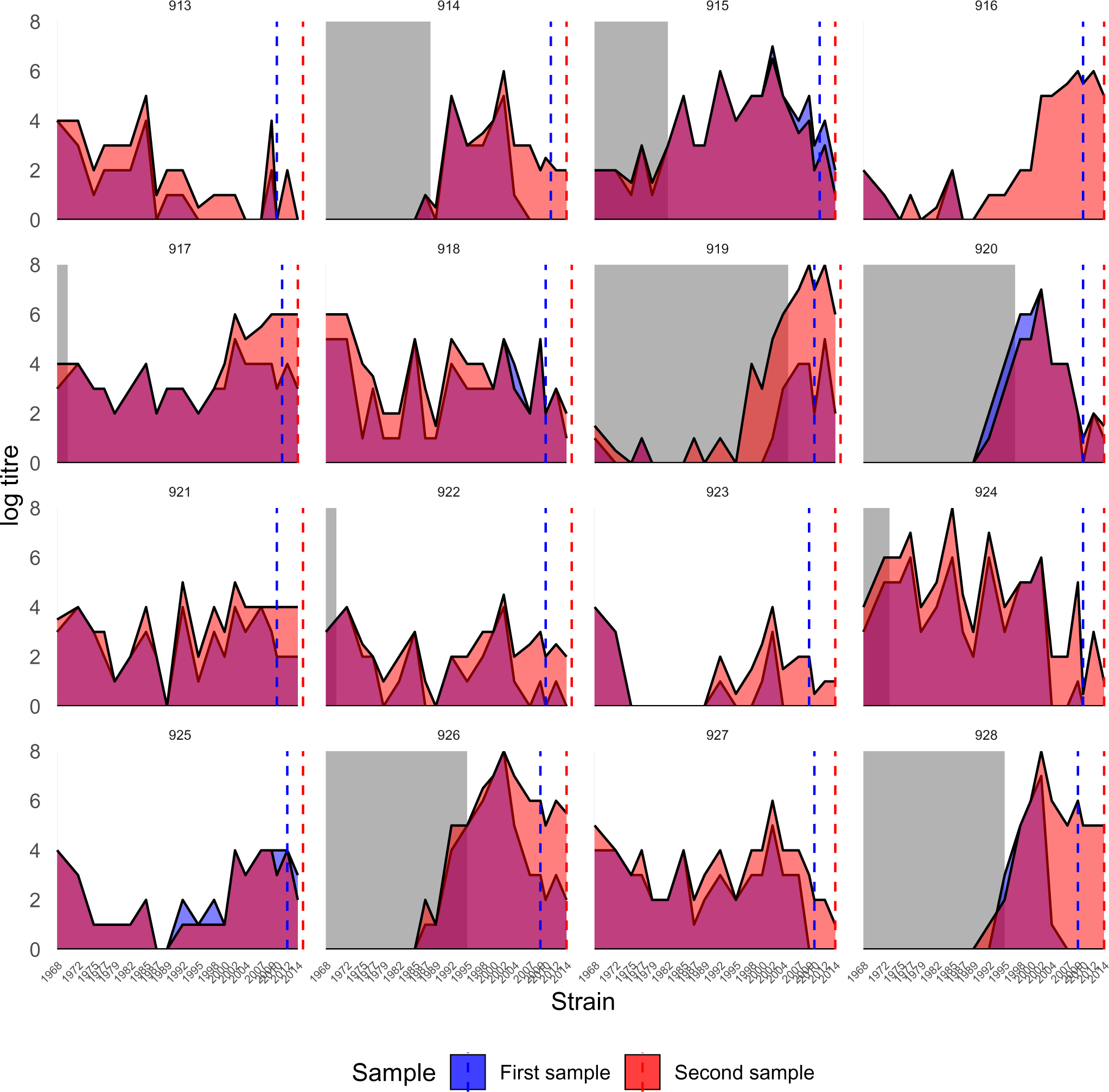
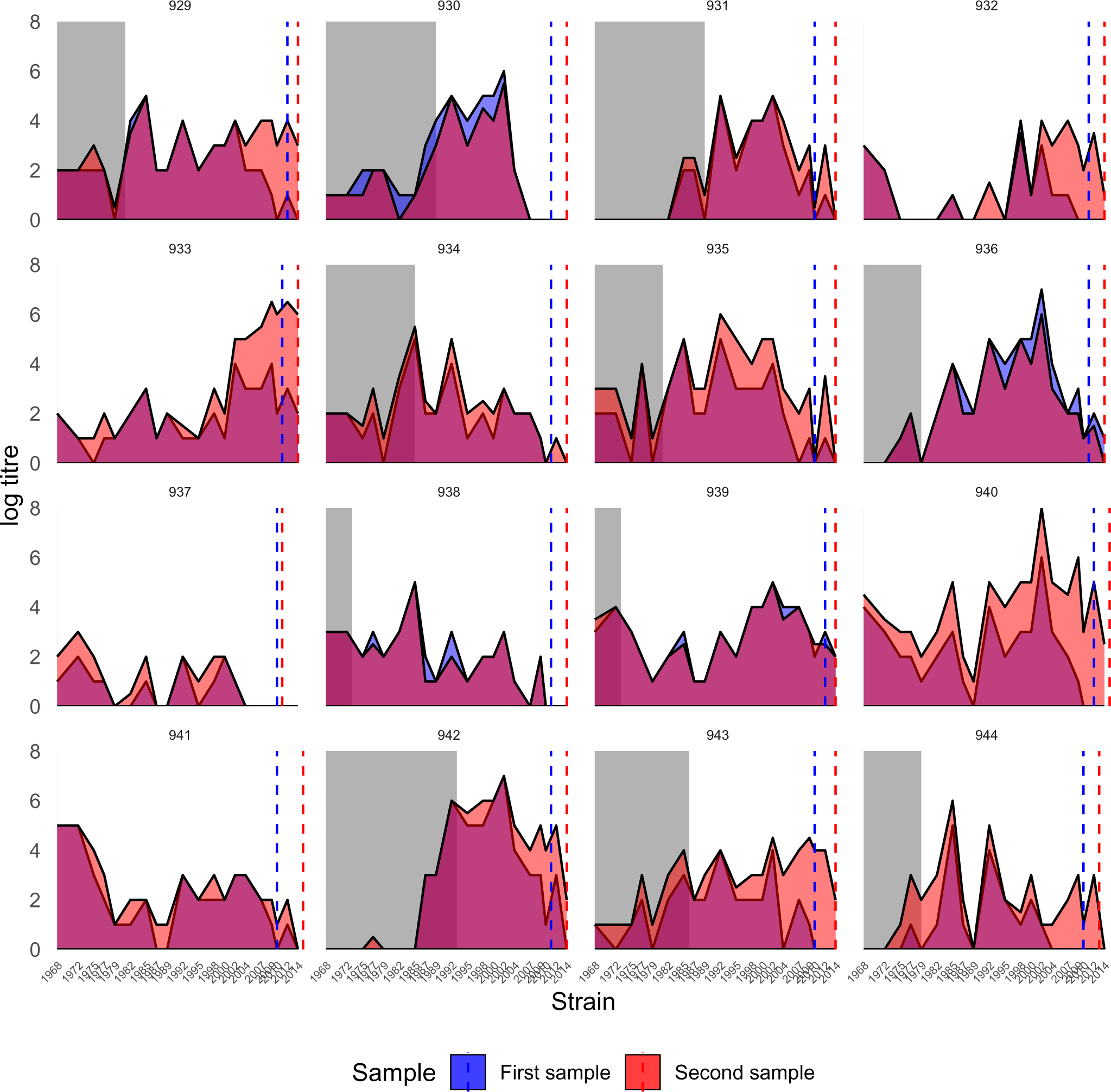
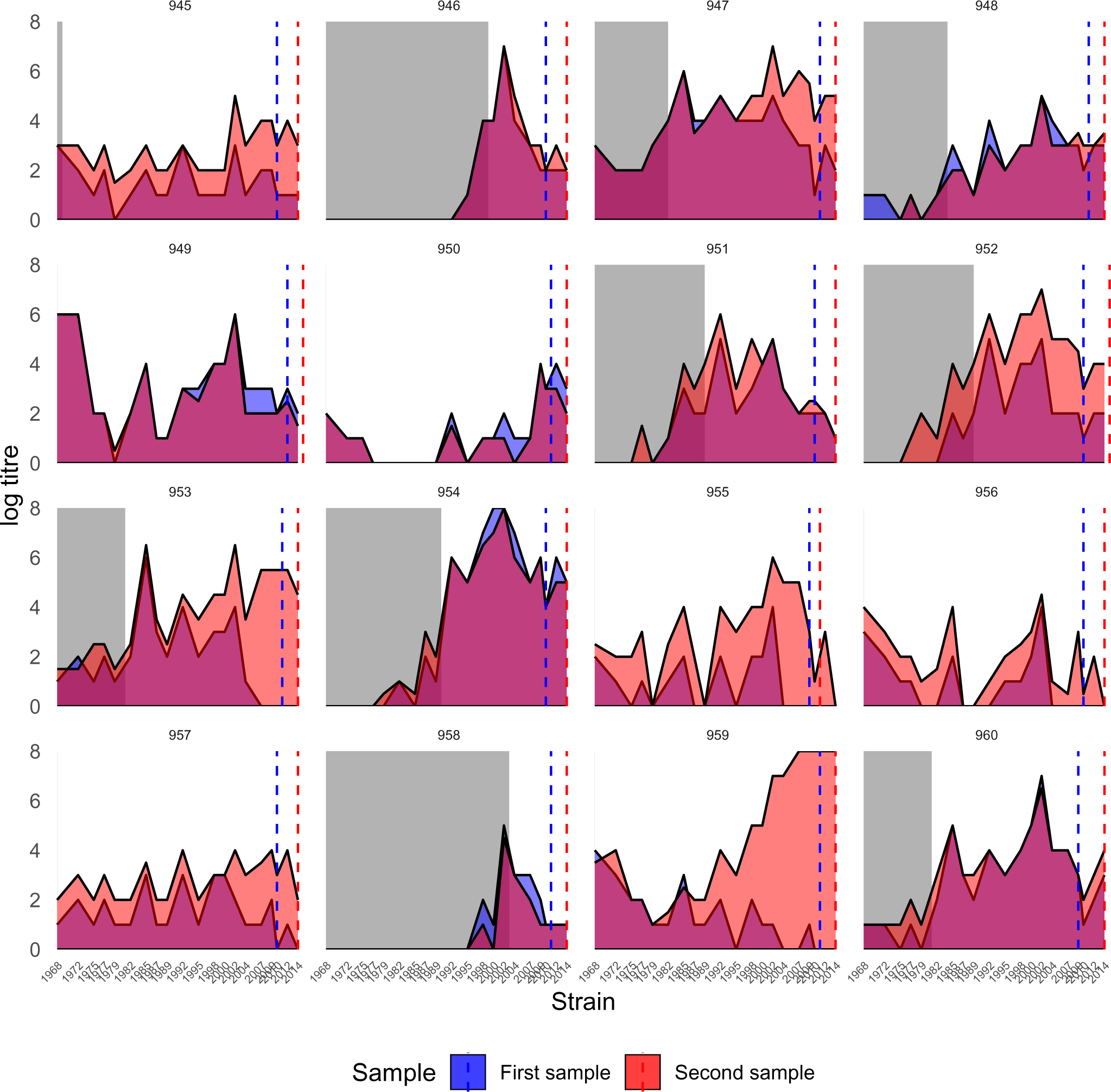
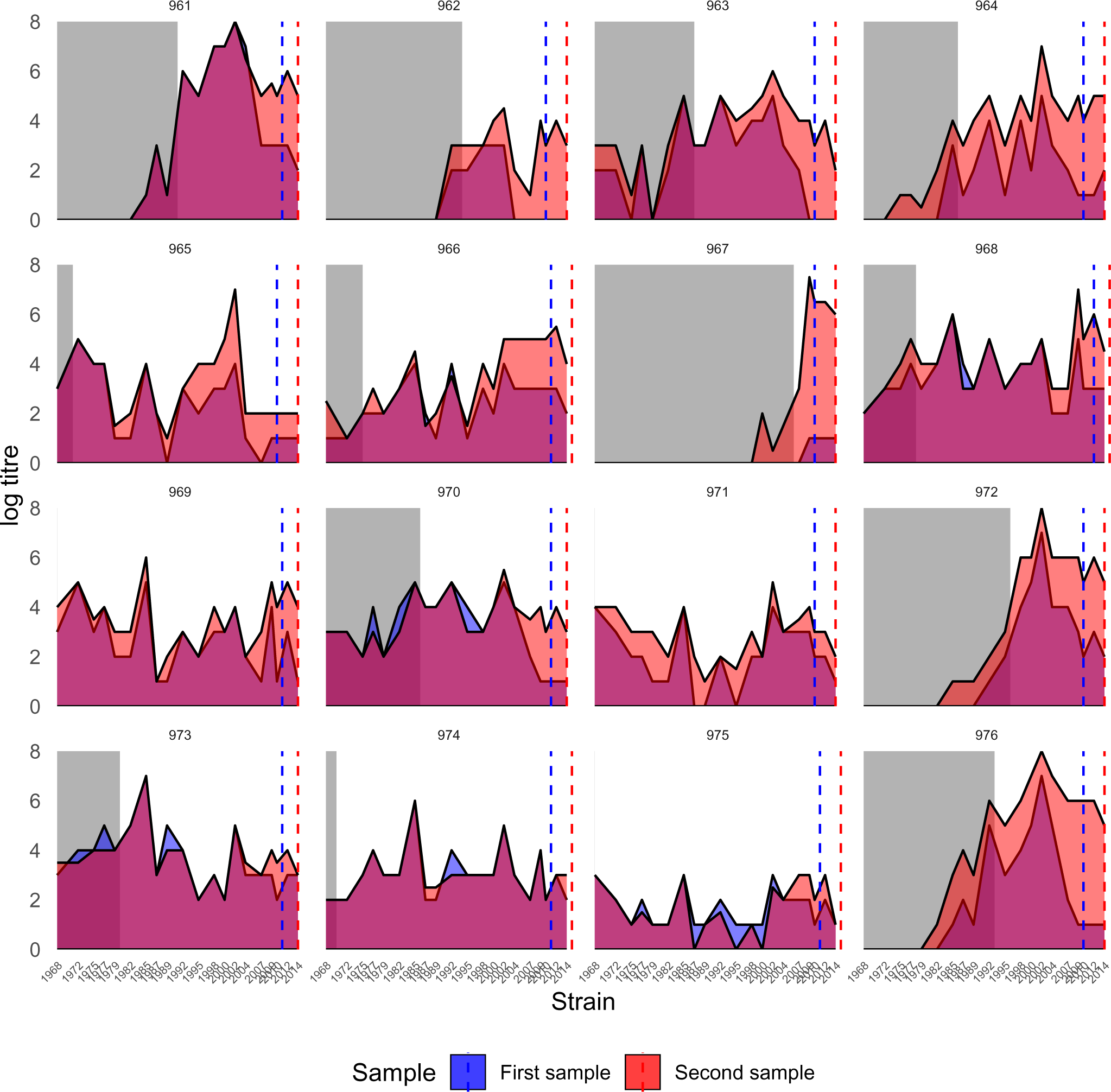
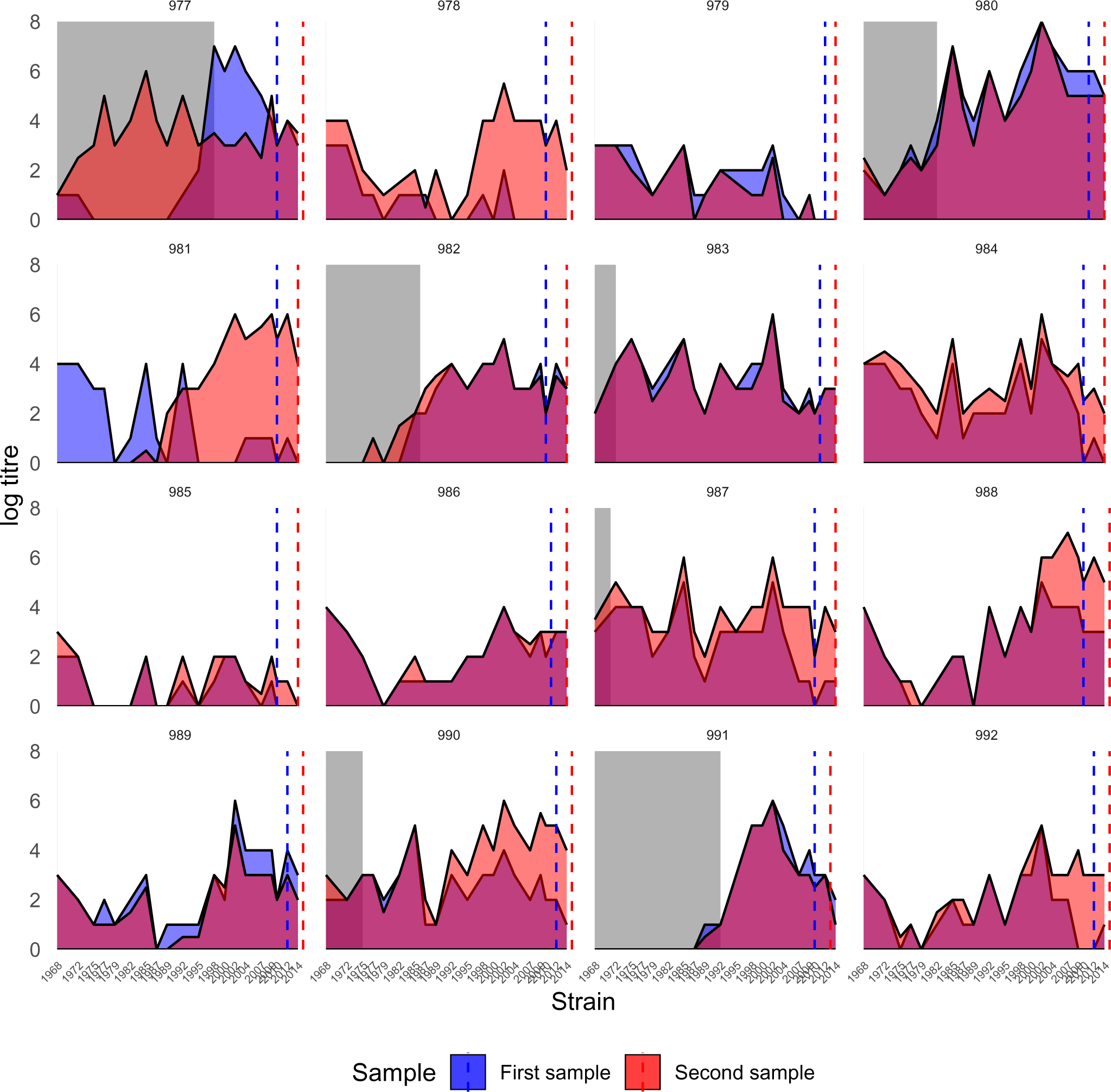
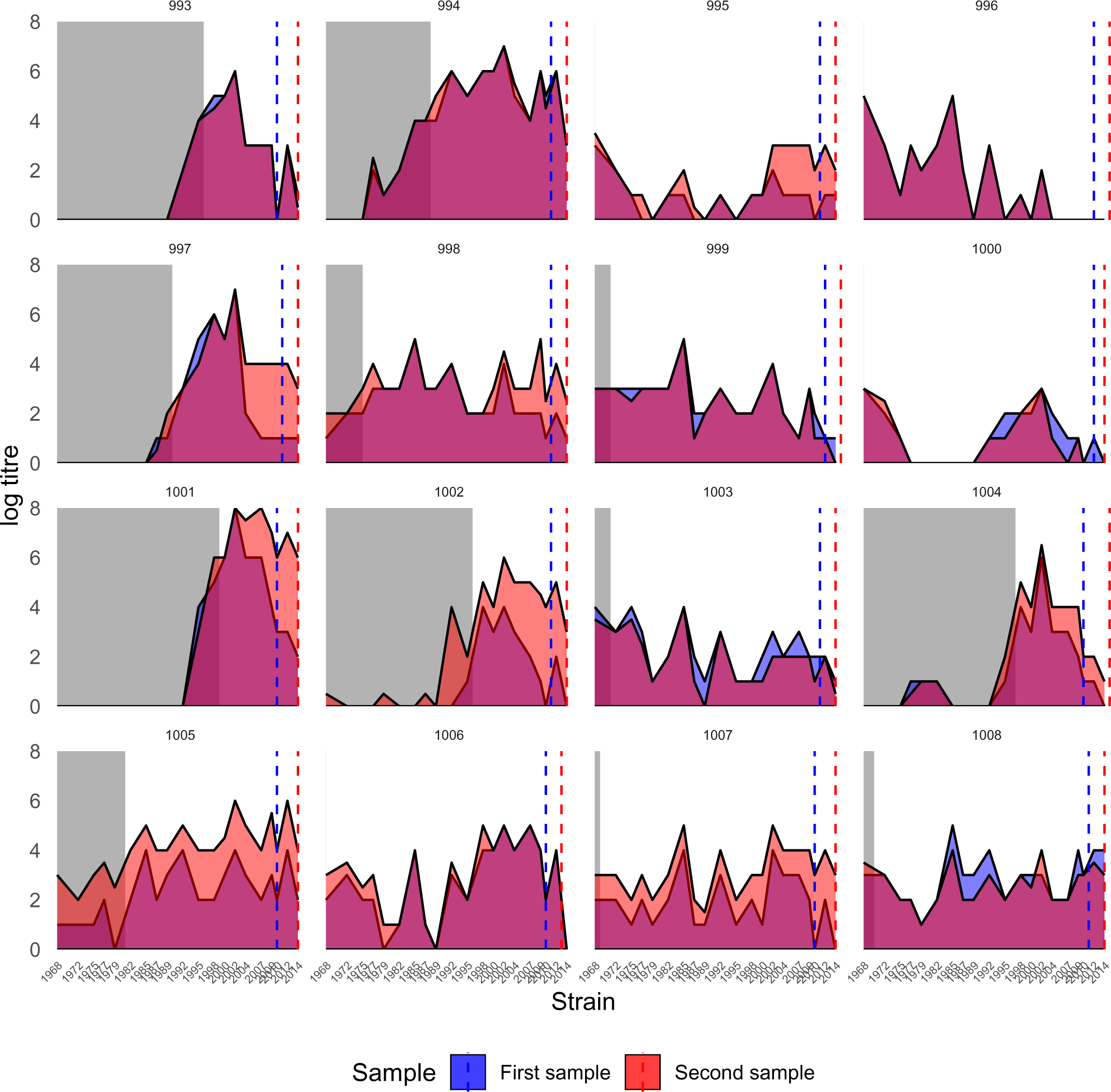
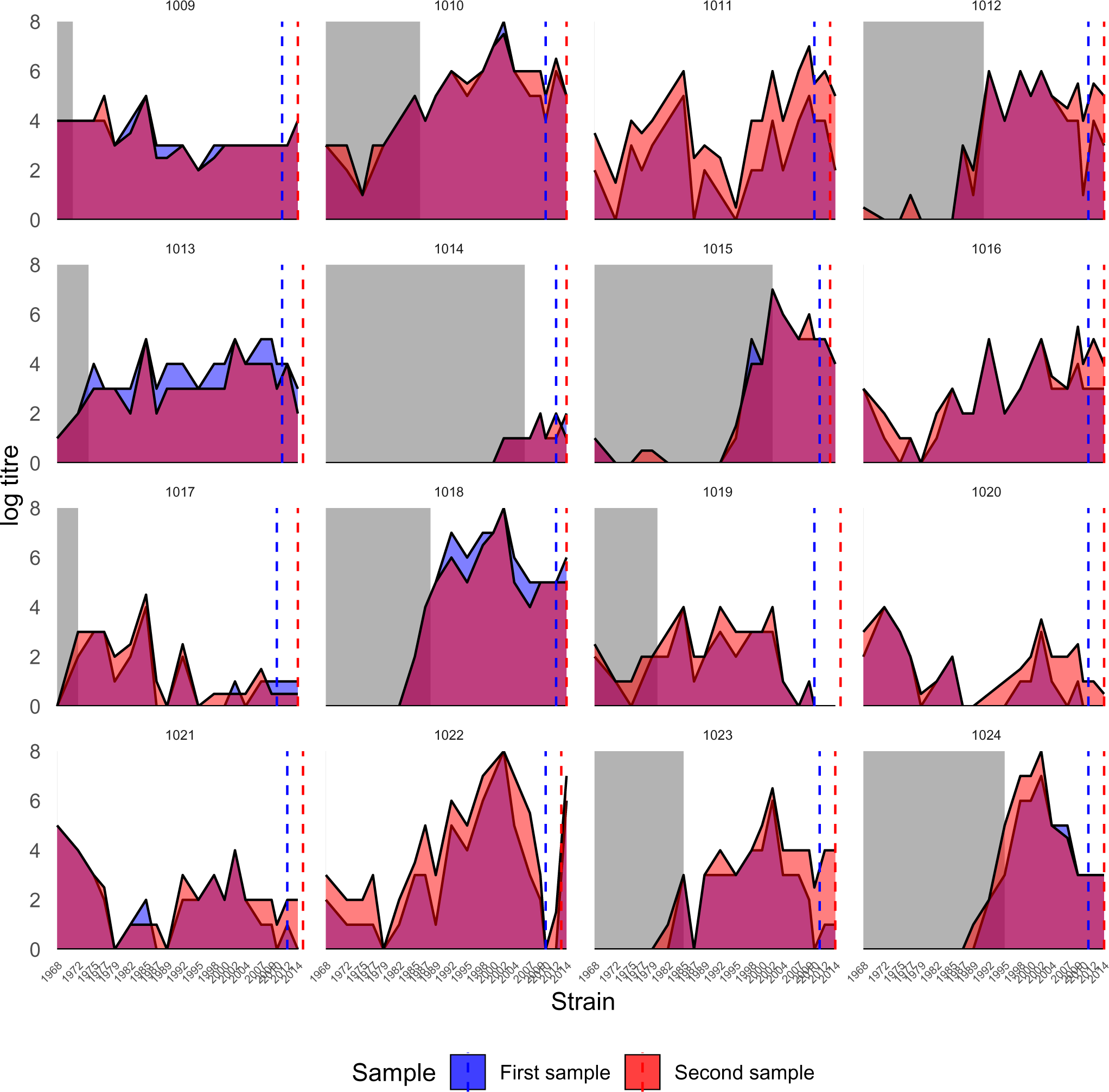
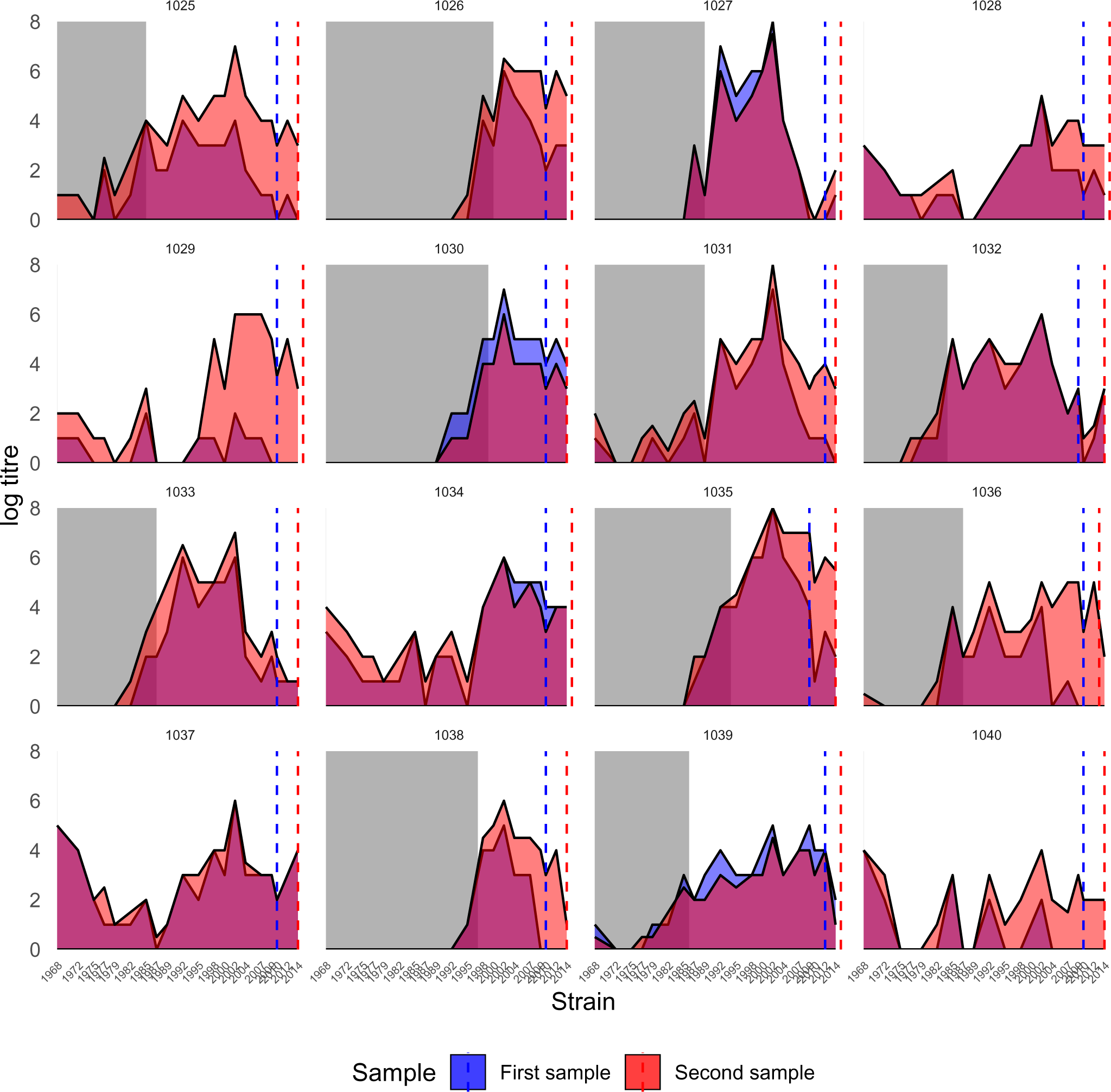
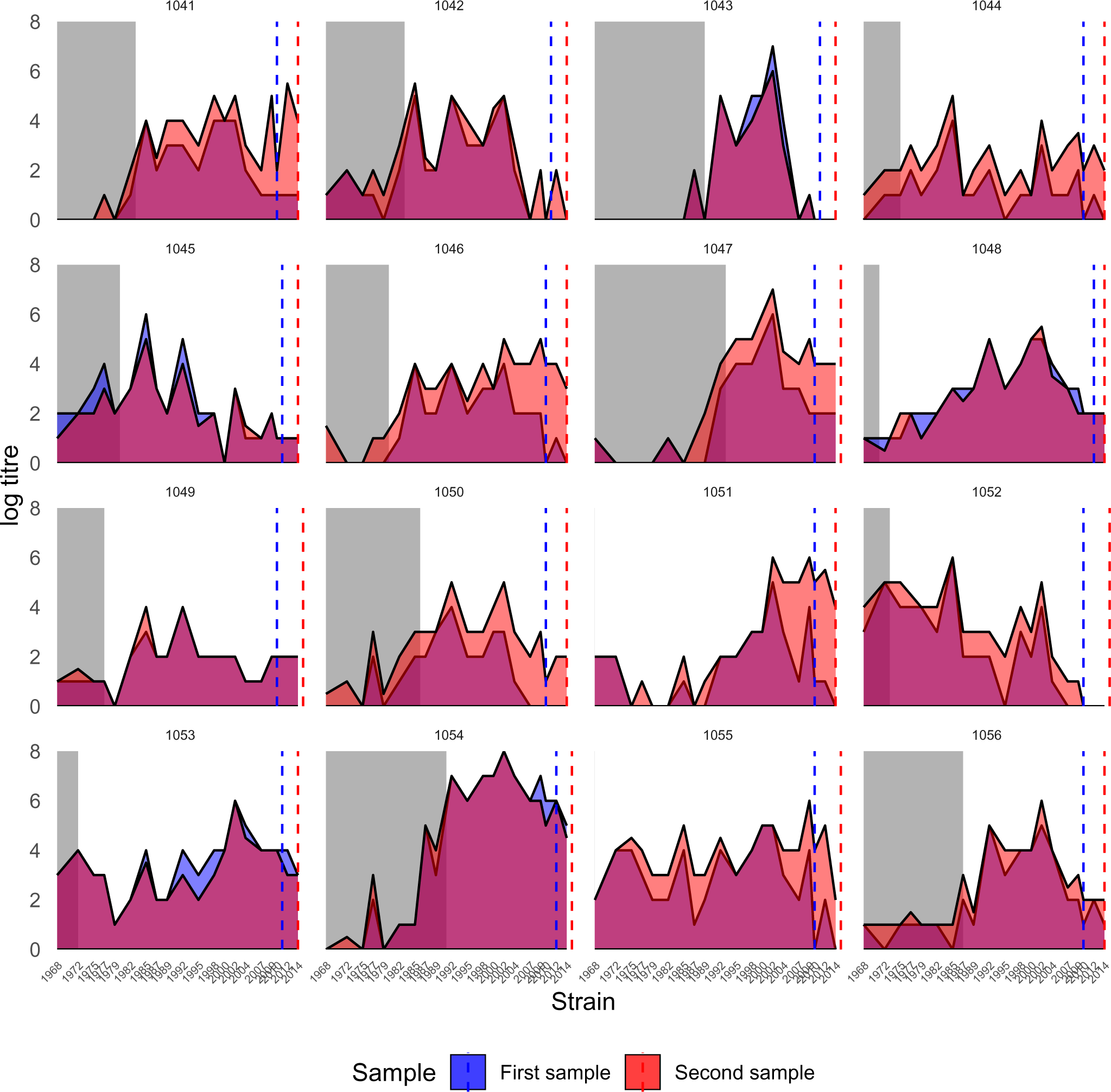
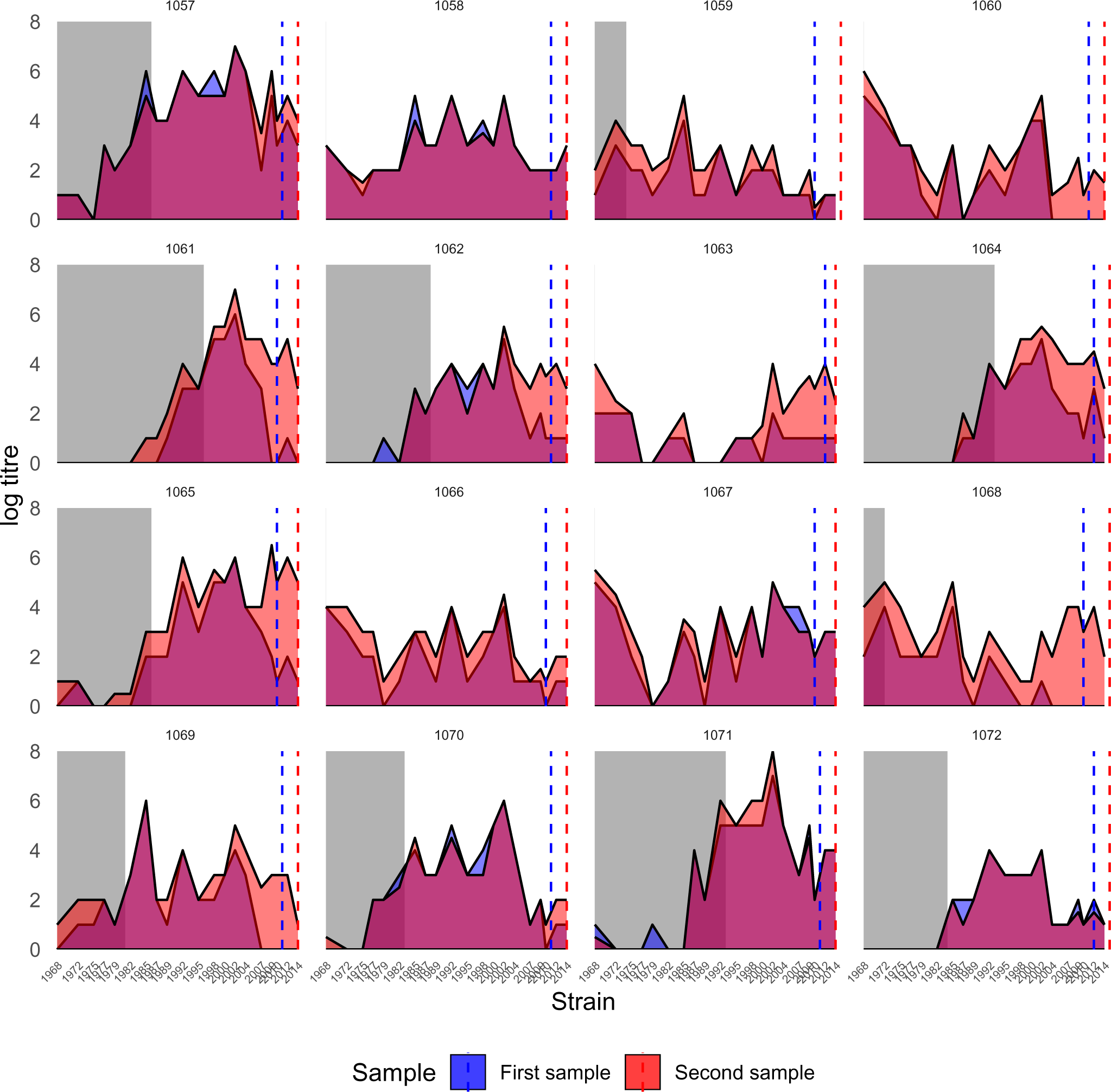
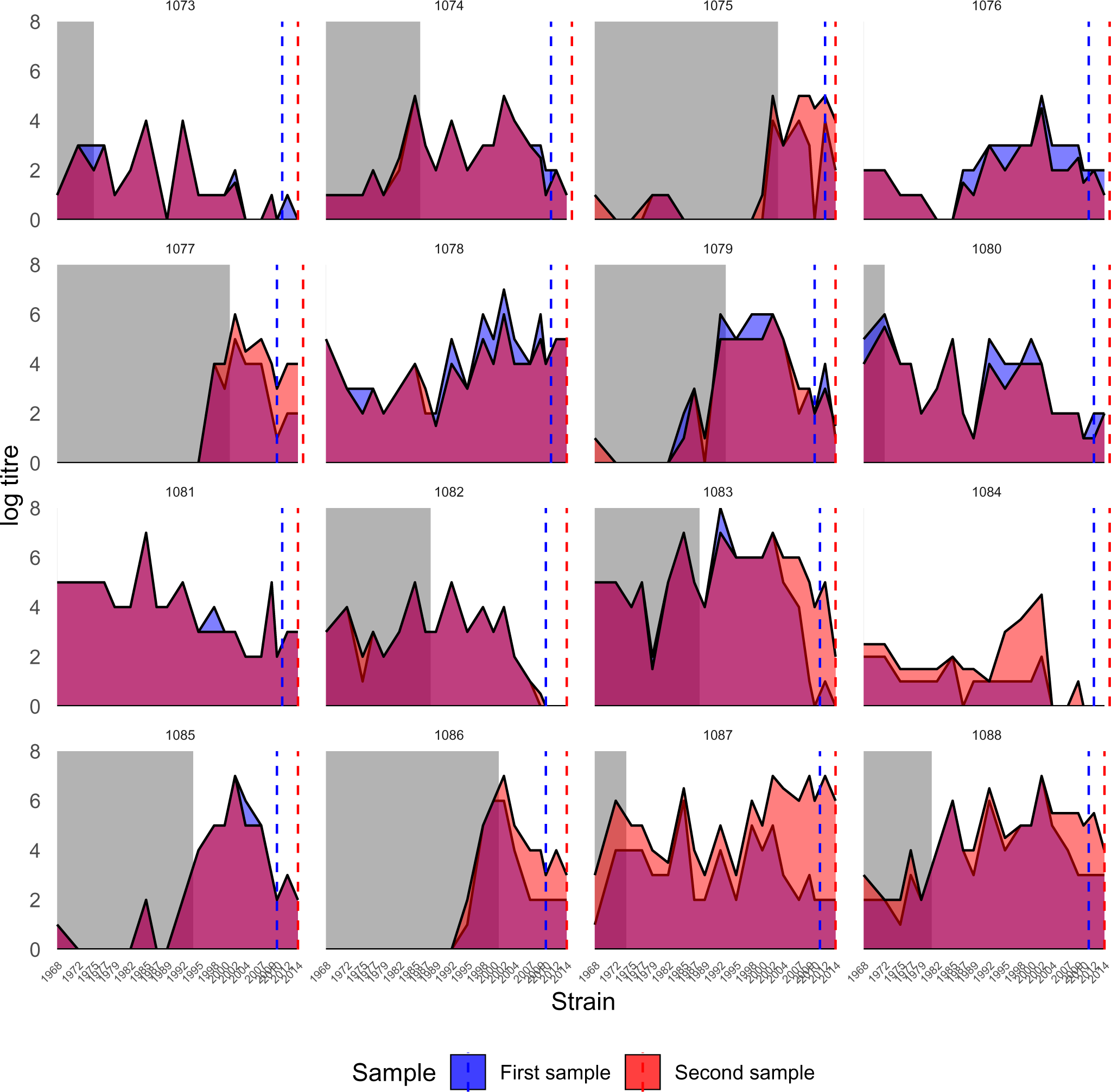
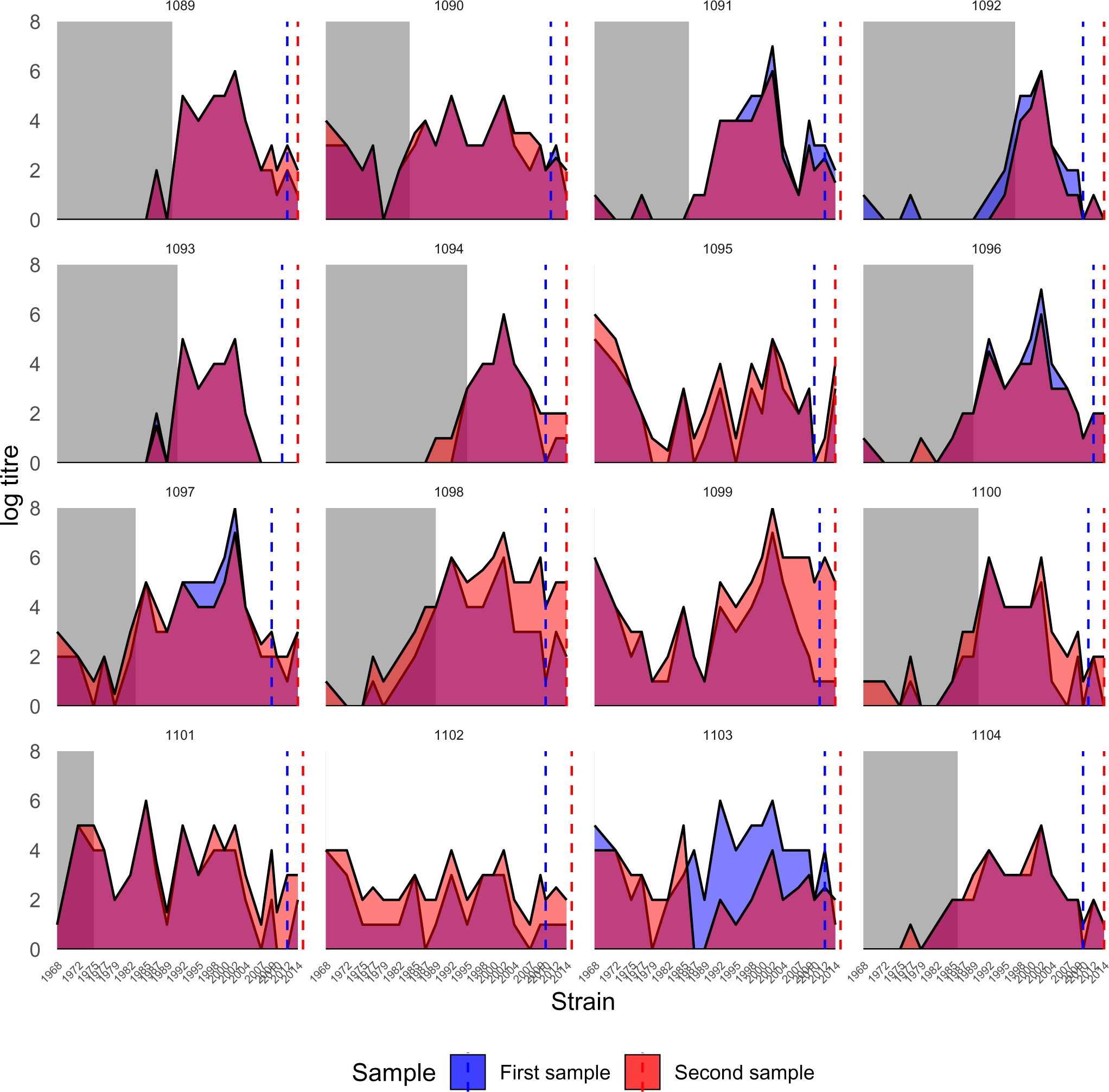
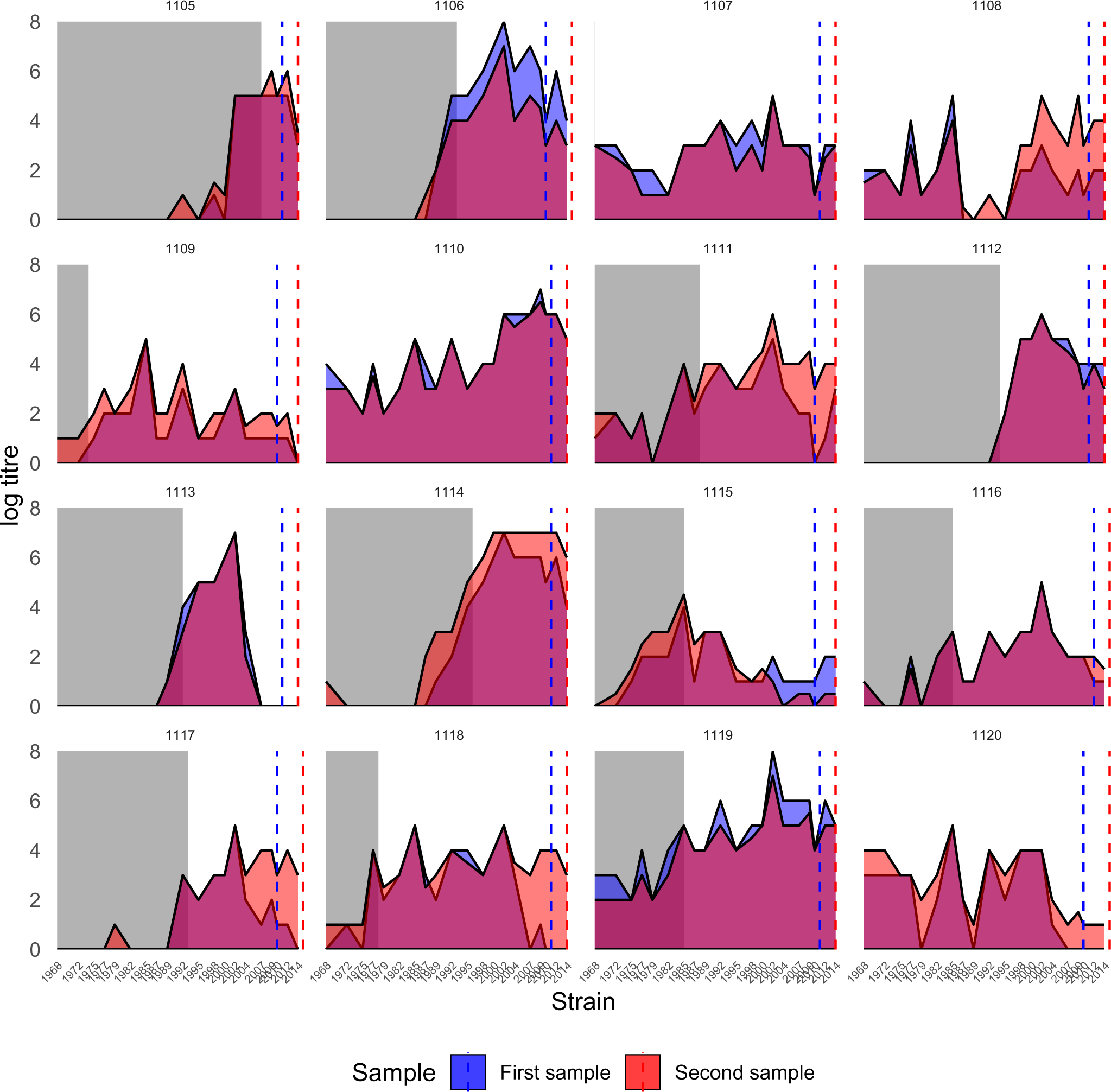
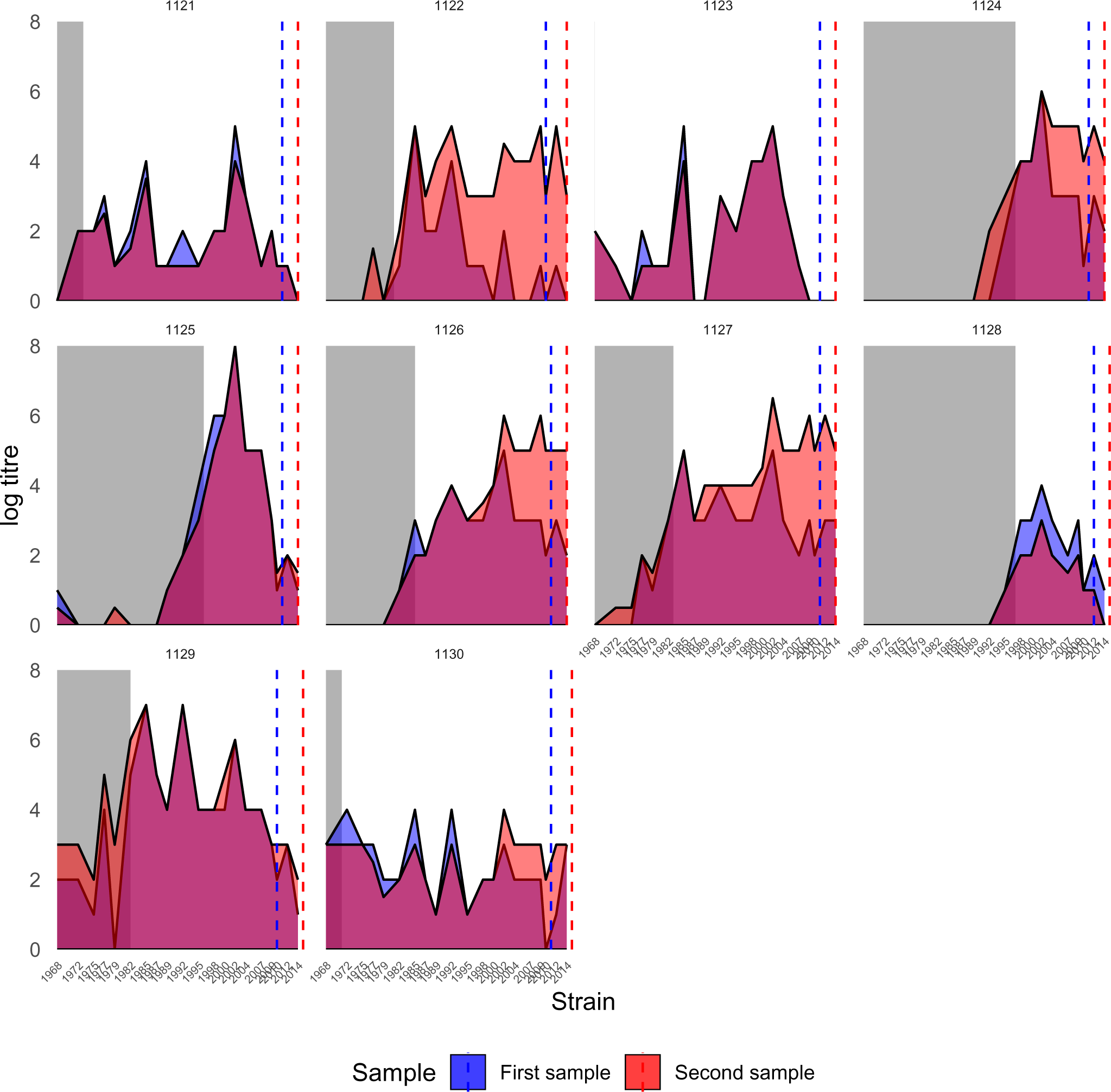
Observed antibody profiles for all individuals in the Fluscape cohort. Each subplot shows antibody levels measured against each of the 20 H3N2 strains. The x-axis shows the isolation year of the measured strain. The areas are shaded by sample number, showing titre measurements from the first (blue) and second (red) samples. Grey rectangles mark strains which circulated before that individual was born. The vertical colored lines show the timing of the serum samples relative to the strain isolation times. Plots where the red region extends above the blue region reflect antibody boosting between the first and second serum sample. Where multiple titres were measured against the same strain from the same serum sample, we plotted the mean of the log titres.

**Figure S21:**
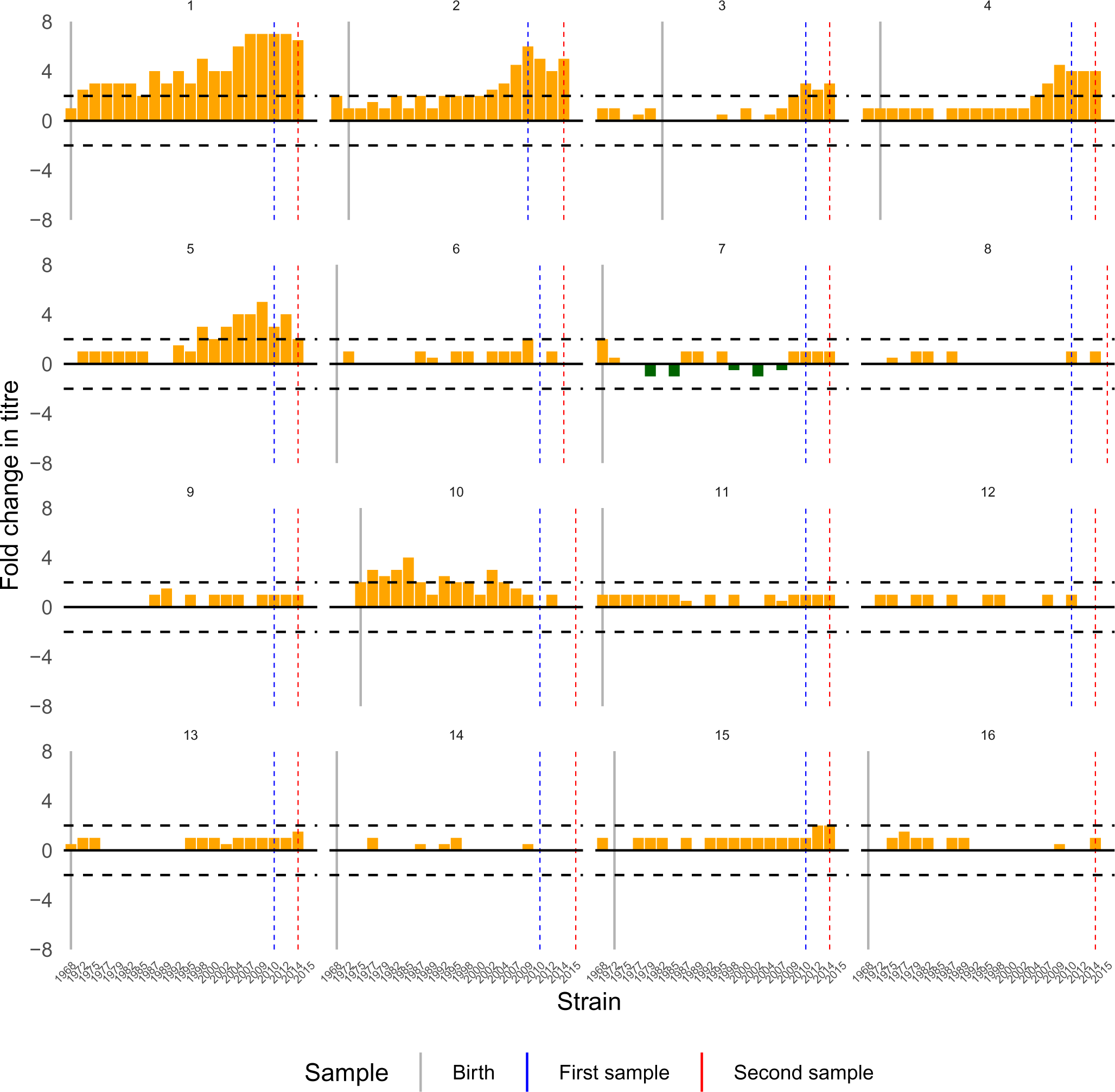
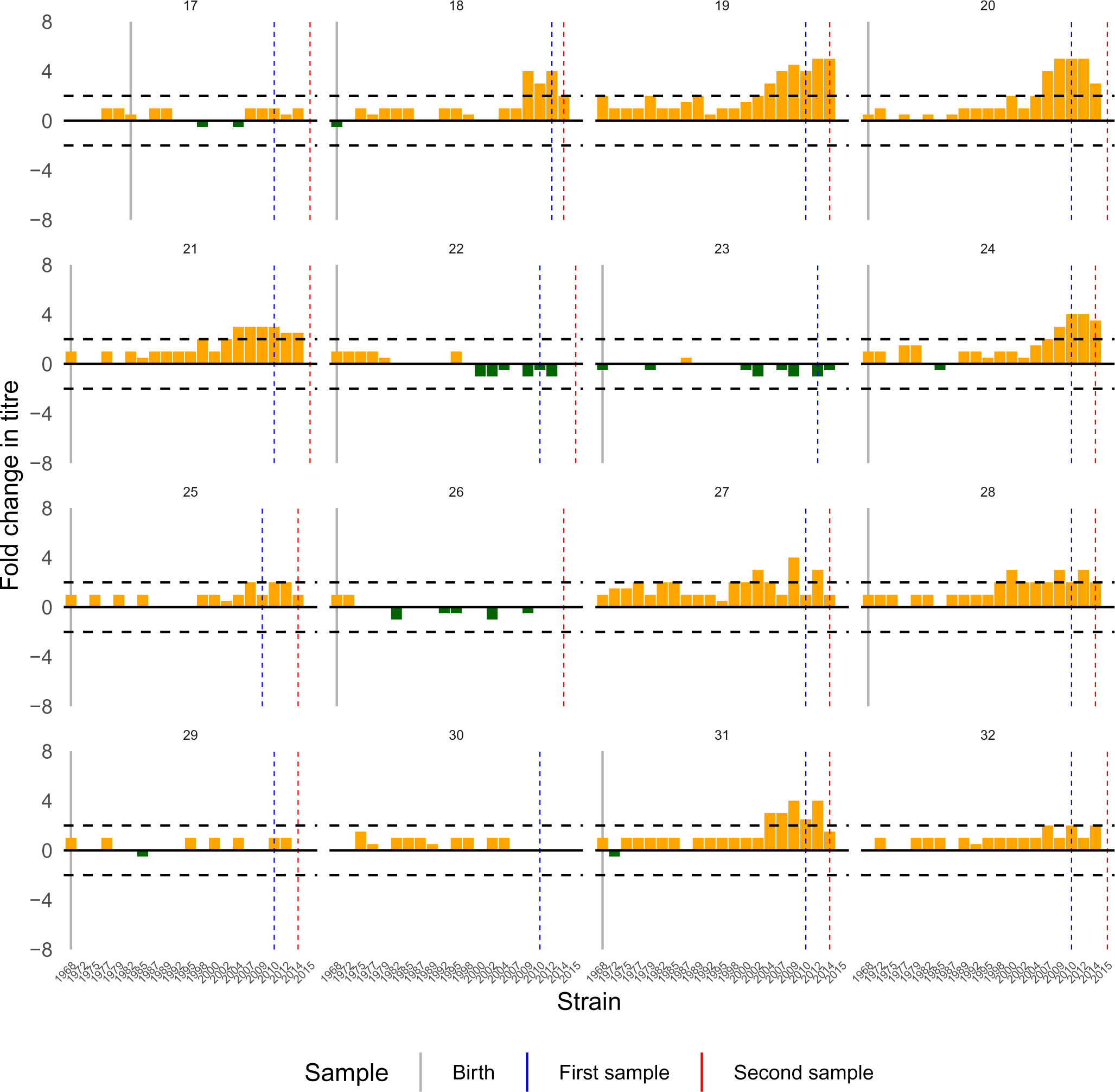
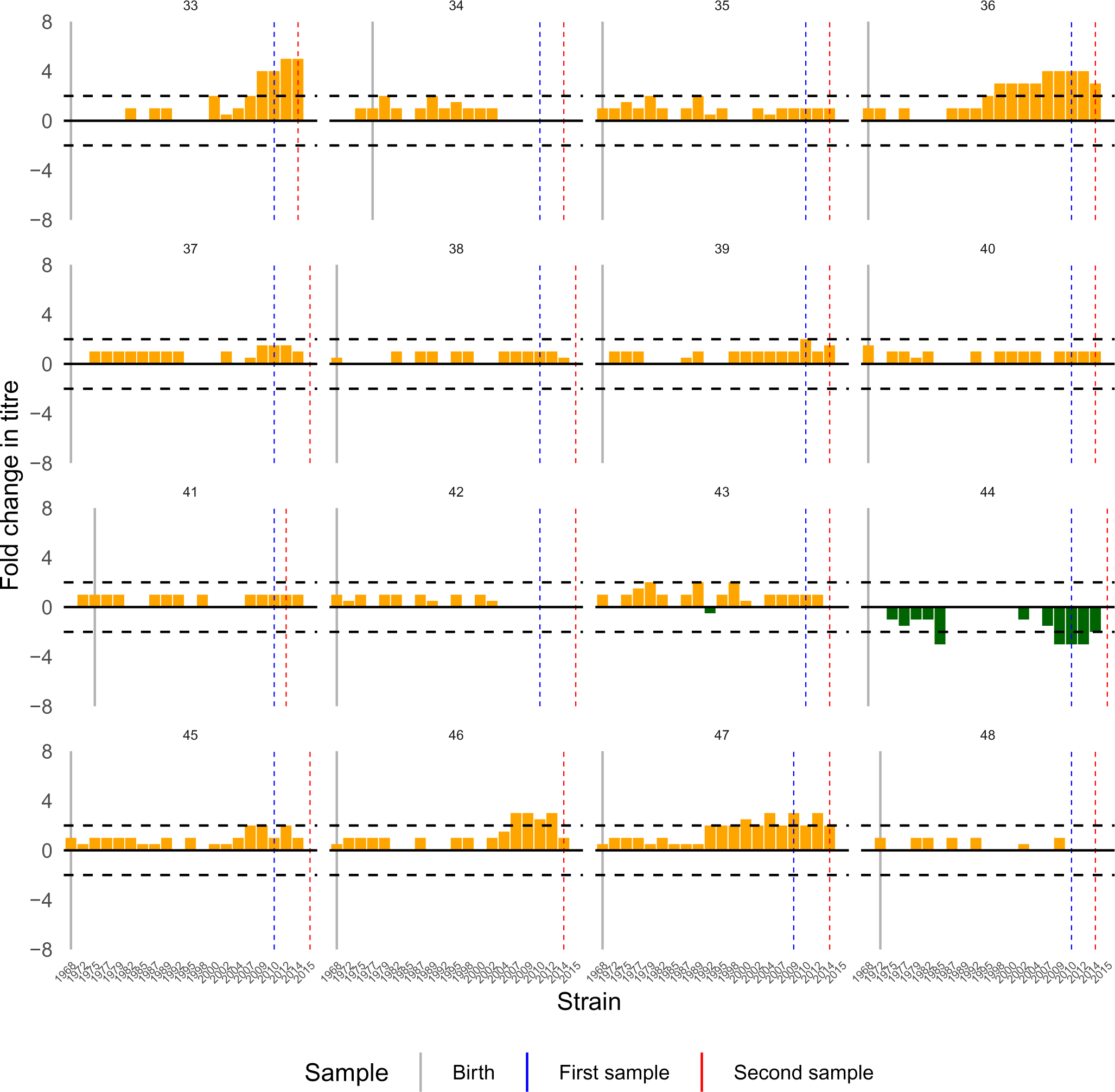
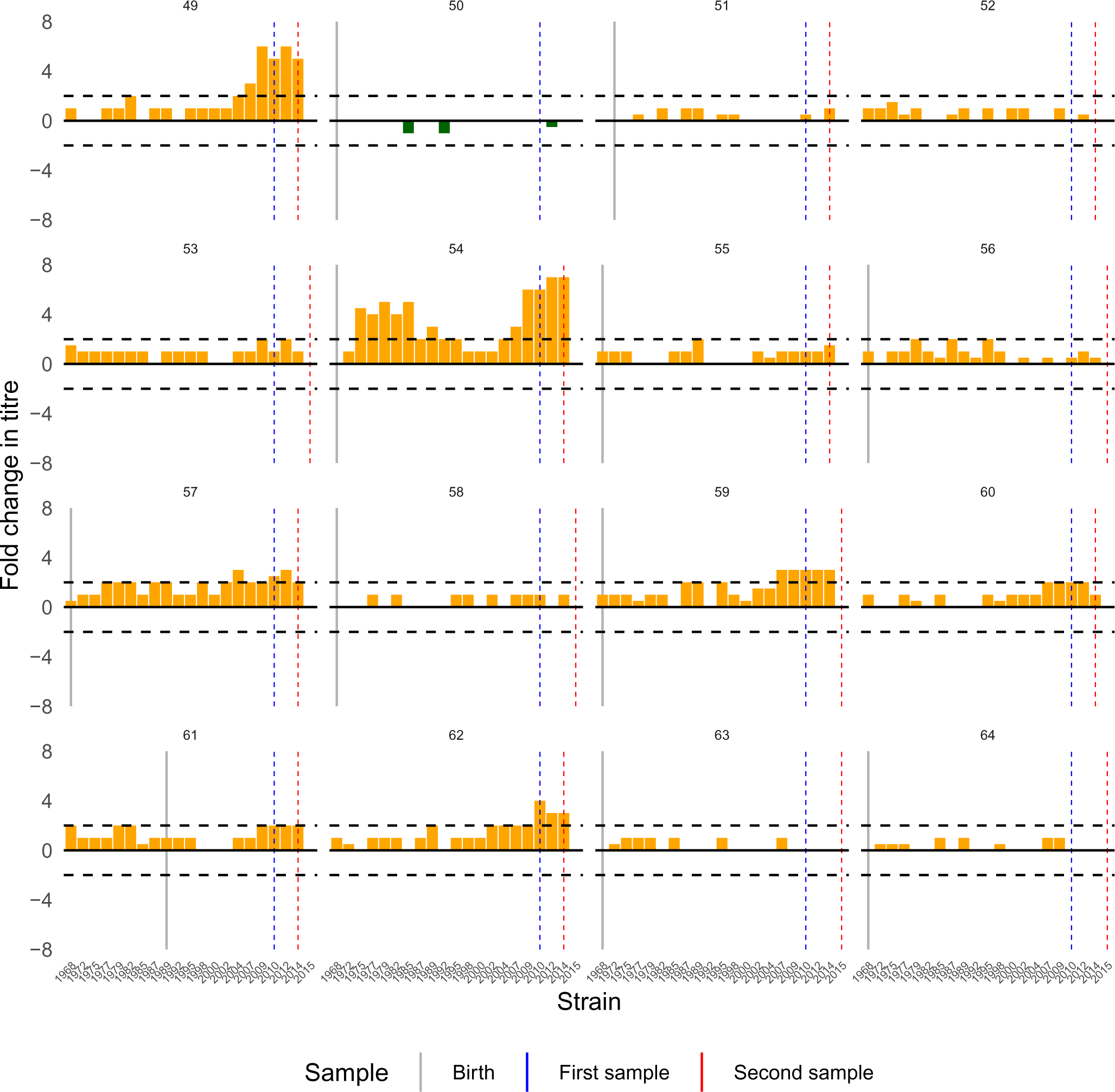
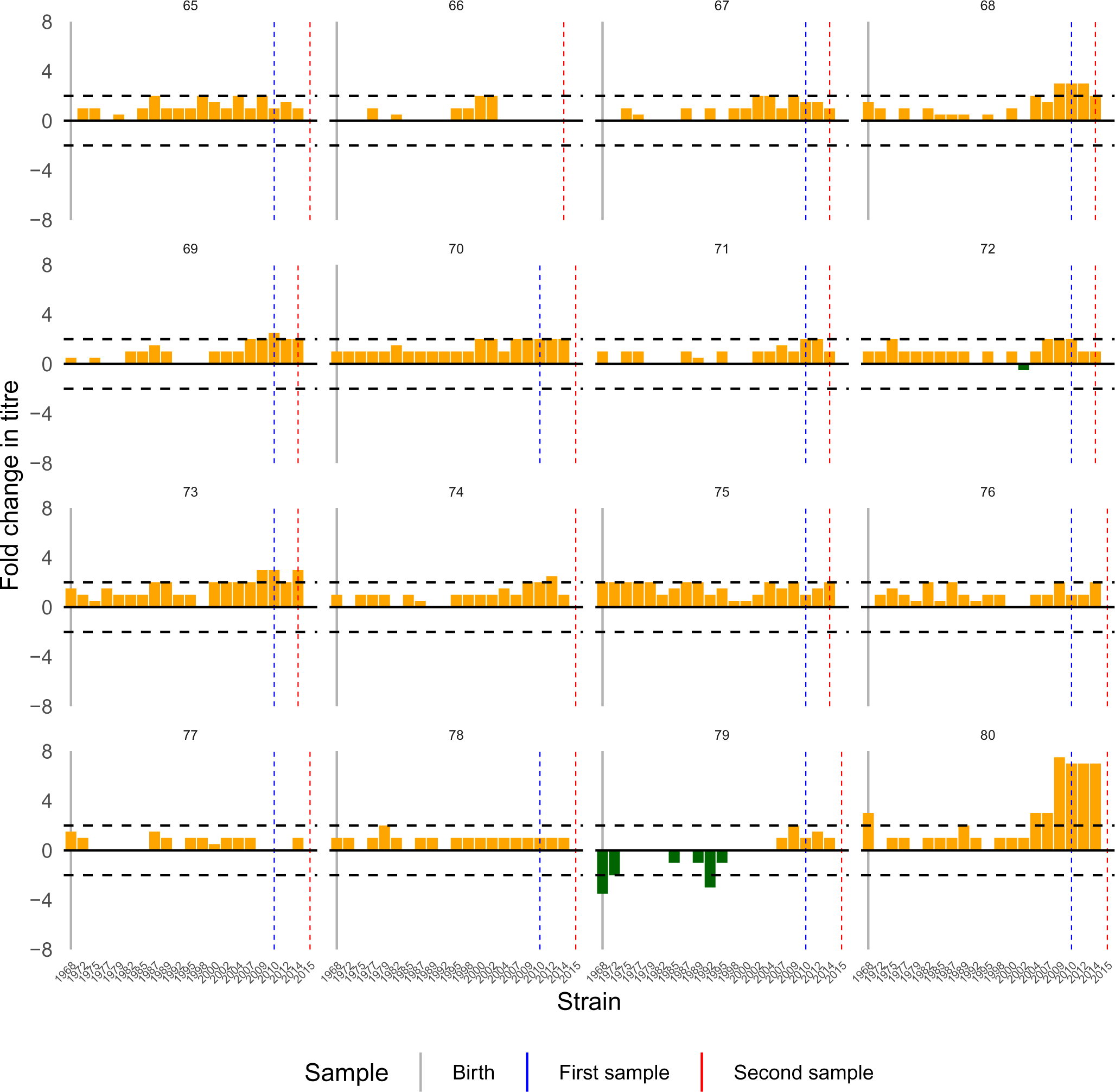
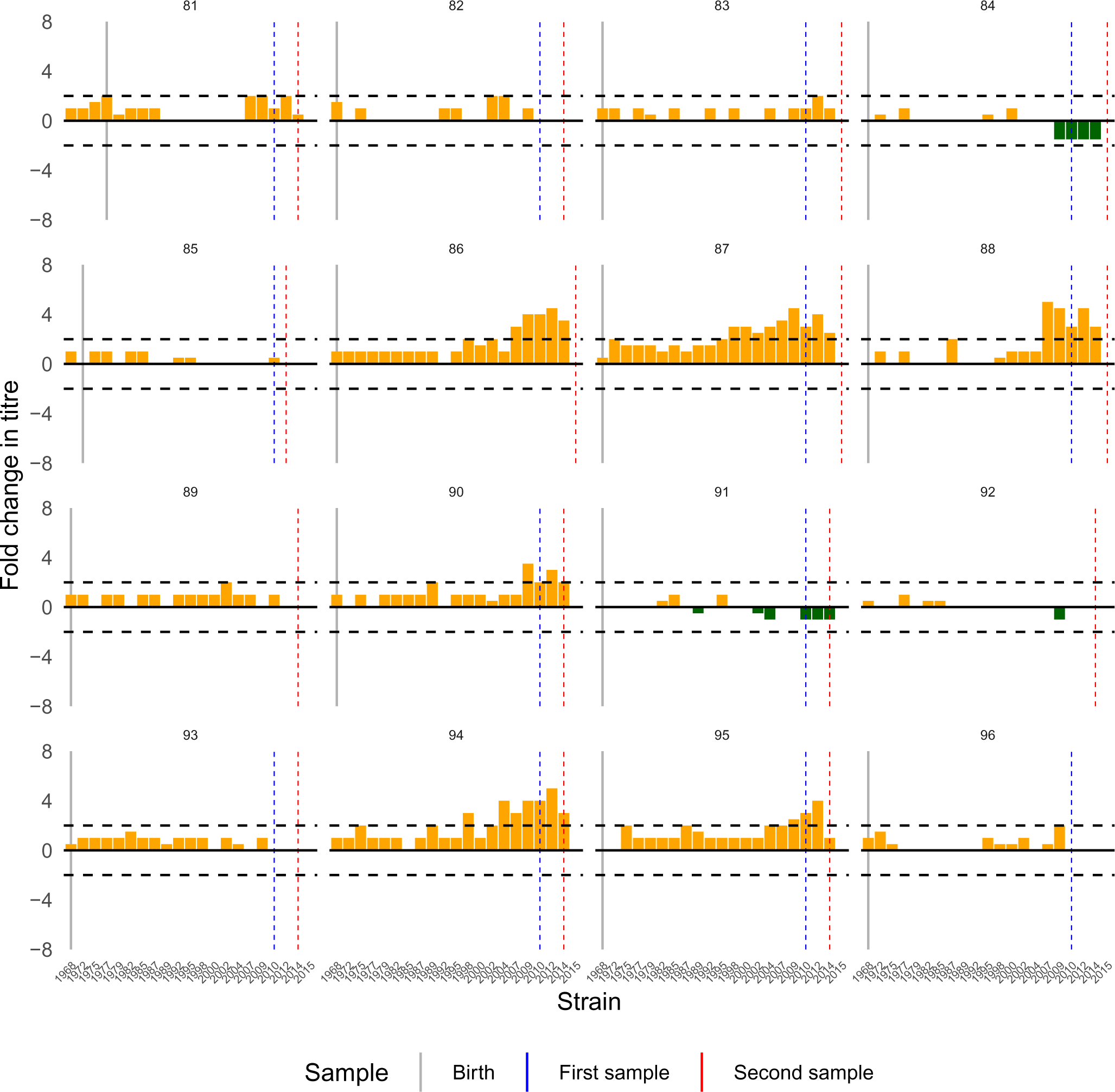
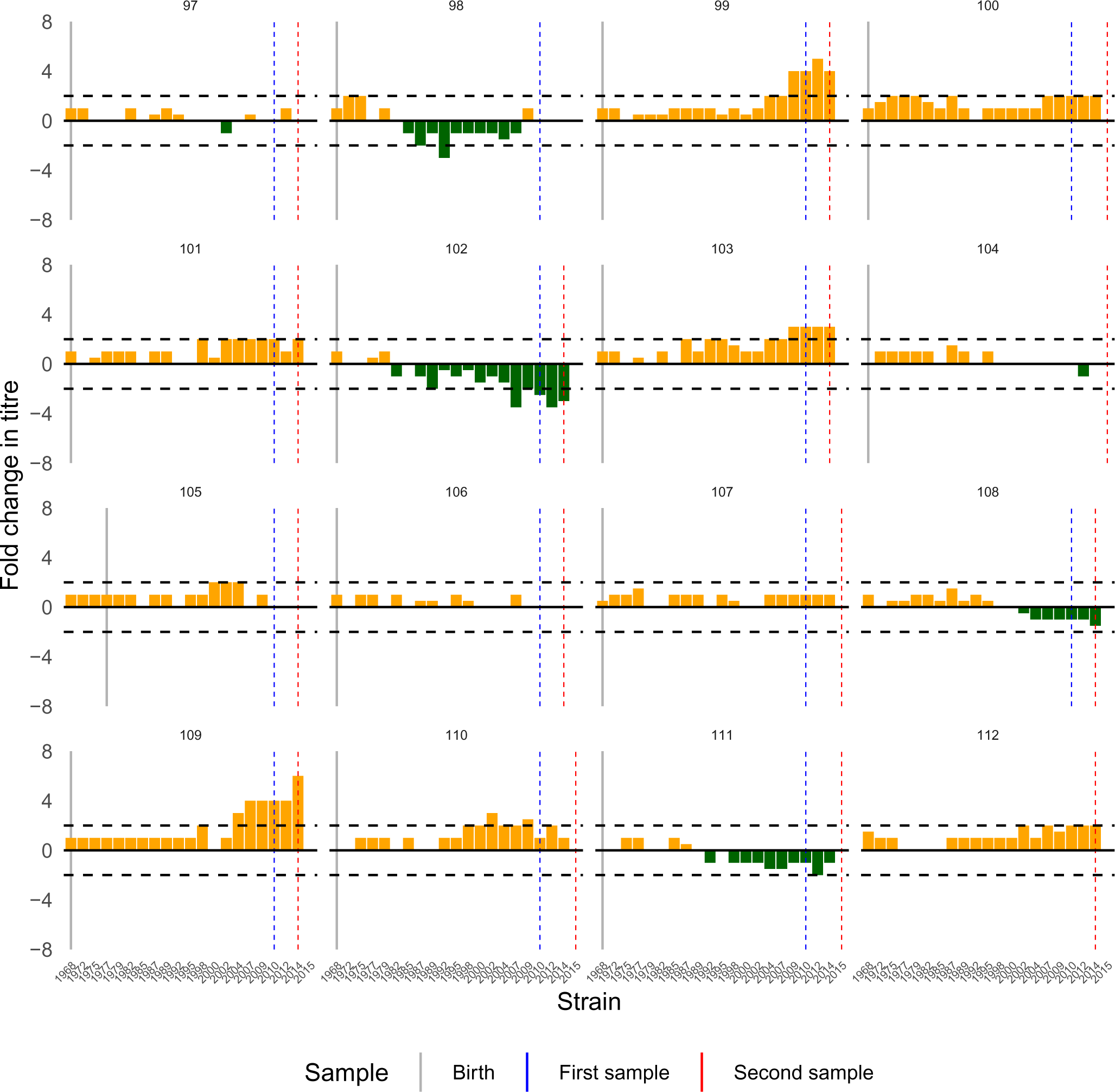
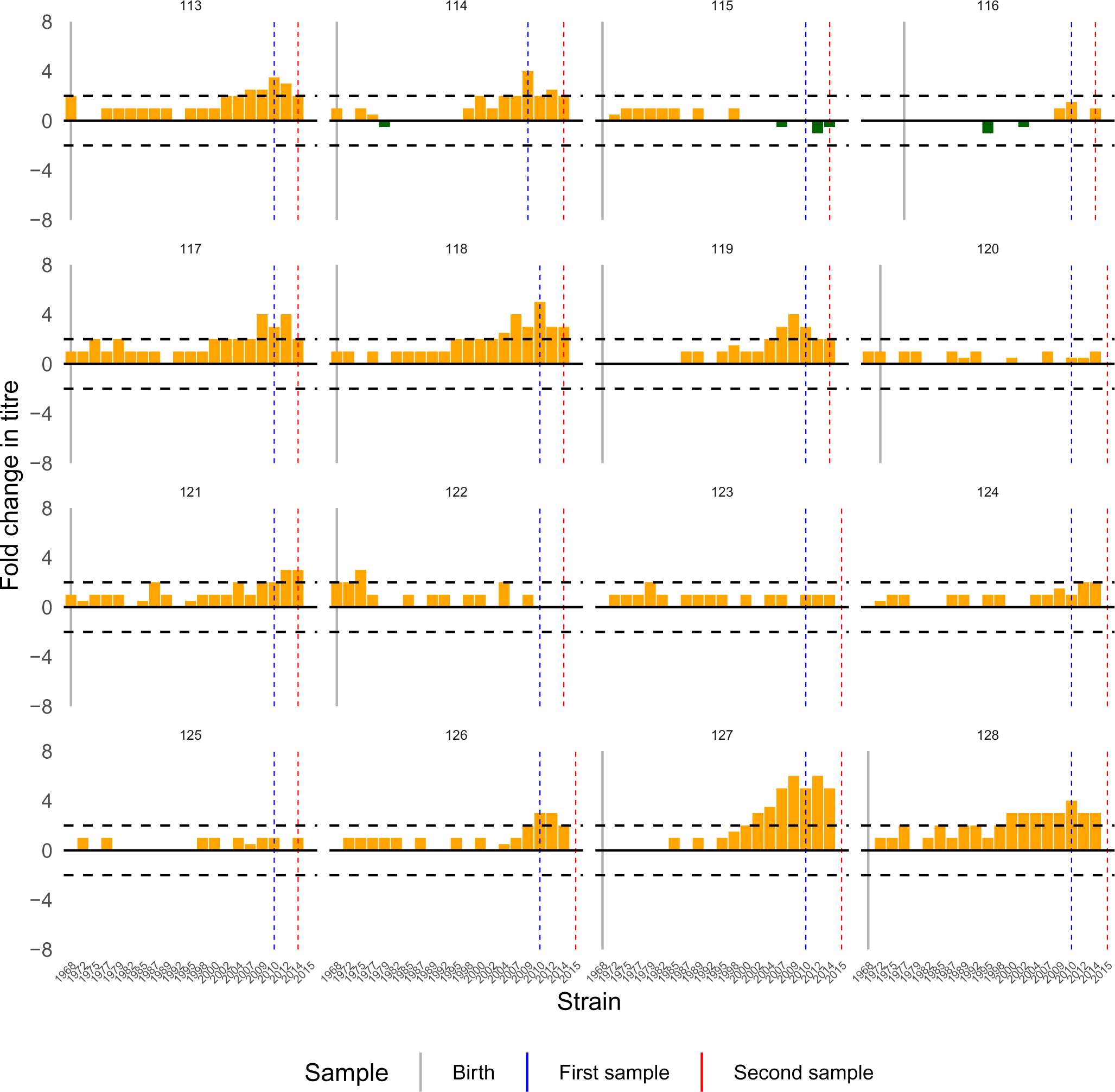
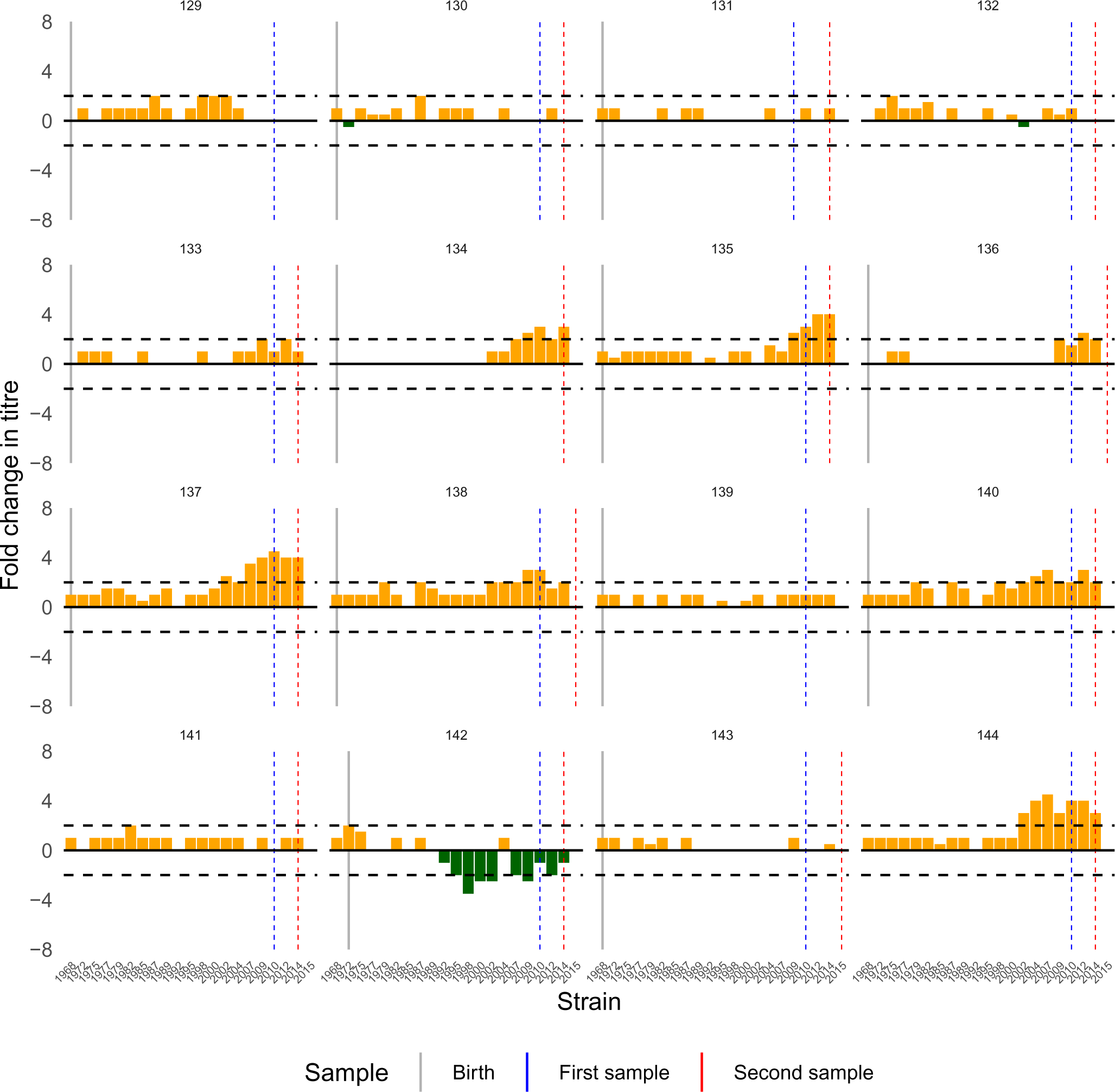
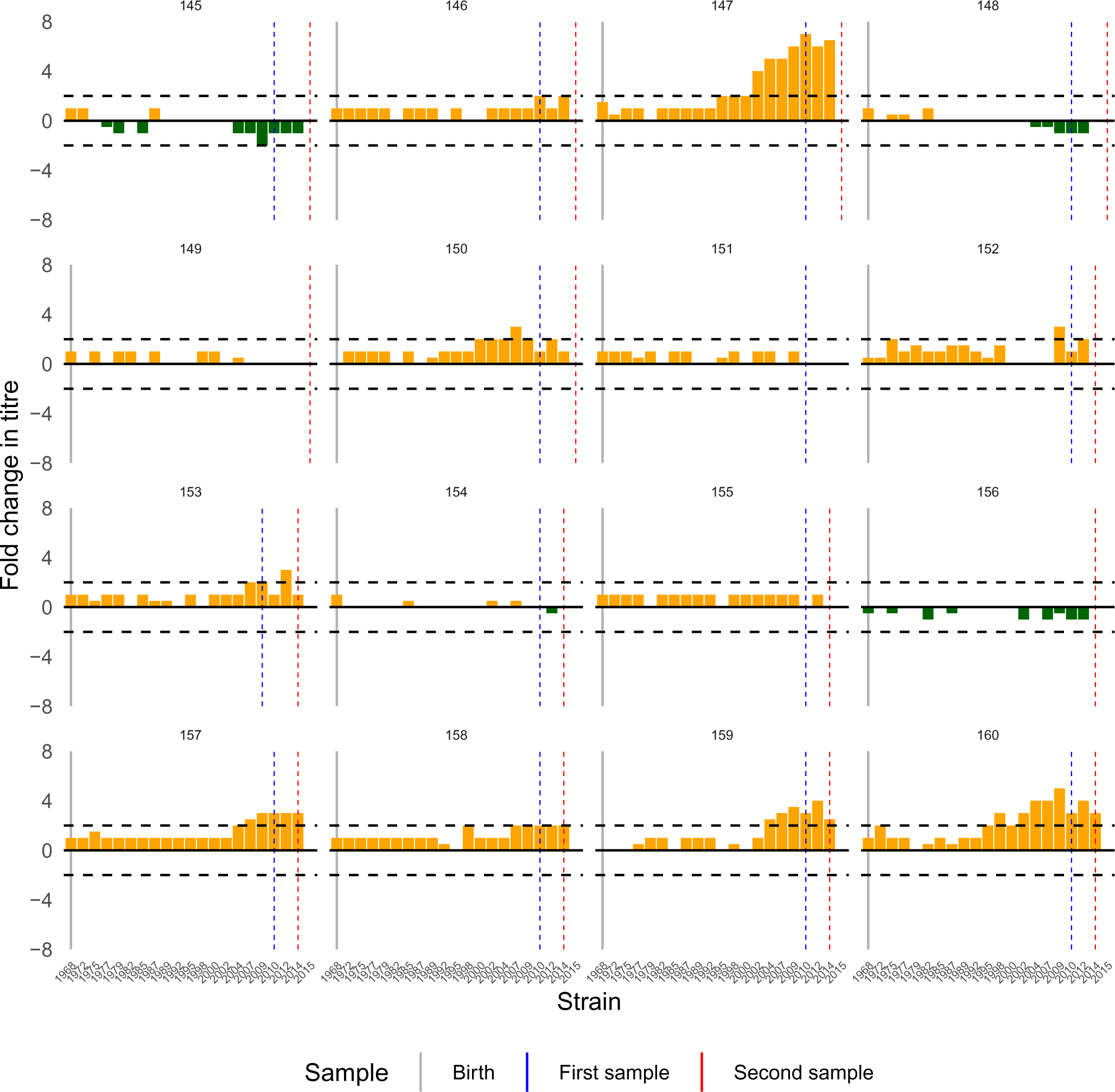
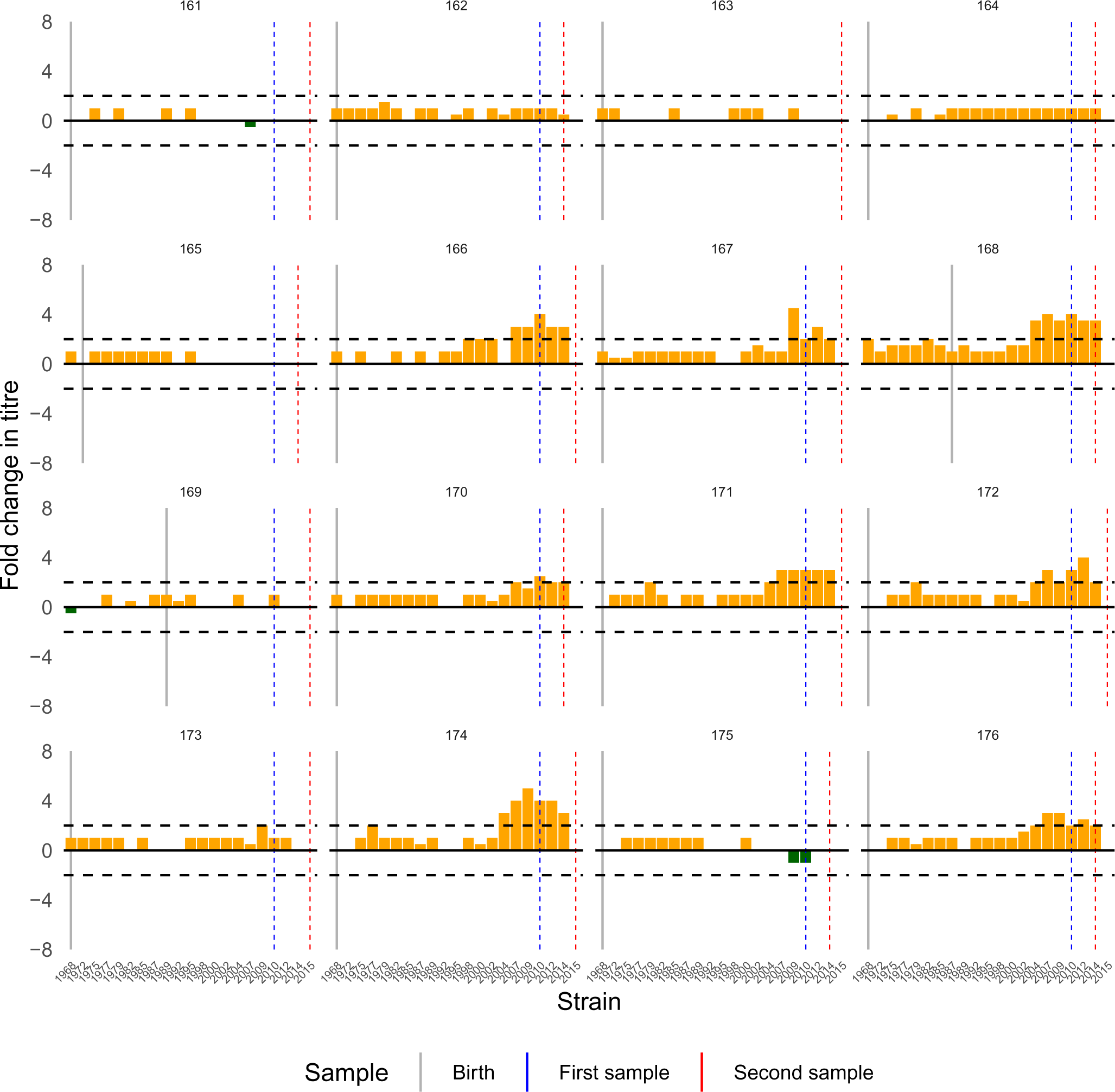
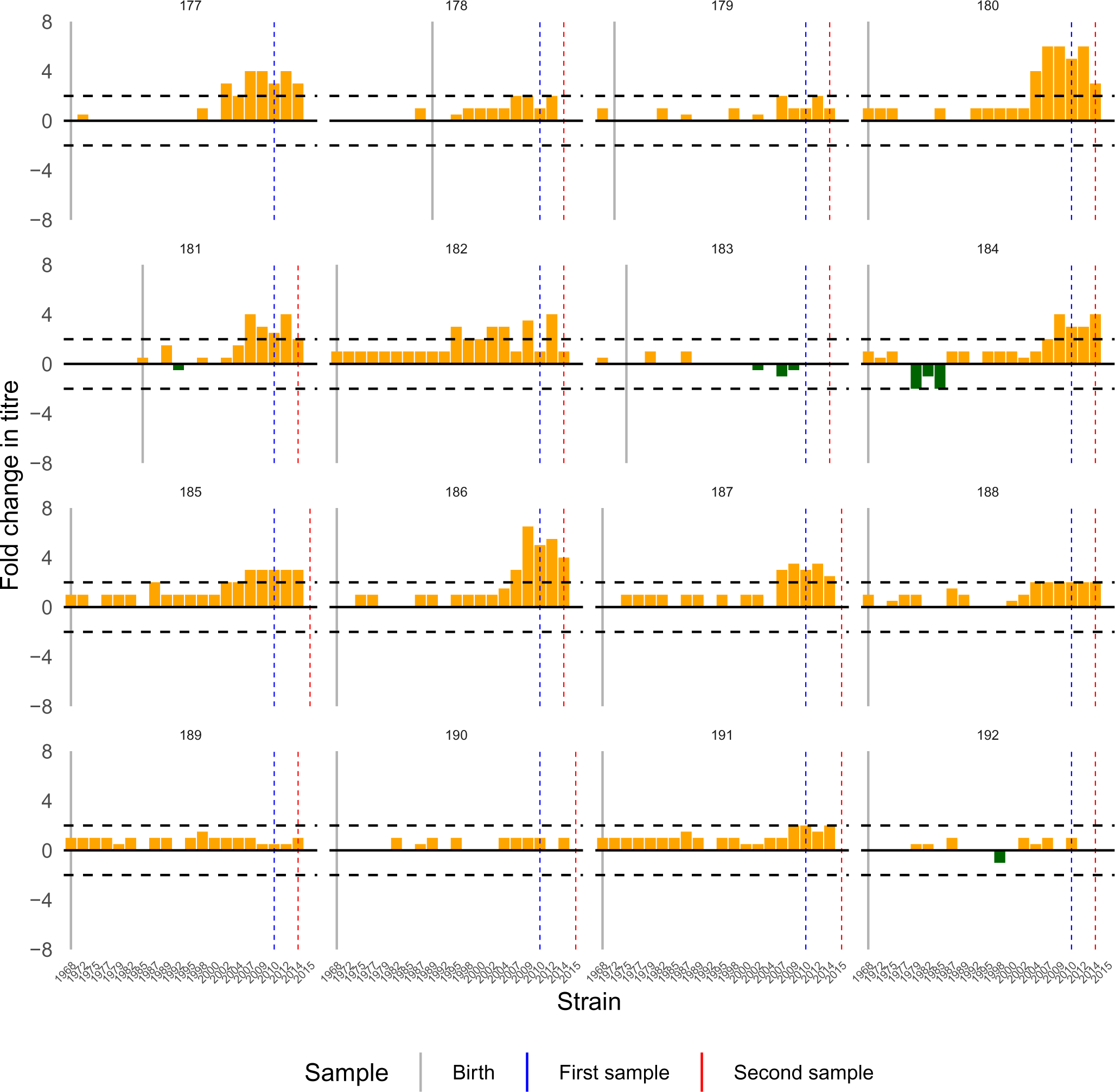
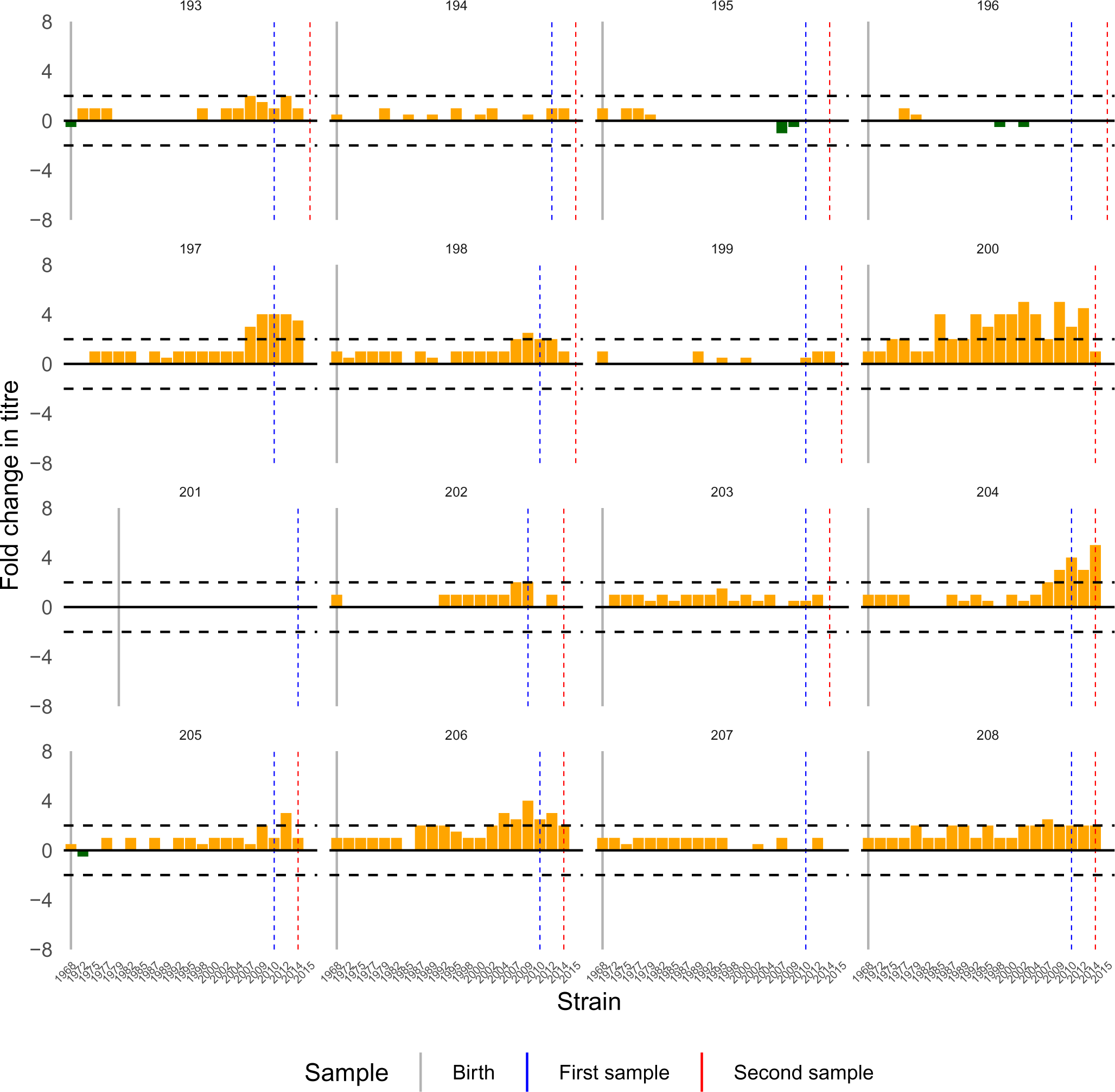
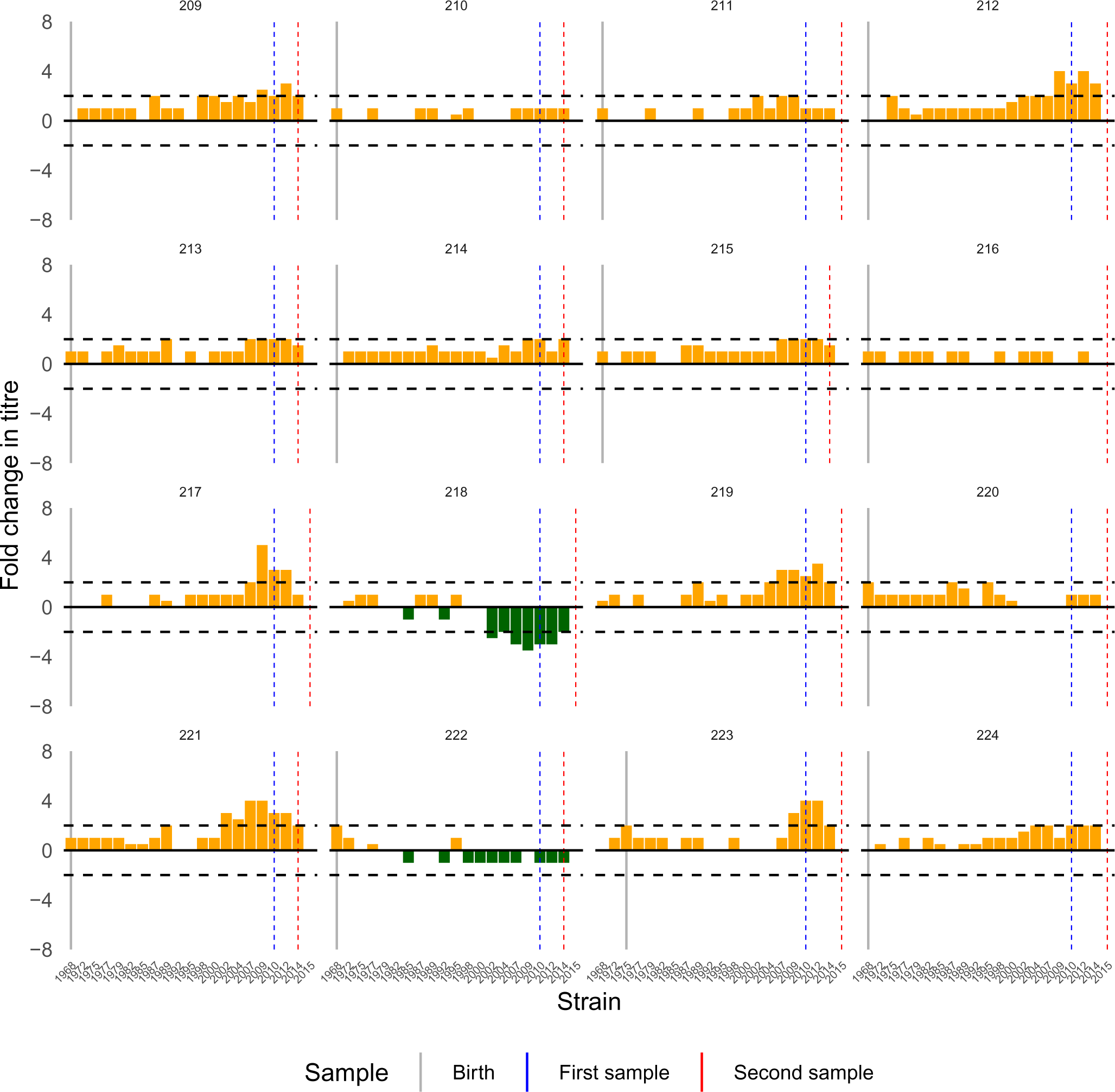
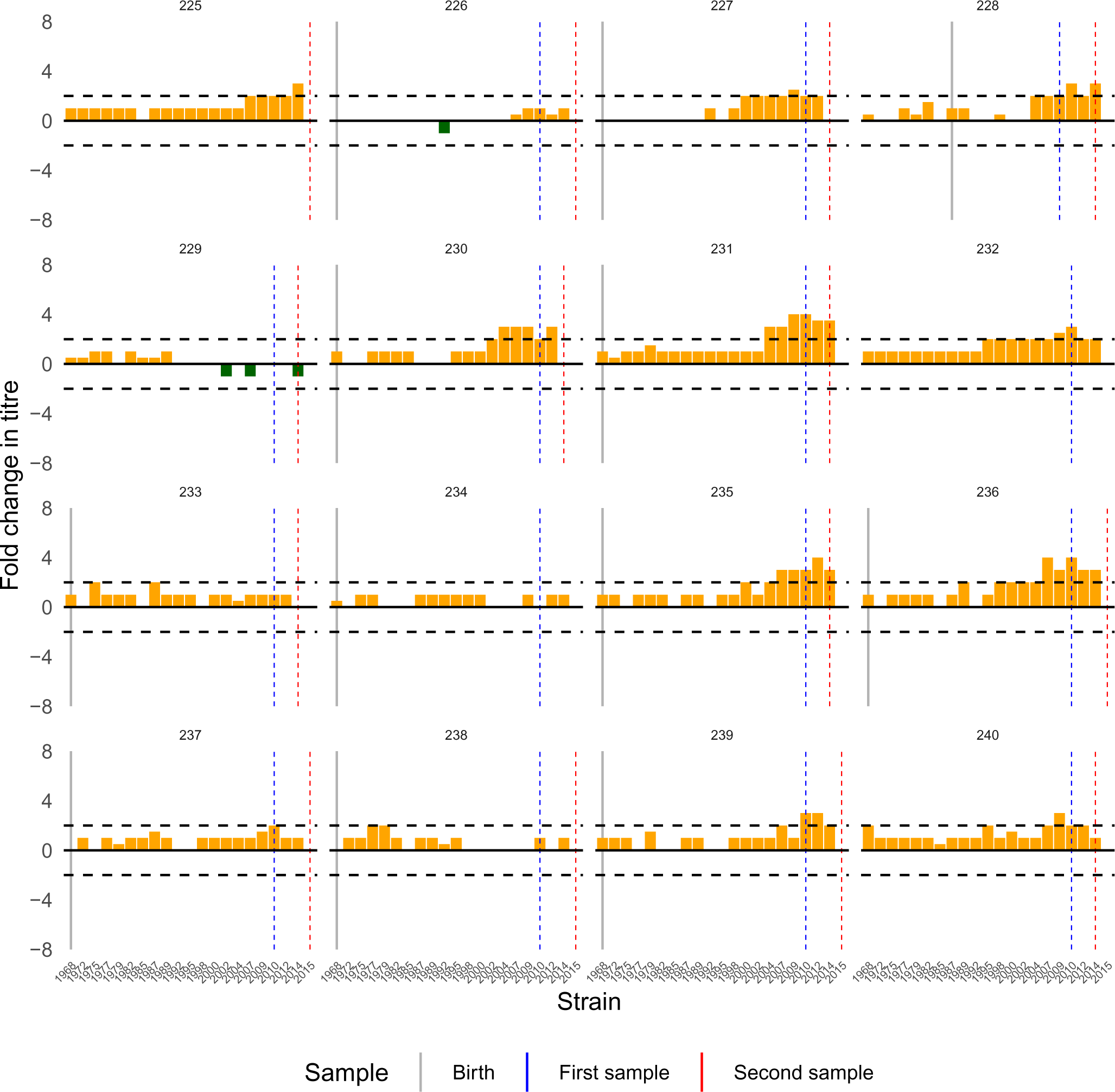
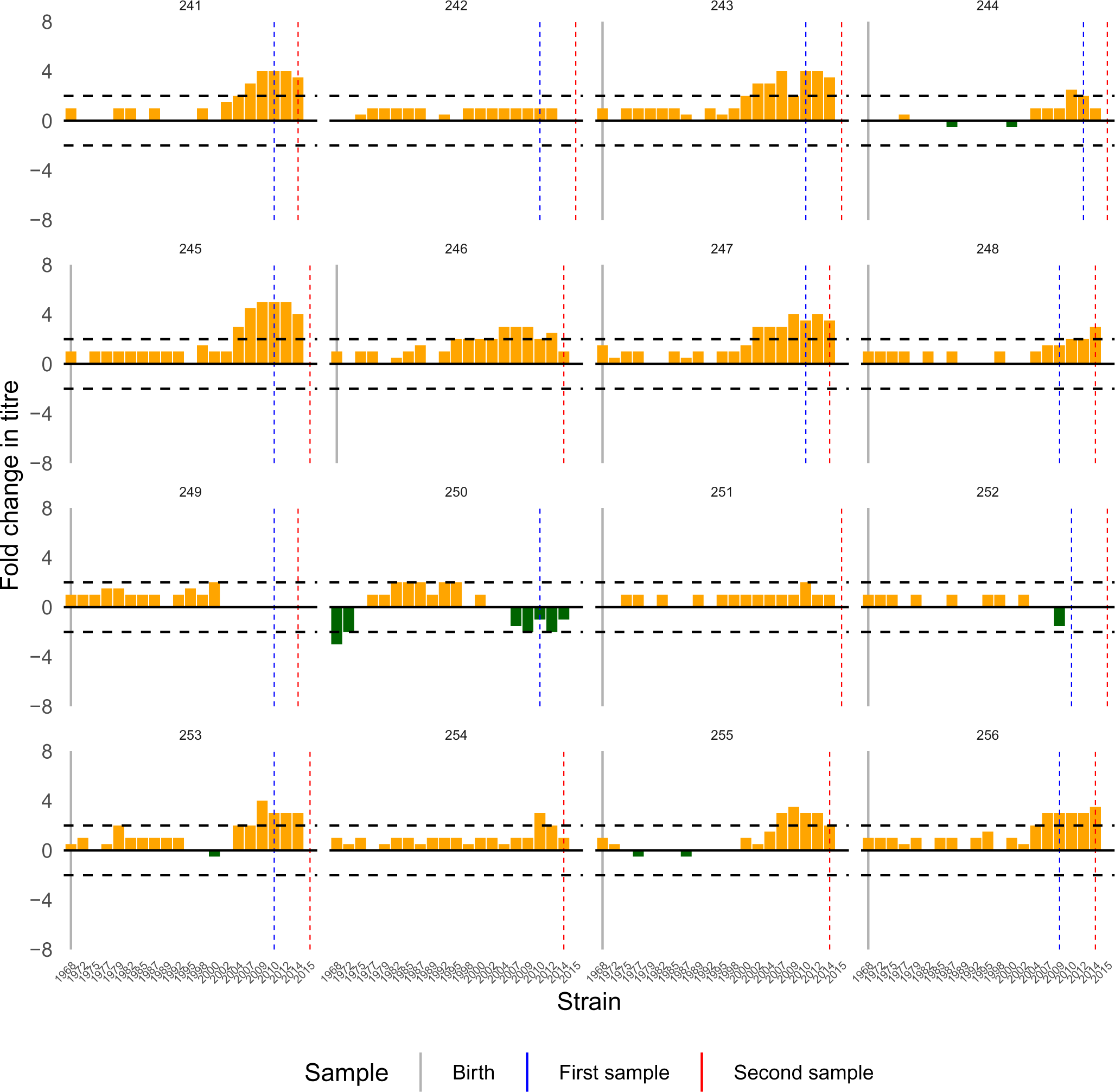
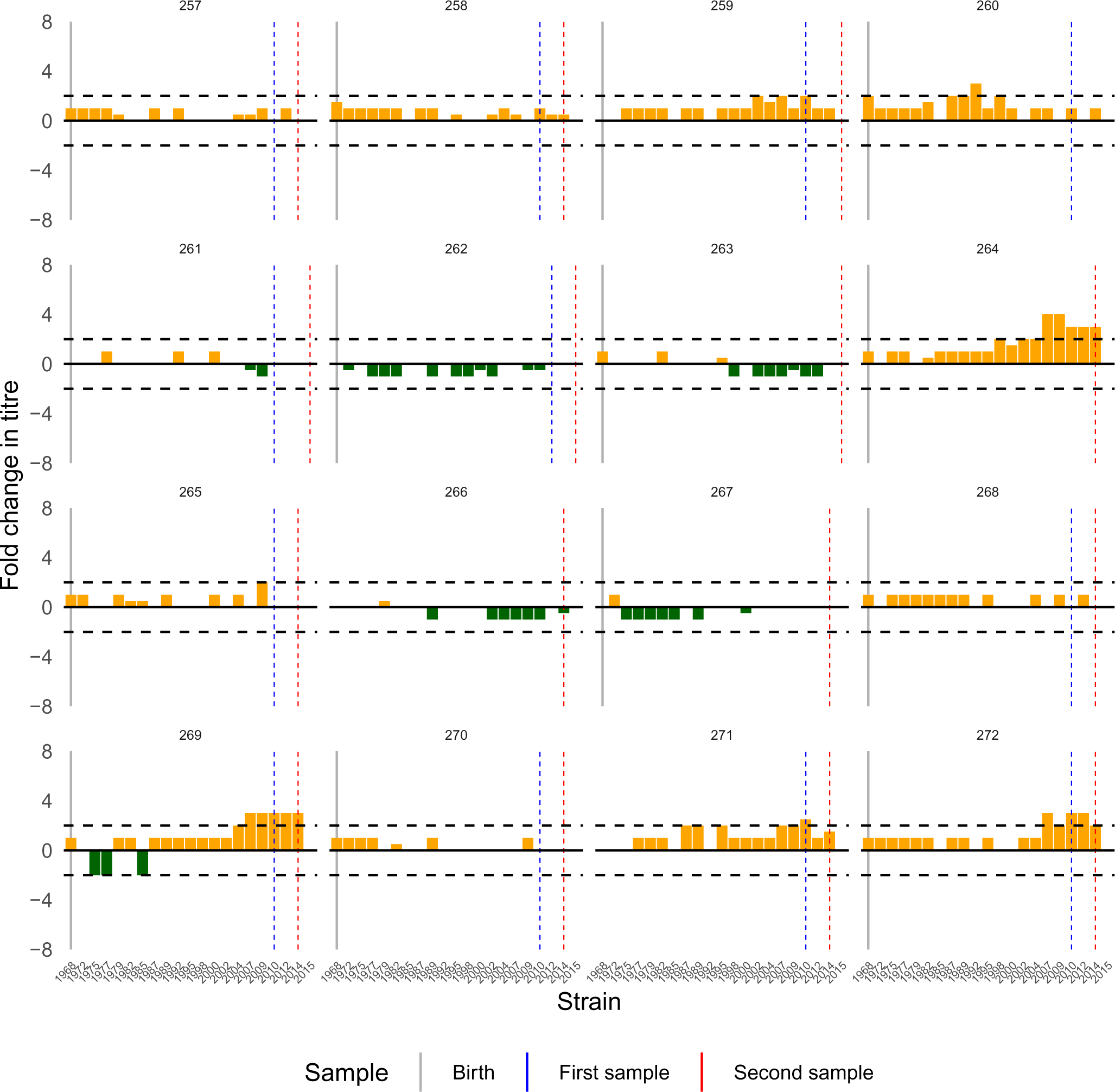
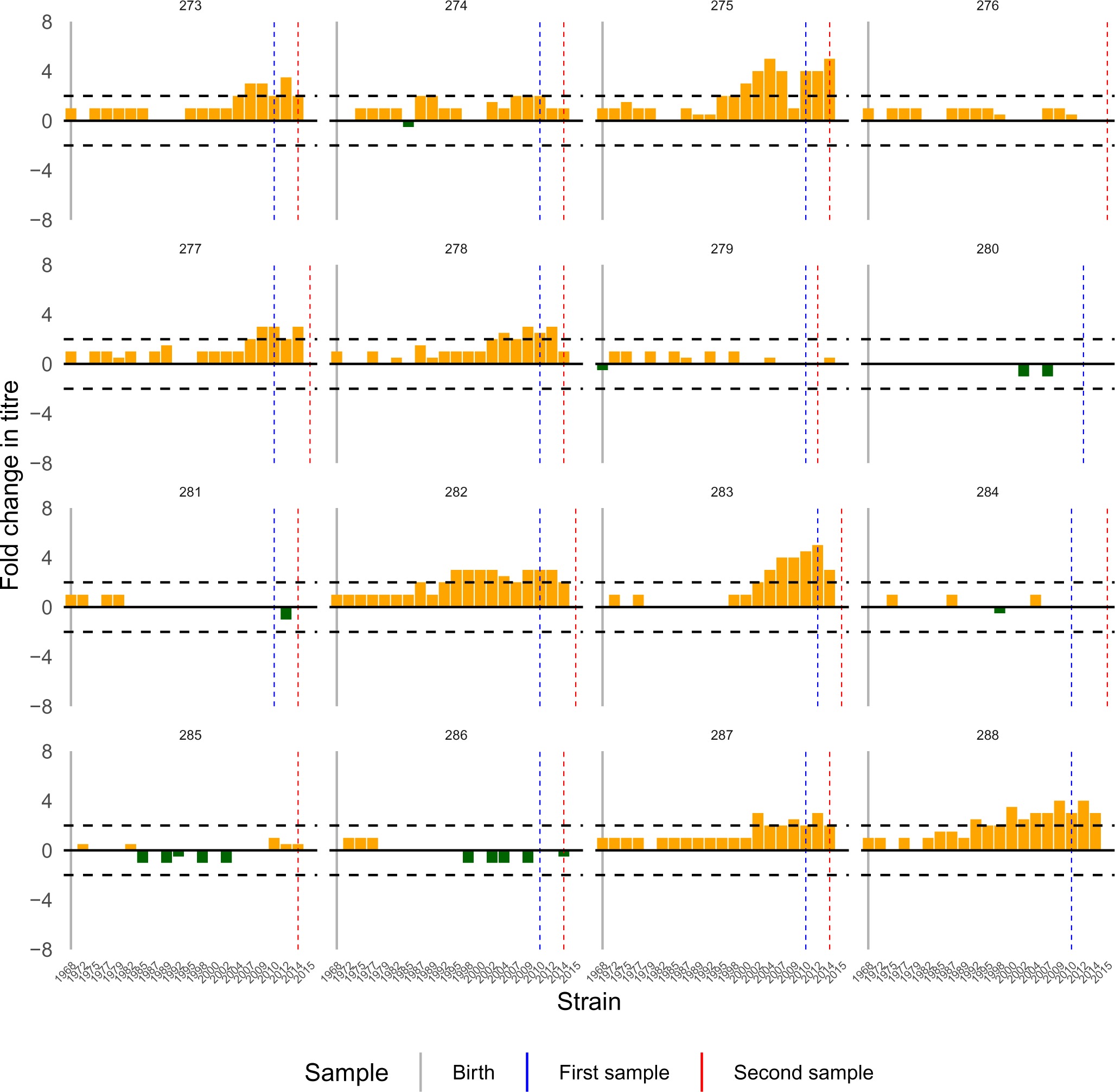
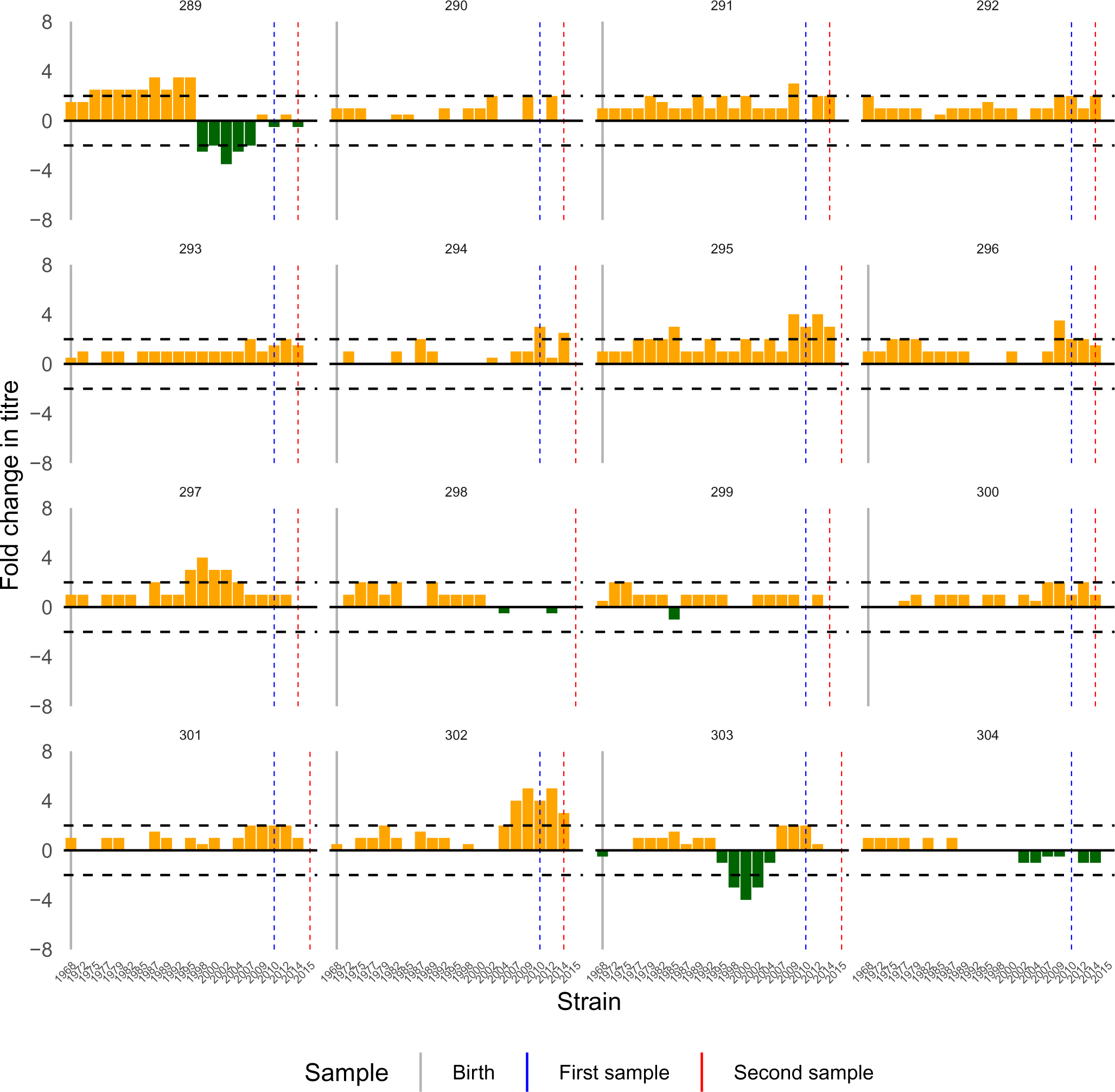
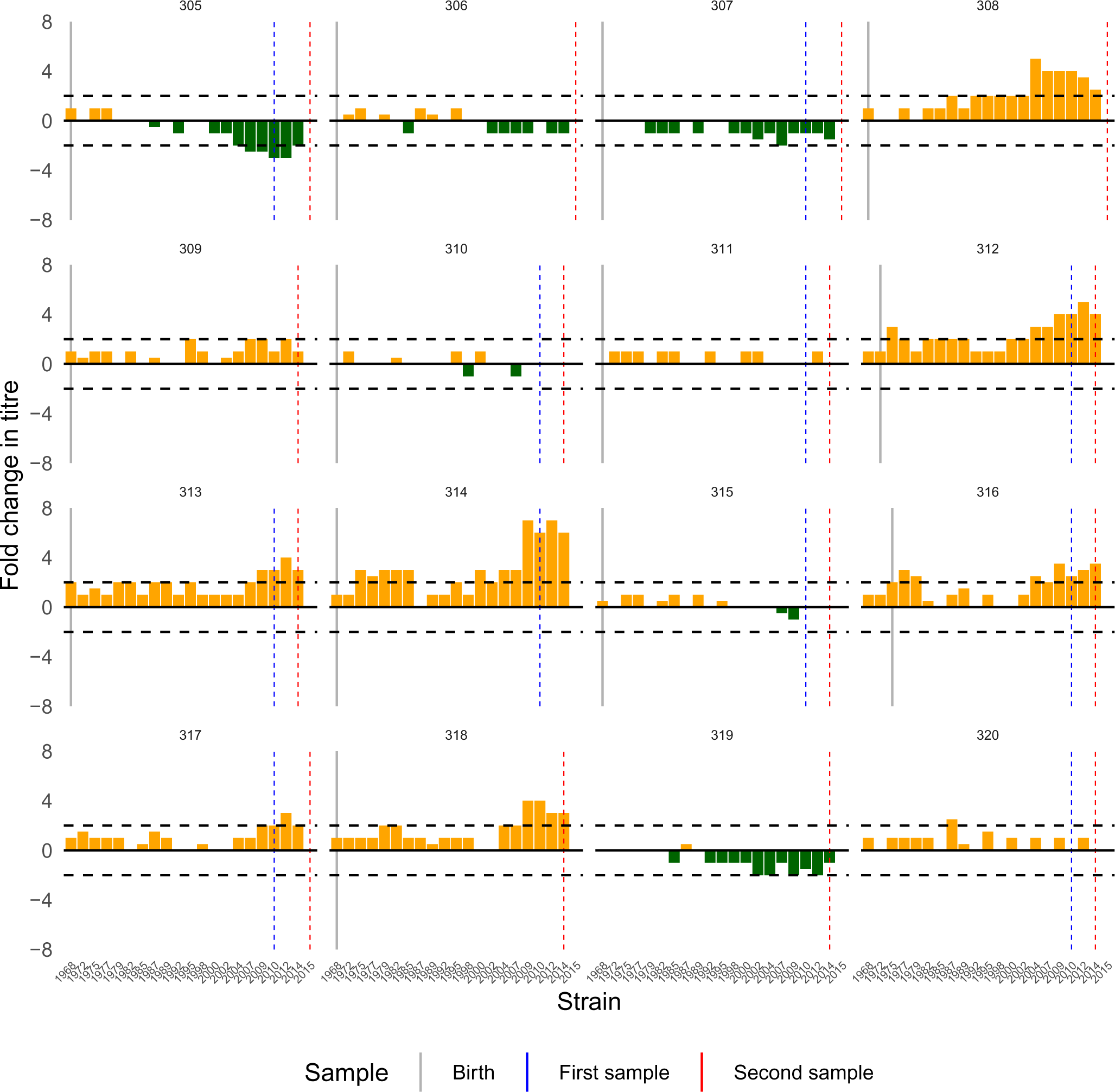
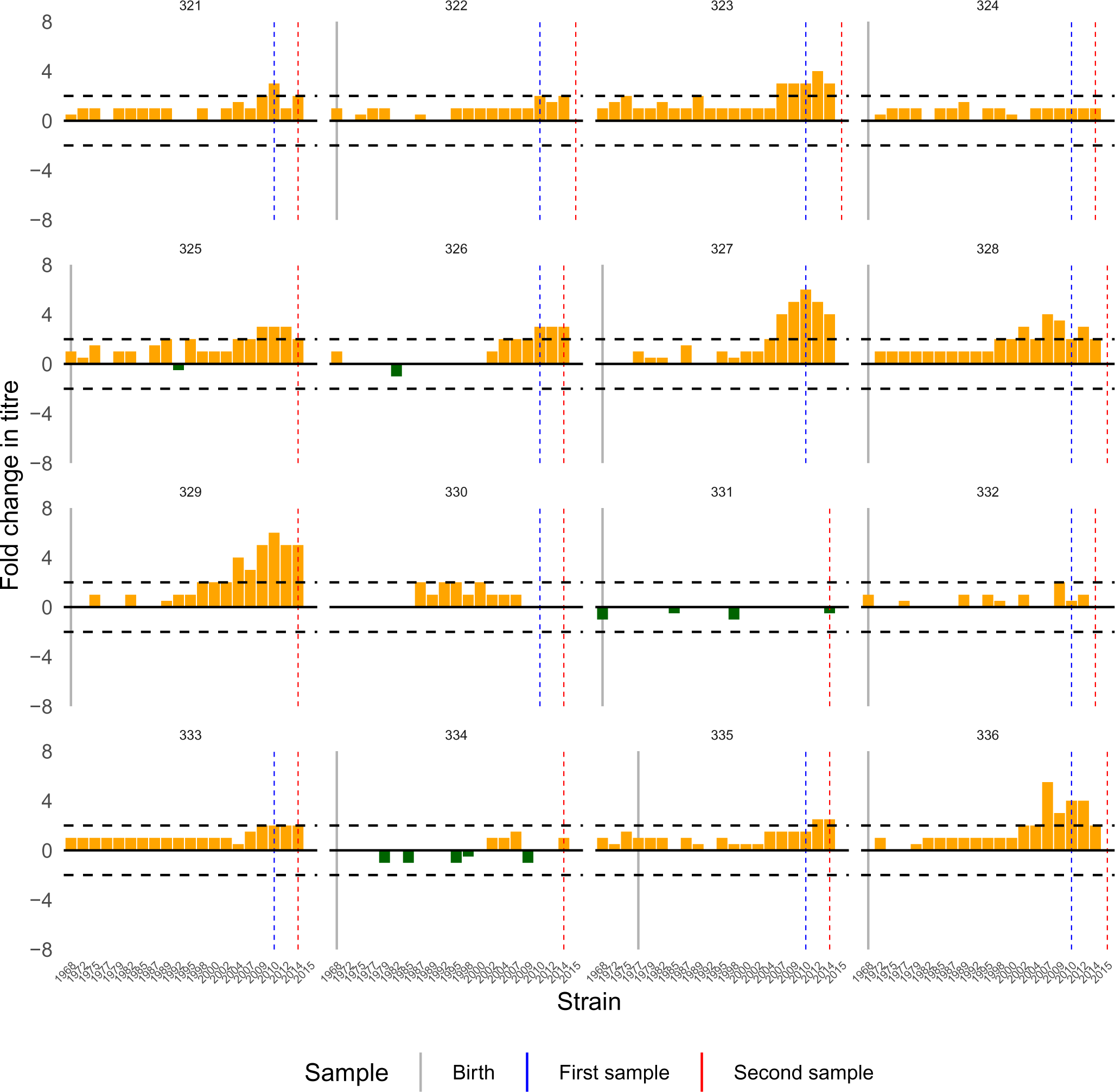
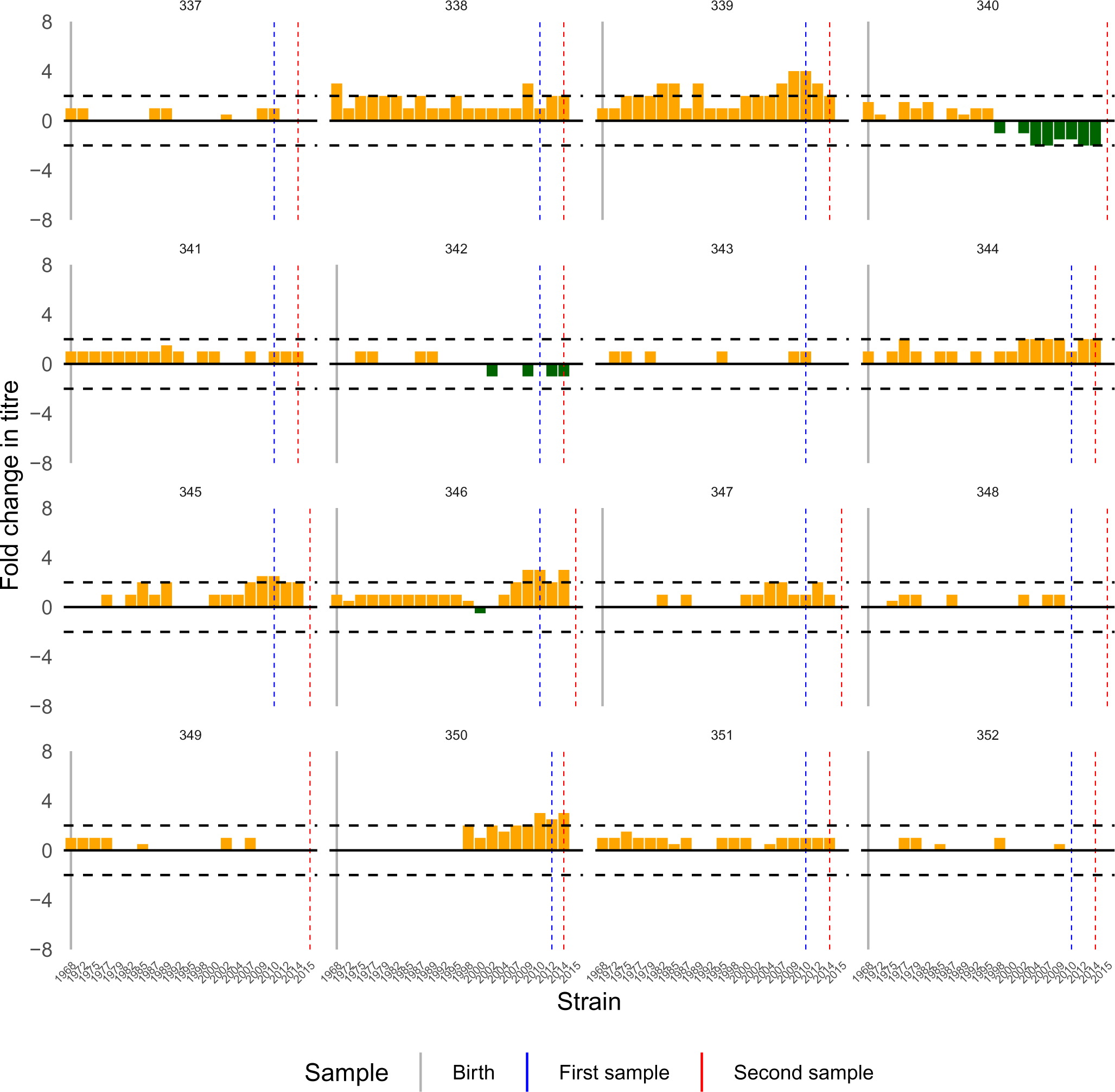
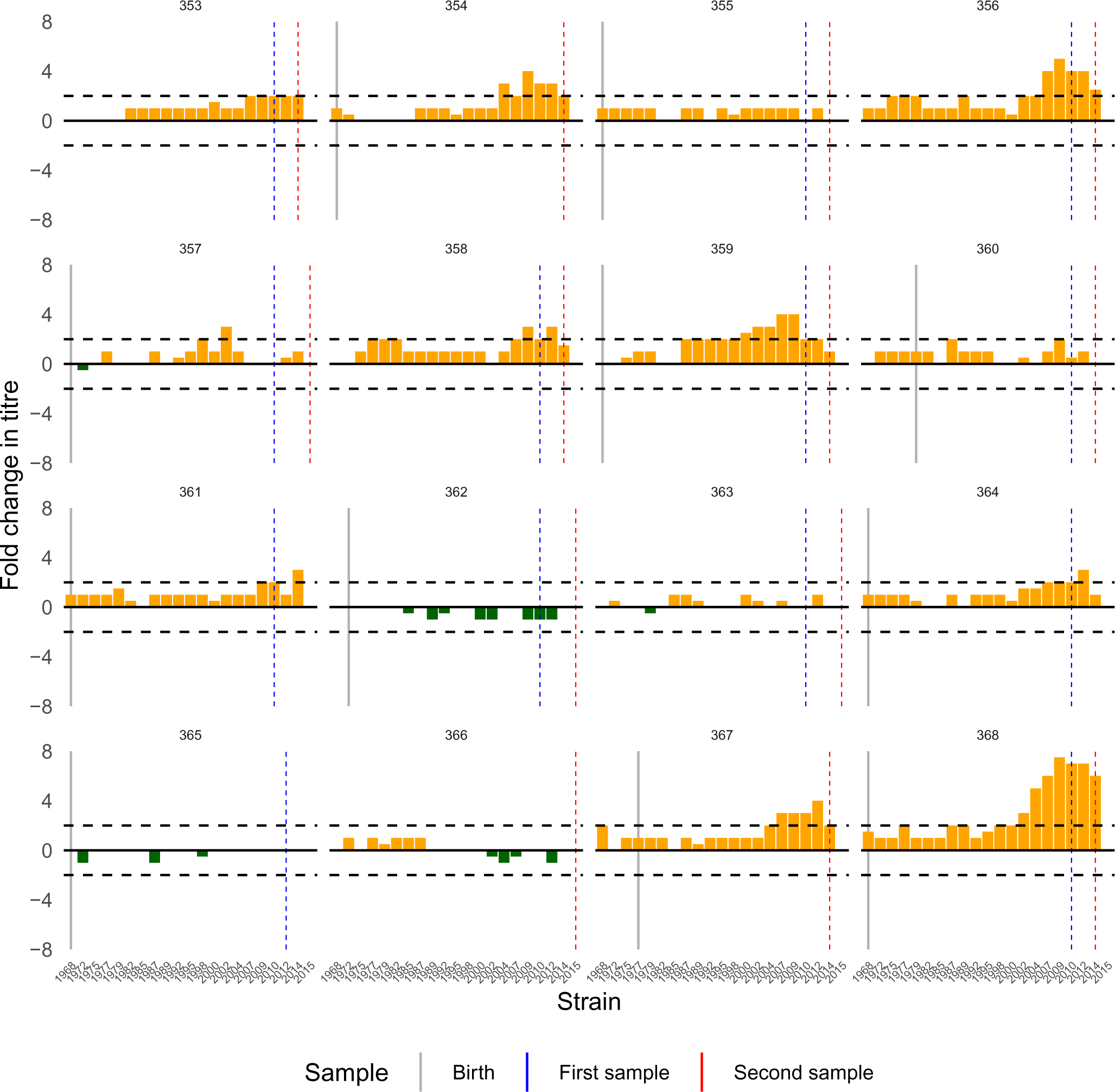
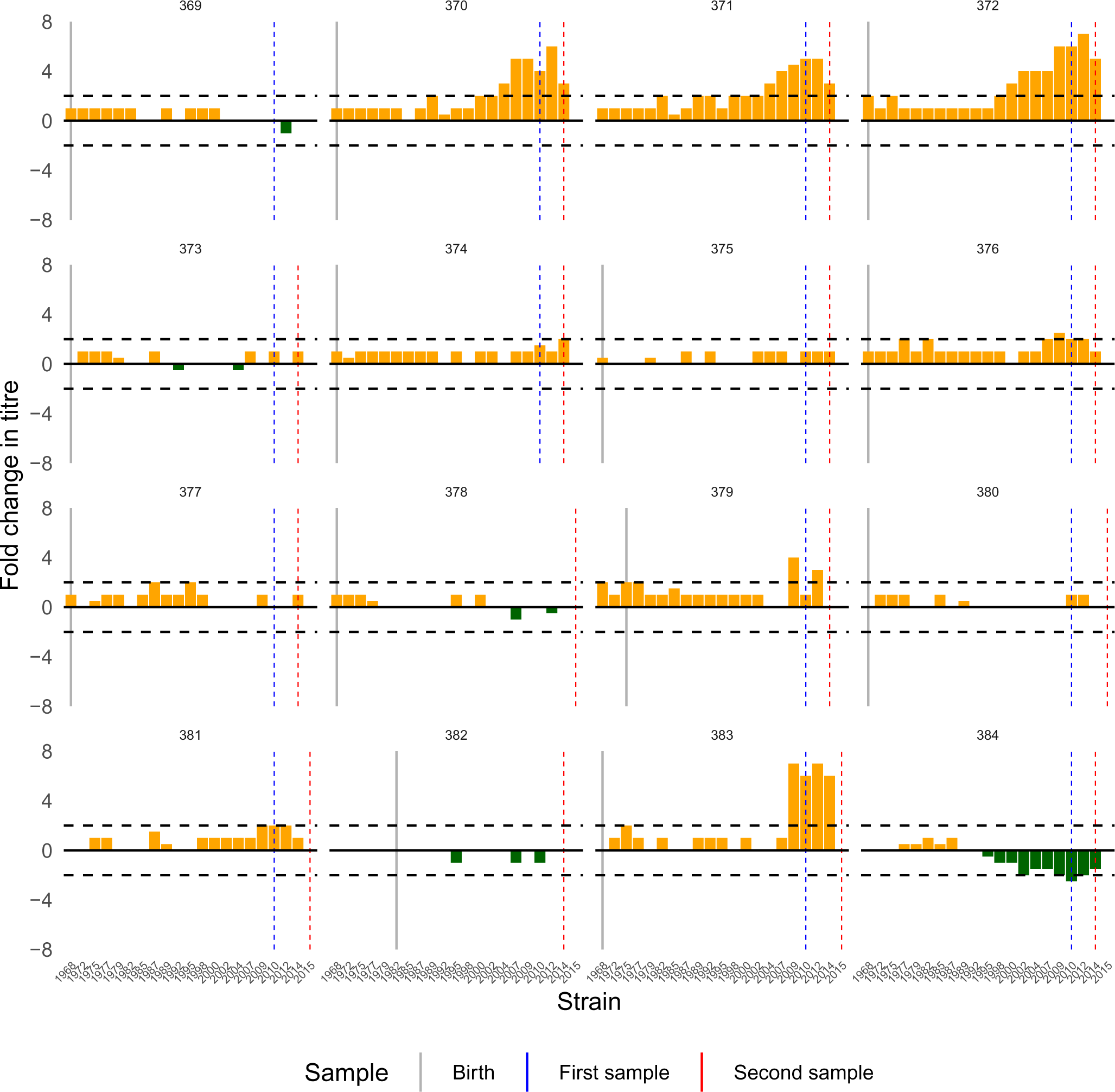
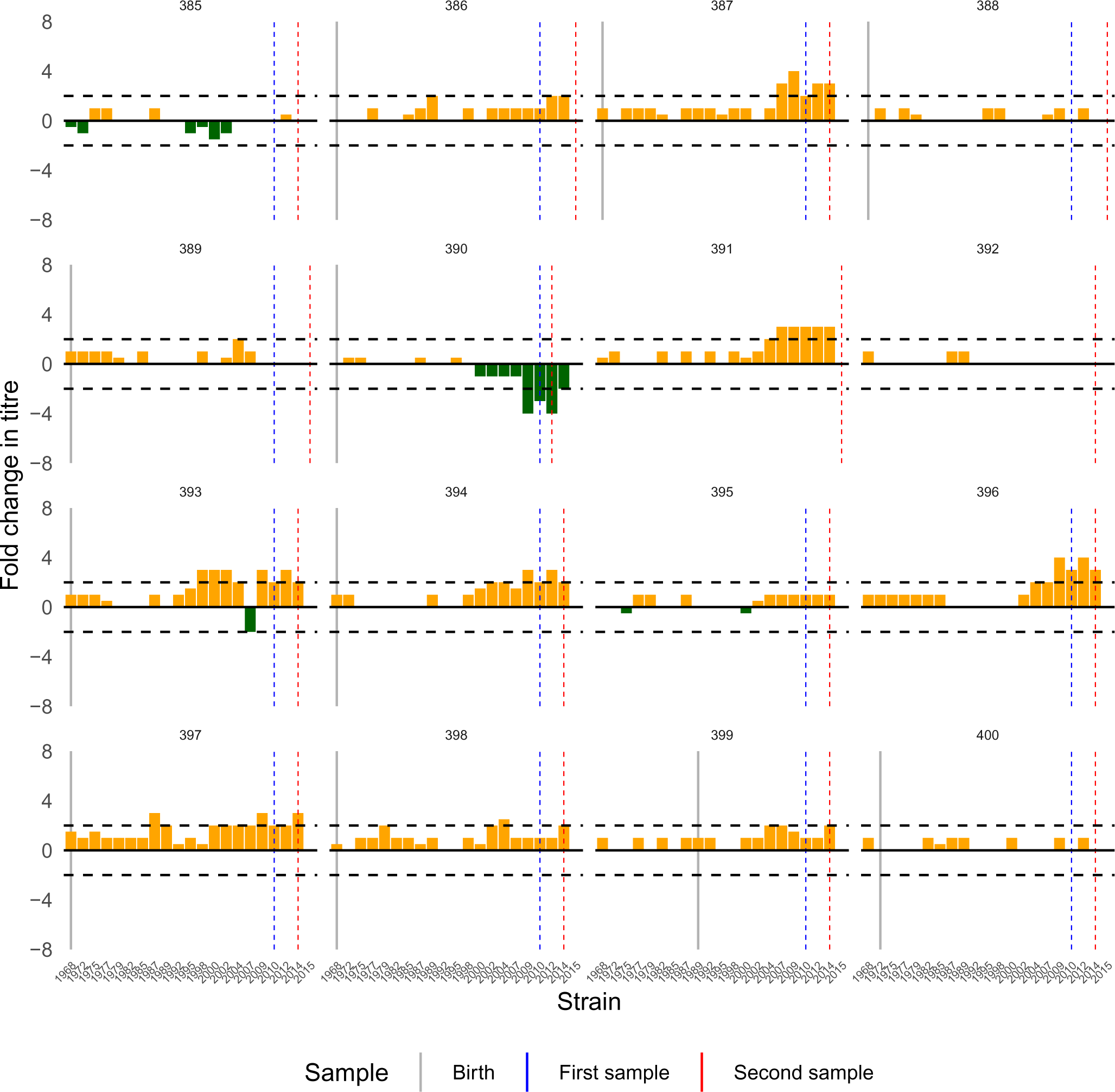
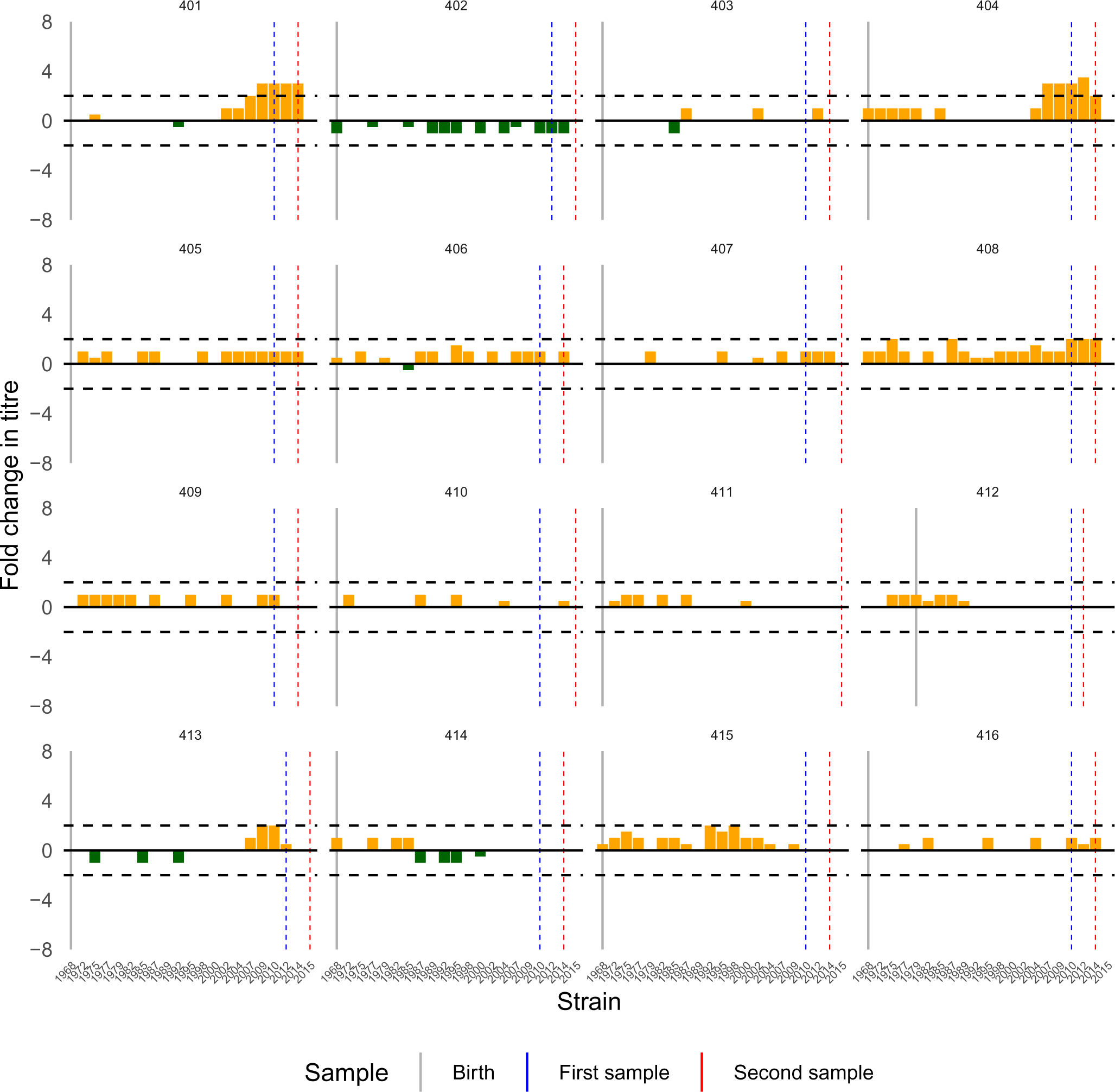
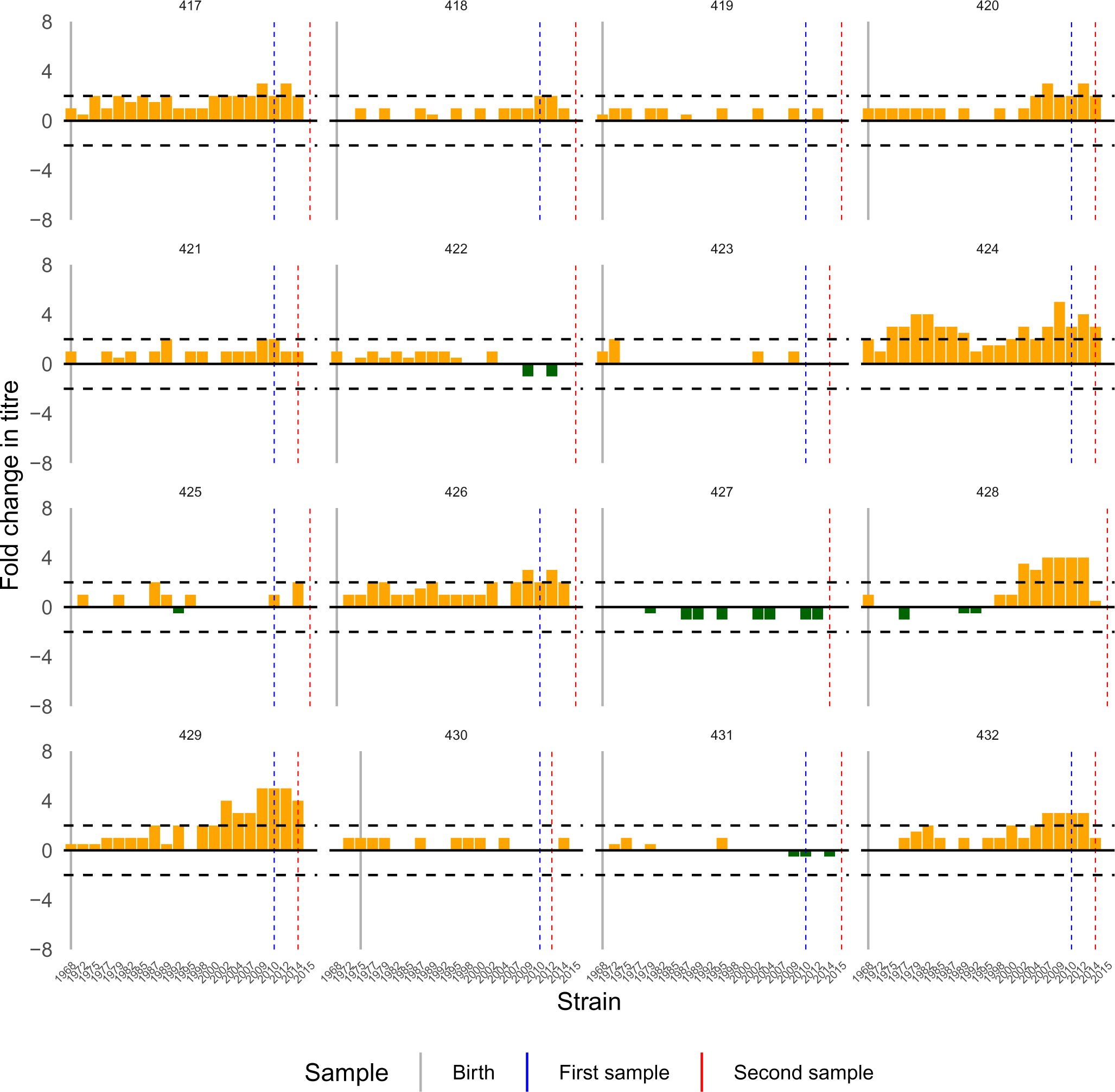
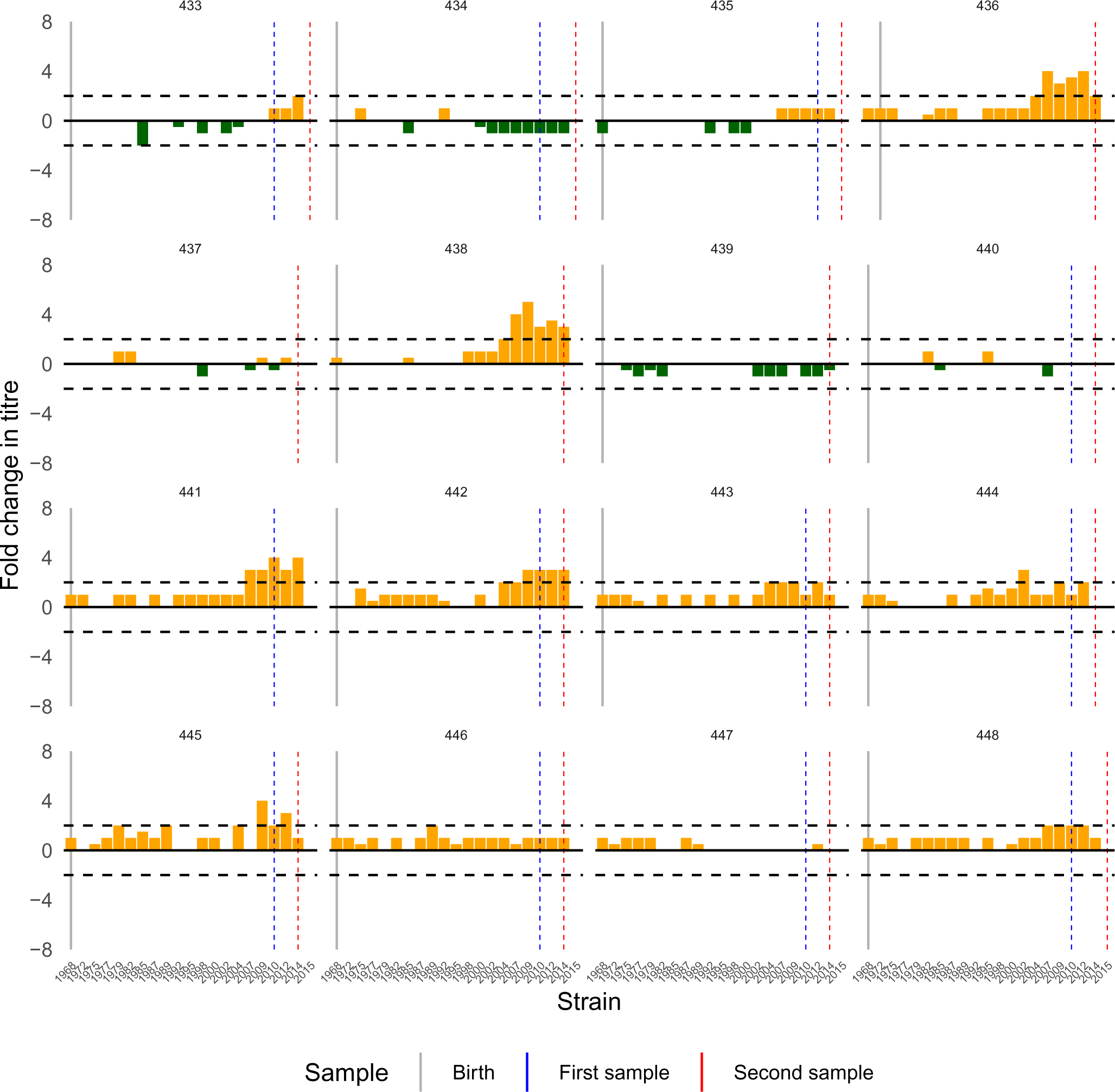
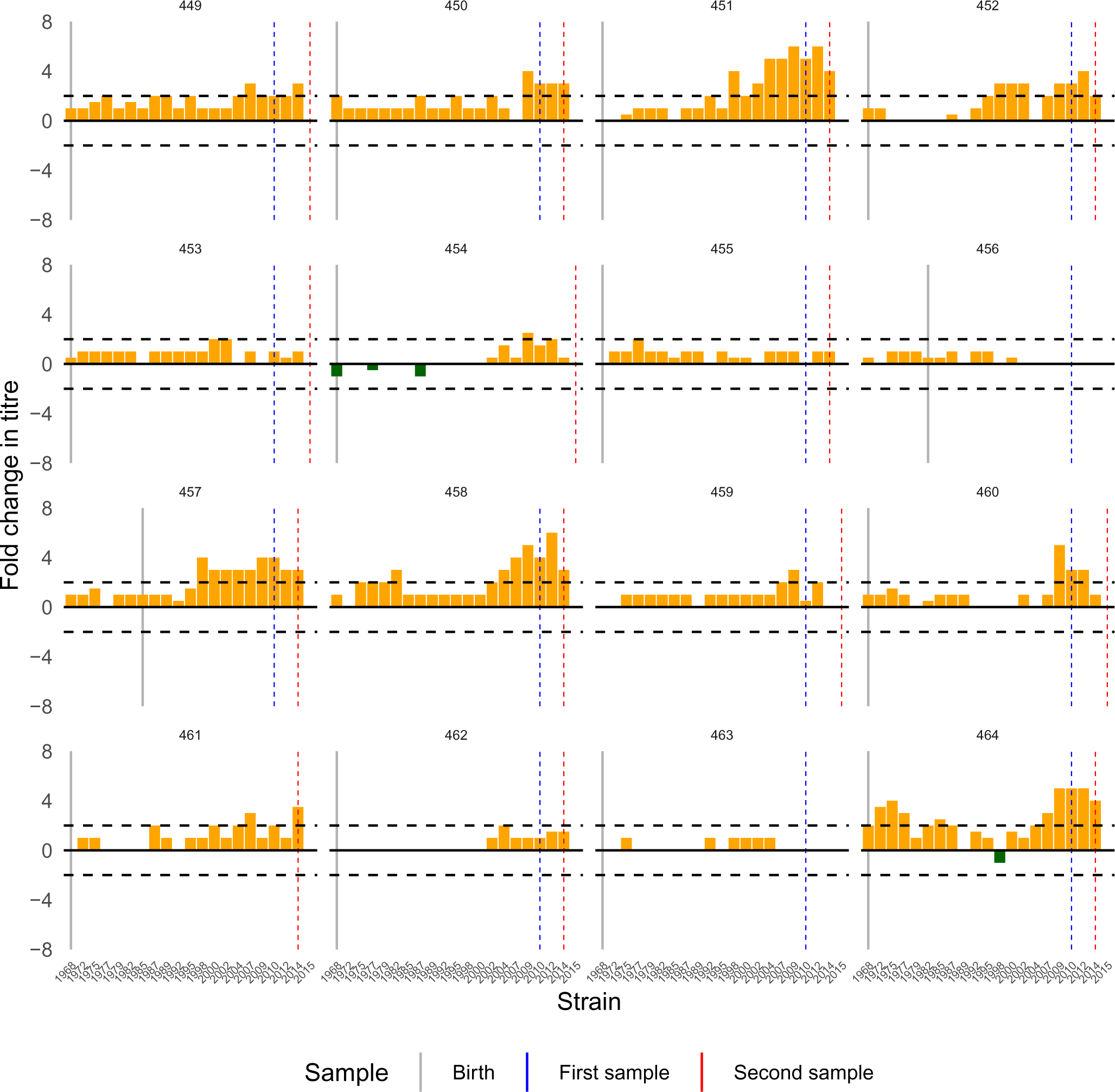
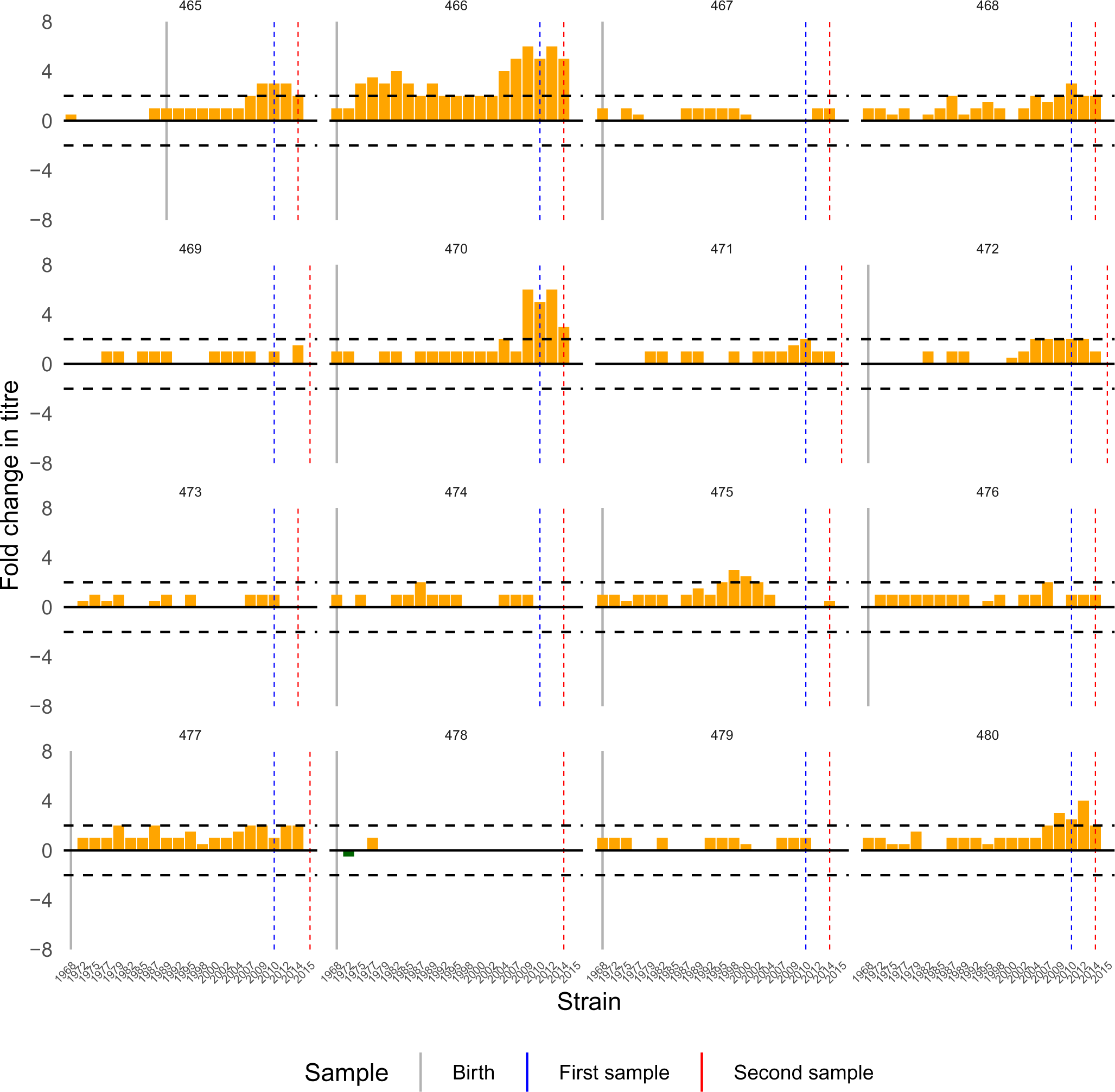
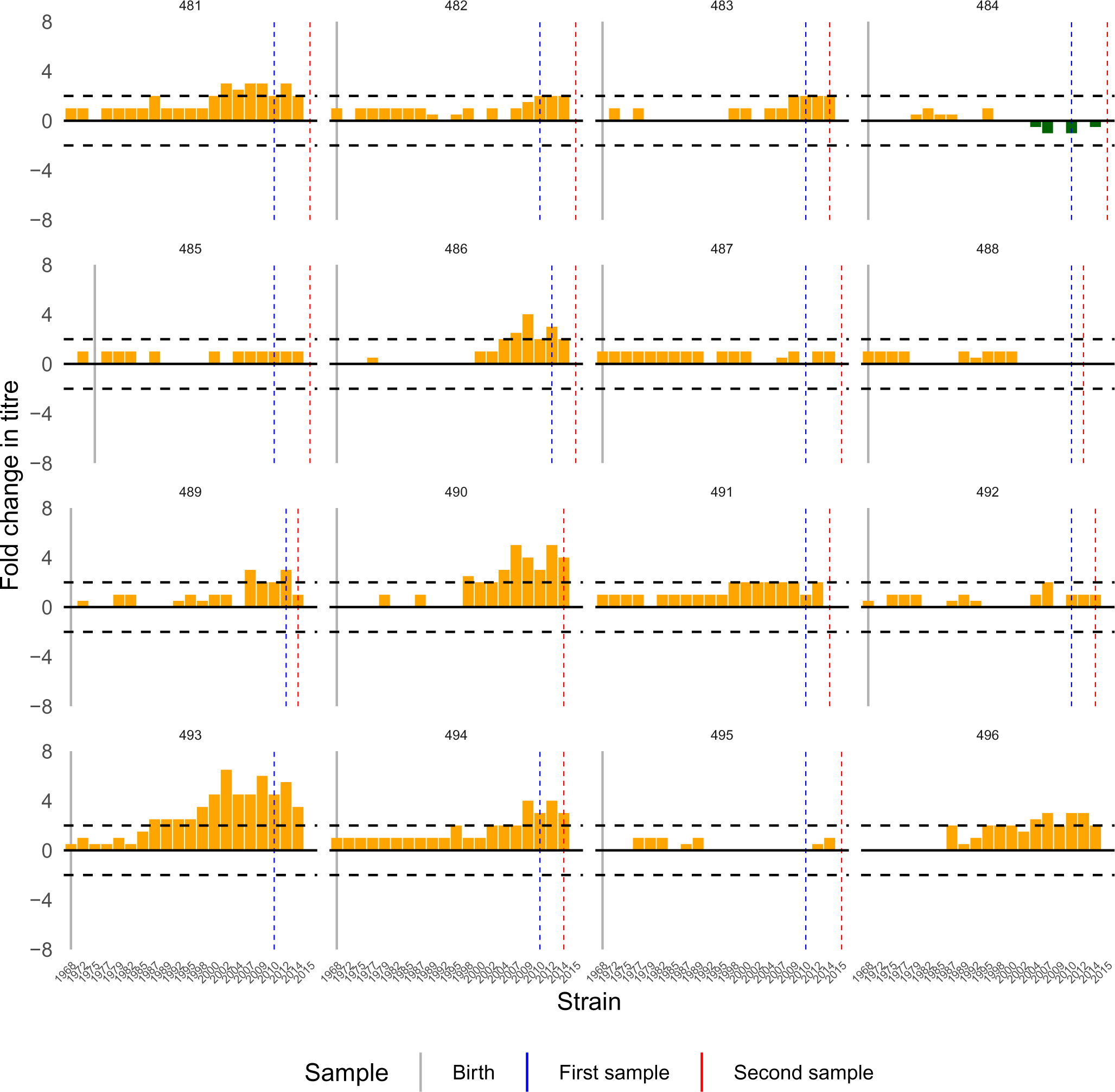
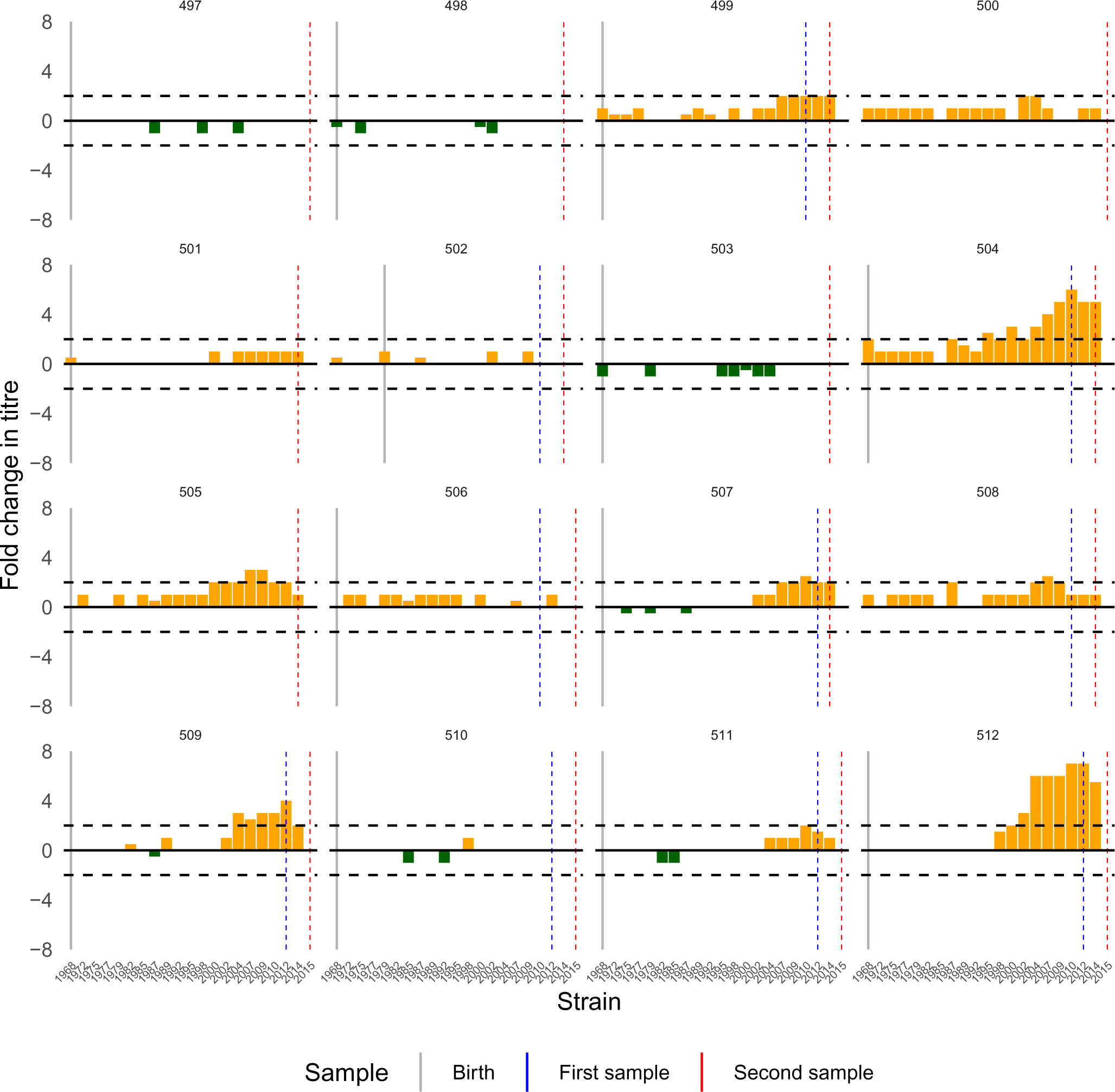
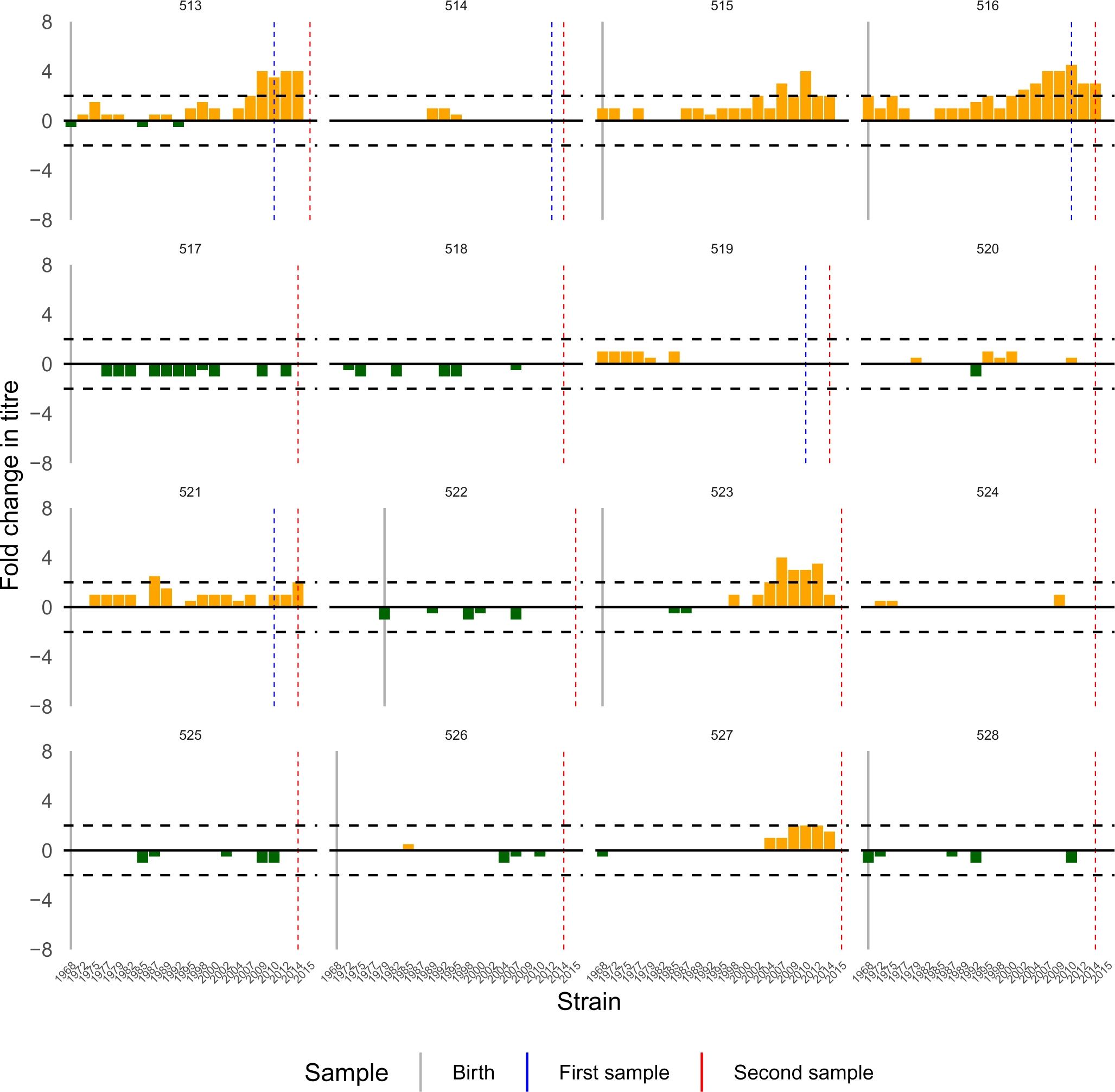
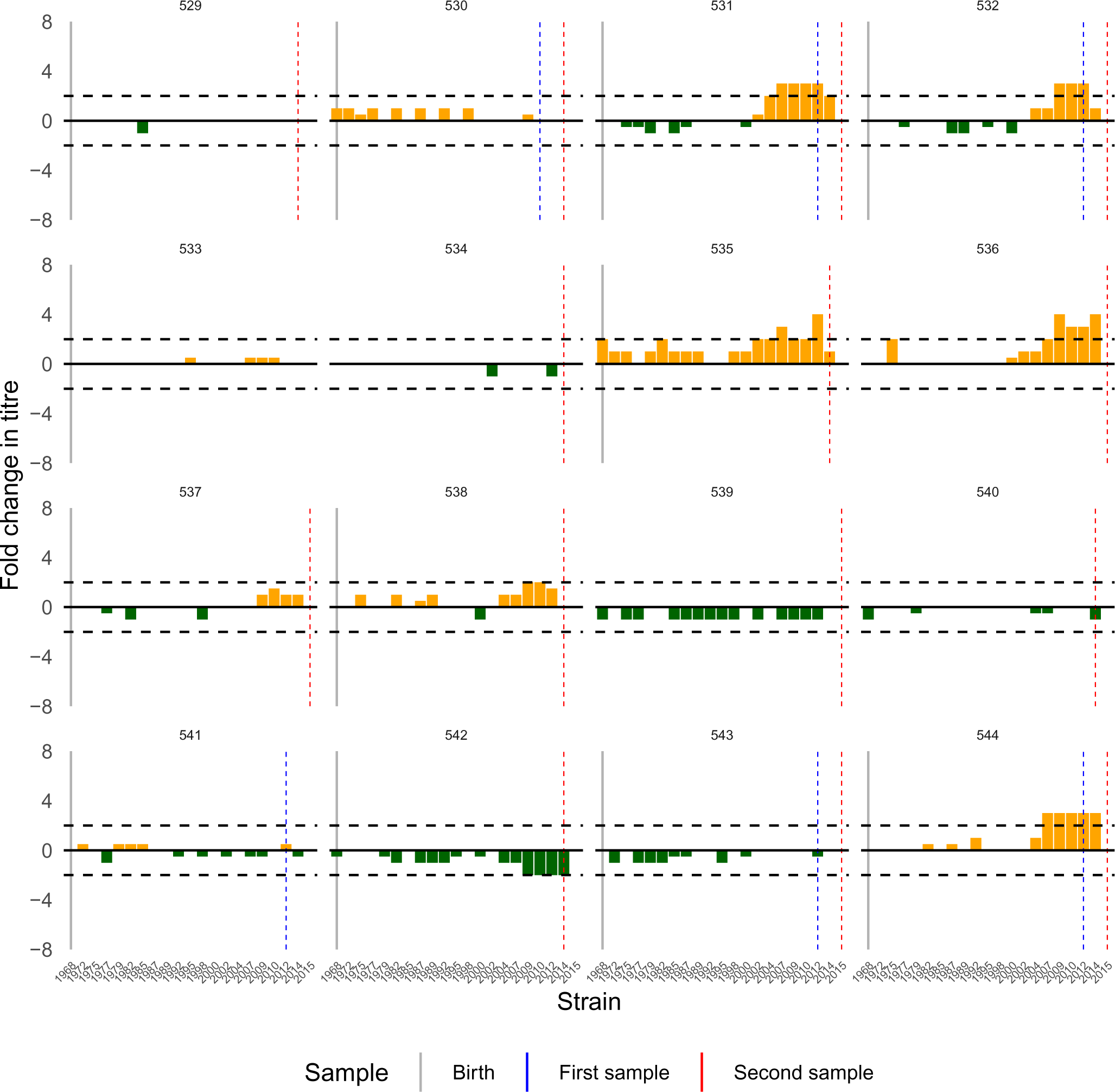
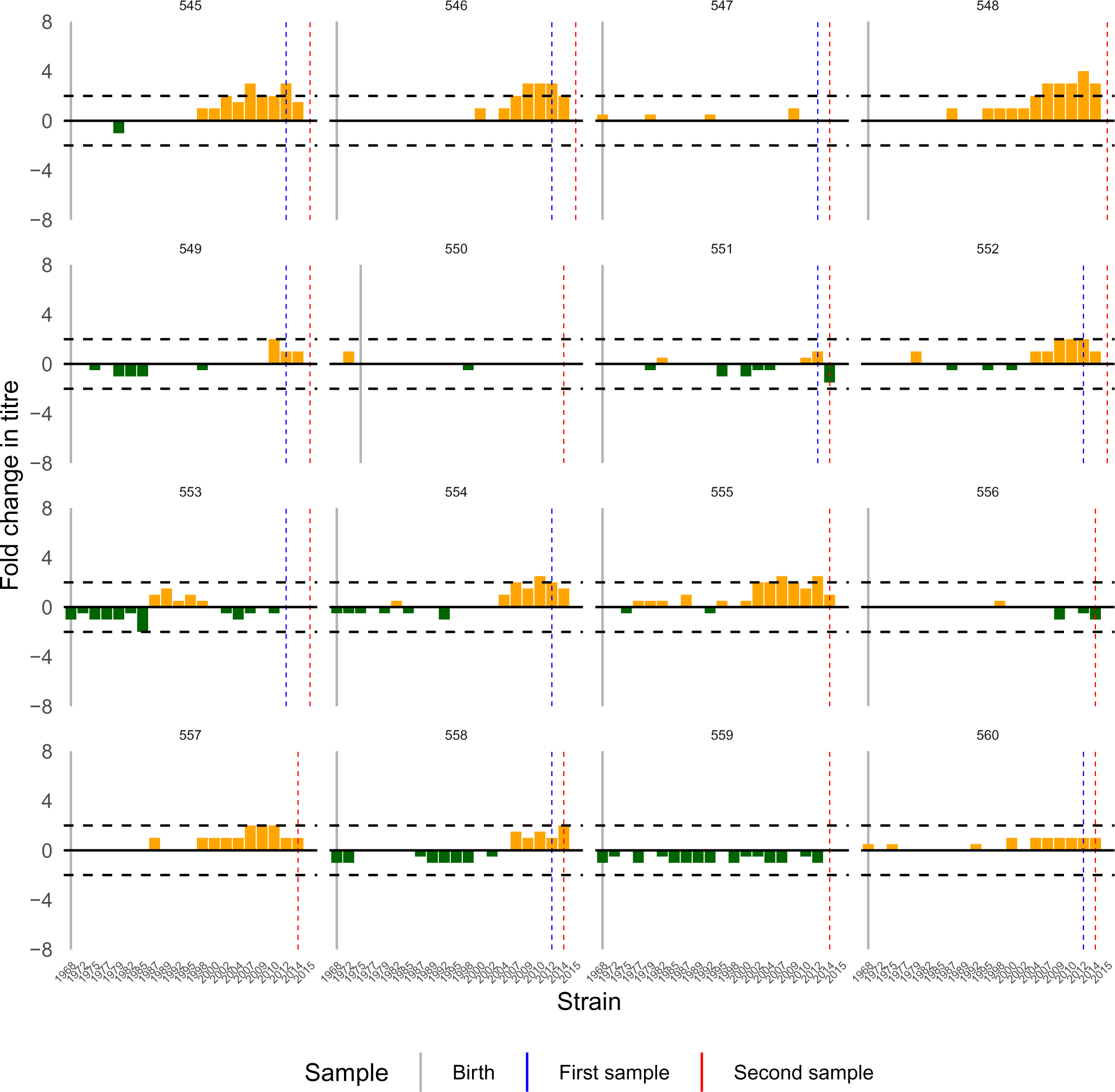
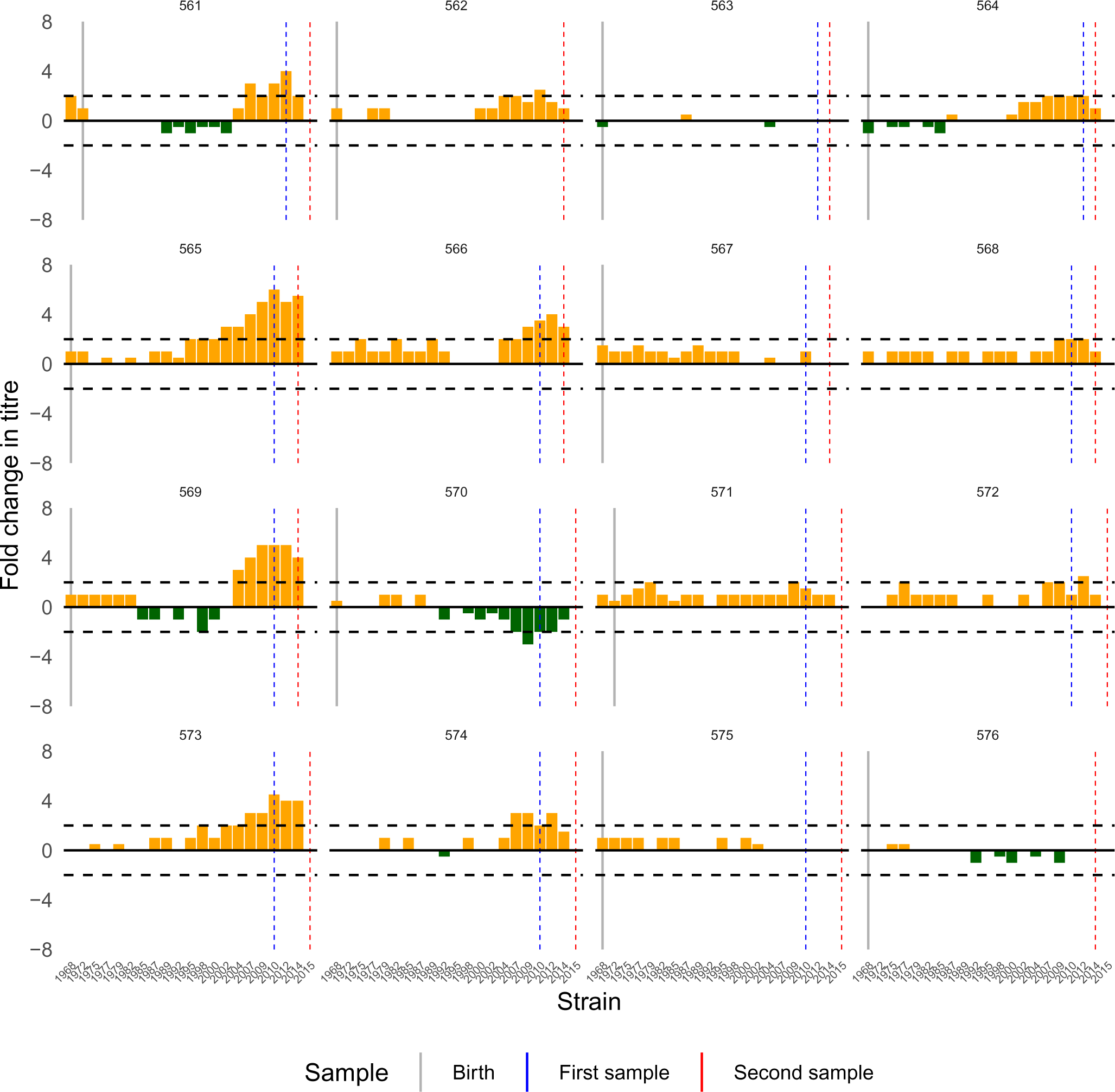
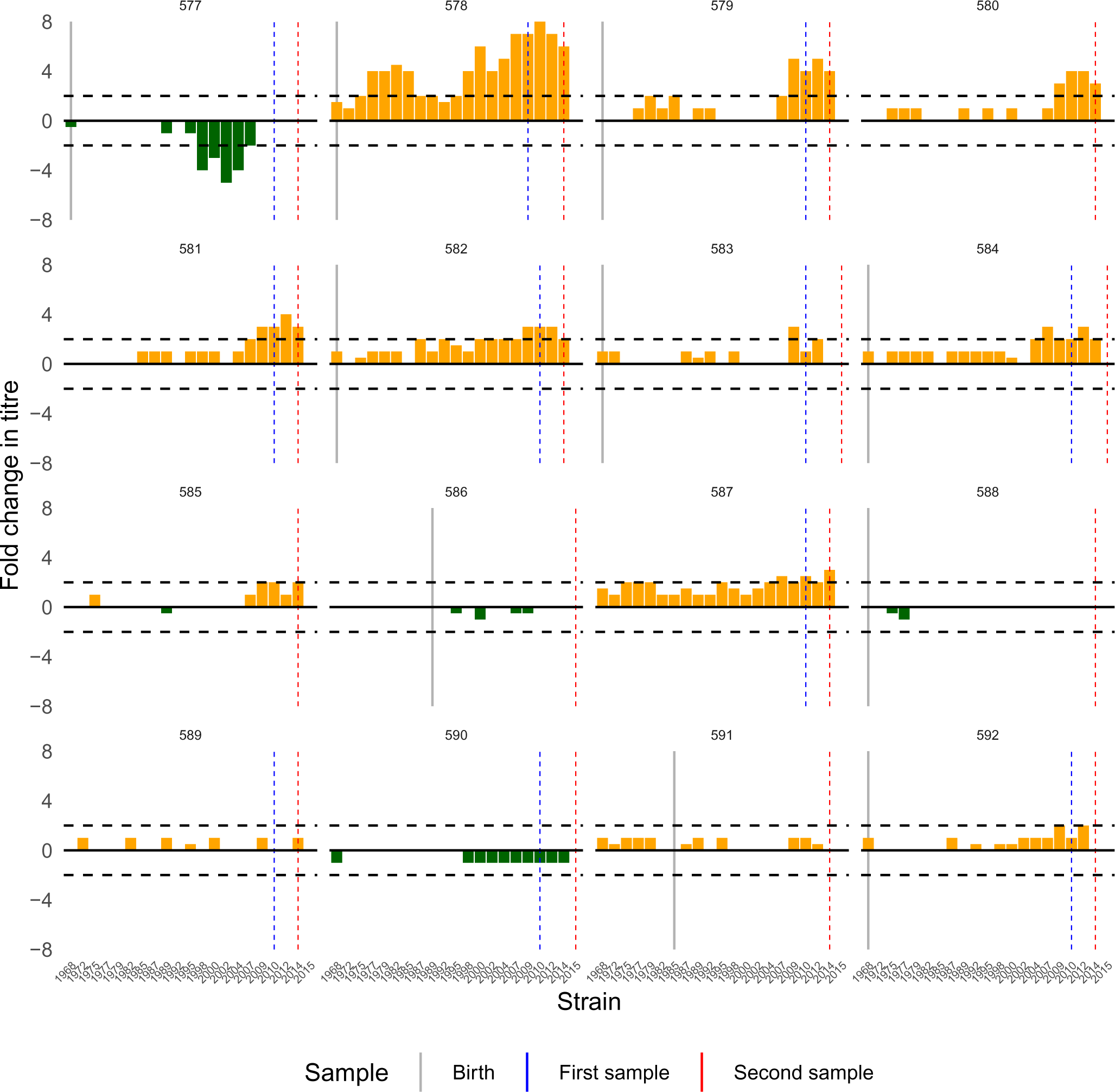
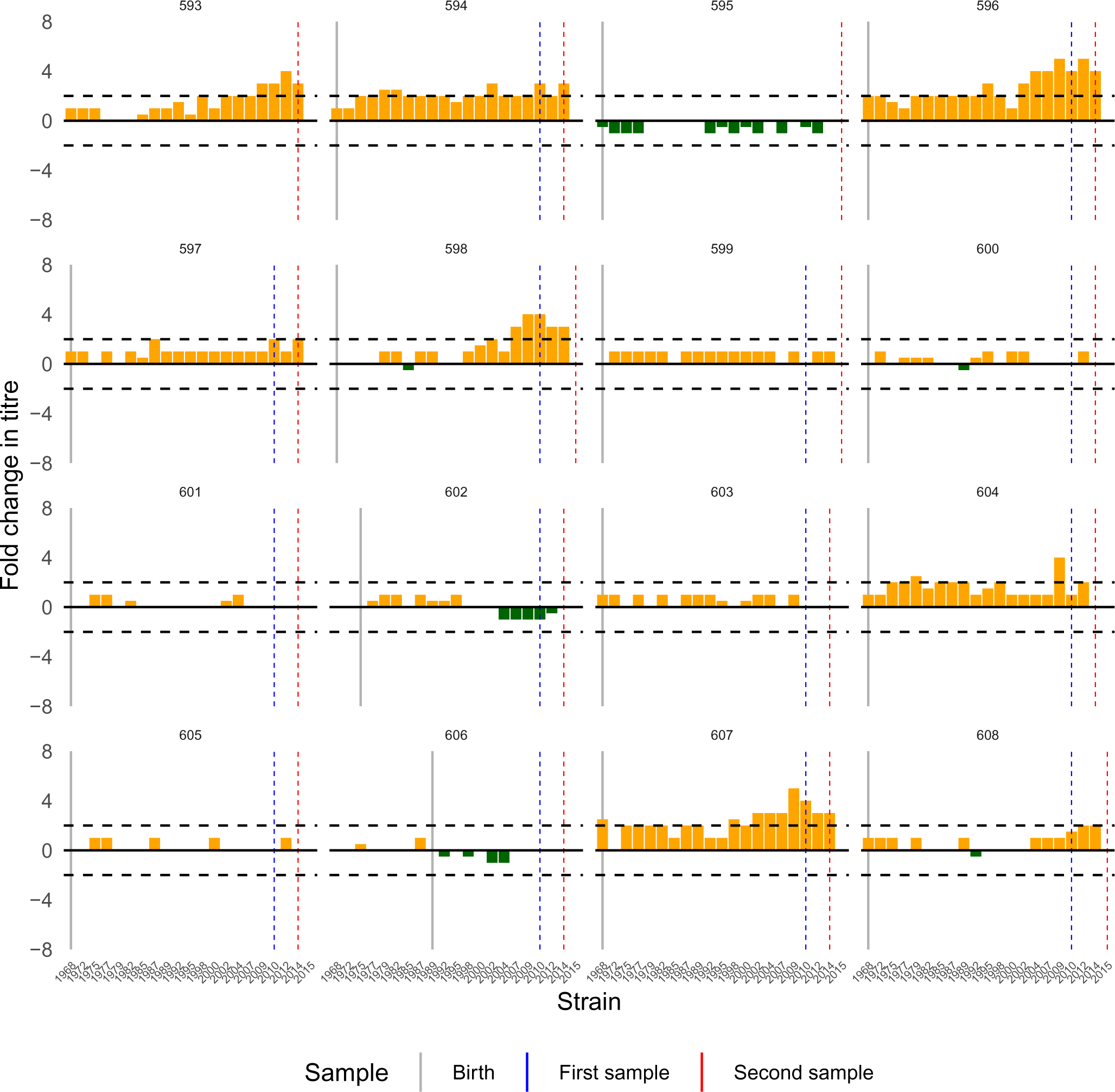
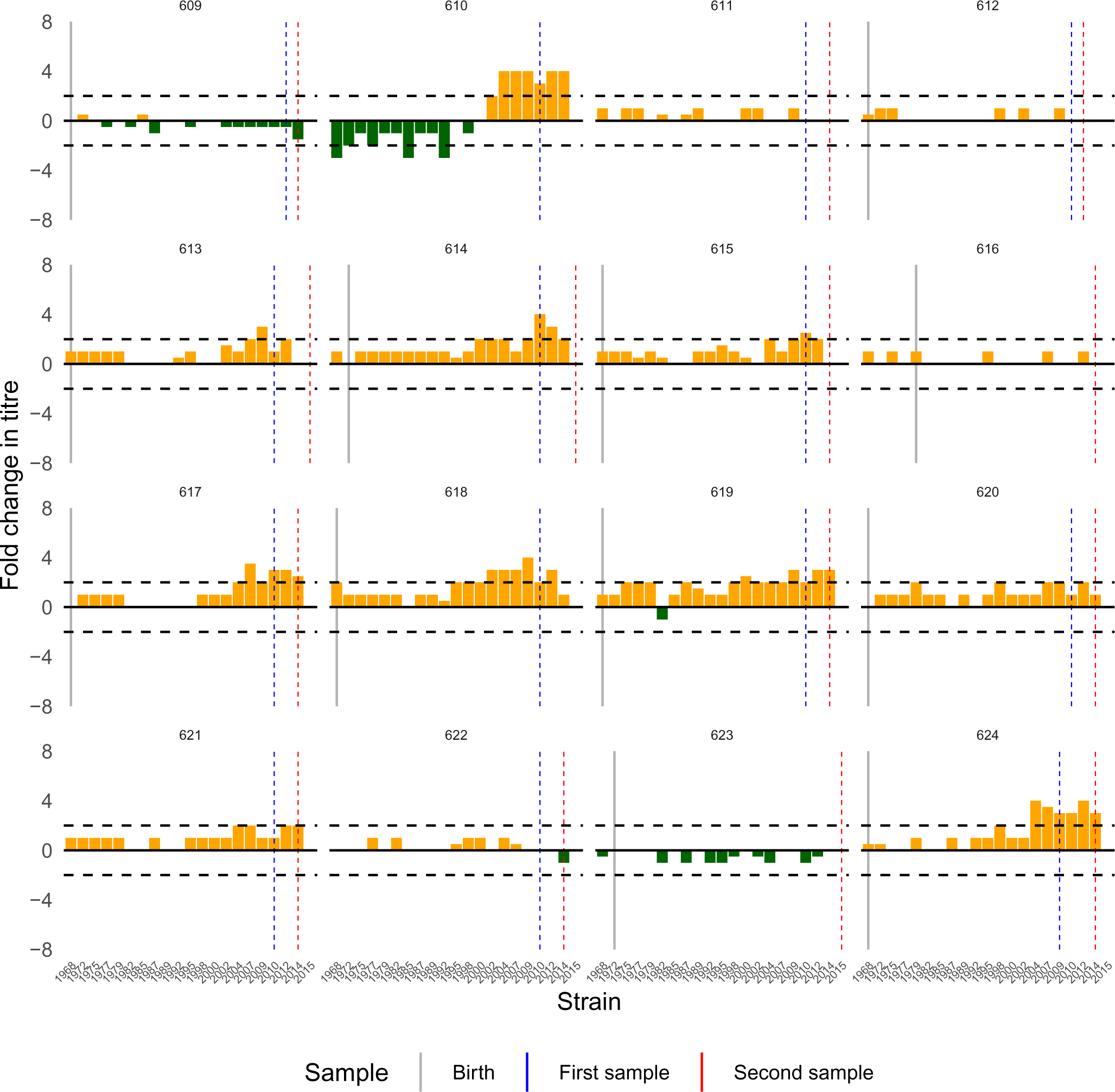
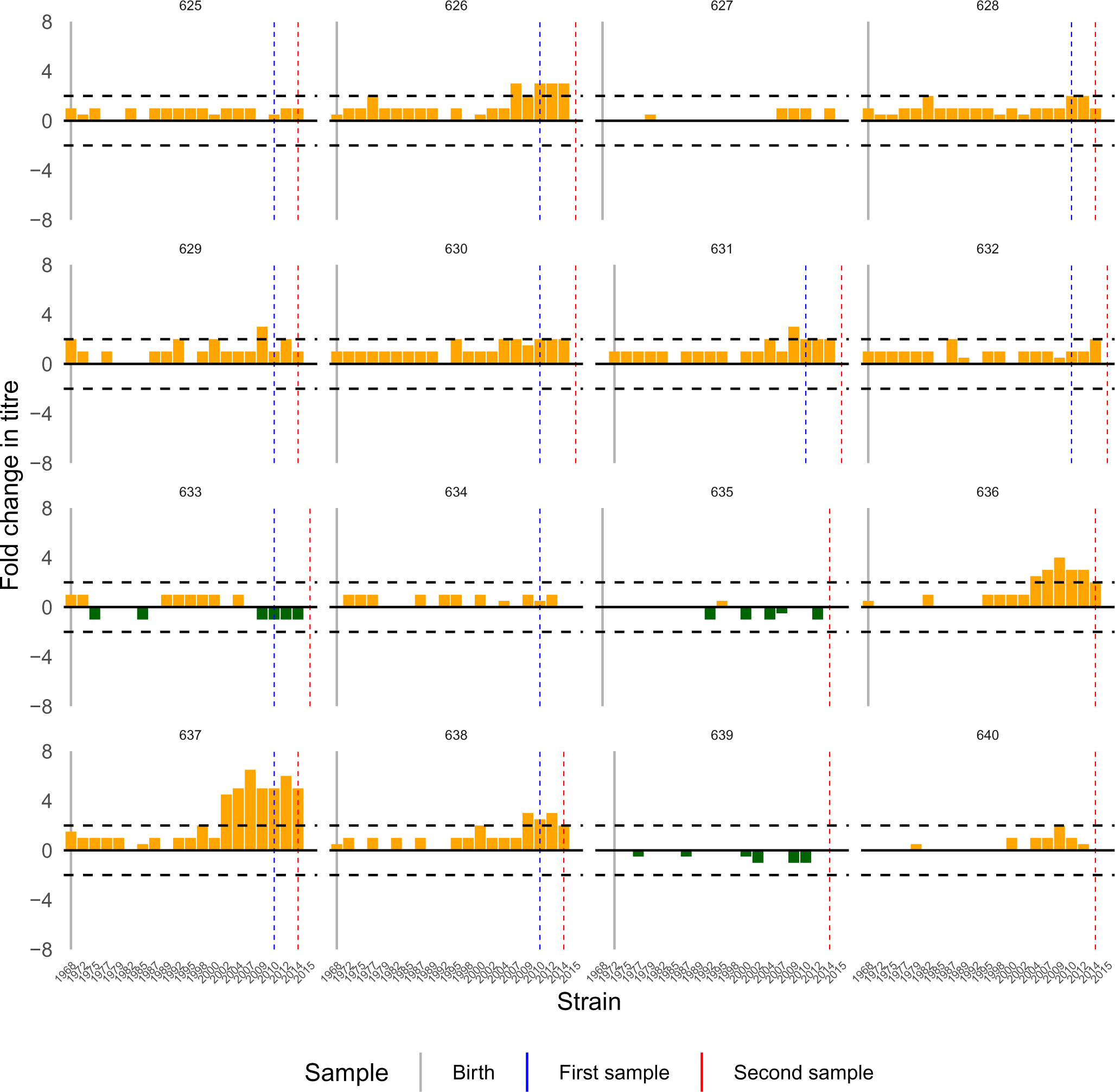
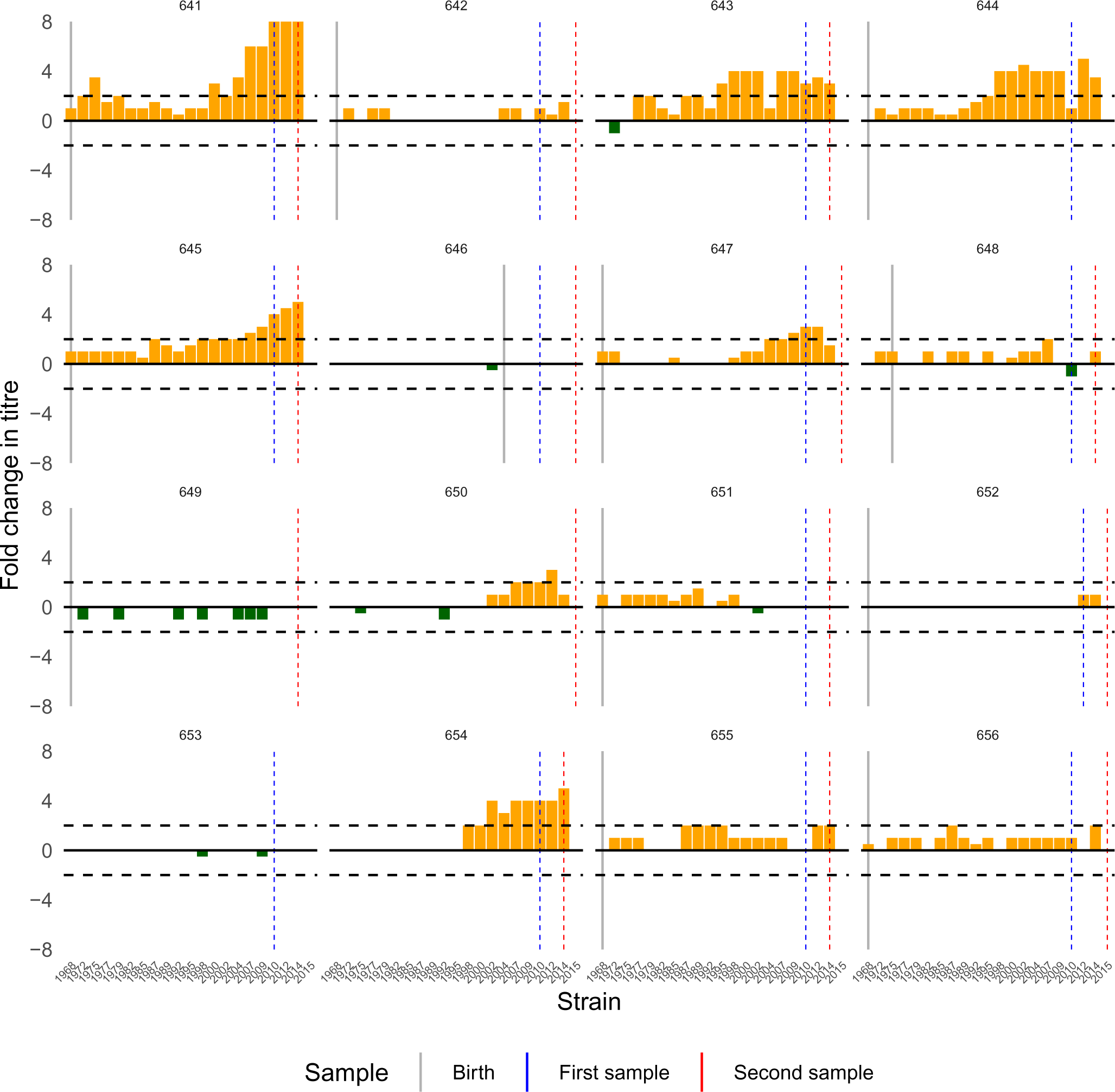
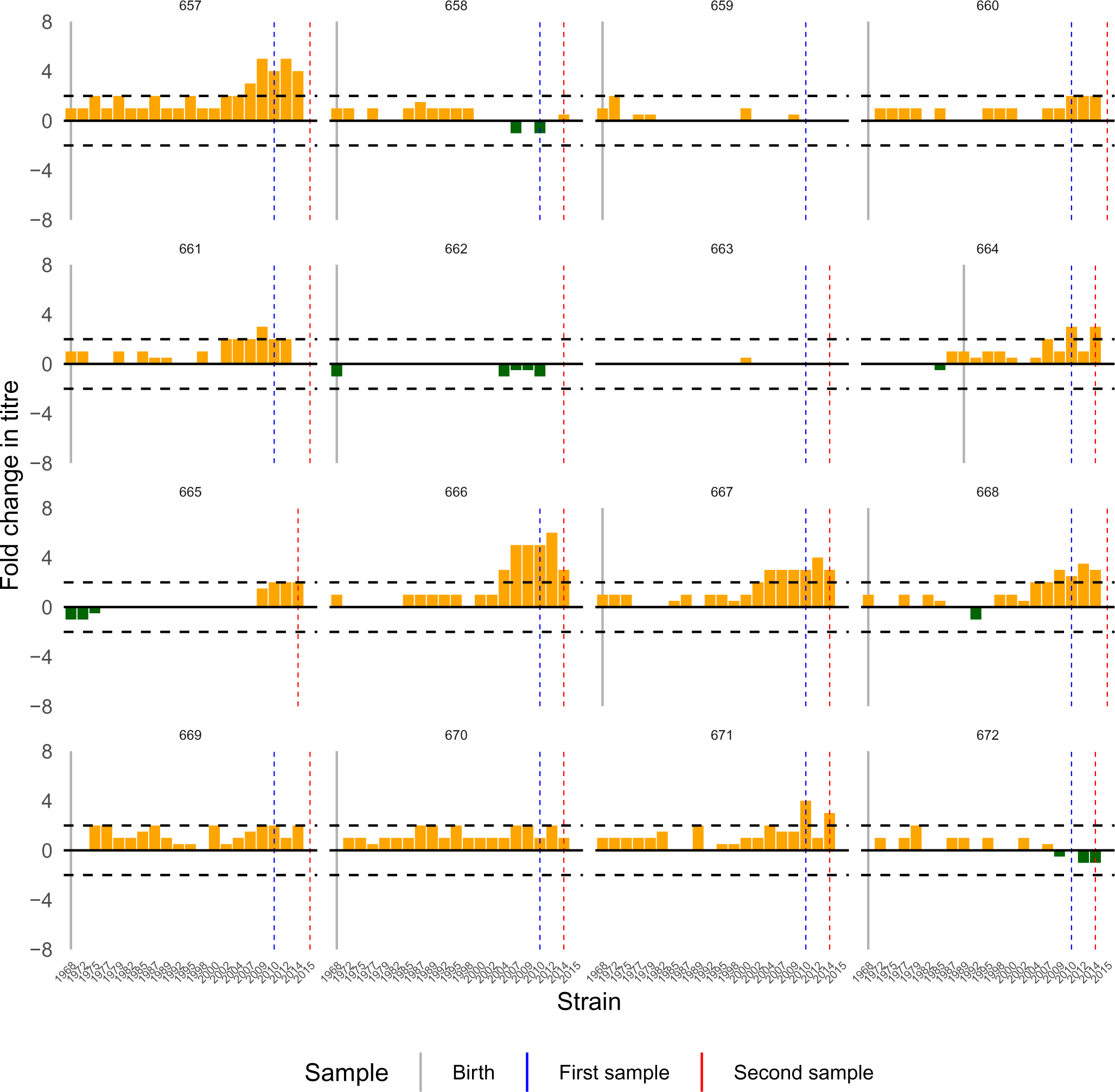
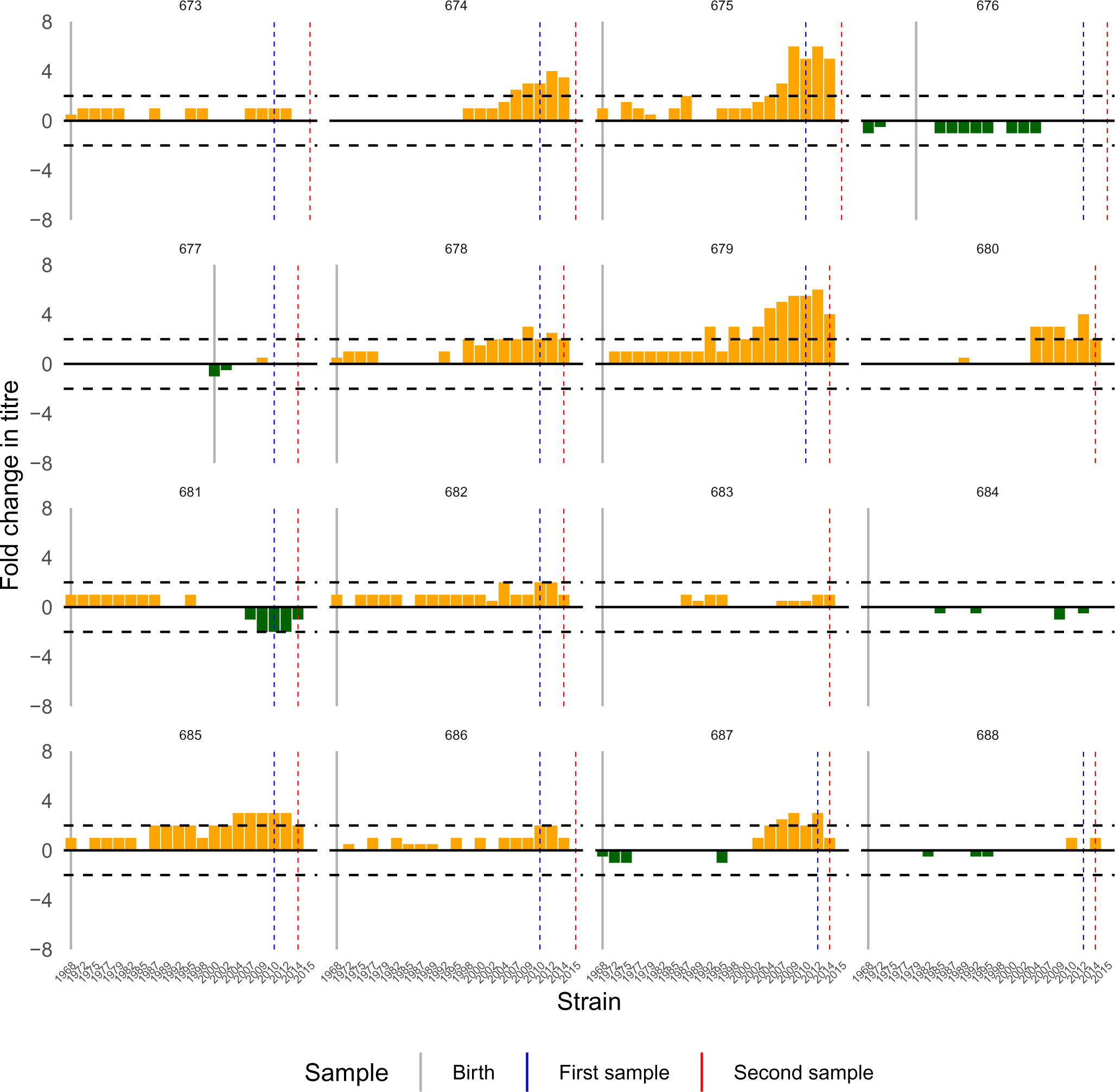
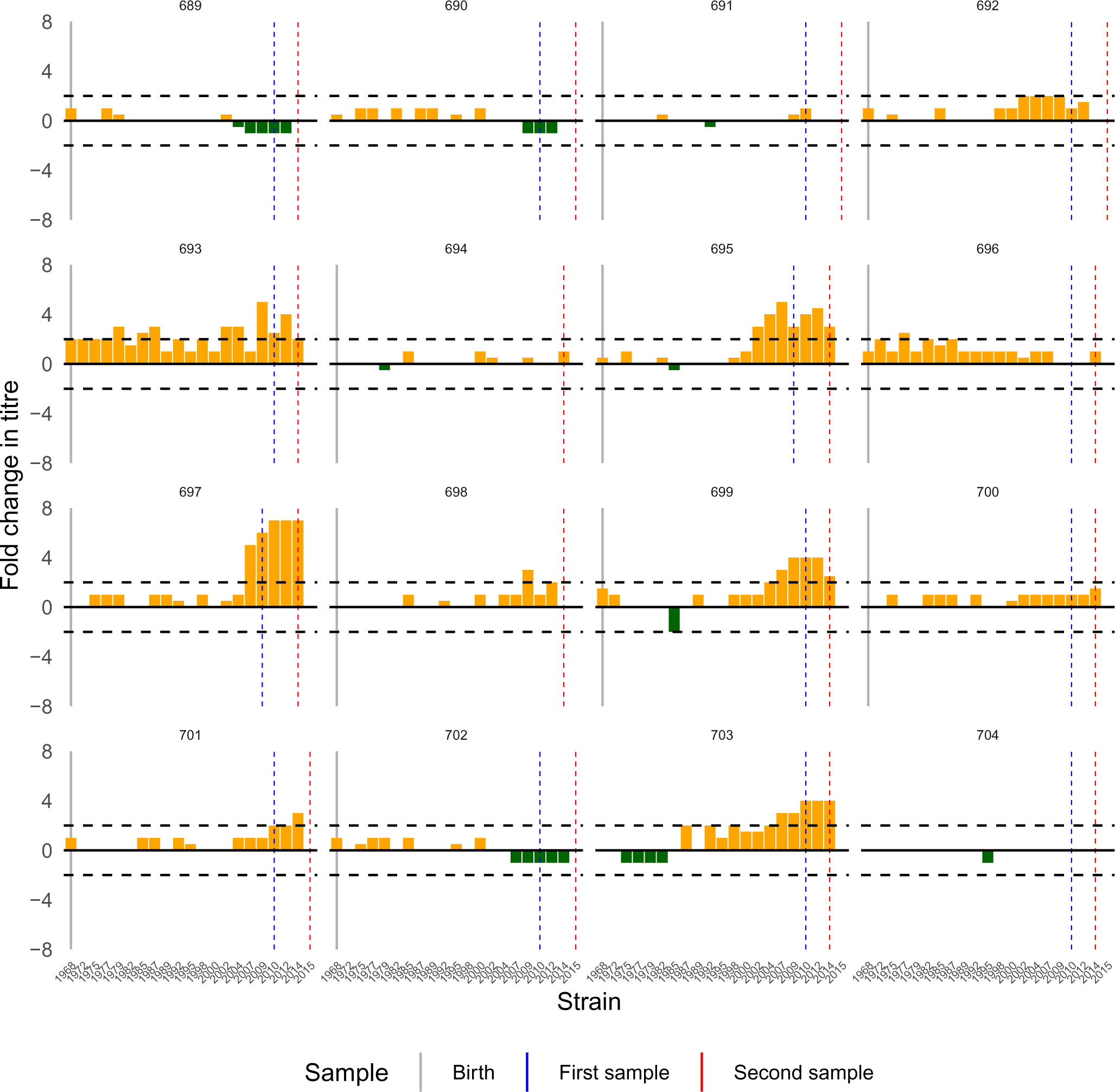
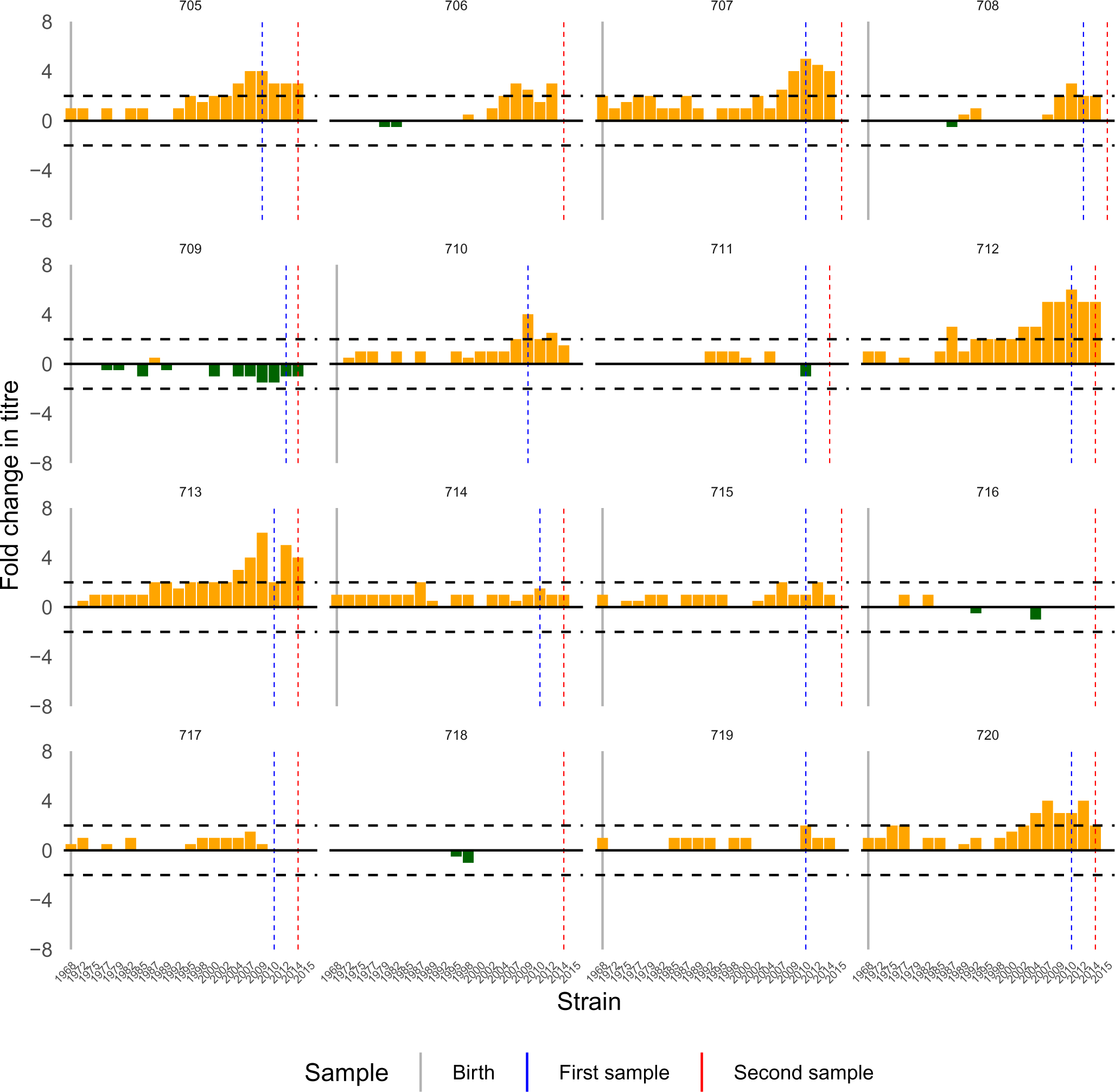
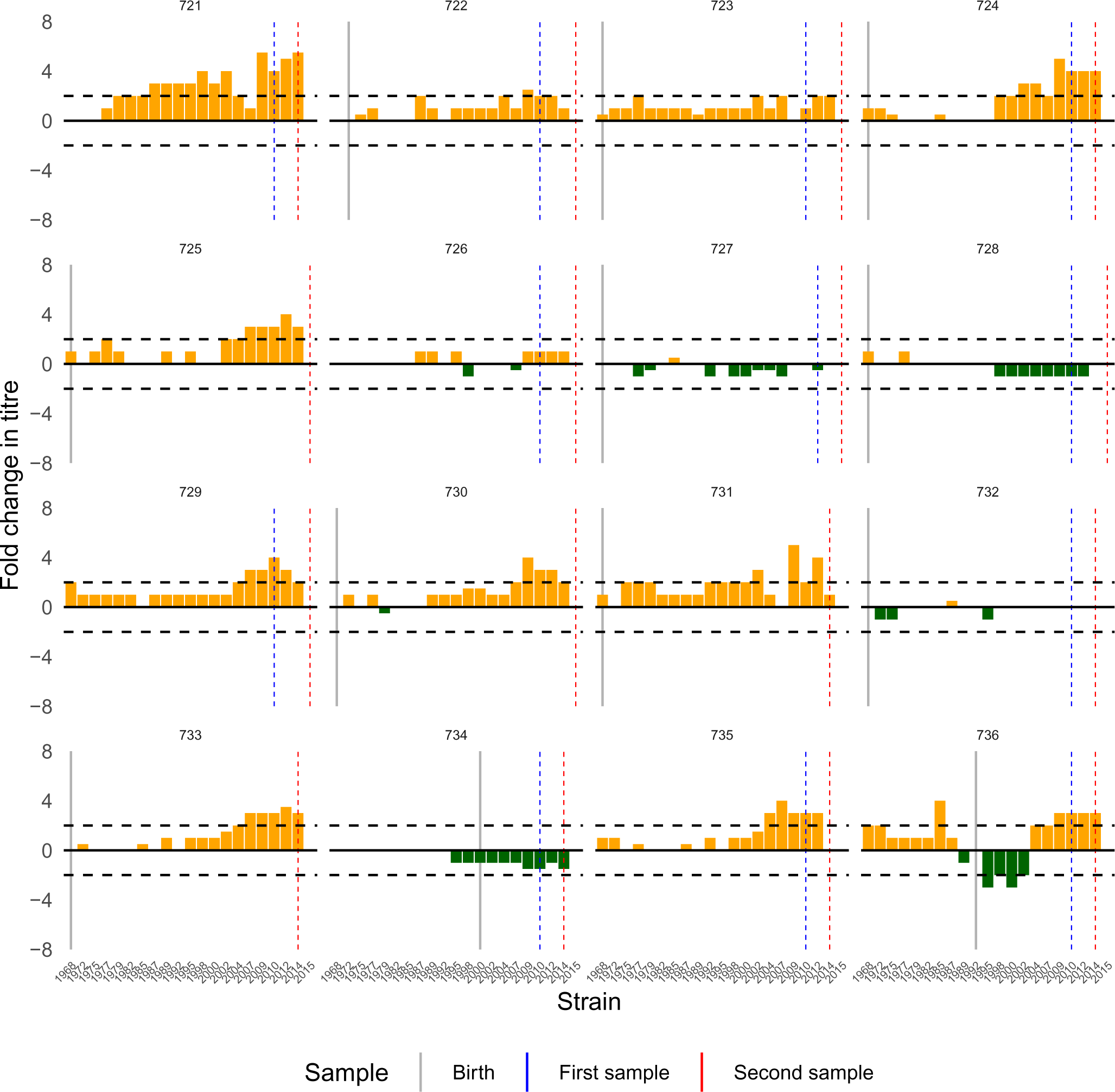
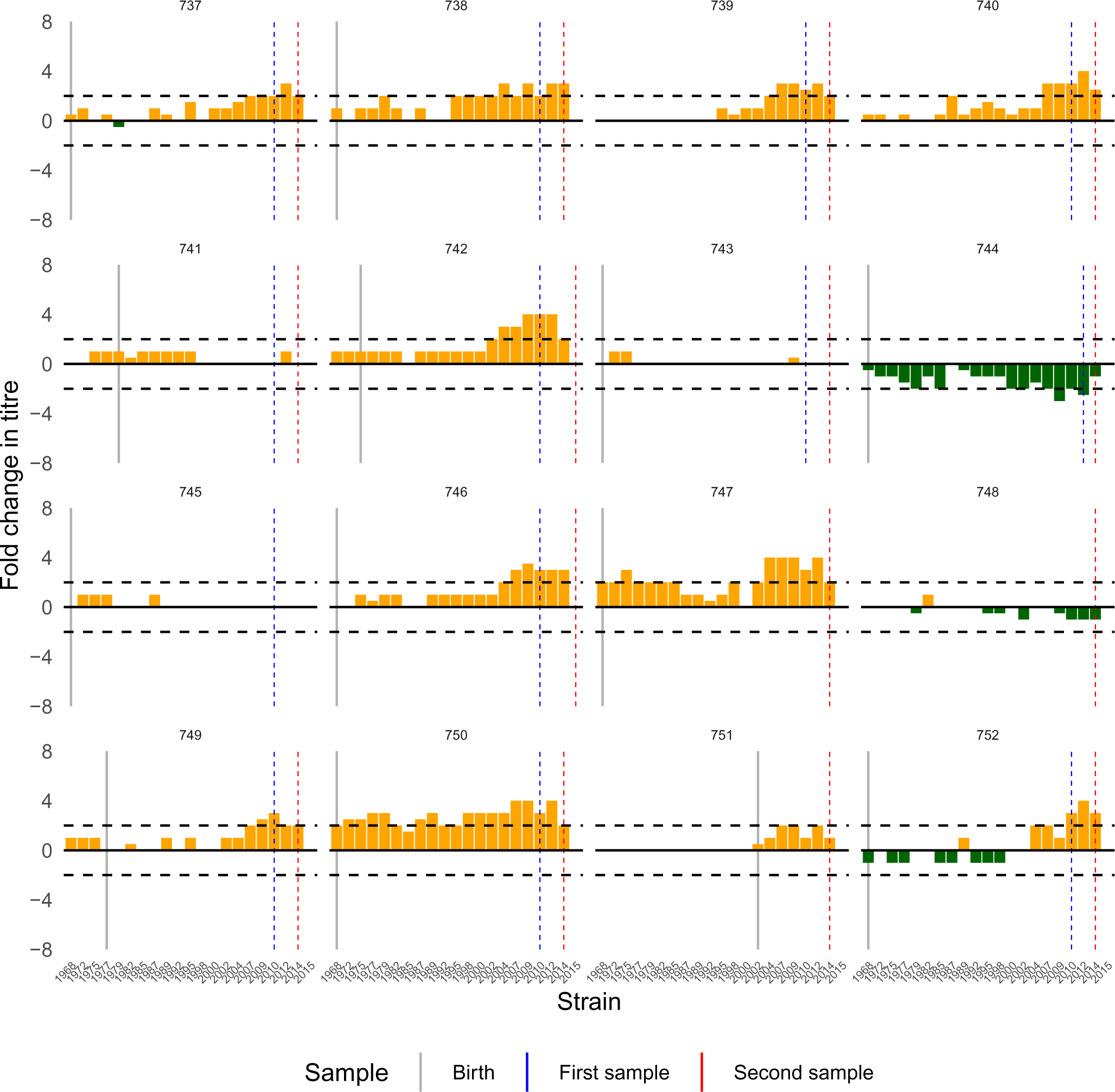
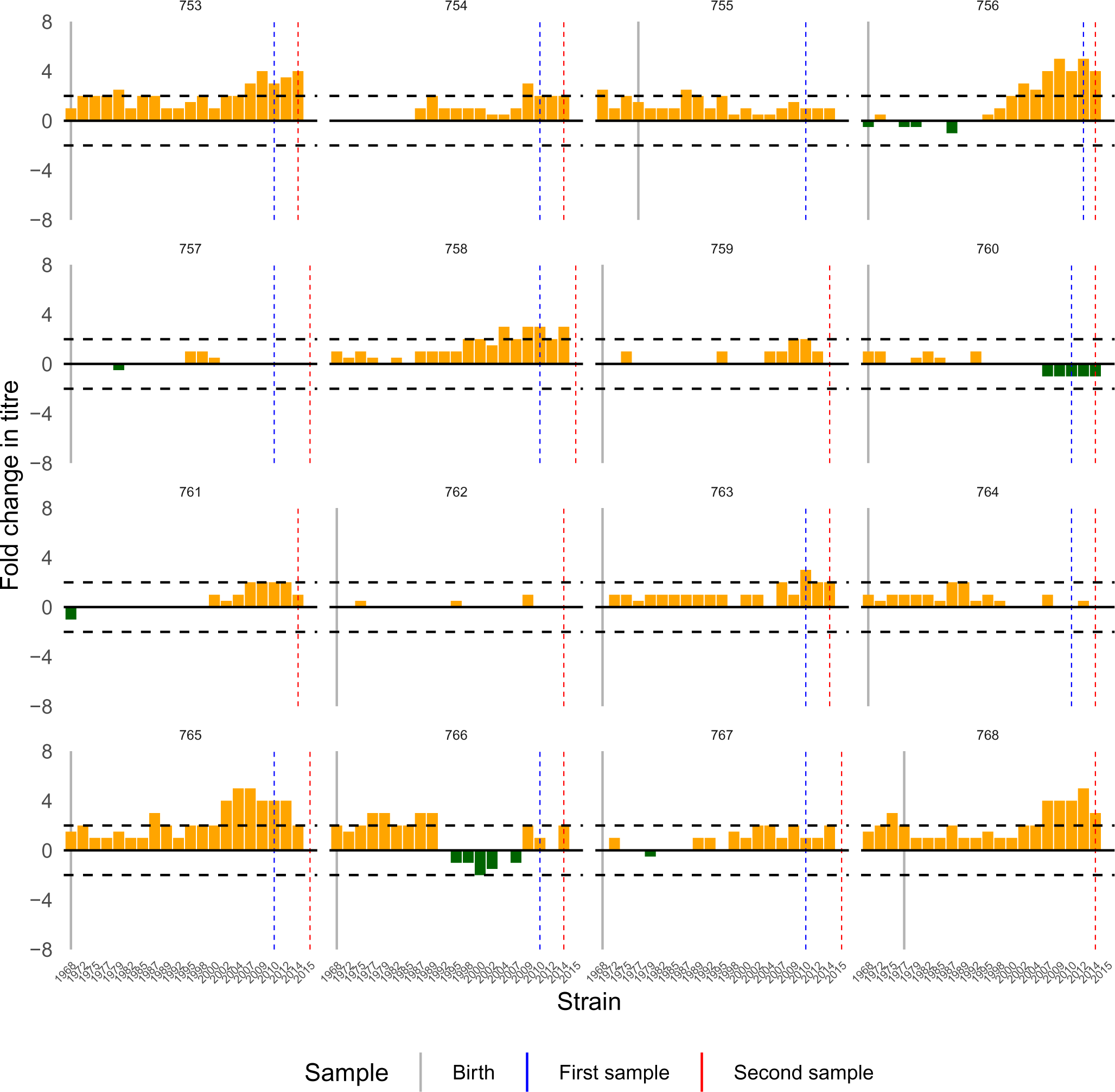
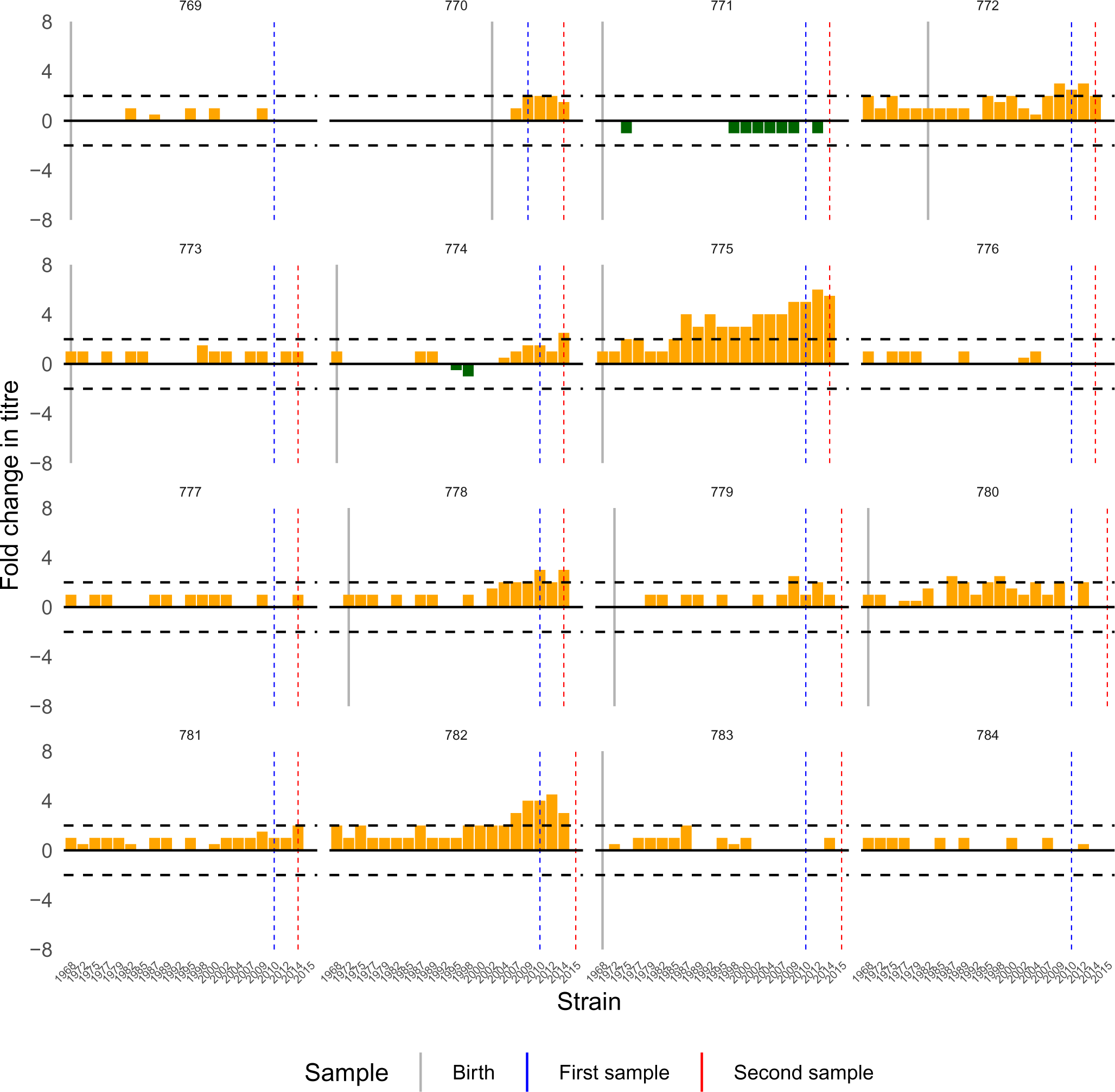
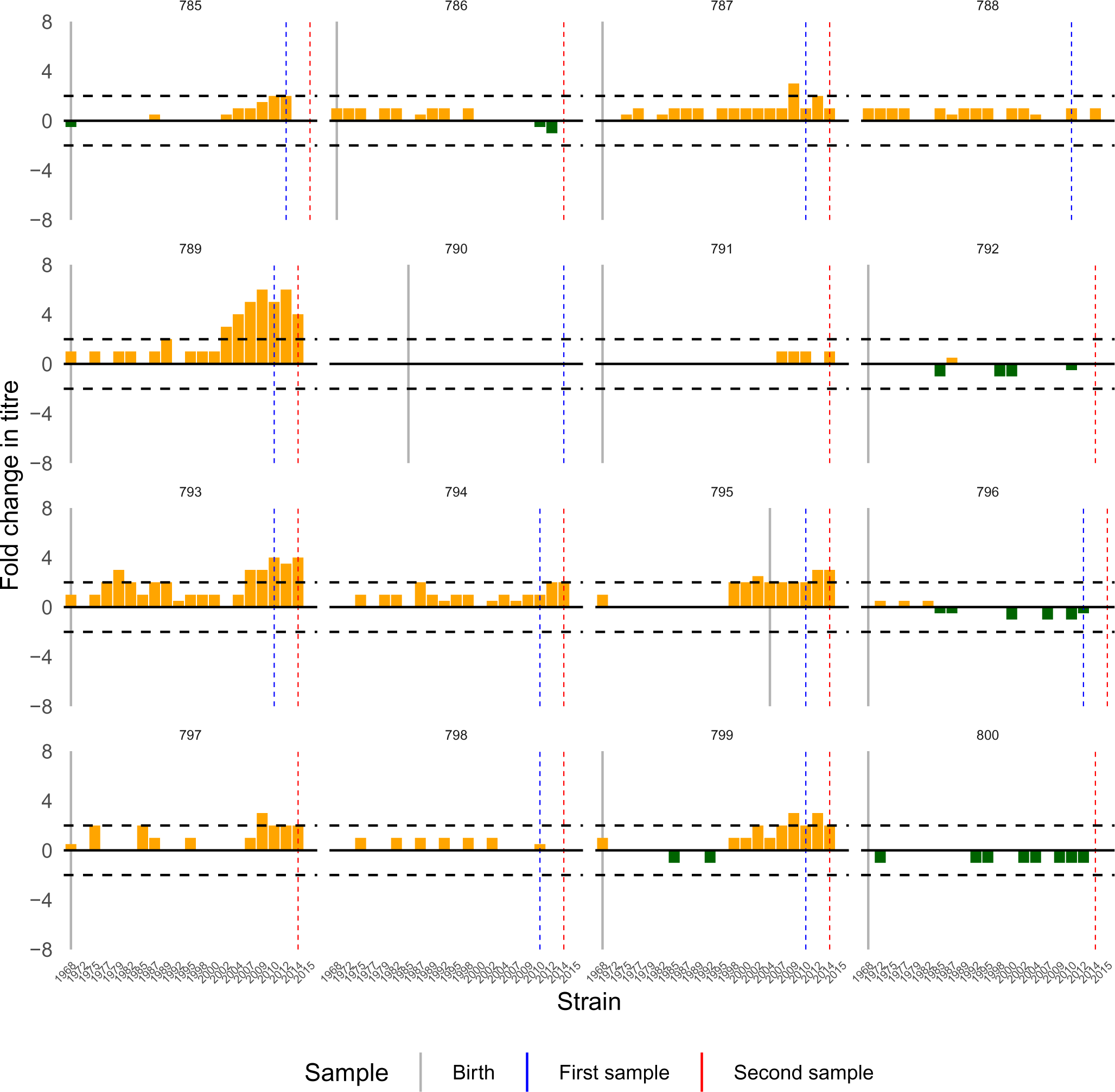
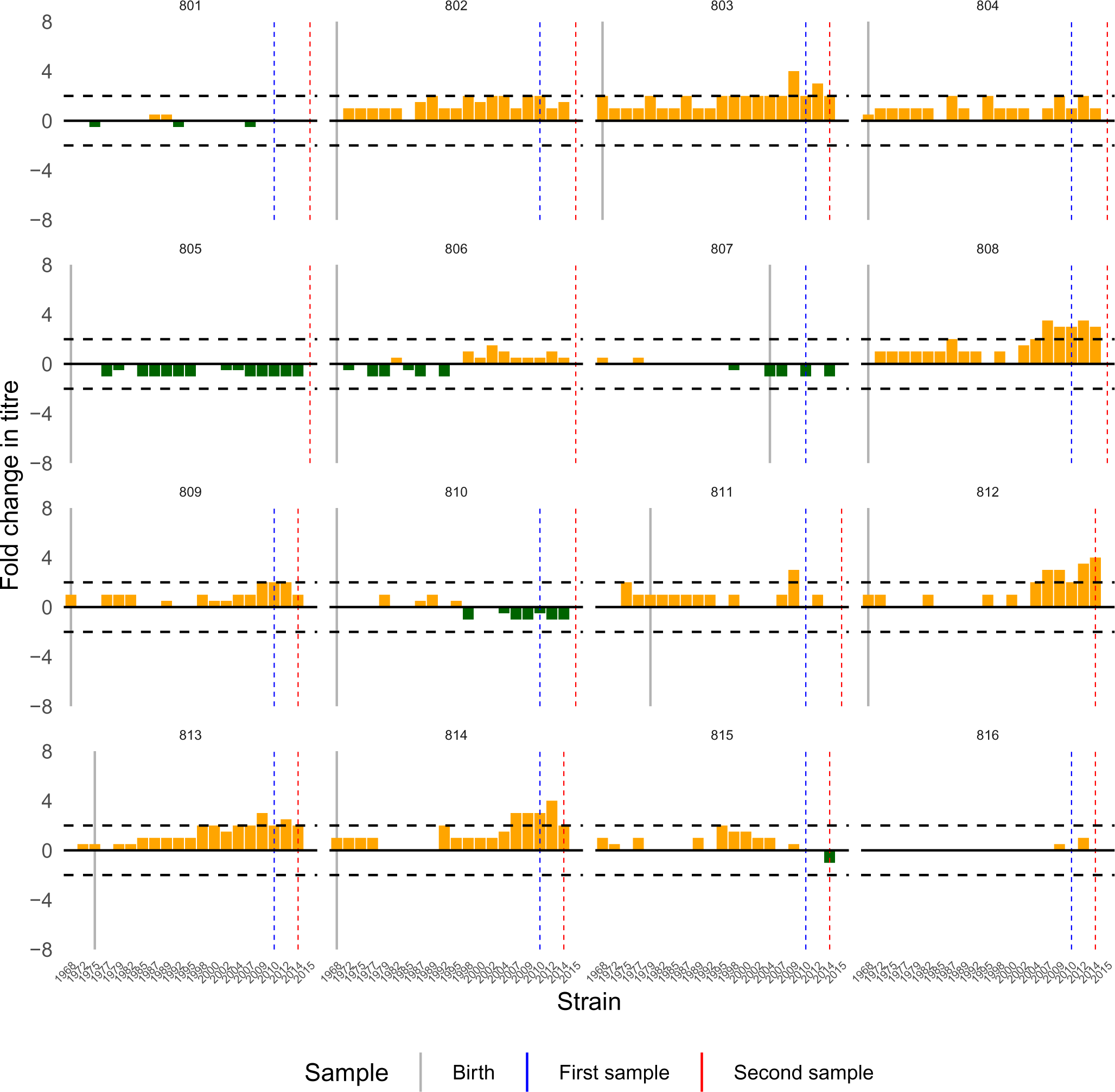
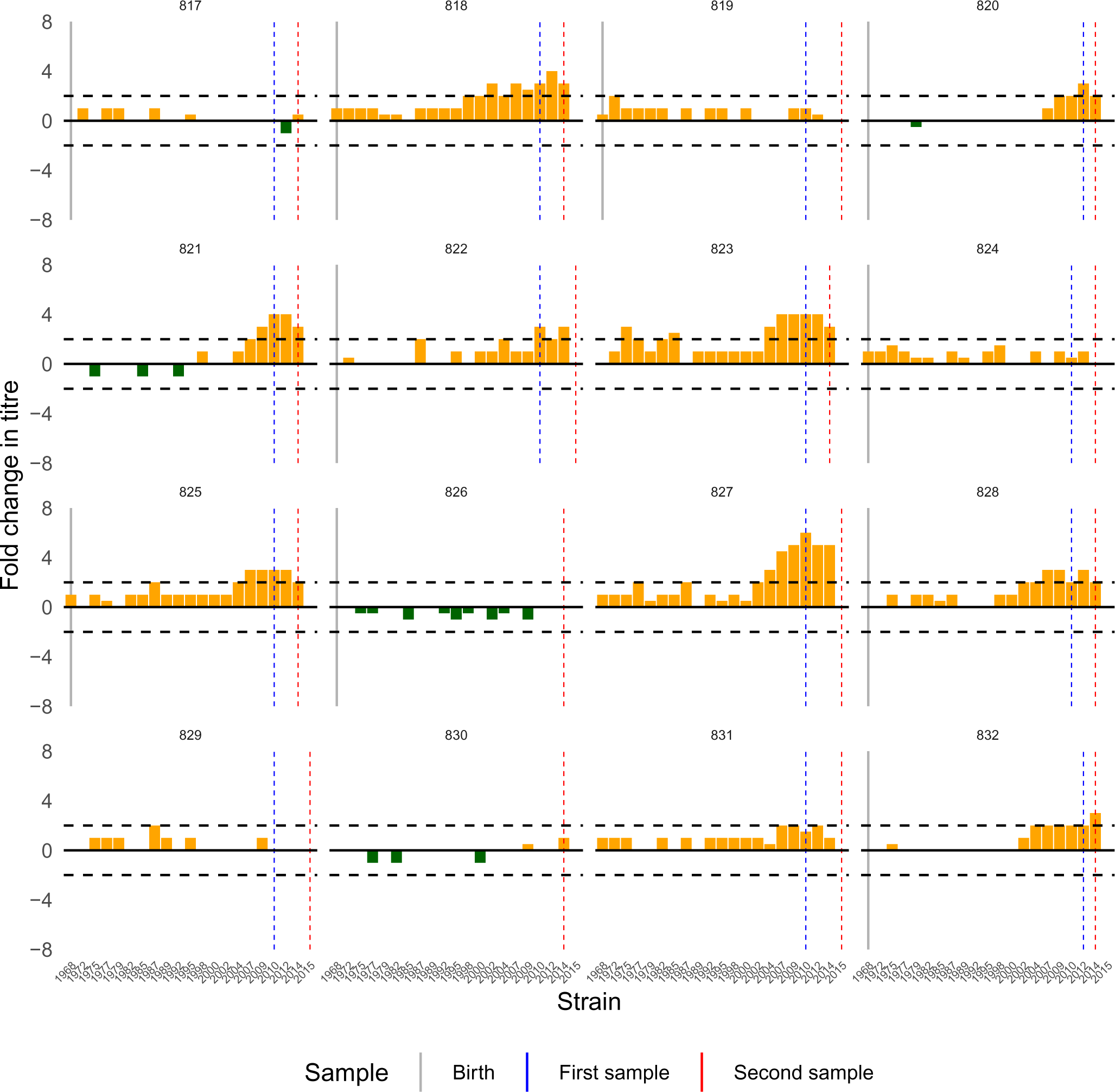
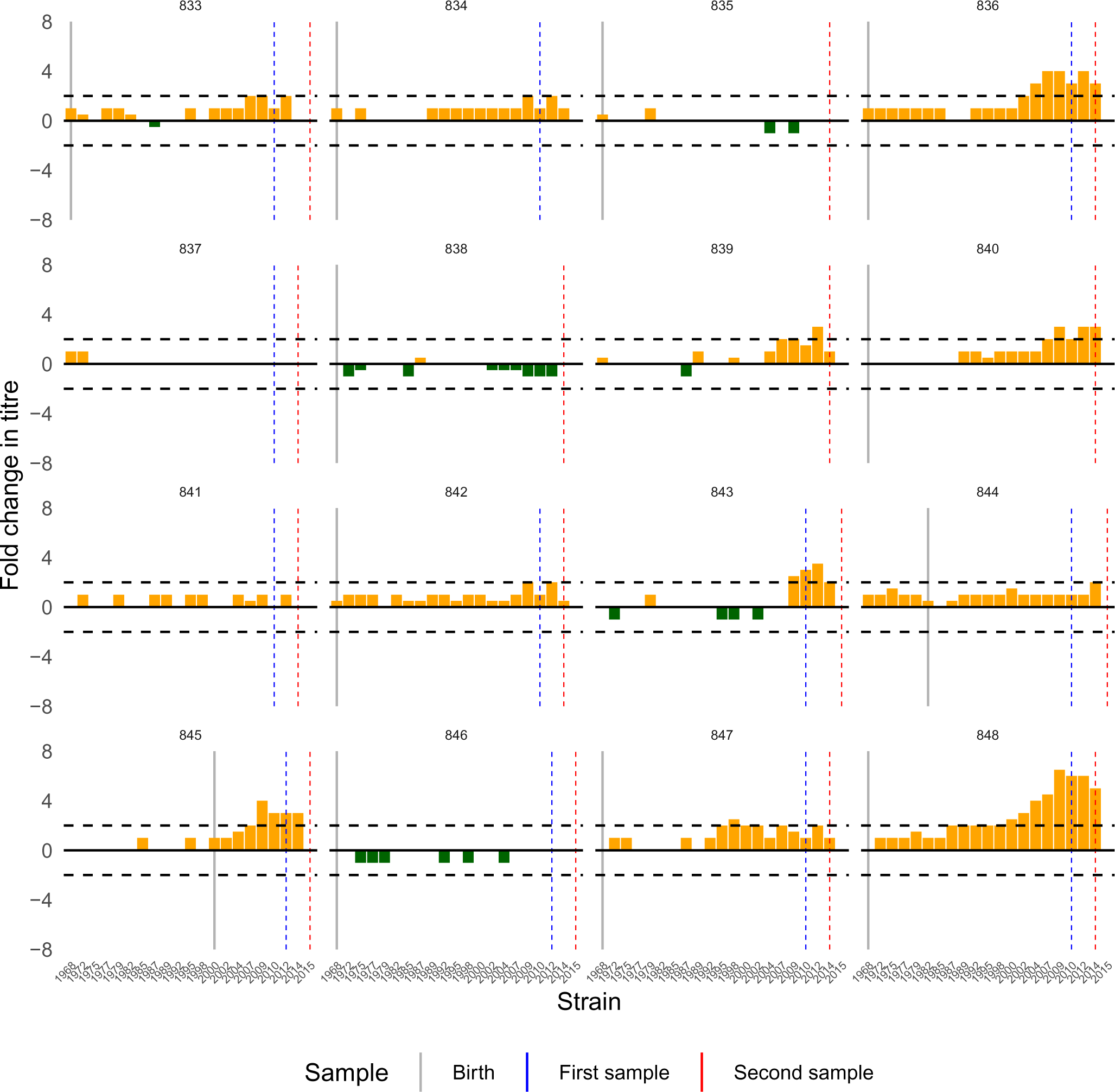
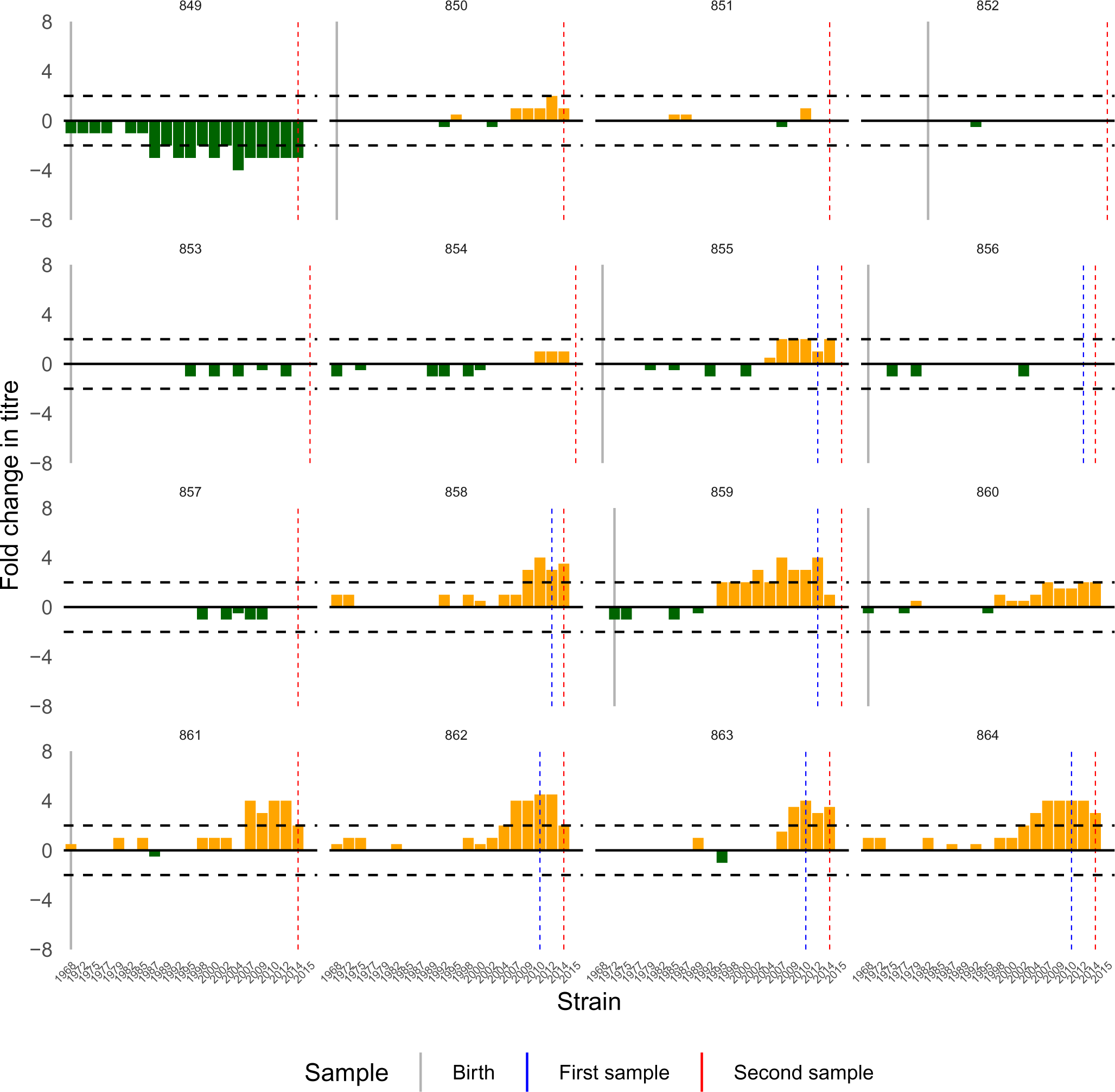
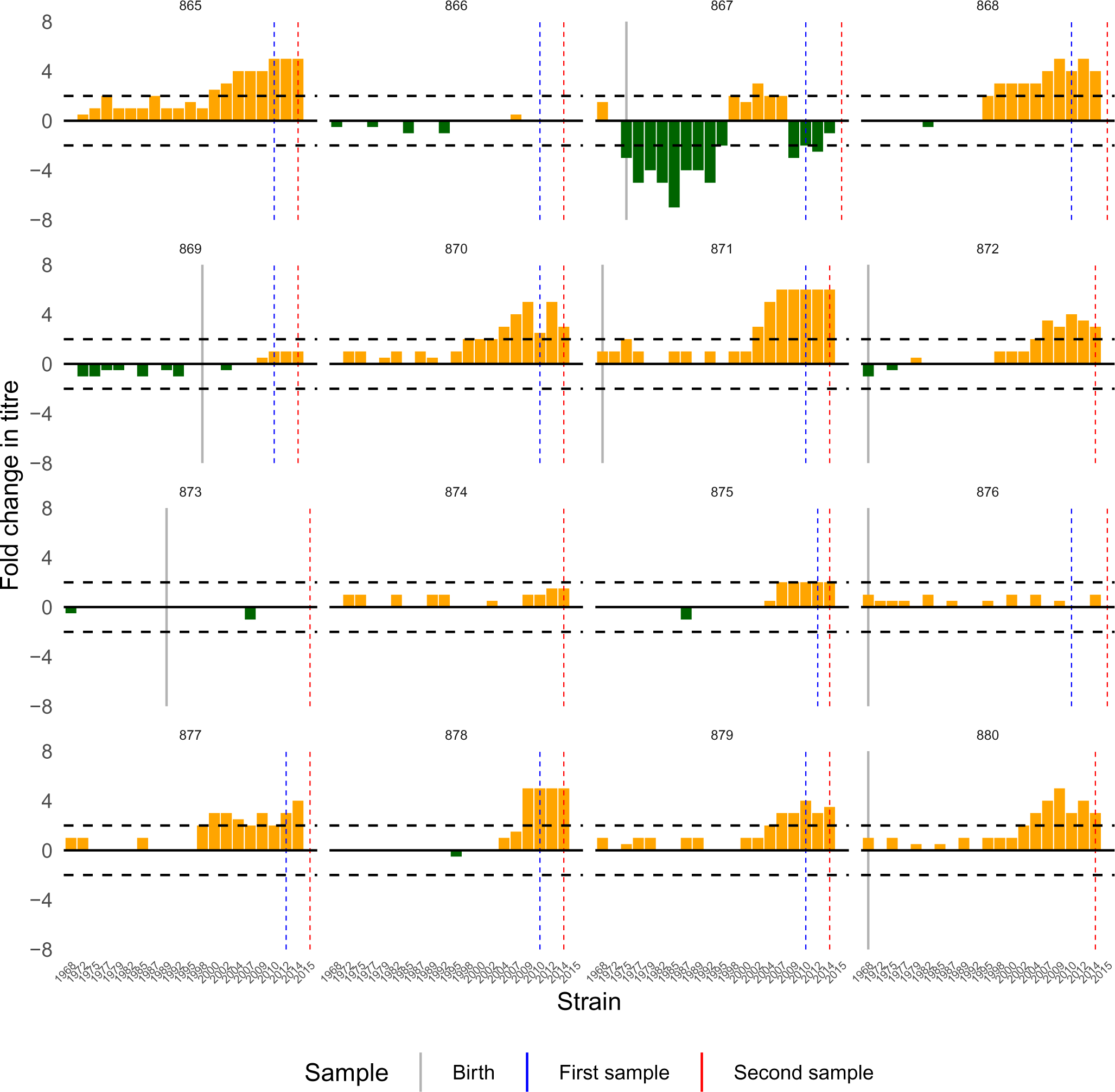
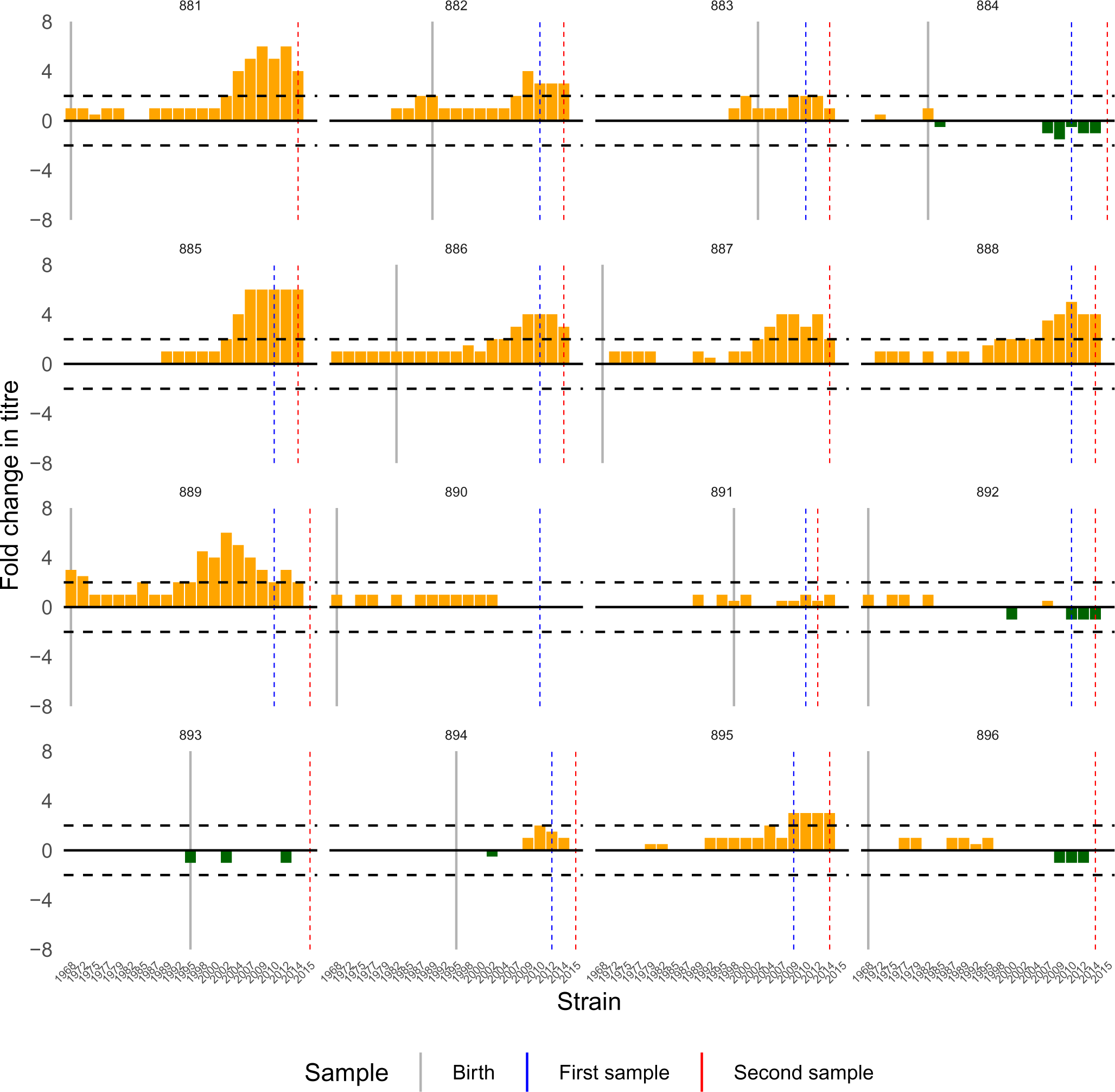
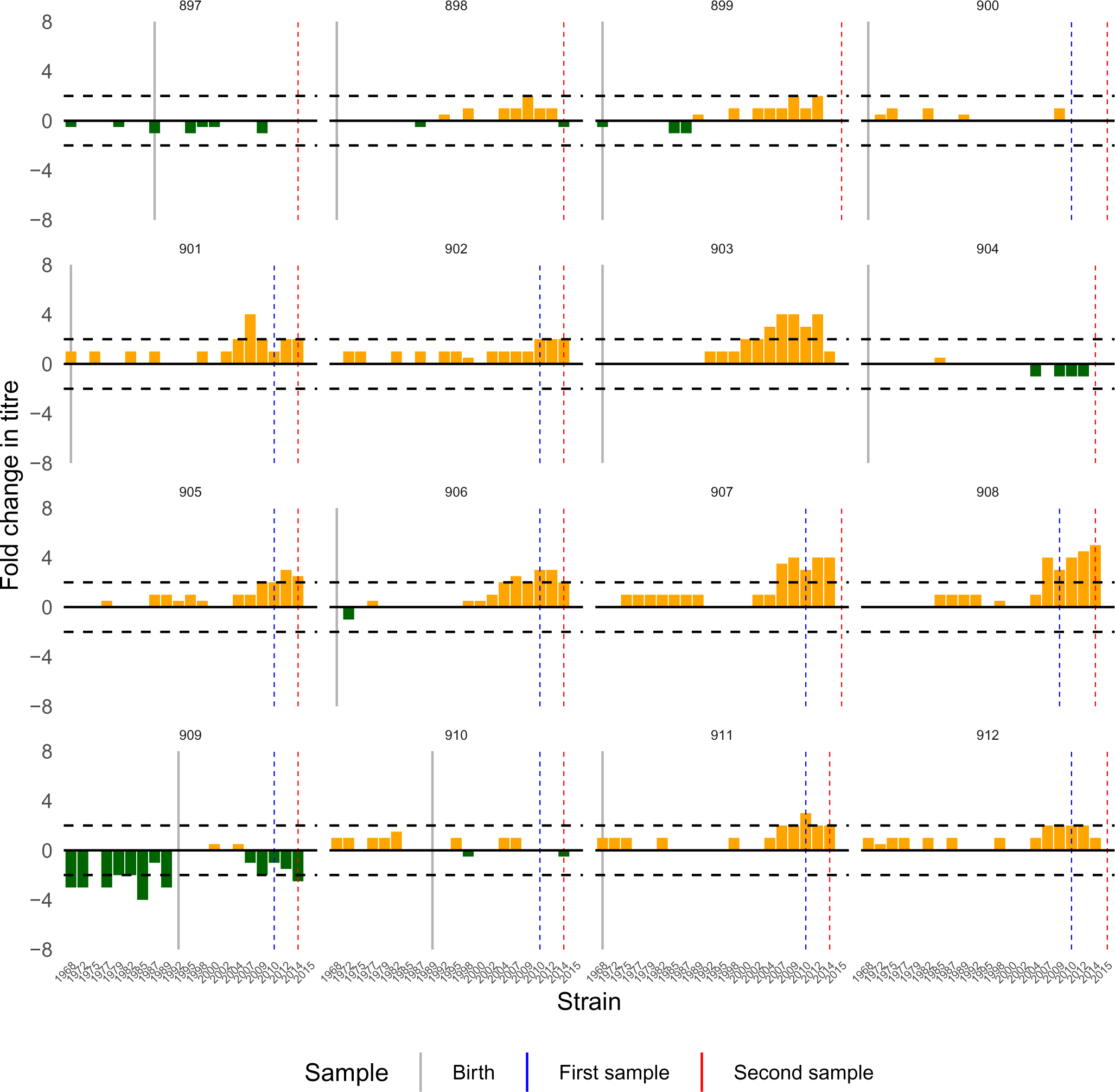
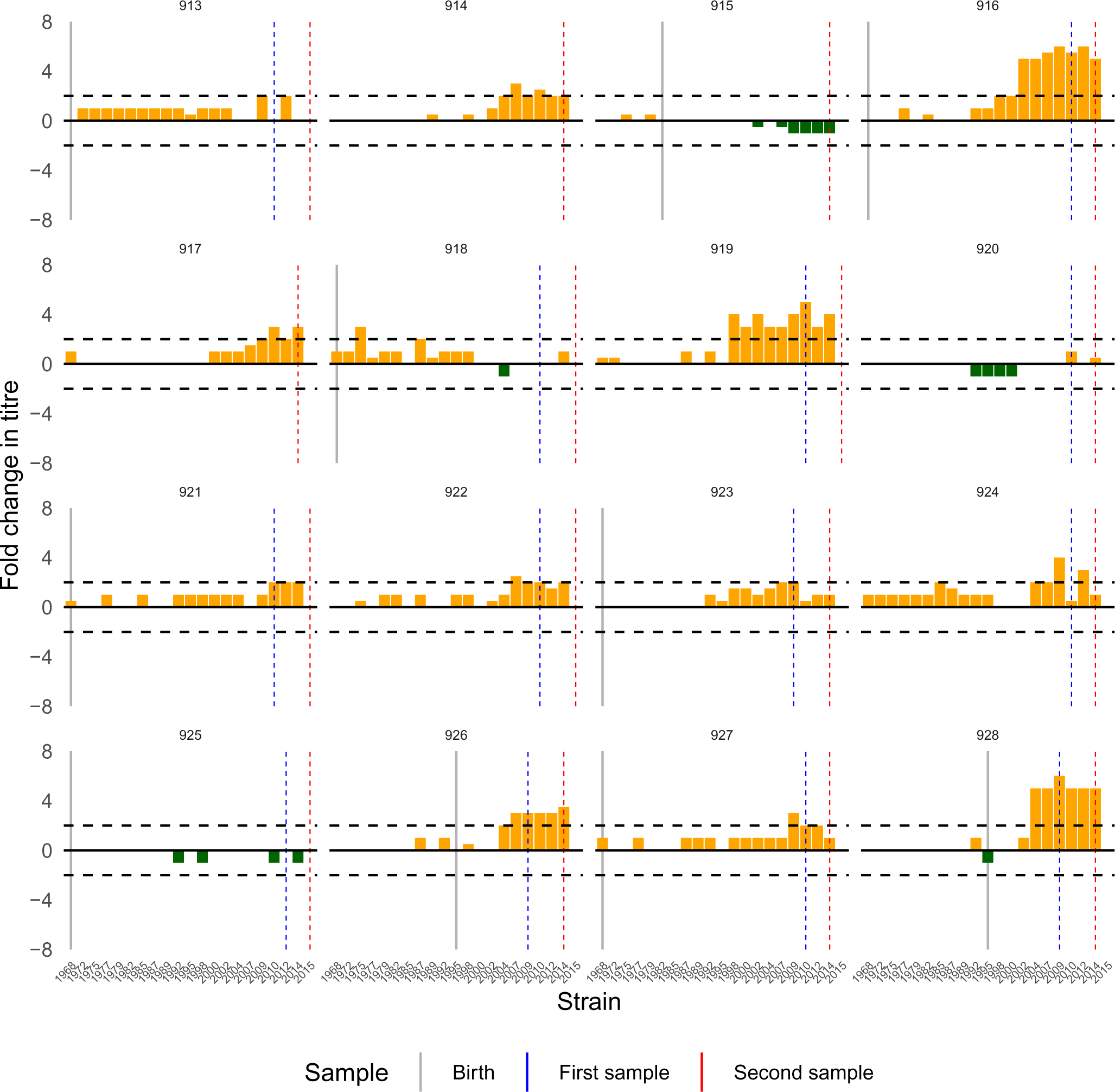
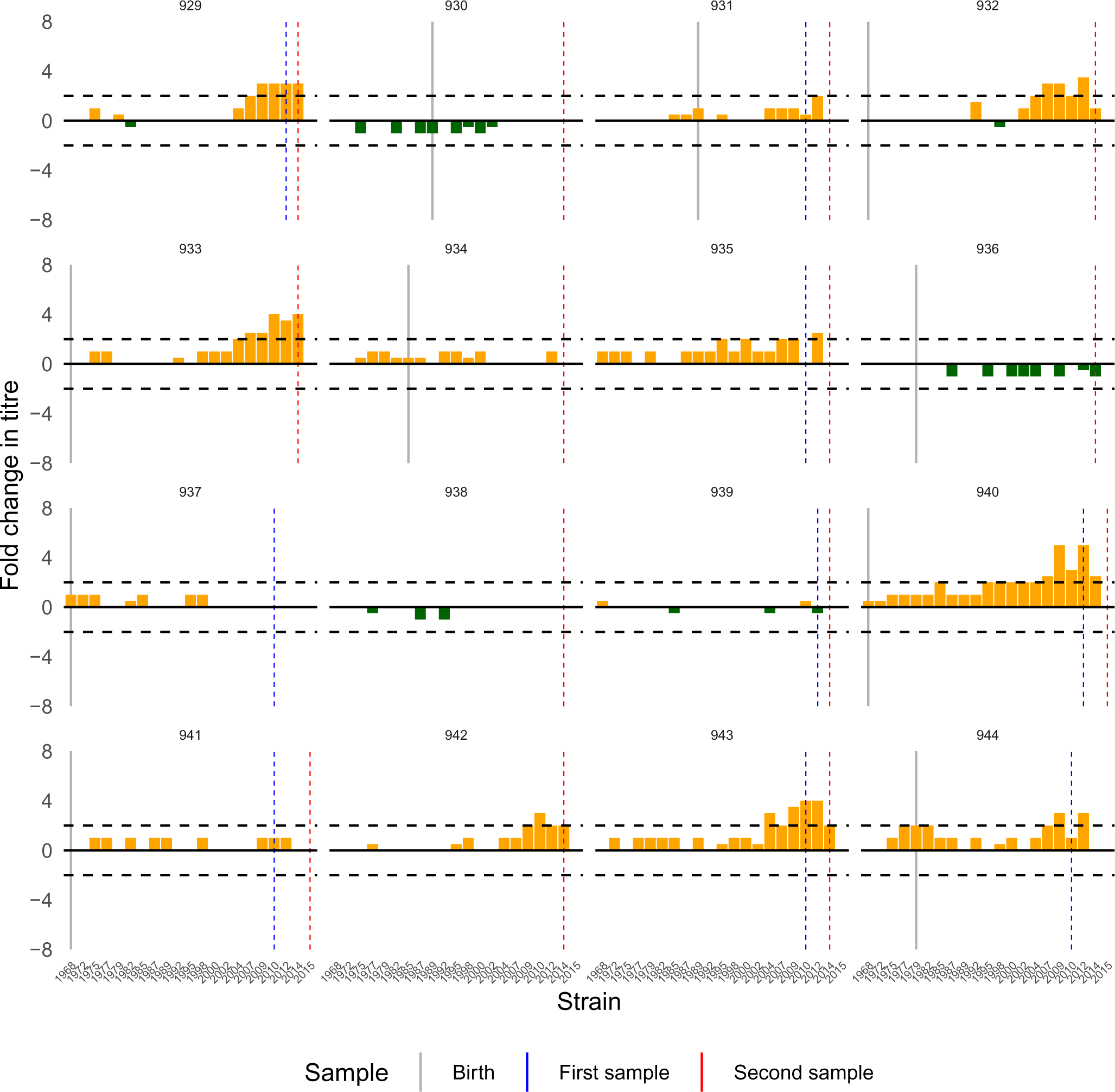
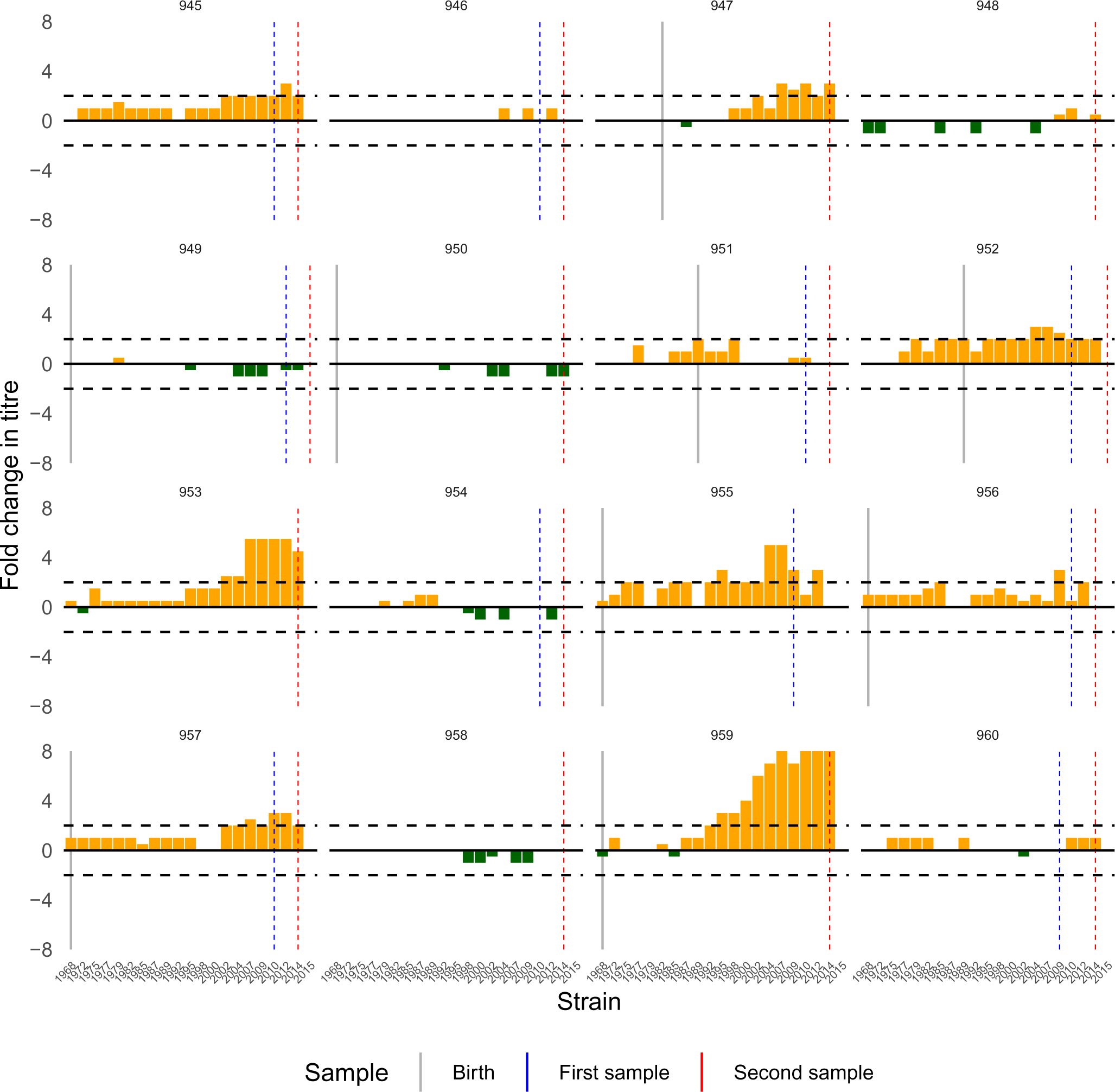
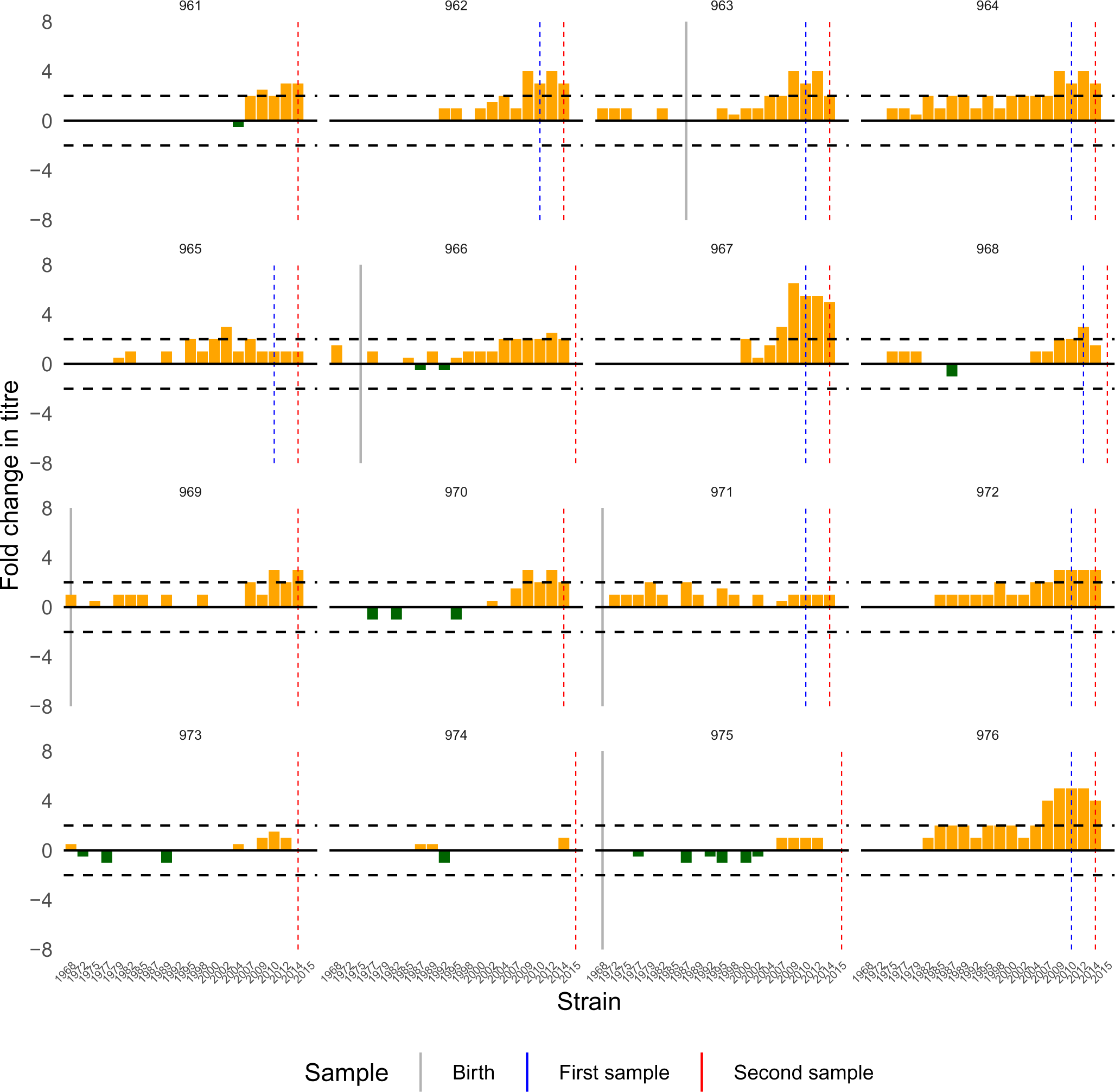
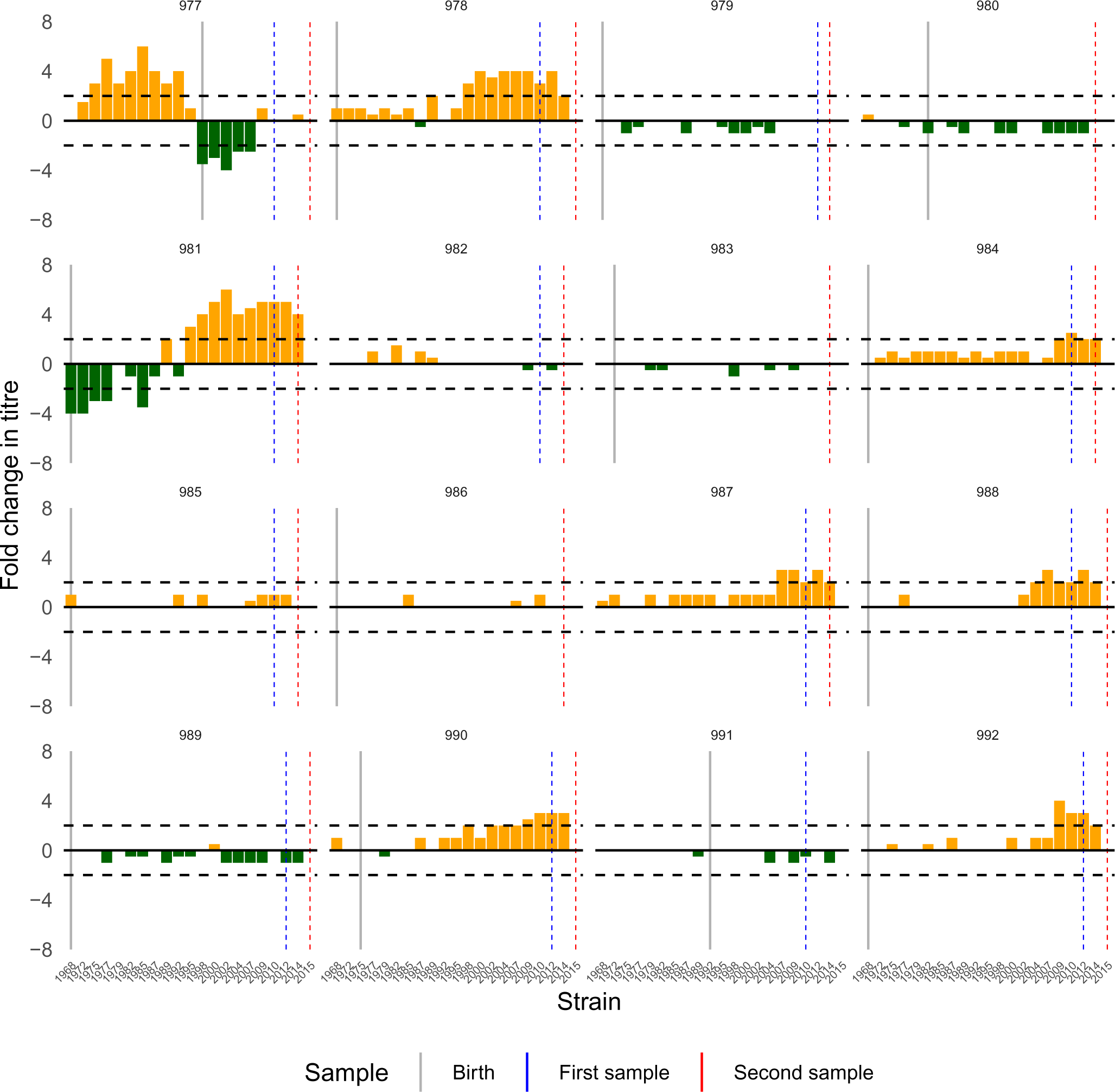
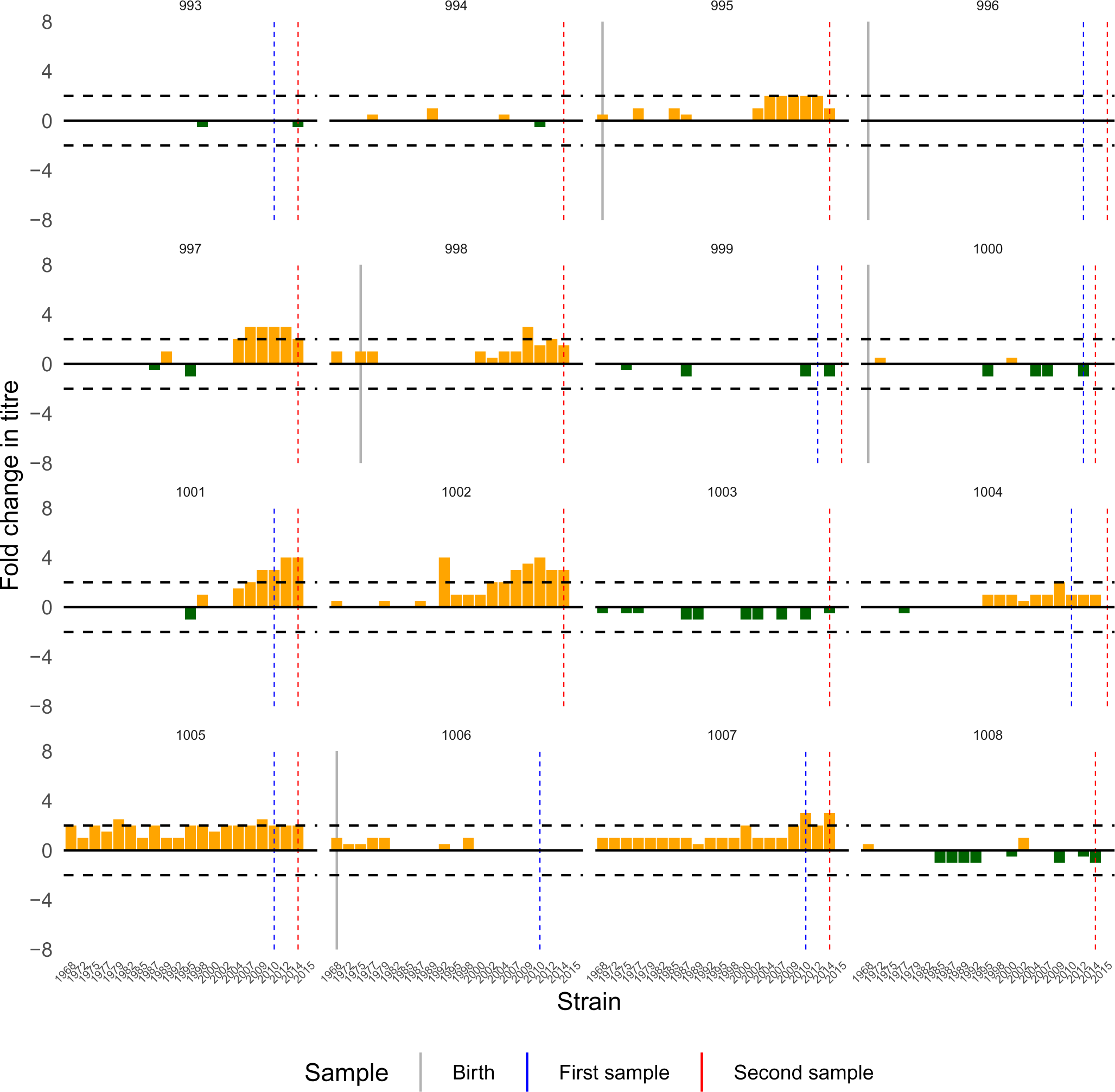
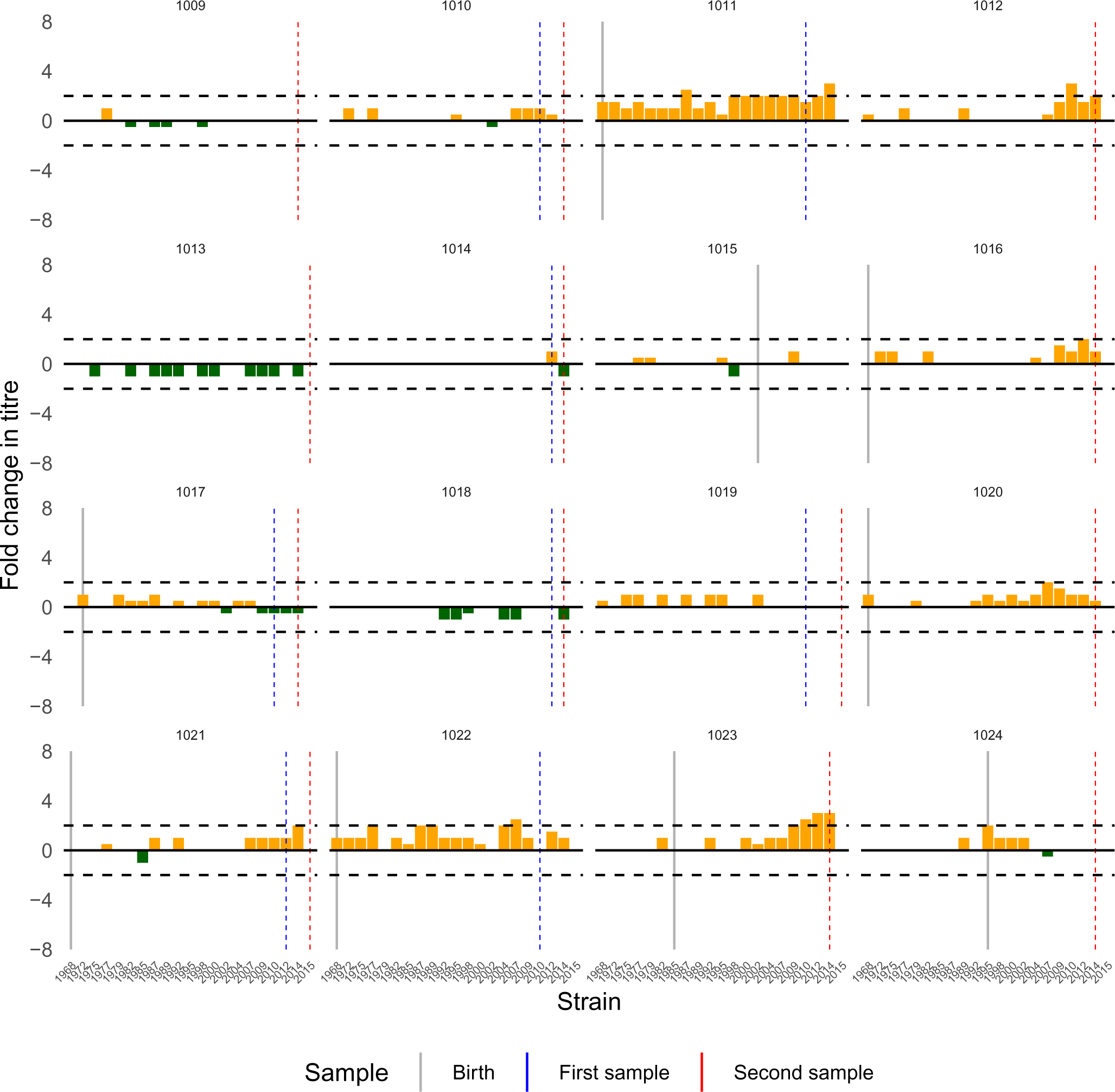
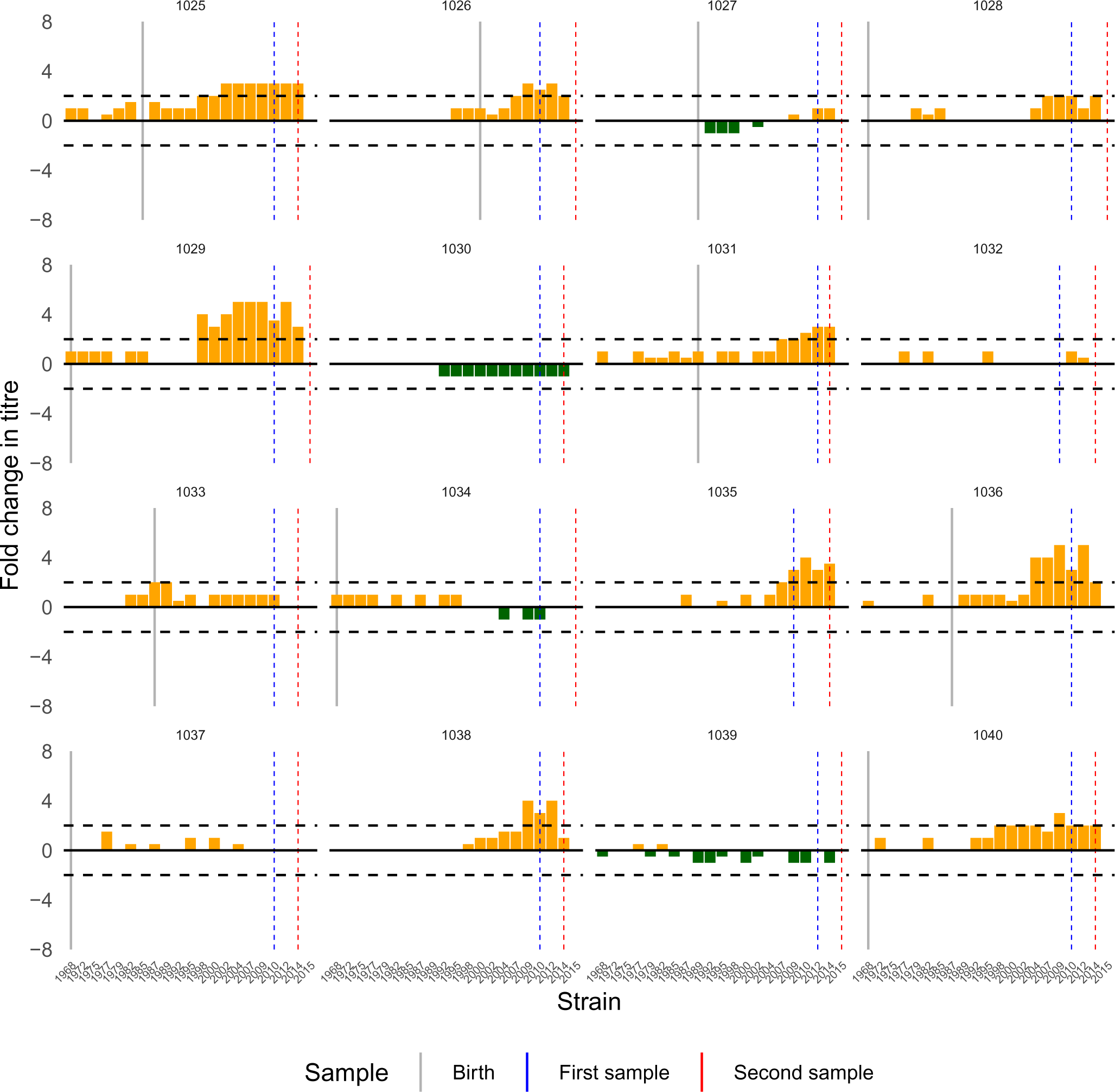
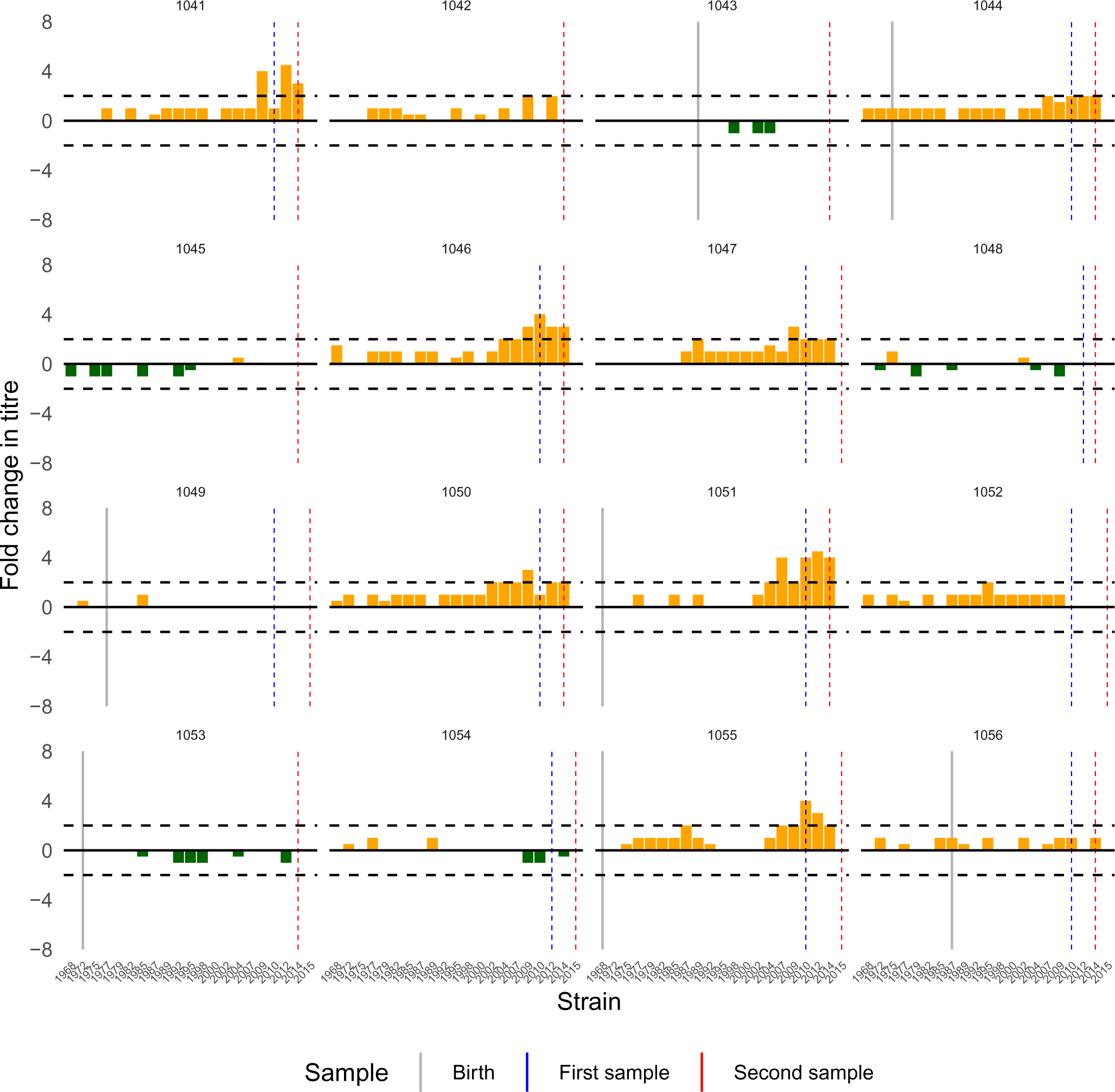
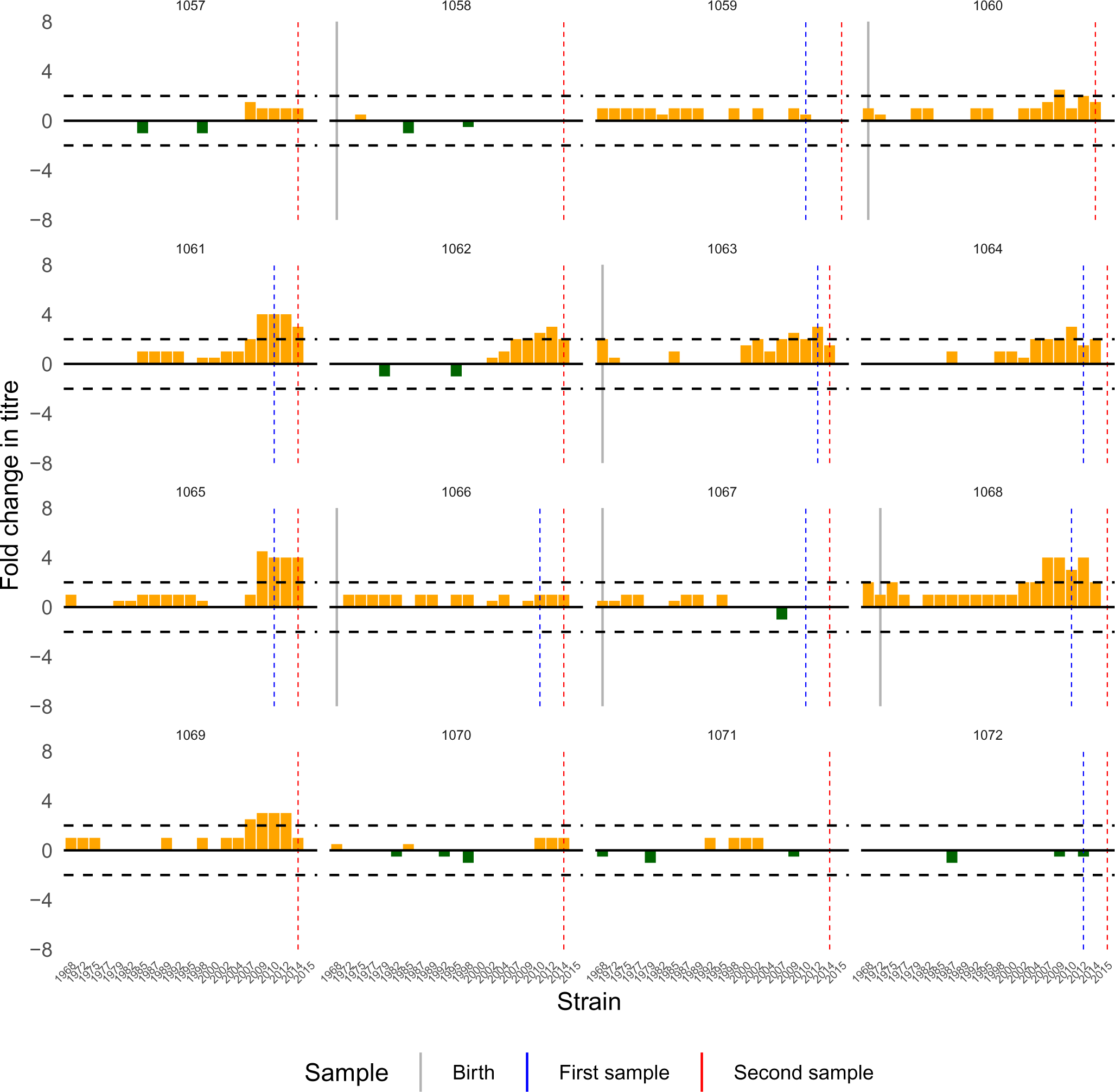
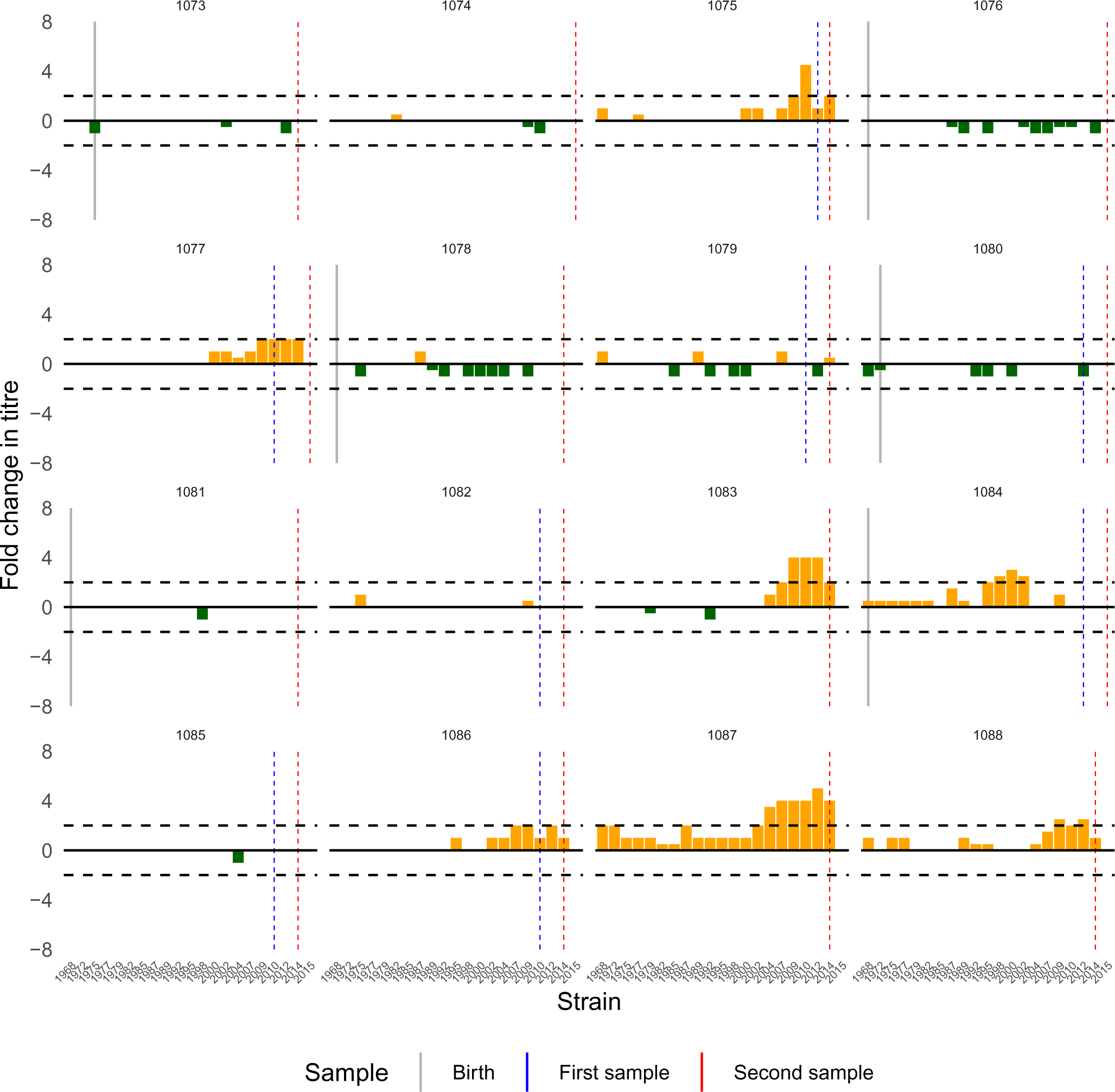
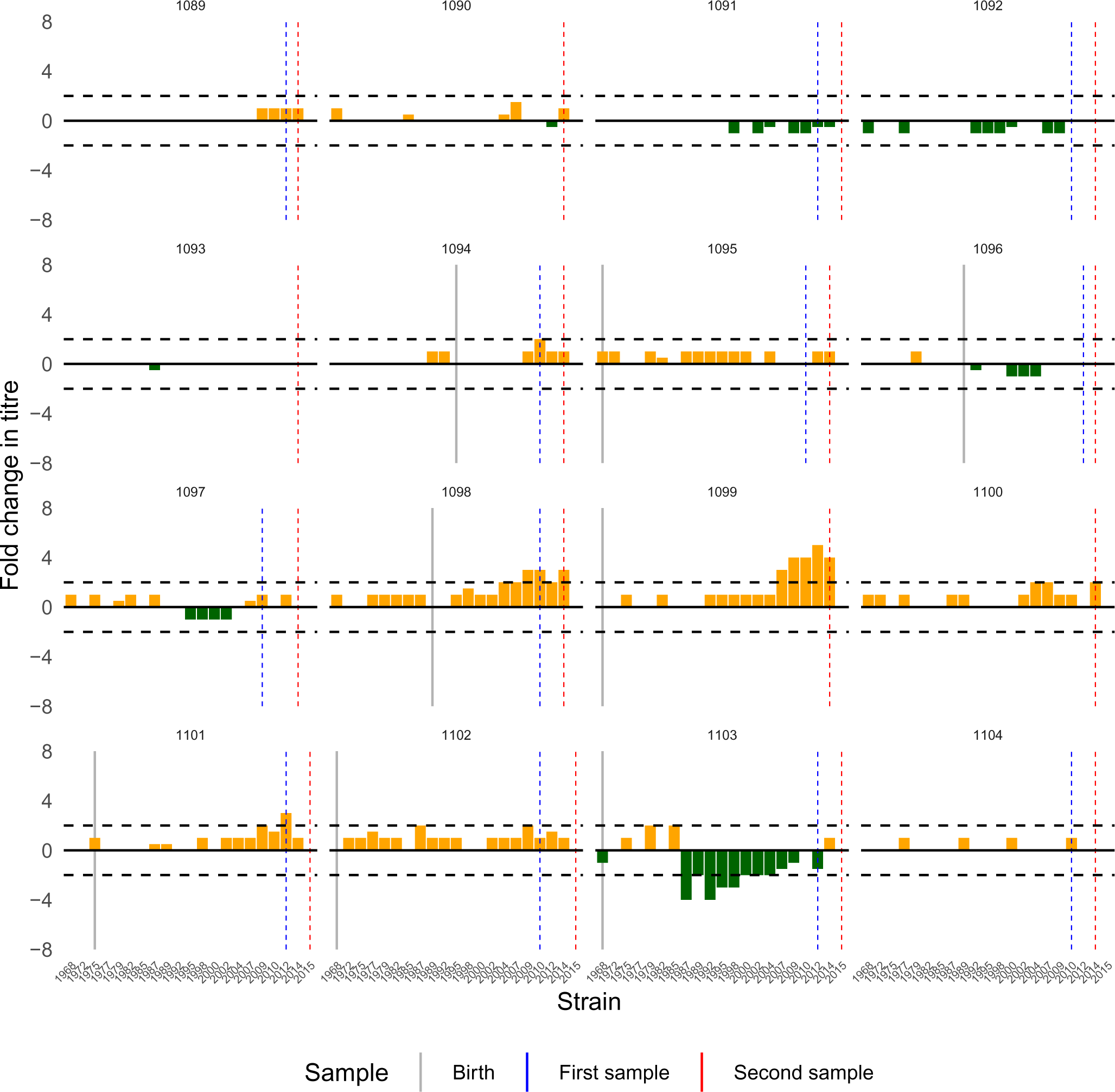
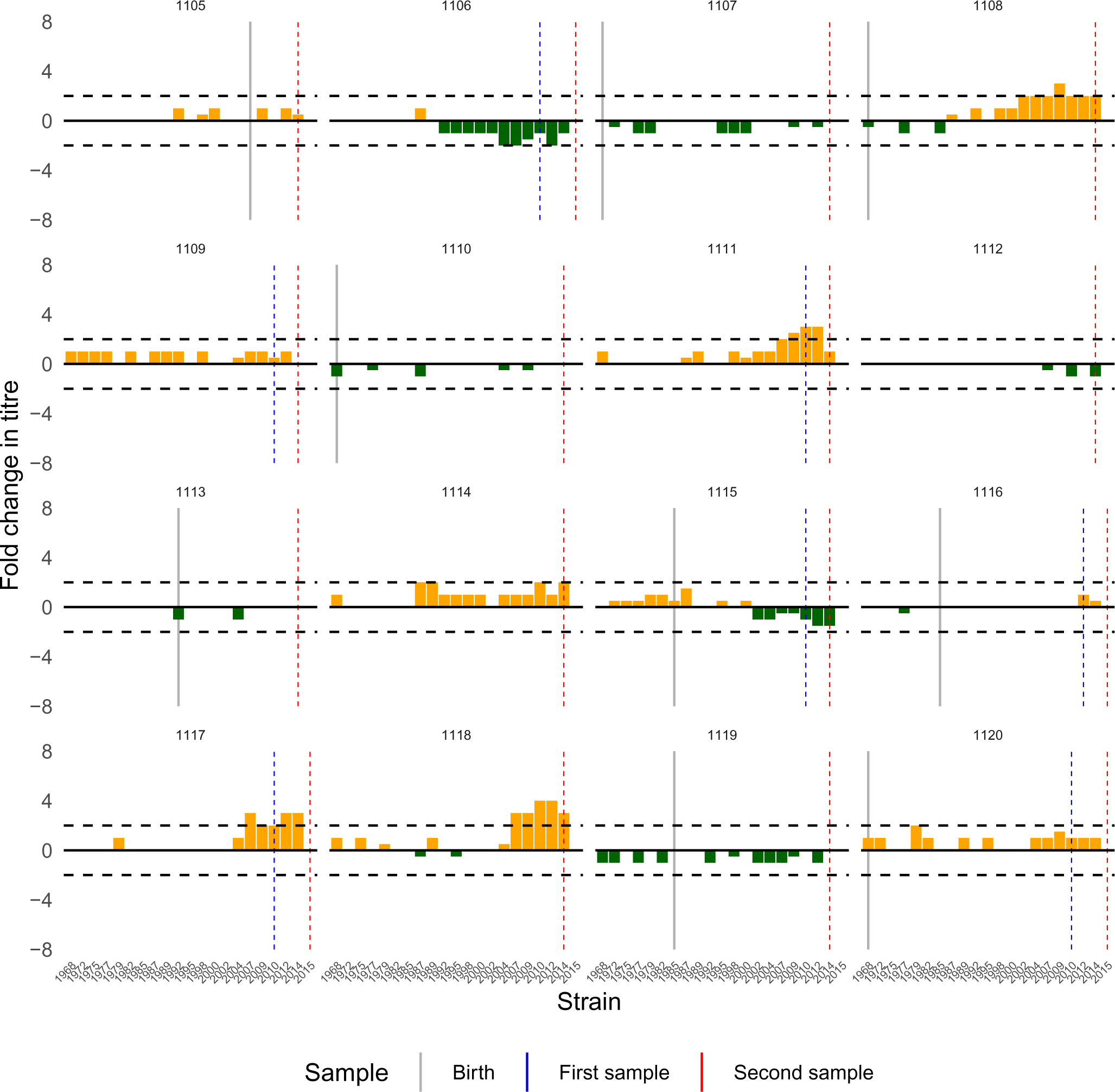
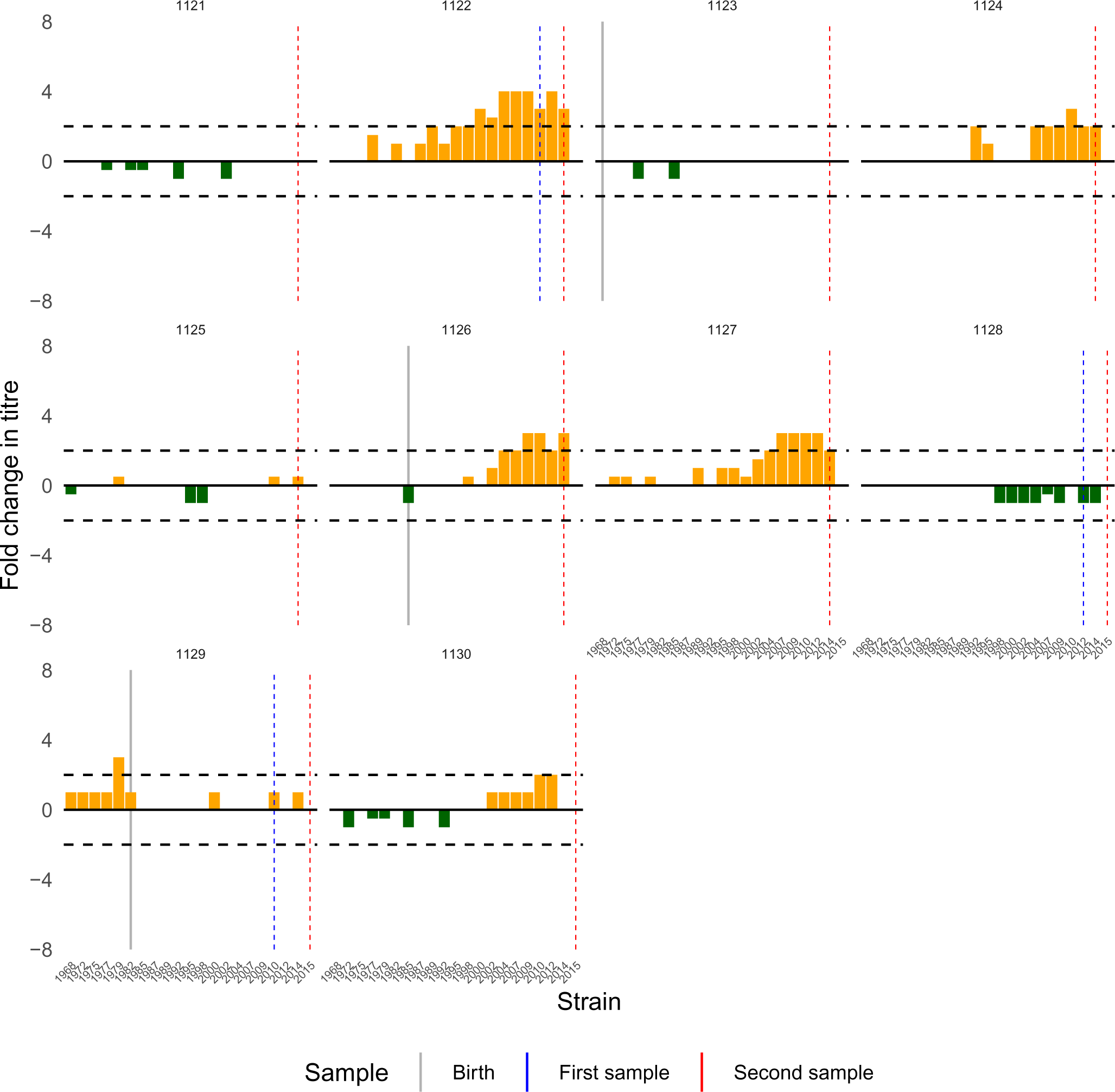
Observed changes in antibody titre for all individuals in the Fluscape cohort. As in Fig S20, but showing fold-change in titre against each strain between samples. Each subplot shows the change in antibody levels measured against each of the 20 H3N2 strains. The x-axis gives the isolation year of the measured strain. The vertical grey line shows the timing of birth or 1968, whichever was later. Bars are shaded orange to denote antibody boosting and green to denote antibody waning. Horizontal dash lines indicate 2-fold boosting or waning.

**Table S1:** self-reported vaccination status at time of first study visit (top) and between study visits (bottom). Percentages exclude individuals who declined to answer, were unsure, or had missing data.

| Self-reported vaccination at time of first serum sample (N=1005) |  |  |  |  |
| --- | --- | --- | --- | --- |
| Never vaccinated | This year | Last year | 2-5 years | 5+ years |
| 88.3% | 1.09% | 3.08% | 2.78% | 4.77% |
| Reported vaccination since last study visit |  |  |  |  |
| Visit 2 (N=822) |  | Visit 3 (N=1020) |  | Visit 4 (N=1119) |
| 1.70% |  | 1.57% |  | 0.536% |

**Table S2:** Estimated attack rates and infection patterns 2010-2014 prior to removing runs of consecutive infections. Percentages shown are posterior median and 95% credible intervals. Attack rate was defined as the proportion of individuals who were infected at least once in that year. “Reinfected” gives the percentage of people that were infected more than once in a year. Bottom table shows the percentage of individuals that were infected 0, 1, 2, 3, 4 or 5 times between 2010-2014 inclusive.

| Year | Measured strain | Estimated attack rate (%) | Reinfected within same year (%) |
| --- | --- | --- | --- |
| 2010 | A/Perth/2009 | 28.8% (25.2%-32.1%) | 4.25% (3.01%-5.4%) |
| 2011 | - | 42.9% (39.9%-46.2%) | 5.01% (3.49%-6.98%) |
| 2012 | A/Texas/2012 | 24.7% (20.9%-28.6%) | 2.26% (1.17%-3.61%) |
| 2013 | - | 20.8% (16.5%-24.9%) | 8.41% (7.09%-9.97%) |
| 2014 | A/Hong Kong/2014 | 57.9% (55.0%-60.5%) | 6.02% (5.00%-7.16%) |

| Proportion of individuals with different total number of infections from 2010-2014 |  |  |  |  |  |
| --- | --- | --- | --- | --- | --- |
| 0 | 1 | 2 | 3 | 4 | 5 |
| 12.2%<br>(11.2%-13.4%) | 32.7%<br>(30.7%-34.7%) | 32.7%<br>(30.8%-35.2%) | 16.8%<br>(15.0%-18.6%) | 4.78%<br>(3.67%-5.93%) | 0.531%<br>(0.177%-0.973%) |

### Supplementary Text 1

#### 1. Estimation of strain-specific measurement offsets

Our initial approach was to fit the full *serosolver* model exactly as described in the Materials and Methods, but without the inclusion of strain-specific measurement offsets (i.e., set all *χ_j_* to 0). Although convergence diagnostics were acceptable, we noted that estimated attack rates were extremely high (or low) in some time periods (Fig S16). These unusually high or low estimates were associated with systematic under- or over-estimation of expected log HI titres compared to the observed values (Fig S17). For example, model predicted titres are systematically lower than observations for the viruses that circulated in years 1985 and 2002, but systematically higher for years 1995 and 2010. These strain-specific biases are potentially problematic for the attack rate estimates, as the model can only account for elevated titres against a particular strain through adding more infections. If some viruses have systematically higher antibody titres simply due to measurement bias, then the model may incorrectly infer more infections than occurred.

These systematic measurement biases have a number of possible biological explanations. Serum potency and virus avidity, resulting in different haemagglutinin (HA) reactivity [88,89], are known sources of variation when performing antigenic cartography using ferret sera. This effect was noted by Fonville et al. when using locally-weighted multiple linear regression to fit antibody landscape surfaces to antibody titre data. Fonville et al. found that some viruses had systematically under-predicted values (e.g., NL/620/89) whereas others were systematically overestimated (e.g., Victoria/361/11) [37]. This phenomenon was also described by Bedford et al. when simultaneously performing antigenic cartography alongside phylogenetic tree reconstruction using both genetic and HI titre data [90]. Bedford et al. found models that explicitly estimated parameters for “virus avidity” to represent the contribution of virus-specific effects to observed titres were better supported than models that did not.

Including a strain-specific offset term as part of the observation process enables the antibody kinetics and infection model described here to account for these systematic biases. In lieu of generating these strain-specific offset terms from experimental data, we would ideally jointly infer these offsets alongside the infection histories and antibody kinetics parameters. However, our attempts to do so using the infection history prior used in the main text were unsuccessful and we could not generate converged chains. This is likely because the offset terms and attack rate estimates were highly correlated; elevated titres against a strain that circulated in a given year may be explained either by high seroresponse rates or systematic overestimation of titres to that strain.

Instead, we generated estimates for the measurement offsets using a modified version of the main text model. We fit the same antibody kinetics and infection histories to the full Fluscape dataset, but with three changes: (1) the observation model included an additional, estimated strain-specific offset parameter, *χ_j_*, for each measured strain as described in the Materials and Methods; (2) we estimated infection histories at an annual rather than 3-monthly resolution to reduce the parameter space to be explored; (3) we used a different infection history prior (prior version 3 in *serosolver*) which led to much better convergence and identifiability of the offset terms at the expense of less interpretable attack rate estimates. This alternative prior version places a Beta-Binomial prior on each individual’s expected number of lifetime infections and, unlike the prior used in the main text, assumes that an individual’s probability of infection in a given time period is unrelated to any other individual:

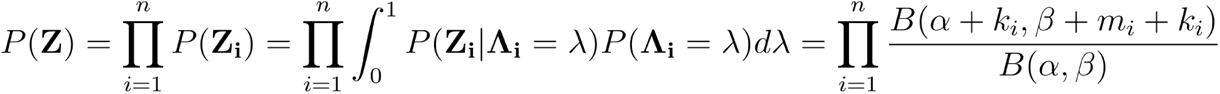

Where *k_i_* is the total number of infections experienced by individual *i*, and *m_i_* is the number of time periods individual *i* could be infected. This prior is suitable for fitting antibody landscapes to each individual’s antibody profile, but generates less interpretable attack rate estimates [35]. Model fitting was exactly the same as described in the main text, but 6 MCMC chains were run for only 12,000,000 iterations with the first 2,000,000 discarded as burn-in. Each strain-specific offset term was estimated under a multi-level model, assuming that each offset term in **χ** was drawn from the same normal distribution with mean 0 and unknown standard deviation.

Fig S22 shows that the model-predicted titres provided a better fit to the data with the added offset terms. We used the maximum posterior probability estimates for each χ as a fixed parameter for the model used in the main text (Table S3). Note that the model without the offset terms produced very similar antibody kinetics parameter estimates (Table S4), as well as age- and location-specific attack rate patterns, to those shown in the main text.

**Figure S22:**
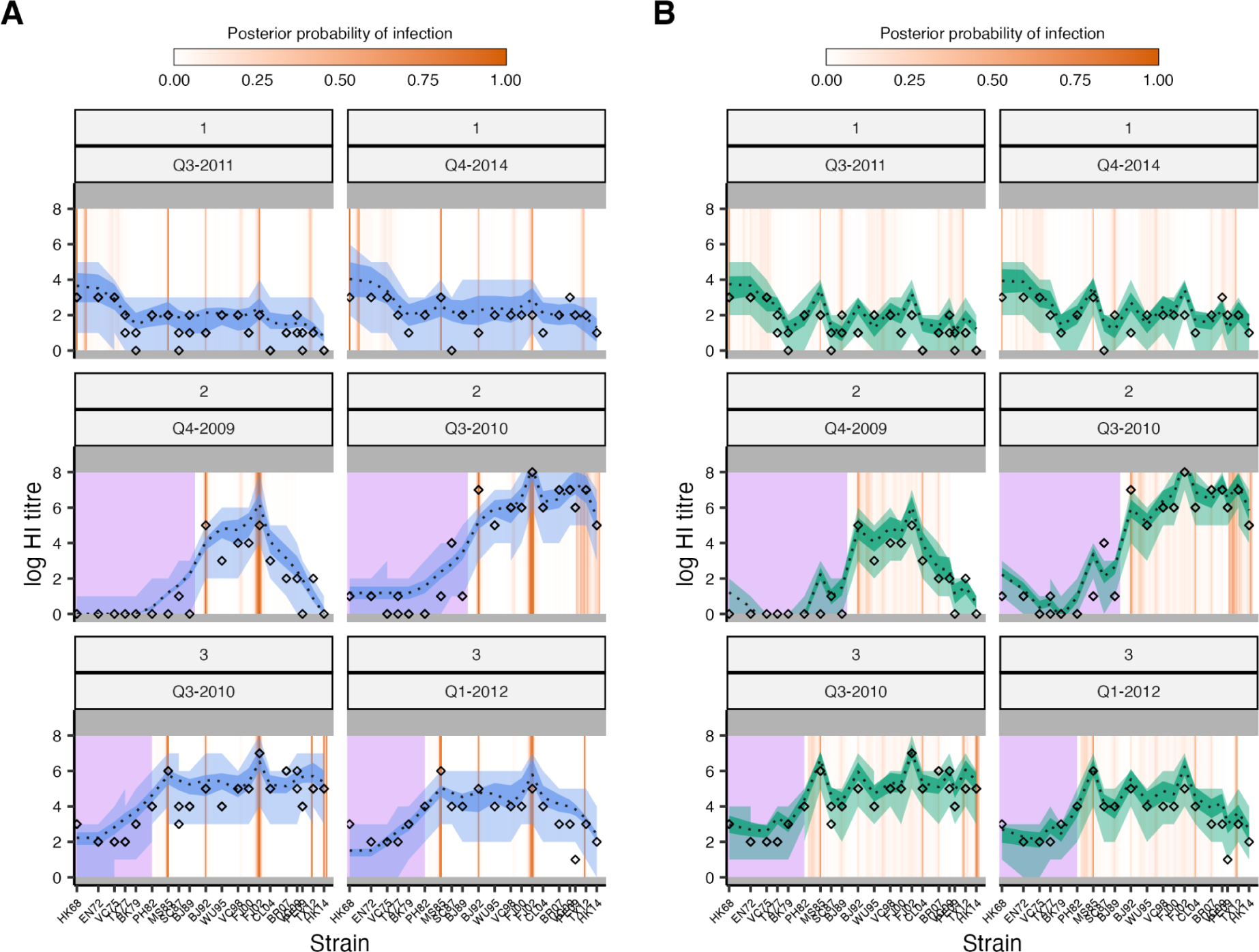
Comparison of estimated antibody landscapes from the same model fit with and without strain-specific measurement offsets. Rows represent individuals. Subplots show antibody titres based on serum samples taken at that time. X-axis represents a position along the antigenic summary path. Black diamonds show observed titres. Black line and blue or green shaded regions show posterior median and 95% credible intervals (CrI) on model-predicted latent titres (dark blue/green) and 95% prediction intervals (light blue/green). Orange bars show posterior probability of infection in that 3-month window. Grey rectangles denote the limit of detection of the HI assay. Purple rectangles show time periods prior to birth. (**A**) Model-predicted titres compared to observed HI titres at each sampling time for three randomly selected individuals, as in Fig S5, but from fitting the model without strain-specific measurement offsets. (**B**) as in (**A**), but with the estimated strain-specific measurement offsets included.

**Table S3:** Strain-specific measurement offset terms used in the main model fits. Values shown are maximum posterior probability estimates from a less flexible version of the model fit to the same data (described in Supplementary Text 1). Values shown to three significant figures.

| Strain | Fixed value |
| --- | --- |
| A/HongKong/1968 | 1.216 |
| A/England/1972 | 0.387 |
| A/Victoria/1975 | -0.643 |
| A/Texas/1977 | -0.476 |
| A/Bangkok/1979 | -1.392 |
| A/Philippines/1982 | -1.139 |
| A/Mississippi/1985 | 0.139 |
| A/Sichuan/1987 | -1.941 |
| A/Beijing/1989 | -2.302 |
| A/Beijing/1992 | -0.386 |
| A/Wuhan/1995 | -1.847 |
| A/Victoria/1998 | -0.387 |
| A/Fujian/2000 | -0.573 |
| A/Fujian/2002 | 0.746 |
| A/California/2004 | -0.515 |
| A/Brisbane/2007 | -0.655 |
| A/Victoria/2009 | 0.028 |
| A/Perth/2009 | -0.984 |
| A/Texas/2012 | 0.217 |
| A/HongKong/2014 | 0.027 |

**Table S4:** Estimated antibody kinetics parameters under the model without strain-specific measurement offsets.

| Parameter | Description | Estimate (posterior median; 95% credible intervals) |
| --- | --- | --- |
| $\mu_l$ | Long-term boosting | 1.54 (1.50-1.58) |
| $\mu_s$ | Short-term boosting | 1.95 (1.82-2.07) |
| $\tau$ | Antigenic seniority term | 0.0329 (0.0307-0.0357) |
| $\omega$ | Waning rate parameter for the short-term response (per 3-months) | 0.872 (0.776-1.02) |
| $\sigma_l$ | Long-term cross reactivity | 0.0896 (0.0889-0.0898) |
| $\sigma_s$ | Short-term cross reactivity | 5.17e-05 (5.7e-06-0.000278) |
| $\varepsilon$ | Standard deviation of observations | 0.874 (0.868-0.881) |

### Supplementary Text 2

#### 2. Scenario analyses using simulation-recovery experiments

##### 2.1 Overview

The full *serosolver* model framework is complex, with many components and parameters to be estimated or fixed. A concern is therefore the identifiability of the estimated model parameters and the potential biases introduced by inappropriate assumptions for the fixed components. To assess the ability of the *serosolver* framework to accurately recover infection histories and antibody kinetics parameters, we simulated a dataset matching the dimensions of the Fluscape survey and performed extensive simulation-recovery experiments testing the robustness of the framework to each step of our inference pipeline. The aim of this analysis was to test how simplifying assumptions and model misspecification might bias our estimates relative to the known ground truth.

##### 2.2 Simulation settings

We generated a simulated serosurvey of 1000 individuals of random ages uniformly sampled between 5 and 75 years. We simulated attack rates in 3-month time windows over a 46-year time period loosely based on estimates from the real data – per-quarter attack rates were randomly drawn from a log-normal distribution with mean 0.0375 and standard deviation 1.5 (on the natural scale). We assumed an attack rate of 0.6 in the first time period to represent the pandemic wave of H3N2 in 1968. Random infection histories for each individual were then simulated for each 3-month time period as Bernoulli trials with infection probability given by the simulated attack rates. Based on these infection histories, we then simulated each individual’s latent antibody kinetics using assumed model parameter values (Table S5). To reflect the Fluscape serosurvey, we assumed that each individual had two serum samples taken at random times from the last 24 time periods of the simulation, with two titre measurements at each sample against 24 strains uniformly distributed across the simulation period. A crucial addition to the simulation is the inclusion of strain-specific measurement offsets – measurements against each strain were assumed to be shifted relative to the true titre, where these measurement shifts were normally distributed with mean 0 and standard deviation 0.5. We generated random measurement shifts for each measured strain drawn from this distribution. Finally, we used the same antigenic map as described in the main text Materials and Methods.

**Table S5:** Description of antibody kinetics parameter values used for the simulation. Uniform priors were used for all parameters.

| Parameter | Description | Value | Prior lower bound | Prior upper bound | Estimate using the correctly specified model (posterior median; 95% credible intervals) |
| --- | --- | --- | --- | --- | --- |
| $\mu_l$ | Long-term boosting | 1.80 | 0 | 8 | 1.74 (1.71-1.77) |
| $\mu_s$ | Short-term boosting | 2.70 | 0 | 8 | 2.74 (2.53-2.91) |
| $\tau$ | Antigenic seniority term | 0.05 | 0 | 1 | 0.0511 (0.0486-0.0541) |
| $\omega$ | Waning rate parameter for the short-term response (per 3-months) | 0.20 | 0 | 1 | 0.216 (0.188-0.231) |
| $\sigma_l$ | Long-term cross reactivity | 0.10 | 0 | 1 | 0.101 (0.100-0.102) |
| $\sigma_s$ | Short-term cross reactivity | 0.03 | 0 | 1 | 0.0310 (0.0293-0.0329) |
| $\varepsilon$ | Standard deviation of observations | 1.00 | 0 | 25 | 1.00 (0.995-1.01) |

#### 2.3 Scenario analyses

##### 2.3.1 Re-estimating the strain-specific measurement offsets

The first step of the inference pipeline is to estimate the strain-specific measurement offset terms, where some strains have systematically higher or lower titres measured in the HI assay after accounting for differences in time-since-infection, exposure history and random measurement error. Following the same approach as outlined in Supplementary Text 1, we fitted the *serosolver* model to the simulated data to re-estimate the strain-specific measurement offsets. The recovered parameter estimates were close to those used to simulate the data, though there were some systematic biases (Fig S23). Most notably the total number of infections was overestimated, the antibody boosting parameters were underestimated and the cross-reactivity parameters were overestimated. The estimated measurement offset parameters were also biased but all in the right direction (Fig S24). Relative to the prior ranges (uniform between −3 and 3), the parameter estimates were close to their true values, giving us confidence that our estimates for the measurement offset terms were still informative and could thus be used as fixed values for subsequent model fits.

**Figure S23:**
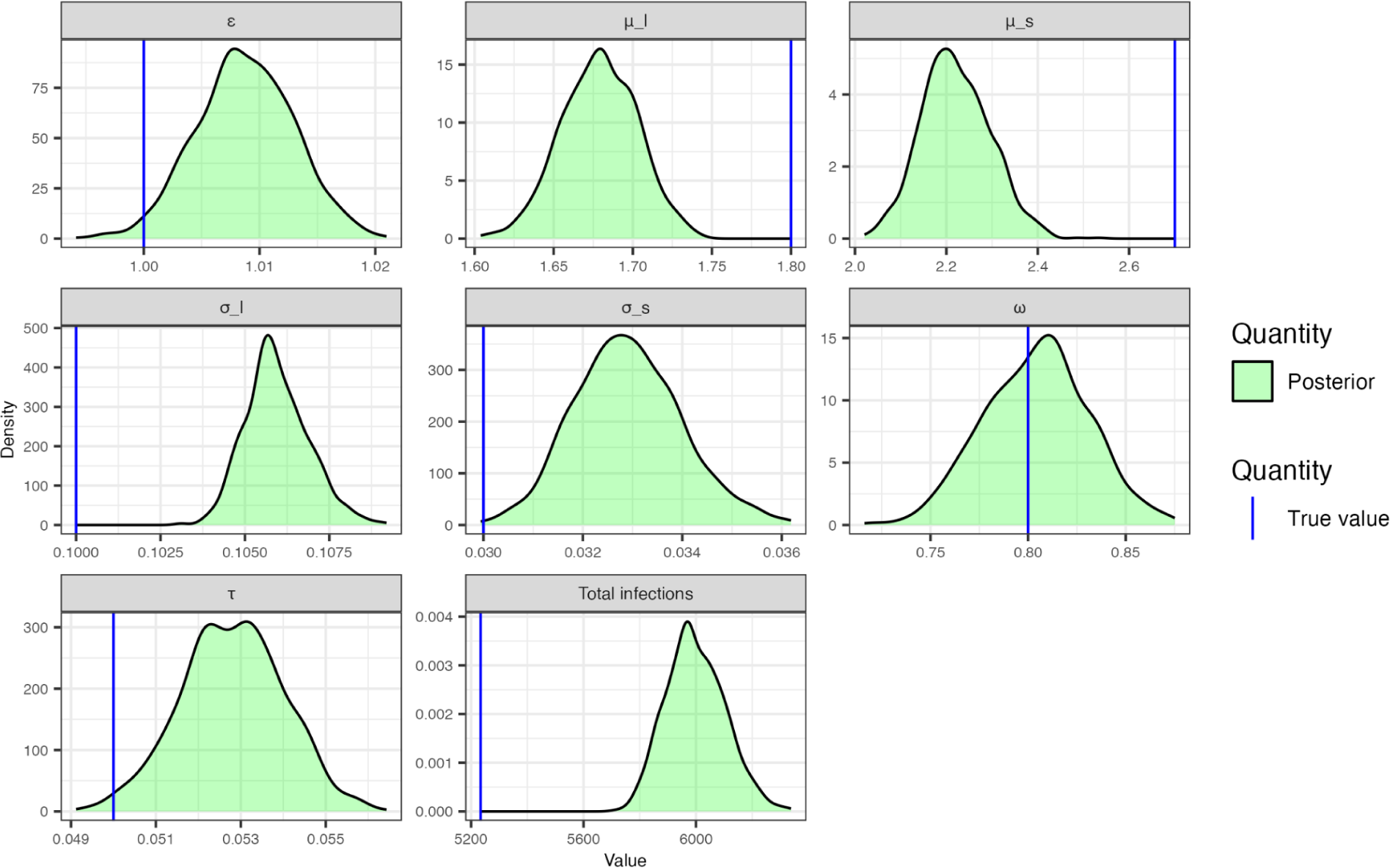
Estimated posterior distributions for antibody kinetics parameters (green shaded region) using the fitted model described in Supplementary Text 1 compared to true values used in the simulation (blue line).

**Figure S24:**
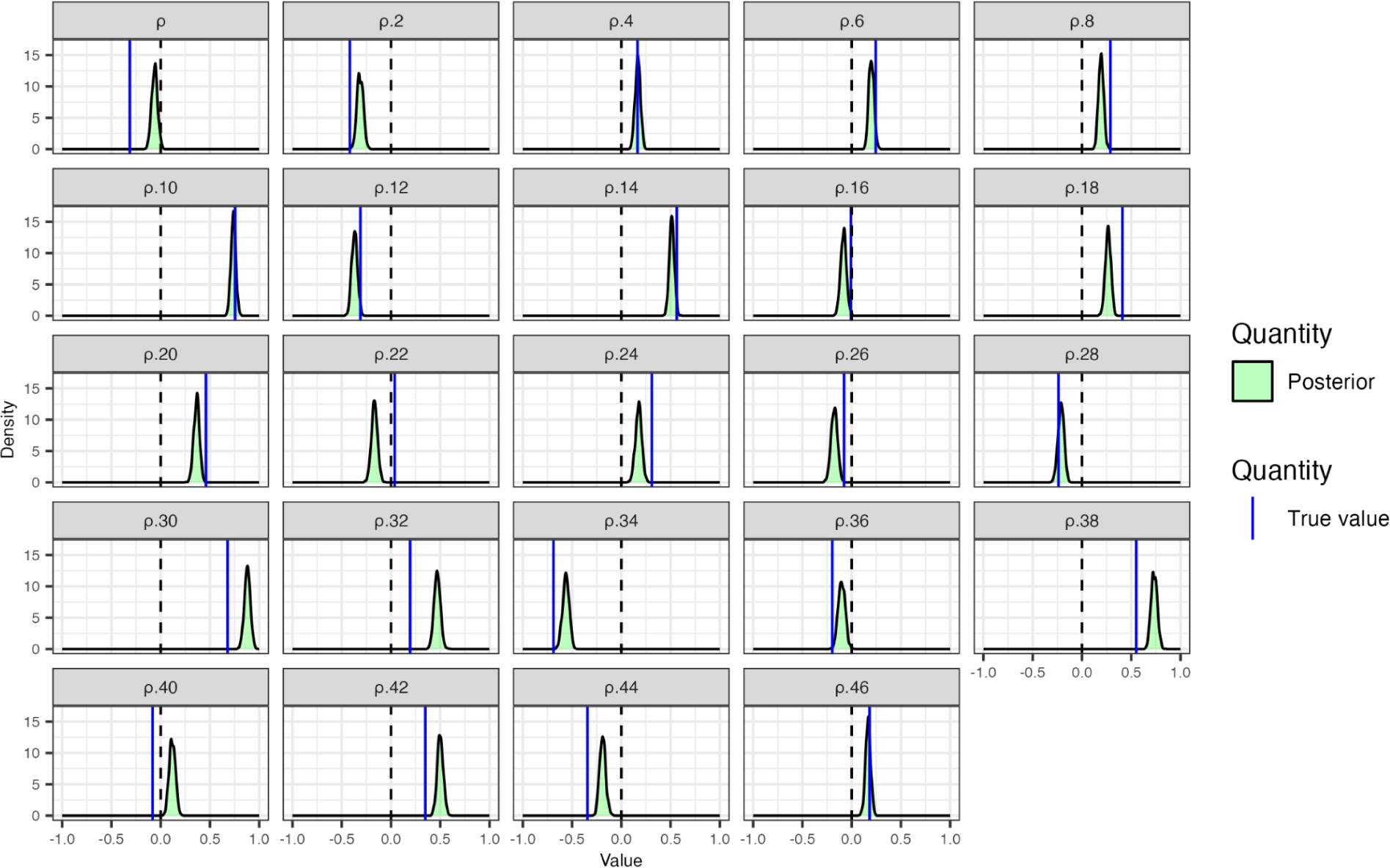
Estimated posterior distributions for strain-specific measurement offsets (green shaded region) using the fitted model described in Supplementary Text 1 compared to true values used in the simulation (blue line).

##### 2.3.2 Correctly specified model

Using the re-estimated strain-specific measurement offsets estimated from Section 2.3.1, we fit the *serosolver* model to the simulated data under the assumption that the model was correctly specified. This provides a sense check that the inference framework is able to accurately recover the antibody kinetics parameter estimates, attack rates and infection histories when the true generative model is known. 5 MCMC chains were run for 1,000,000 iterations with a 200,000 iteration burn in period. The model accurately re-estimated the true simulation parameter values (Table S5), though with a slight underestimation of the long-term boosting parameter (Fig S25).

**Figure S25:**
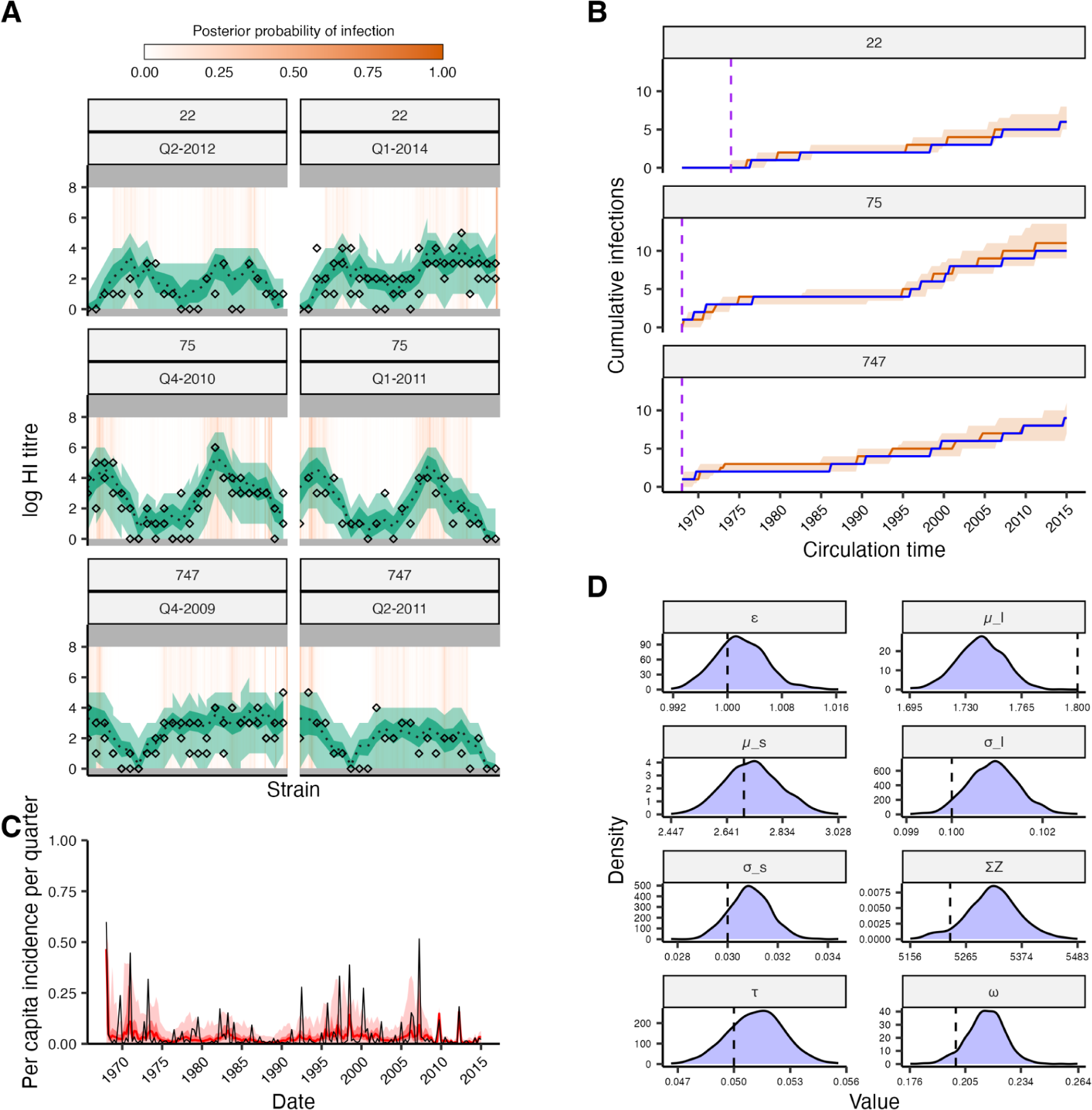
Assessment of model fitting accuracy based on simulated data. Results shown are from fitting the full model to simulated infection histories and antibody titres with known parameters. (**A**) Model-predicted titres compared to observed HI titres at each sampling time for three individuals. (**B**) Posterior median and 95% credible intervals (CrI) for the cumulative number of infections over time from birth (orange). Blue solid line shows the true, known cumulative number of infections. Purple dashed line shows the time of birth. (**C**) Posterior estimated per-capita per-3-month attack rates. Red line and shaded region shows posterior median and 95% CrI. Grey line shows the true values used for the simulation. (**D**) Shaded regions show posterior distributions of estimated antibody kinetics parameters. Dashed lines show the true value used for simulation. Note the x-axis range is small relative to the prior ranges in Table S4.

##### 2.3.3 Fitting the model ignoring strain-specific measurement offsets

To demonstrate the importance of considering the strain-specific measurement offsets, we compare the model estimates from Section 2.3.2 to results from fitting the model without accounting for the strain-specific measurement offsets, matching the workflow of the main text model. Both versions of the model were able to accurately re-estimate individual-level infection histories and fit to the antibody data well (Fig S25A/B & Fig S26A/B). However, the version without the measurement offset term led to biased attack rate estimates in some time periods (Fig S26C, e.g., 1969, 1975-1980, 1995-2000, 2005-2010). Furthermore, the version without the measurement offset terms led to greater bias in the estimated antibody kinetics parameters, particularly the observation error parameter, ε (Fig S26D). Overall, these results demonstrate that attempting to account for strain-specific measurement biases, even if incompletely, leads to parameter estimates which are closer to the true values than if these biases are ignored.

**Figure S26:**
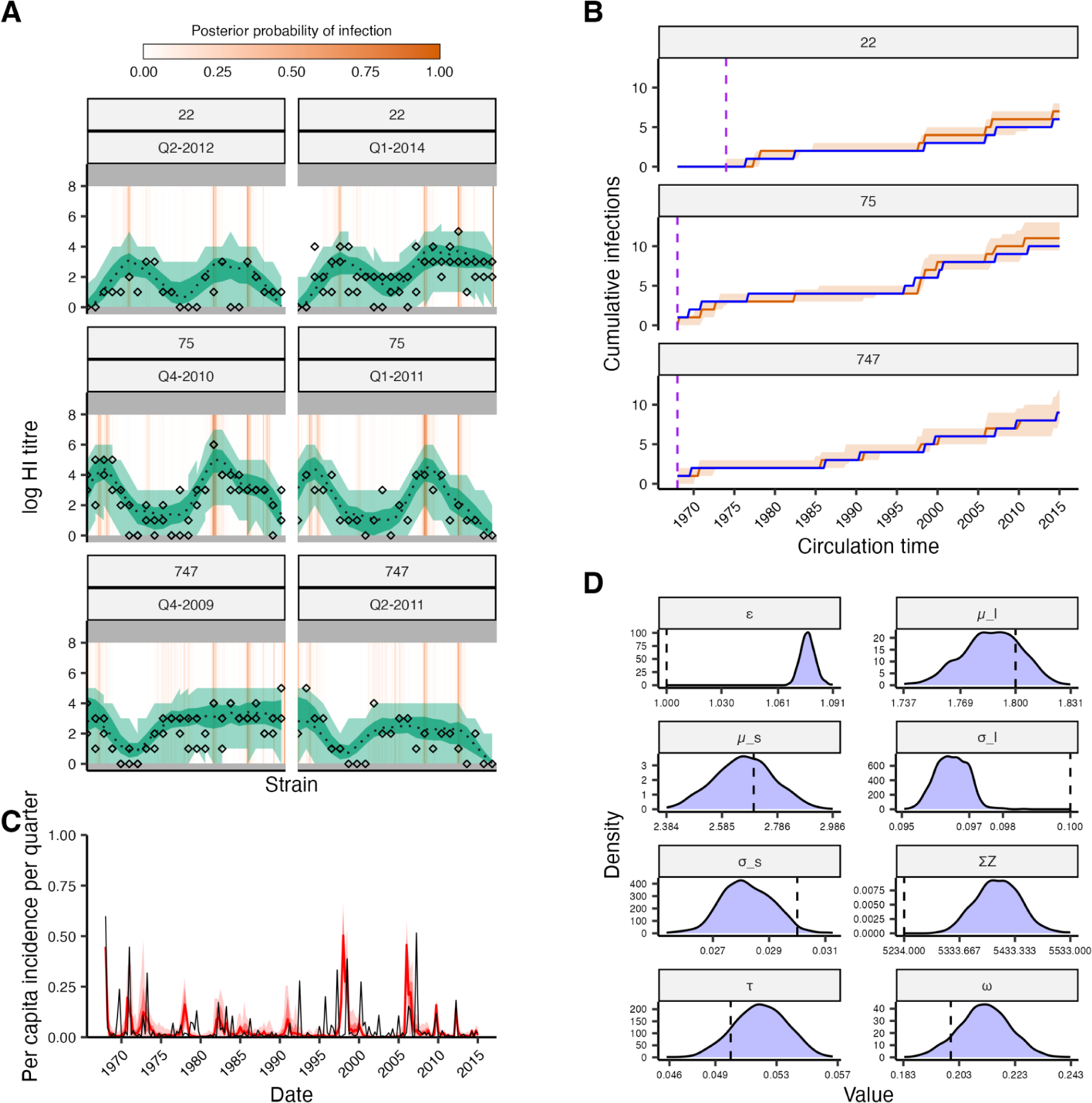
Assessment of model fitting accuracy based on simulated data when strain-specific measurement offsets are ignored. Results shown are from fitting the full model to simulated infection histories and antibody titres with known parameters where strain-specific measurement offsets are used in the simulation, but ignored in the fitted model. (**A**) Model-predicted titres compared to observed HI titres at each sampling time for three individuals. (**B**) Posterior median and 95% credible intervals (CrI) for the cumulative number of infections over time from birth (orange). Blue solid line shows the true, known cumulative number of infections. Purple dashed line shows the time of birth. (**C**) Posterior estimated per-capita per-3-month attack rates. Red line and shaded region shows posterior median and 95% CrI. Grey line shows the true values used for the simulation. (**D**) Shaded regions show posterior distributions of estimated antibody kinetics parameters. Dashed lines show the true value used for simulation. Note the x-axis range is small relative to the prior ranges in Table S5.

##### 2.3.6 Misspecifying the antigenic map

We do not know the strains which each individual was potentially exposed to, only the most likely antigenic cluster circulating in each time period. Thus, an individual’s infection history inferred using *serosolver* depends on the assumed strain an individual could be infected with in each time period, and how it contributes cross-reactive antibodies to their antibody profile. In theory, we might jointly estimate the antigenic map coordinates alongside the other model parameters, but at present this is computationally infeasible and thus we assume a fixed antigenic map for model fitting. As our estimates rely on the assumed position of each strain on the antigenic map, we performed sensitivity analyses to test how our estimates are affected by misspecifying the antigenic map.

First, we refit the model as described in Section 2.3.1 to the same simulated dataset, but instead of using the antigenic map used to simulate the data, we used an alternative antigenic map produced by Bedford et al for model fitting [90] (comparison in Fig S27). As with the antigenic map used in the main text, to generate a single antigenic position representing each strain we fitted a cubic smoothing spline through the antigenic coordinates of all strains on the map (here, smoothing parameter = 0.8). This scenario tests the assumption that the antigenic map used for fitting does not match the antigenic map underlying the data generating process. Parameter estimates, infection histories and attack rates were all largely accurate despite using the wrong antigenic map, though the timing of some elevated attack rates were slightly off (e.g., 1973, 1992, 2007). There were some small biases in the long-term boosting, cross-reactivity and observation error parameters (Fig S28). This suggests that although our quantitative estimates are affected by the assumed antigenic map, the overall trends are largely preserved.

**Figure S27:**
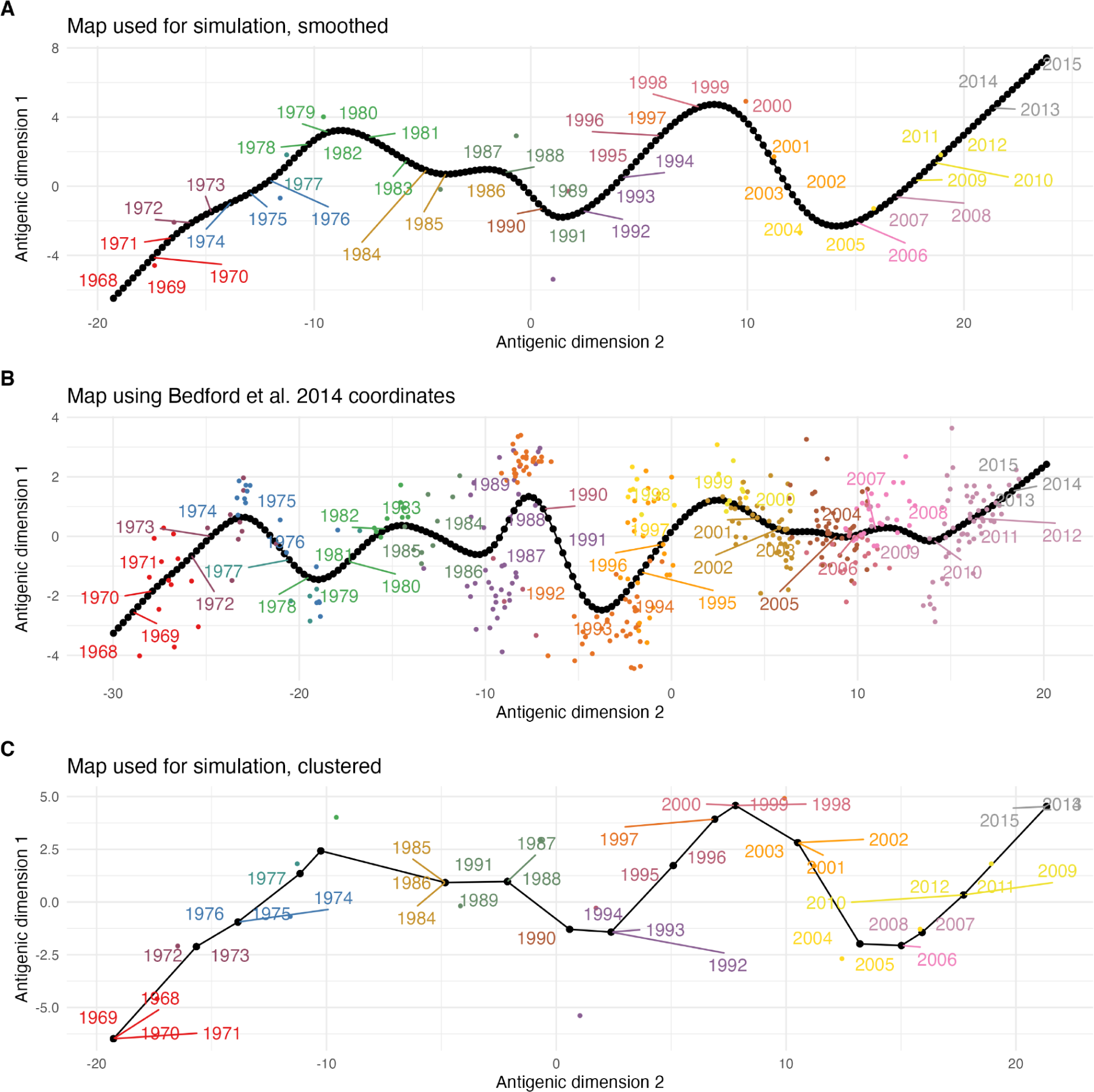
Antigenic maps used for the simulation (A) compared to the map produced using data from Bedford et al. (B), and the map which assumes a punctuated path through antigenic space (C). Axes show arbitrary antigenic dimensions. Coloured points show the position of individual strains – all labels of the same color correspond to the same antigenic cluster. Black line shows the fitted antigenic summary path for each map. Black dots show the antigenic position of the strain corresponding to each time period; (**A**) and (**B**) assume continuous evolution through antigenic space whereas (**C**) assumes that the same position is used for all strains within a cluster.

**Fig S28:**
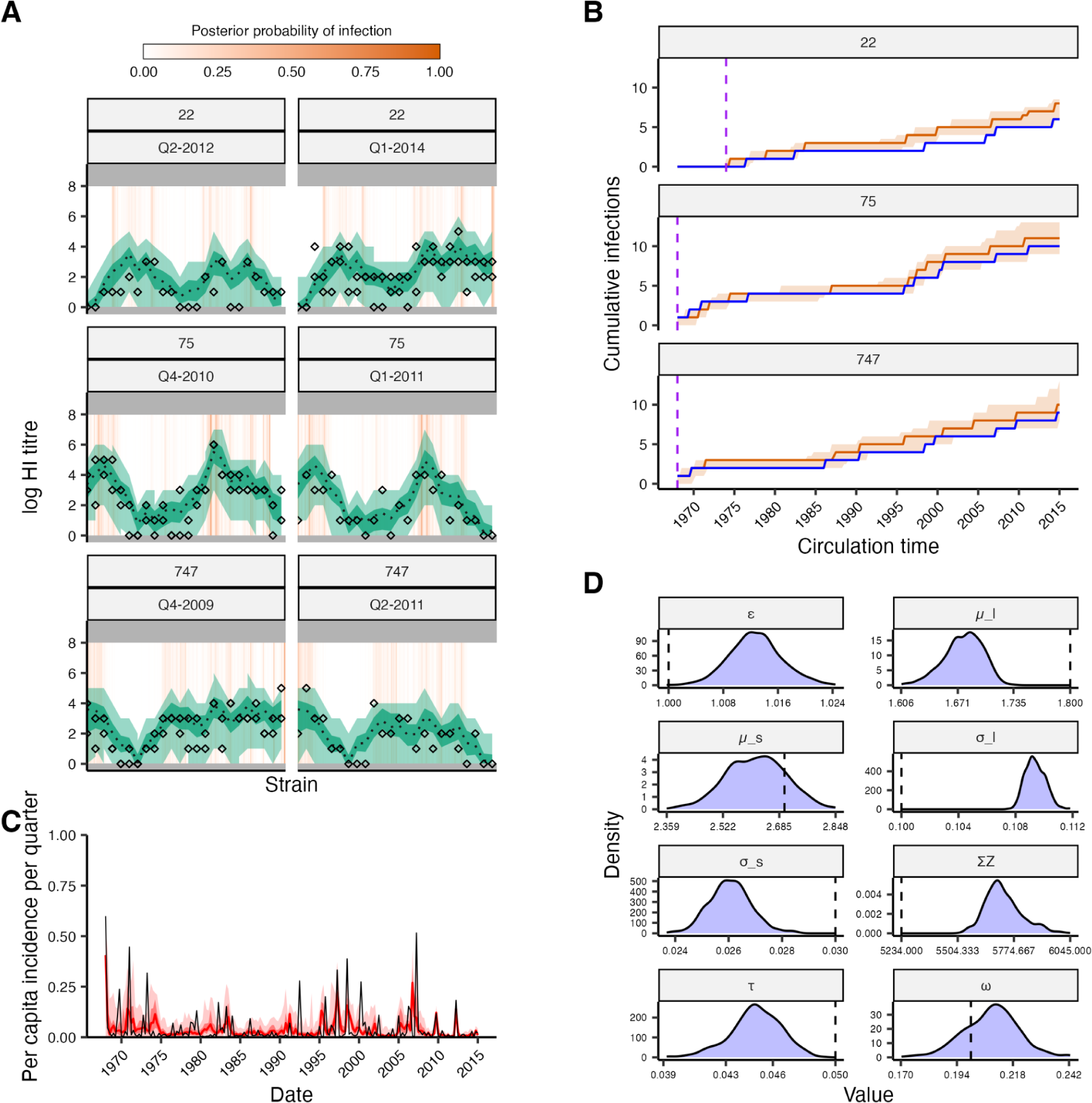
Assessment of model fitting accuracy when the antigenic map used for model fitting does not match the one used for simulation. Results shown are from fitting the full model to simulated infection histories and antibody titres with known parameters. (**A**) Model-predicted titres compared to observed HI titres at each sampling time for three individuals. (**B**) Posterior median and 95% credible intervals (CrI) for the cumulative number of infections over time from birth (orange). Blue solid line shows the true, known cumulative number of infections. Purple dashed line shows the time of birth. (**C**) Posterior estimated per-capita per-3-month attack rates. Red line and shaded region shows posterior median and 95% CrI. Grey line shows the true values used for the simulation. (**D**) Shaded regions show posterior distributions of estimated antibody kinetics parameters. Dashed lines show the true value used for simulation. Note the x-axis range is small relative to the prior ranges in Table S4.

Second, we generated an alternative simulated dataset using an antigenic map where antigenic evolution was assumed to follow punctuated jumps between clusters with all strains belonging to the same cluster having the same antigenic coordinates matching the first strain isolated from each cluster (Fig S27C), and then refit the model described in Section 2.3.1 using the smoothed antigenic map rather than the clustered map used for the simulation. This scenario tests the impact of our assumption that sequential strains follow a smooth and continuous path through antigenic space despite the true data generating process following punctuated jumps between clusters. We used the estimates from Du et al. to determine the time periods of cluster dominance – Du et al. predicted which A/H3N2 antigenic cluster likely circulated in China in each time period based on HA sequence data. For example, where Du et al. present a time range e.g., BE92 (1992-1995), we assumed that cluster circulated up to and not including the final time point in the range e.g., assume that BE92 circulated from January 1992 up to and including December 1994. We do not have cluster information after the PE09 cluster, and thus we assumed a new cluster emerged in 2013 (i.e., the PE09 cluster dominated for 4 years). Du et al. demonstrate that clusters are almost entirely dominant in the period in which they are circulating, though they rarely reach 100% frequency.

Parameter, infection history and attack rate estimates were mostly close to their true values despite misspecifying the antigenic map, with some differences in the estimated attack rates. Infection histories and model fits matched their true values well (Fig S29A&B), though the timing of elevated attack rates did not always align (Fig S29C). For example, the model missed the high attack rate in 1971, likely attributing these infections to the consecutive high attack rate periods from 1968. The high attack rates in 1992 and 2007 were also slightly misaligned, though the model was able to detect high incidence around those time periods. This misalignment is expected given how the antigenic map was misspecified. In the clustered map, we assumed that all strains in a cluster had the antigenic coordinates matching the first strain. However, in the smoothed map, each strain was instead assumed to be antigenically different from the previous one. Thus, when fitting with the smooth map, the model tends to attribute infections in a cluster to the first strain, as its antigenic coordinates are the closest to their true values used in the simulation. This tends to shift attack rates in those cluster periods earlier. Reassuringly, the model was still able to accurately recover the total number of infections in the simulation.

Some of the antibody kinetics parameters were slightly biased, with notable overestimation of the observation error parameter and waning rate, and slight underestimation of the long-term cross-reactivity parameter (Fig S29D). Bias in the waning rate parameter estimates is unsurprising given that attack rates were estimated to be slightly later than the truth for recent time periods which provide the model with information on these short-term kinetics. If infections are estimated later than their true timing, then a higher waning rate is required to reach the same low titres in a shorter time since infection. Bias in the observation error parameter is also unsurprising, as bias from model misspecification in the measurement offset term estimation stage (Section 2.3.1) becomes compounded when fitting the full model.

**Fig S29:**
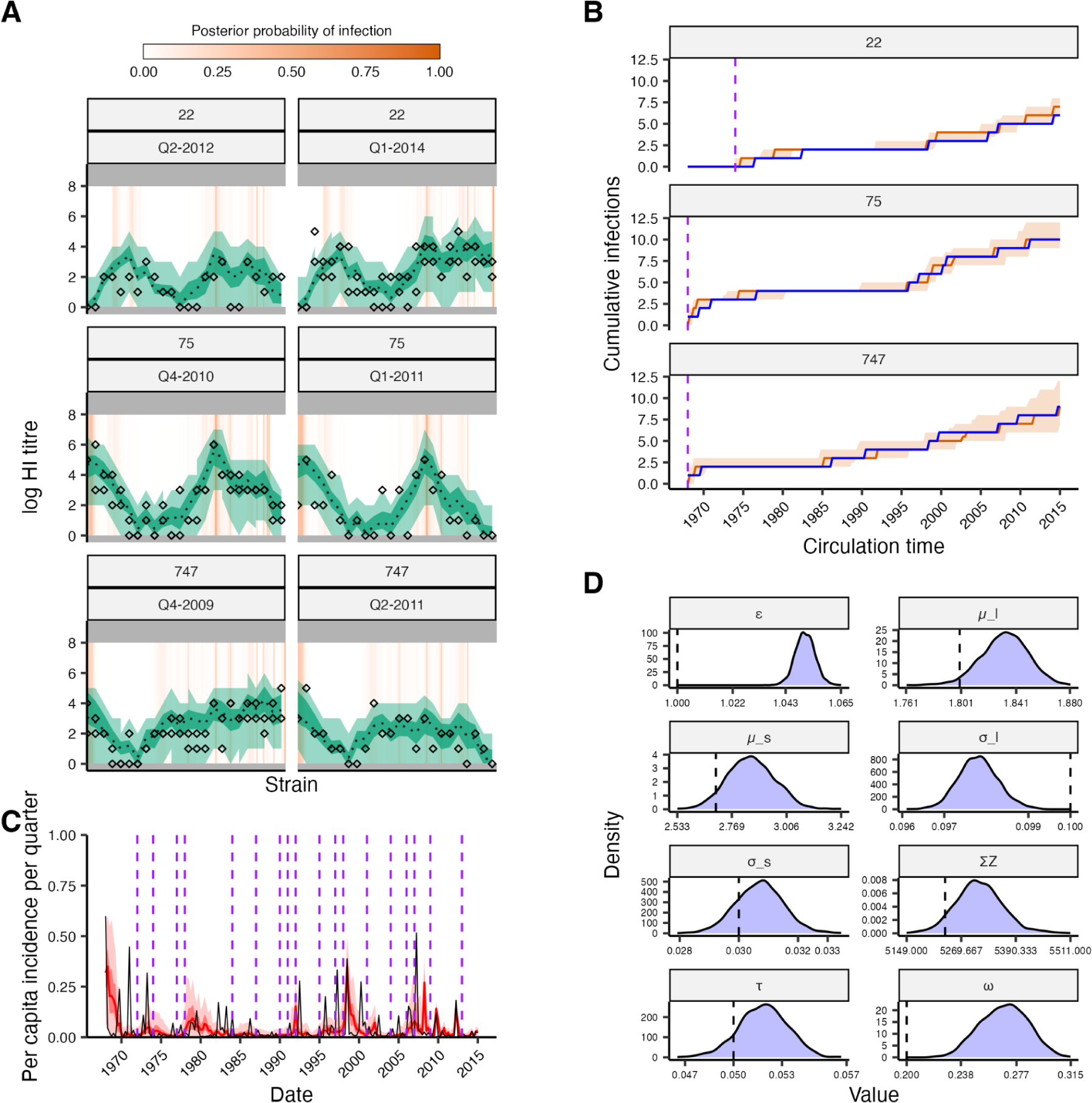
Assessment of model fitting accuracy based when fitting a model with a smoothed antigenic map to data simulated using a punctuated antigenic map. (**A**) Model-predicted titres compared to observed HI titres at each sampling time for three individuals. (**B**) Posterior median and 95% credible intervals (CrI) for the cumulative number of infections over time from birth (orange). Blue solid line shows the true, known cumulative number of infections. Purple dashed line shows the time of birth. (**C**) Posterior estimated per-capita per-3-month attack rates. Red line and shaded region shows posterior median and 95% CrI. Grey line shows the true values used for the simulation. Purple dashed lines show cluster transitions. (**D**) Shaded regions show posterior distributions of estimated antibody kinetics parameters. Dashed lines show the true value used for simulation. Note the x-axis range is small relative to the prior ranges in Table S5.

<< Separate file >>

**Supplementary Material 3: Distribution of quarterly attack rates by location over time.** Each colored point shows the inferred attack rate in each of the 40 locations, with size and shading reflecting the posterior median attack rate. Underlying the plot is a map of the study area, with each grid cell shaded by its log10 population density.

