## Supplementary figures and images for "Reconstructed influenza A/H3N2 infection histories reveal variation in incidence and antibody dynamics over the life course"

### Supplementary Material 3

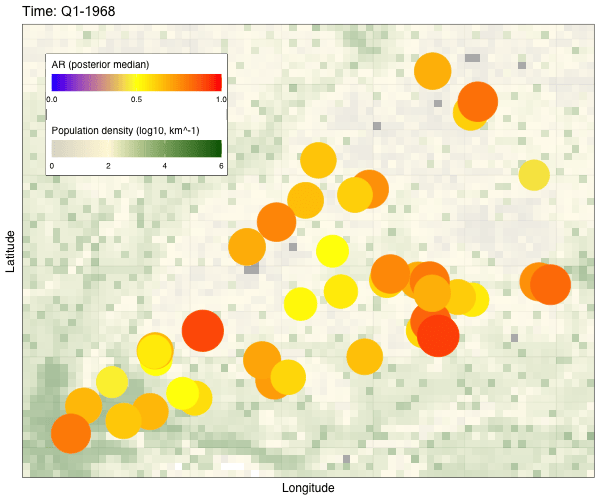
